# Trends in cancer incidence in younger and older adults: an international comparative analysis

**DOI:** 10.1101/2025.08.04.25332933

**Authors:** Amy Berrington de Gonzalez, Martina Brayley, Reuben Frost, Neal Freedman, Marc J. Gunter, Isobel Jackson, Patricia Lapitan, Meredith S. Shiels, Montserrat García-Closas

**Author notes:** Corresponding author: Amy Berrington de Gonzalez, DPhil, 15 Cotswold Rd, Sutton, SM2 5NG, UK.

## Abstract

**Background:** There is concern about widespread increases in cancer incidence rates in younger adults.

**Objective:** To compare international cancer incidence trends in younger adults (aged 20-49 years) and older adults (aged 50+ years).

**Design:** Surveillance study

**Setting:** 42 countries from Asia (n=11), Europe (n=22), Africa (n=1), North and South America (n=6), and Australasia (n=2) with annual cancer incidence data from 2003-2017 in the IARC Globocan database.

**Participants:** Adults aged 20+ years.

**Measurements:** Joinpoint regression to estimate the average annual percent change (AAPC) in cancer incidence rates for 13 cancers previously reported as increasing in multiple countries in younger adults (leukemia, breast, endometrial, colorectal, oral, kidney, liver, pancreatic, gallbladder, prostate, stomach, esophageal and thyroid cancer).

**Results:** Cancer incidence rates increased in younger adults in most (>75%) countries between 2003-2017 for six of the 13 cancers: thyroid (median AAPC=3.57%), breast (0.89%), colorectal (1.45%), kidney (2.21%), endometrial cancer (1.66%) and leukaemia (0.78%). Incidence rates for these cancers also increased in older adults in most countries (median AAPC thyroid=3%, breast=0.86%, kidney=1.65%, endometrial=1.20% and leukemia=0.61%). The exception was colorectal cancer which only increased in older adults in around half the countries (median AAPC=0.37%), and the AAPC was greater in younger than older adults in 69% of countries. For liver, oral, oesophageal and stomach cancer, rates decreased in younger adults in more than half the countries.

**Limitations:** Most countries with available data were high-middle income and results might not be generalizable.

**Conclusions:** Cancer incidence rates increased for several cancers in many of the countries studied; however, other than colorectal cancer, these increases occurred in both younger and older adults These findings can help inform future research, clinical and public health guidelines.

**Primary funding source:** The Institute of Cancer Research, UK and the NIH Intramural Research Program.

## Introduction

There is growing concern about widespread increases in cancer in younger adults following several studies that have reported increasing cancer incidence rates in those aged under 50 in many countries (1, 2). A wide range of potential causes have been proposed such as changes in diet, childhood obesity and antibiotic use (1, 2). Implications under discussion include new screening or surveillance guidelines and new research programmes to identify novel carcinogens in younger adults. However, the studies of international trends have mostly restricted their evaluation to cancer incidence in younger adults (1, 2), and have not directly compared these trends with those in older adults. Some studies have evaluated the trends in cancer incidence in younger and older adults in individual countries (3–5) but have not included an international comparison. Differences in calendar periods and age groups in these studies hinder comparisons across countries.

To address these limitations, we conducted an international comparative study where we estimated and compared the direction and magnitude of trends in cancer incidence in both younger (age 20-49 years) and older adults (age 50+ years). We evaluated thirteen cancers that were previously highlighted as increasing in younger adults in multiple countries between 2002 and 2012 (2). We used the IARC Global Cancer Observatory database (GLOBOCAN; https://gco.iarc.fr) and evaluated trends up to 2017, which enabled us to include 42 countries (6).

## Methods

### Data sources

We used the IARC GLOBOCAN cancer over time database and included all countries with cancer incidence data for the 15-year period 2003-2017 (2017 was the most recent data for the majority of countries)(20/06/2025). This resulted in 42 eligible countries; see Appendix Table 1 for details of the high-quality population-based national and regional cancer registries used by IARC Globocan for each country. We evaluated 13 cancers previously reported to be increasing in many countries in younger adults: colorectal, stomach, breast, prostate, endometrial, gallbladder, kidney, liver, esophageal, oral, pancreas, thyroid cancer and leukemia (2).

### Statistical methods

We defined cancer in younger adults as diagnoses at ages 20-49 years and in older adults as age 50+ years. Joinpoint regression was used to estimate the average annual percent change (AAPC) and 95% confidence intervals for cancer incidence trends (7–10). The AAPC is calculated by first fitting a piecewise linear function on log transformed age-standardised rates (ASR), allowing for the trends in cancer rates to vary across time. Models with different numbers of segments are fitted, and the one that best describes the data is chosen. The AAPC is then calculated as an average of the linear trends, weighted by the lengths of time that the trends spanned. A maximum of three joins were allowed for the 15-year period. The AAPC could not be estimated if there were 0 cases in any year, which did occur for the age range 20-49 years for the rarer cancers in many smaller countries. We present the full Joinpoint results to show if there were significant changes in the trends identified over the 15-year study period.

We provide several summary statistics for each cancer including: 1) the number of countries with increasing rates (defined as AAPC>0) for younger adults, 2) the number of countries with increasing rates in both younger and older adults (AAPC>0 for age<20-49 and AAPC>0 for age 50+), and 3) the number of countries where the rate of increase was greater for younger adults than older adults (defined as a +ve AAPC age 20-49y > AAPC age 50+). In this last comparison the AAPC age 50+ could be +ve or -ve.

We also compared the AAPCs for younger and older adults to test for significant differences (regardless of direction of the trend). We used a two-tailed z-test to test for differences in the AAPCs by age group when both models for each age group had joins. If there were no joins in either age-group we used a Monte Carlo simulation method (N = 10^7^) to test for the difference between the AAPCs (11). This was necessary as under 0 joins, the AAPC’s distribution is not normal, but based on a t-distribution. We used a significance threshold of p<0.05 to identify statistically significant findings. The combination of 42 countries, and 13 cancers resulted in more than 500 tests. To account for multiple testing and the increased risk of type I error rate, we calculated the Bayesian false-discovery probability (BFDP) for all tests with a p<0.05, following Wakefield’s approach (12) Findings with a BFDP<0.8 were considered noteworthy. We calculated BFDP for a prior probability of 0.05 for a true difference in log(AAPC), as differences by age group are moderately likely based on past evidence. The prior variance used in BFDP was calculated using the formula: 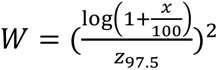 In this formula, *z*_97.5_ corresponds to the 97.5^th^ percentile of the standard normal distribution. This was chosen as it corresponds to the prior belief that an increase in AAPC from 0 to an AAPC greater than *x* occurs with a probability of 2.5%. We used a range of values of *x* (1%, 2.5%, 5%) to test the sensitivity of our results to the prior variance used.

We used Joinpoint Regression Program (version 5.0.2) (13). For all other statistical analysis and data manipulation we used R (R version 4.4.1; 2024-06-14 ucrt) (14).

### Role of the Funding source

The funders had no role in the design, conduct, interpretation, or publication of the study.

## Results

### Global analysis of 13 cancers

There were 42 countries eligible for inclusion in the international comparison located in Asia (n=11), Europe (n=22), Africa (n=1), North and South America (n=6), and Australasia (n=2).

Cancer incidence rates were increasing (AAPC>0) in younger adults in most (>75%) countries for 6 of the 13 cancers during 2003-2017 (Table 1, Figure 1): thyroid (median AAPC=3.57%), breast (0.89%), colorectal (1.45%), kidney (2.21%),, endometrial cancer (1.66%) and leukaemia (0.78%). For all these cancers, except colorectal cancer, incidence rates were also generally increasing in older adults (median AAPC thyroid=3%, breast=0.86%, colorectal=0.37%, kidney=1.65%, endometrial=1.20% and leukemia=0.61%). The rate of increase was greater in the younger adults for more than half the countries but 95% confidence intervals for the trends in the younger adults were often wide and included zero (Figure 1). Therefore, for these six cancers there were relatively few countries where the magnitude of the annual change in incidence rates in younger adults was statistically significantly different than those in older adults (Appendix Table 3). Furthermore, when accounting for multiple comparisons, very few countries showed noteworthy (BFDP<0.8) differences in trends between younger and older adults. The exception was colorectal cancer where the increasing trends in younger adults were greater than the trends in older adults for 69% of countries, and this difference was statistically significant in 38% of countries (19-26% for a range of priors with the BFDP<0.8).

**Table 1:** Number and % of eligible countries with increasing cancer rates (AAPC>0) in younger adults, and also in older adults and the number of countries with cancer rates in younger adults increasing at a faster rate than in older adults

|  | Eligible countries (age 20-49y) | ↑ age 20-49y | ↑ age 20-49y & ↑ 50+y | ↑ age 20-49y > 50+y |
| --- | --- | --- | --- | --- |
| Cancer | n (%)<sup>1</sup> | n (%) | n (%) | n (%) |
| Thyroid | 41 (100) | 38 (93) | 34 (83) | 25 (61) |
| Breast | 42 (100) | 38 (90) | 30 (71) | 23 (55) |
| Colorectum | 42 (100) | 37 (88) | 23 (55) | 29 (69) |
| Kidney | 40 (100) | 34 (85) | 29 (72) | 25 (62) |
| Endometrium | 36 (100) | 27 (75) | 22 (61) | 22 (61) |
| Leukaemia | 40 (100) | 30 (75) | 22 (55) | 21 (52) |
| Prostate | 35 (100) | 23 (66) | 17 (49) | 17 (49) |
| Gallbladder | 29 (100) | 18 (62) | 12 (41) | 11 (38) |
| Pancreas | 36 (100) | 22 (61) | 21 (58) | 10 (28) |
| Liver | 35 (100) | 17 (49) | 16 (46) | 7 (20) |
| Oral | 39 (100) | 18 (46) | 15 (38) | 13 (33) |
| Oesophagus | 32 (100) | 11 (34) | 8 (25) | 6 (19) |
| Stomach | 39 (100) | 9 (23) | 3 (8) | 8 (21) |

<sup>1</sup>AAPC was not estimable if countries had 0 cases in any year.

**Figure 1:**
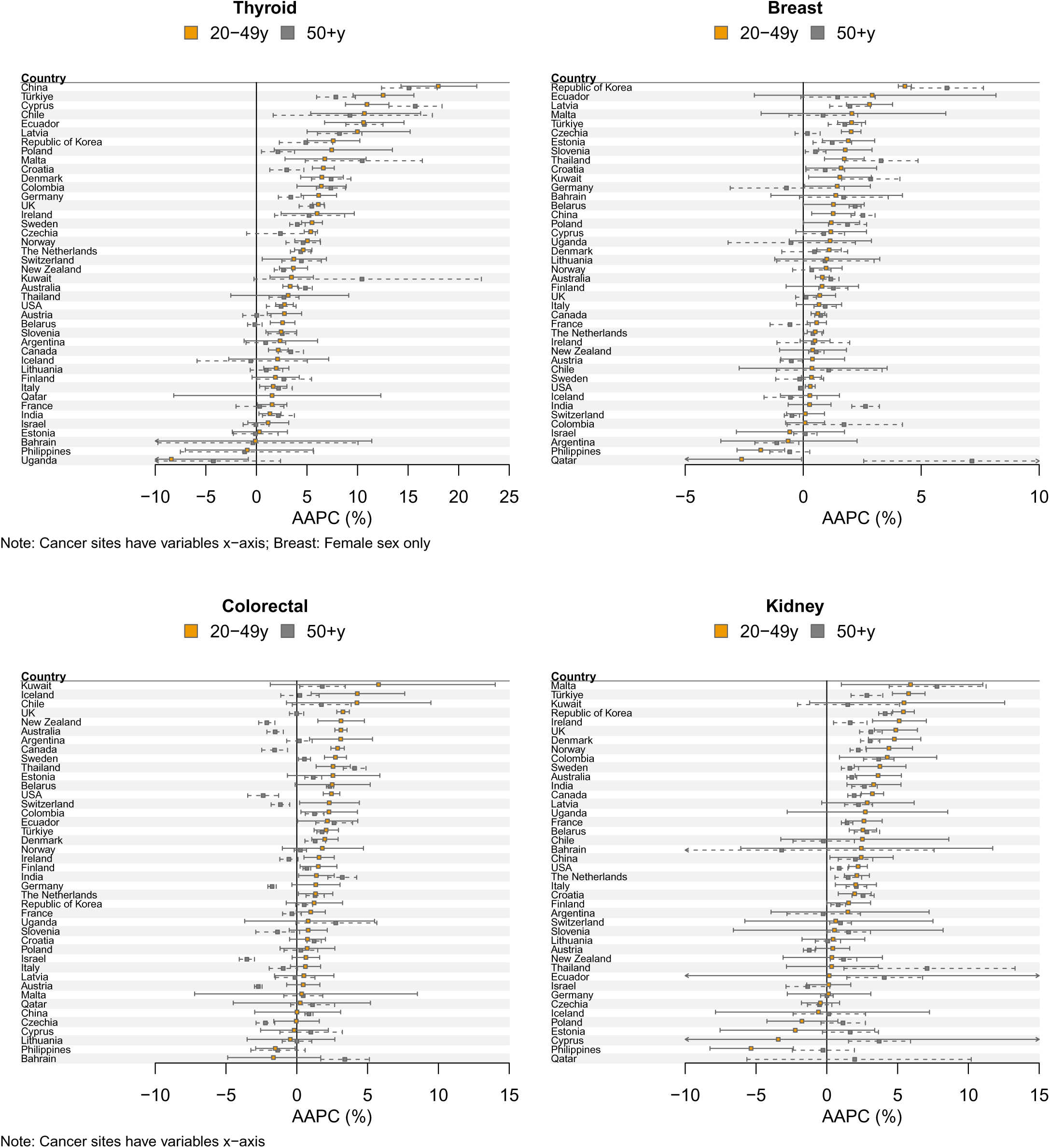
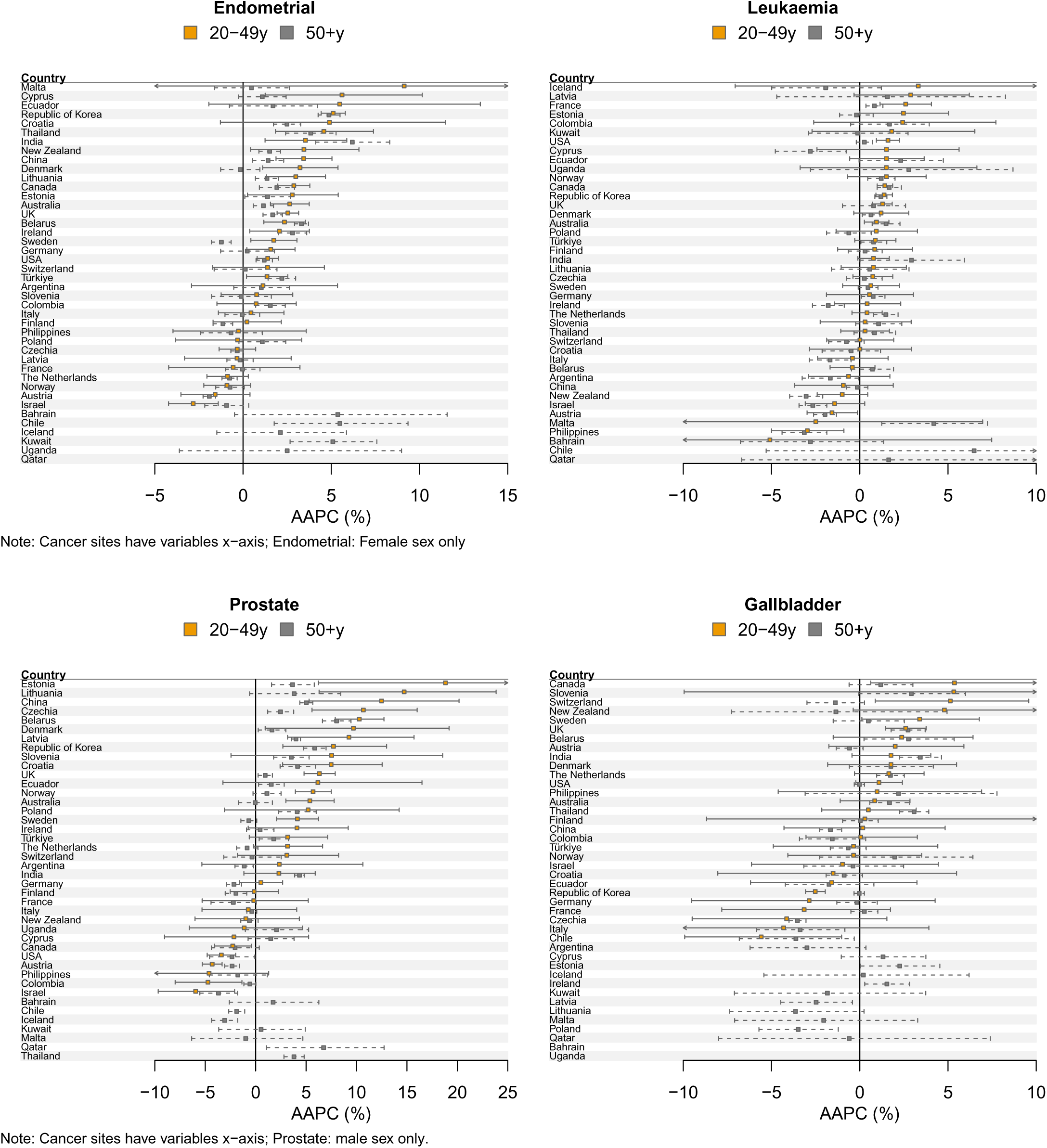
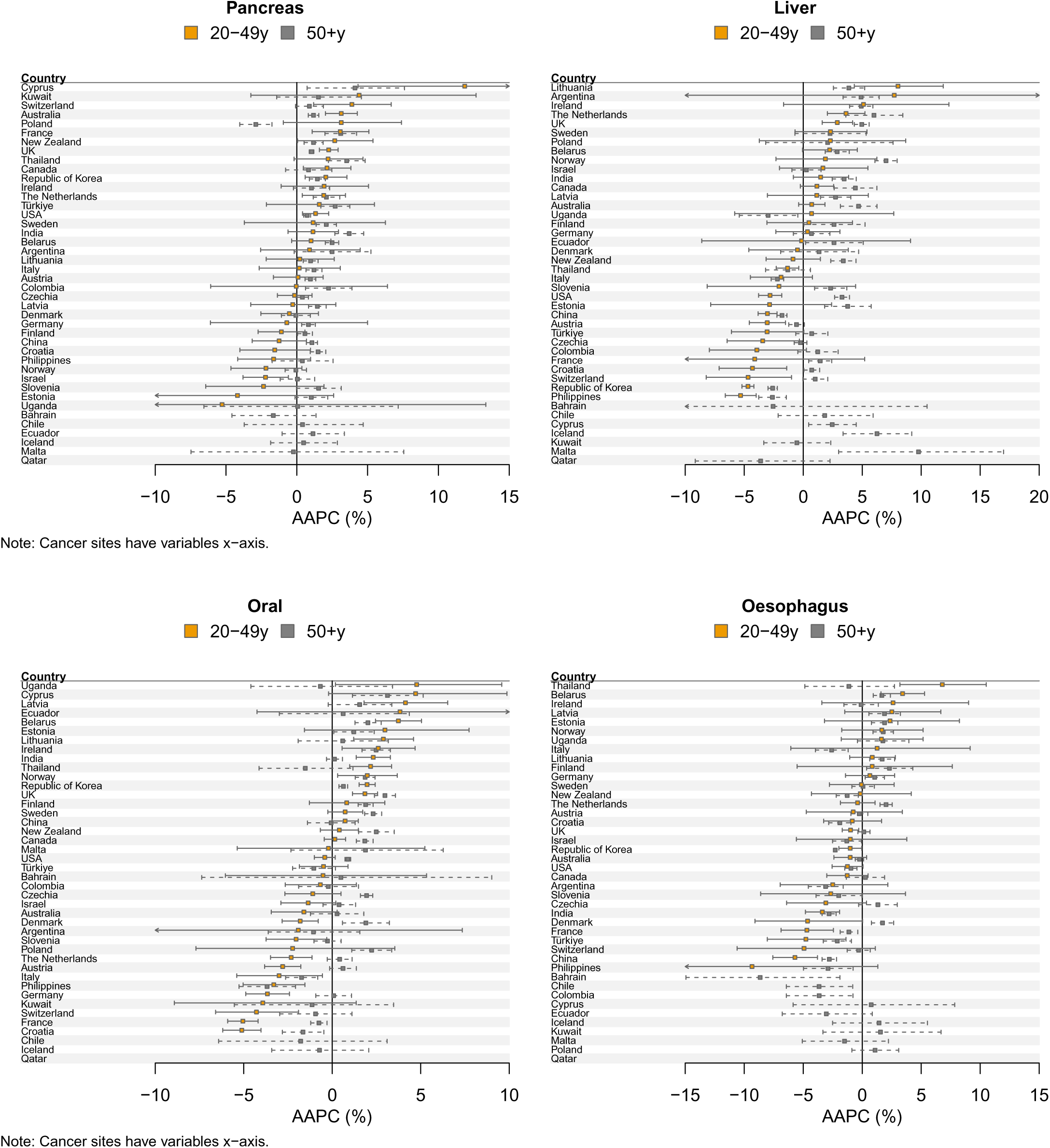
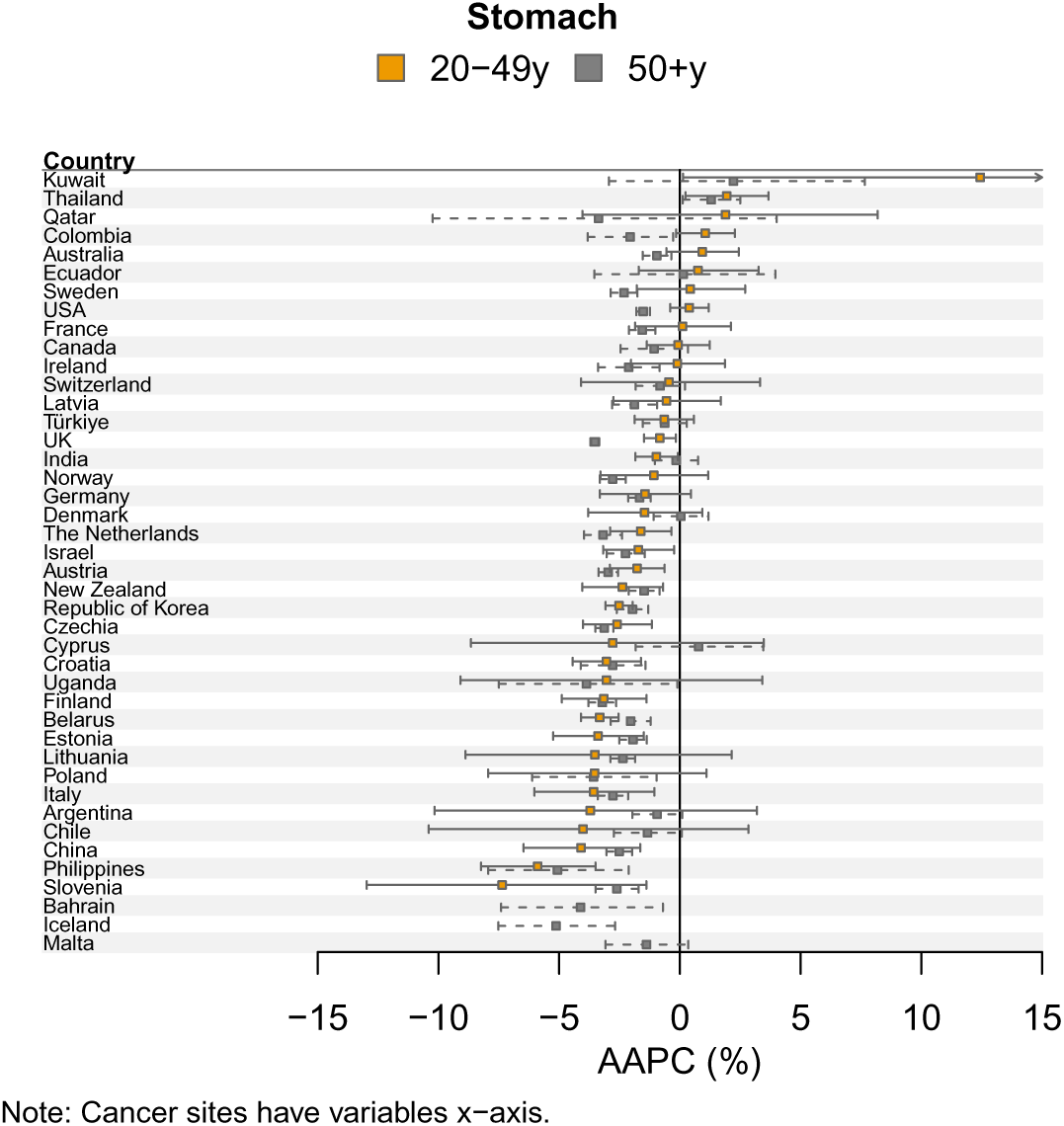
Average annual percent change (AAPC) in cancer incidence (2003-2017) by country and age group (sorted by AAPC in age group 20-49 years)

For prostate, gallbladder and pancreatic cancers, incidence rates were increasing in younger adults in just over half the countries (median AAPC prostate=3.2%, gallbladder=0.49% and pancreatic cancer=1% (Table 1, Figure 1). Often incidence rates were increasing also in older adults (median AAPC prostate=0.75%, gallbladder=-0.10% and pancreatic cancer=0.96%). The percentage of countries where incidence rates were increasing faster in younger adults was 49% for prostate cancer, 38% for gallbladder and 28% for liver cancer (Table 1). There was wide variation in the trends for prostate cancer in younger adults (AAPC range: to −6.0% to 18.9%) although confidence intervals were also wide.

For liver, oral, oesophageal and stomach cancers, incidence rates were decreasing in younger adults in more than 50% of the countries studied (median AAPC liver=-0.14%, oral=-0.42%, oesophageal=-0.92%, stomach=-1.62%) (Table 1, Figure 1). In older adults liver and oral cancer incidence rates were increasing in more than half of countries (median AAPC liver=2.17%, oral=0.49%) whereas for stomach and oesophageal cancer cancer incidence rates were decreasing in more than half of countries (median AAPC stomach=-2.05%, oesophageal=-0.25%).

When we examined the full Joinpoint results there were relatively few instances of more recent trends being qualitatively different from the AAPC. The exception was for thyroid cancer where in younger adultsJoinpoint identified a recent decreasing trend when the AAPC was positive in 9 of the 42 countries (Appendix Tables 4-16 and Appendix Figures 1-13).

When comparing results within UN regions the findings were also broadly similar (Appendix Figures 14-26). For example, thyroid and kidney cancer were increasing in younger adults in most countries with available data (>75%) in Asia, Europe and the Americas (Appendix Tables 17-19). Similarly, stomach, oral and esophageal cancer incidence rates were decreasing in younger adults in the majority of countries (<=50%) in these regions. Patterns differed for prostate cancer which was increasing in young adults in most European countries (80%) but not in the Americas (40%) or Asia (57%).

When comparing results for females and males the findings were broadly similar. For example, thyroid, colorectal and kidney cancer were increasing in younger adults in most countries and at a faster rate than in older adults in both females and males (Appendix Tables 20-21). Similarly for stomach and esophageal cancer these were decreasing in younger adults in the majority of countries for both females and males.

## Discussion

We conducted a systematic assessment of trends in cancer incidence in younger (20–49) and older adults (50+) for 13 cancer types that were previously reported as increasing in younger adults in multiple countries. In this international analysis of 42 countries with 15-year data from 2003-2017, we found that cancer incidence was increasing among young adults in the majority of countries for 6 of the 13 cancers: thyroid, breast, colorectal, kidney, endometrial cancer and leukaemia. Incidence rates for these cancers were also mostly increasing in older adults although with a lower trajectory than in younger adults. The exception was colorectal cancer where incidence rates were increasing in younger adults in most countries, but only increasing in older adults in about half these countries.

Ugai, Sasamoto (2) previously used the same database (IARC Globocan) to study the trends in cancer incidence in adults aged 20-49 years in 44 countries from 2002-2012 and reported that all 13 cancers were increasing in younger adults in multiple countries. Our analysis of these 13 cancers in 42 countries (38 included also in (2)) used similar methods (Joinpoint regression) but with more recent data up to 2017. For 6 of the 13 cancers highlighted in their study we found that the incidence rates were increasing in younger adults in the majority of countries (defined as >75%). However, for liver, stomach, oral and oesophageal cancer we found that rates were decreasing in more than half the countries. The prior study did not include an assessment of cancer incidence trends in older adults. This comparison provides important context that these increases are mostly not specific to younger adults. A recent analysis of trends in cancer incidence and mortality since 2010 in the US reported similar findings – most cancers that were increasing in younger adults were also increasing in older adults (5).

Zhao, Xu (1) used the Global Burden of Disease (GBD) 2019 study including 29 cancers, and estimated an absolute increase of total cancer cases in younger adults (age 14-49 years) of 79% from 1990 to 2019. They also described the increase in morbidity and mortality in the 204 countries studied, but did not provide a comparison of the cancer trends and increases in older adults. Our smaller sample size of 43 countries is due to the requirements in the Globocan database for countries to have high-quality cancer registry data. GBD uses extensive modelling to estimate rates from sparse data and this is likely to be a particular problem in cancer in younger people (15). Given the very different data sources and methods it is difficult to formally compare our findings.

Our findings indicate that changes in exposures rising cancer incidence rates for cancers such as kidney, thyroid and breast cancer are likely to be common across age groups, rather than specific to cancer in younger adults, since there were increases in both younger and older adults. Several studies have suggested that rising rates of obesity could be causing the increases in cancer in younger adults (2, 16). The cancers that we identified as increasing in younger and older adults in the majority of countries were all obesity-related cancers, and included some of those most strongly related to obesity (eg endometrial and kidney cancer) (17). Although some other obesity-related cancers were decreasing in younger adults in more than half the countries (eg stomach and esophageal cancer) (16). Another exception was breast cancer for which obesity is associated with lower risk in pre-menopausal women. There were no other clear patterns with respect to geographic regions or cancer types e.g. all digestive tract cancers. Formal analysis of obesity or other risk factors in these increasing cancer rates is warranted and requires long-term national survey data. .

Colorectal cancer was the only cancer that was increasing in younger adults in many countries and decreasing in older adults. This finding was consistent with a recent analysis of longer-term trends (1943–2017) in colorectal cancer across 50 countries (including the 42 countries included here) (18). This pattern could be due to exposure to novel carcinogens and/or effective screening in older adults. Colorectal cancer screening not only aids in early detection but also in prevention through the removal of pre-malignant lesions. This could distort comparisons in trends between younger and older populations by potentially masking increases in cancer incidence that might have occurred in the absence of screening. Screening or improved detection could also partly explain the reports that a higher proportion of colorectal cancers in younger adults are more aggressive and that the subtype distribution is different (19–22). The most notable qualitative differences in colorectal cancer trends by age group were found for the US, where there is widespread use of colorectal cancer screening by colonoscopy and where colorectal cancer incidence in screen-eligible age groups has been declining for several decades (23). This trend is now being mirrored in other countries that have implemented colorectal cancer screening programmes (24) where we observed divergent trends in younger and older adults (e.g. Canda, Australia, New Zealand, Austria, Switzerland and the UK). Increased surveillance, improved detection, or opportunistic testing could also explain some of the increases in other cancers. For example, a recent analysis highlighted the wide variation in prostate cancer incidence rates across European countries in contrast to much less variation in mortality rates and concluded that PSA testing could partly explain these differences (25). Detailed analyses of trends in cancer incidence rates (as opposed to proportions) by stage at diagnosis and age in countries, where these data are available, would help to further evaluate the role of screening and surveillance.

The strengths of our study include the systematic evaluation of trends in cancer incidence in younger and older adults for 13 cancers in 42 countries. This included systematic comparison for the difference in the trends by age group, which provided the important context of the increasing rates also in older adults. In addition, the Globocan database is restricted to countries with high-quality cancer registry data. Although, this and our requirement for data for data up until 2017 (resulted in most data being from high-middle income countries. Some of the countries do not have national cancer registries and rely instead on regional registries (e.g. SEER registries for the USA) and even within these high-quality registries there are differences in approaches to ascertaining cancer incidence that could have changed over time but should not differ within countries between younger and older adults (Appendix Table 1).

Other limitations include the use of the AAPC summary statistic when sometimes more complex trends were found by Joinpoint. This was necessary for parsimony given the large number of countries and cancers that were being studied. However, the AAPC better describes these shifting trends than alternative metrics such as the conventional estimated annual percentage change, which is based on fitting only one trend line to the data (8). We included the full Joinpoint results in the appendices and these show that there are few instances where there are qualitative changes in recent trends. We also did not examine trends in specific younger age groups (eg 20-29 year olds) again for parsimony and stability. Although this would not have altered our broad conclusion regarding which cancers were increasing in older as well as younger adults. Finally, the latest data for most countries in Globocan was 2017, which means that we could not study the most recent trends, but this still provides an additional five years of data compared to the previous international analysis (2).

Results from previous studies focused on cancer trends in younger adults have led to calls for research, treatment and public health interventions specifically for early-onset cancers (26). Our analysis suggests that in many countries these cancers are increasing in younger and older adults and that the rationale and potential implications of focusing new research studies only on younger adults should be carefully considered. Public heath guidelines also need to consider the absolute rates and number of cancer diagnoses. For example, in 2022 there were approximately 50,000 breast cancers diagnosed in women under age 50 in the US compared to 210,000 in women over aged 50 (6). In contrast there were only 4,000 prostate cancers diagnosed in men <age 50 versus 220,000 in men over aged 50. For colorectal cancer the number of cancer diagnoses were 21,000 (age<50) and 126,000 (age 50+). Despite some large relative increases in rates, therefore, most cancers remain rare in younger adults. Shiels et al (5) estimated the additional number of cancers diagnosed in younger adults in the US in 2019 compared to the expected number based on rates in 2010 with the largest increase for breast cancer of about 5,000 cancers and about 2,000 additional colorectal cancers.

In conclusion, cancer incidence rates increased for several cancers in many of the countries studied; however, other than colorectal cancer, increases occurred in both younger and older adults. Further research is needed into the causes of these increases. These results can help guide future research priorities and clinical priorities such as the need for special treatment strategies and supportive care needs for younger cancer patients.

## Data Availability

All data produced are available online at https://gco.iarc.who.int/overtime/en

https://gco.iarc.fr/overtime/en

## Funding

This work was funded by The Institute of Cancer Research, UK and the NIH Intramural Research Program.

## Author contributions

All authors contributed to conceptualization of the study and editing and review of the manuscript. ABdG and MGC led the statistical analysis and wrote the initial manuscript draft.

## Competing Interests

Authors declare no competing interests.

## Data and materials availability

All data are publicly available from the IARC Globocan webisite.

## Notes

### Competing Interest Statement

The authors have declared no competing interest.

### Author Declarations

Ervik M LF, Laversanne M, Colombet M, Ferlay J, Miranda-Filho A, Bray F. Global Cancer Observatory: Cancer Over Time Lyon, France: International Agency for Research on Cancer; 2024 [15 June 2025]. Available from: https://gco.iarc.who.int/overtime/en.

## References

1. Zhao J, Xu L, Sun J, Song M, Wang L, Yuan S, et al. Global trends in incidence, death, burden and risk factors of early-onset cancer from 1990 to 2019. BMJ Oncology. 2023;2(1). doi: 10.1136/bmjonc-2023-000049.

2. Ugai T, Sasamoto N, Lee HY, Ando M, Song M, Tamimi RM, et al. Is early-onset cancer an emerging global epidemic? Current evidence and future implications. Nat Rev Clin Oncol. 2022;19(10):656–73. Epub 20220906. doi: 10.1038/s41571-022-00672-8. PubMed PMID: 36068272; PubMed Central PMCID: PMC9509459.

3. Kehm RD, Yang W, Tehranifar P, Terry MB. 40 Years of Change in Age- and Stage-Specific Cancer Incidence Rates in US Women and Men. JNCI Cancer Spectr. 2019;3(3):pkz038. Epub 20190610. doi: 10.1093/jncics/pkz038. PubMed PMID: 31414075; PubMed Central PMCID: PMC6686848.

4. Shelton J, Zotow E, Smith L, Johnson SA, Thomson CS, Ahmad A, et al. 25 year trends in cancer incidence and mortality among adults aged 35-69 years in the UK, 1993-2018: retrospective secondary analysis. BMJ. 2024;384:e076962. Epub 20240313. doi: 10.1136/bmj-2023-076962. PubMed PMID: 38479774; PubMed Central PMCID: PMC10935512.

5. Shiels MS, Haque, Anika T., Berrington de González, Amy, Camargo, M. Constanza, Clarke, Megan A., Davis Lynn, Brittny C., Engels, Eric A., Freedman, Neal D., Gierach, Gretchen L., Hofmann, Jonathan N., Jones, Rena R., Loftfield, Erikka, Sinha, Rashmi, Morton, Lindsay M., Chanock, Stephen J. Trends in Cancer Incidence and Mortality Rates in Early-Onset and Older-Onset Age Groups in the United States, 2010–2019. Cancer Discovery. 2025;15(1):1–14. doi: 10.1158/2159-8290.CD-24-1678.

6. Ervik M LF, Laversanne M, Colombet M, Ferlay J, Miranda-Filho A, Bray F. Global Cancer Observatory: Cancer Over Time Lyon, France: International Agency for Research on Cancer; 2024 [15 June 2025]. Available from: https://gco.iarc.who.int/overtime/en.

7. Kim HJ, Fay MP, Feuer EJ, Midthune DN. Permutation tests for joinpoint regression with applications to cancer rates - PubMed. Statistics in medicine. 2000;19(3). doi: 10.1002/(sici)1097-0258(20000215)19:3<335::aid-sim336>3.0.co;2-z.

8. Clegg LX, Hankey BF, Tiwari R, Feuer EJ, Edwards BK. Estimating average annual per cent change in trend analysis. Stat Med. 2009;28(29):3670–82. doi: 10.1002/sim.3733. PubMed PMID: 19856324; PubMed Central PMCID: PMC2843083.

9. Fay MP, Tiwari RC, Feuer EJ, Zou Z. Estimating average annual percent change for disease rates without assuming constant change - PubMed. Biometrics. 2006;62(3). doi: 10.1111/j.1541-0420.2006.00528.x.

10. National Cancer Institute. Joinpoint Trend Analysis Software 2024 [29 August 2024]. Available from: https://surveillance.cancer.gov/joinpoint/.

11. National Cancer Institute. Average Annual Percent Change (AAPC) and Confidence Interval n.d. [22 August 2024]. Available from: https://surveillance.cancer.gov/help/joinpoint/setting-parameters/method-and-parameters-tab/apc-aapc-tau-confidence-intervals/average-annual-percent-change-aapc.

12. Wakefield J. A Bayesian measure of the probability of false discovery in genetic epidemiology studies - PubMed. American journal of human genetics. 2007;81(2). doi: 10.1086/519024.

13. Statistical Research and Applications Branch. Joinpoint Regression Program. Version 5.0.2 ed: National Cancer Institute; 2023.

14. R Core Team. R: A Language and Environment for Statistical Computing. R version 4.4.1 (2024-06-14 ucrt) ed: R Foundation for Statistical Computing; 2024.

15. Global Burden of Disease. Cancer Incidence, Mortality, Years of Life Lost, Years Lived With Disability, and Disability-Adjusted Life Years for 29 Cancer Groups From 2010 to 2019: A Systematic Analysis for the Global Burden of Disease Study 2019. JAMA Oncology. 2021;8(3). doi: 10.1001/jamaoncol.2021.6987.

16. Sung H, Siegel RL, Rosenberg PS, Jemal A. Emerging cancer trends among young adults in the USA: analysis of a population-based cancer registry. The Lancet Public Health. 2019;4(3). doi: 10.1016/S2468-2667(18)30267-6.

17. Kyrgiou M, Kalliala I, Markozannes G, Gunter MJ, Paraskevaidis E, Gabra H, et al. Adiposity and cancer at major anatomical sites: umbrella review of the literature. BMJ. 2017;356. doi: 10.1136/bmj.j477.

18. Sung H, Siegel RL, Laversanne M, Jiang C, Morgan E, Zahwe M, et al. Colorectal cancer incidence trends in younger versus older adults: an analysis of population-based cancer registry data. The Lancet Oncology. 2025;26(1). doi: 10.1016/S1470-2045(24)00600-4.

19. Yeo H, Betel D, Abelson JS, Zheng XE, Yantiss R, Shah MA. Early-onset Colorectal Cancer is Distinct From Traditional Colorectal Cancer. Clinical Colorectal Cancer. 2017;16(4). doi: 10.1016/j.clcc.2017.06.002.

20. Antelo M, Balaguer F, Shia J, Shen Y, Hur K, Moreira L, et al. A High Degree of LINE-1 Hypomethylation Is a Unique Feature of Early-Onset Colorectal Cancer. PLOS ONE. 2012;7(9). doi: 10.1371/journal.pone.0045357.

21. Lawler T, Parlato L, Warren Andersen S. Frontiers | The histological and molecular characteristics of early-onset colorectal cancer: a systematic review and meta-analysis. Frontiers in Oncology. 2024;14. doi: 10.3389/fonc.2024.1349572.

22. Willauer AN, Liu Y, Pereira AAL, Lam M, Morris JS, Raghav KPS, et al. Clinical and molecular characterization of early - onset colorectal cancer. Cancer. 2019;125(12). doi: 10.1002/cncr.31994.

23. National Cancer Institute. Cancer Trends Progress Report Bethesda, MD: NIH; 2024 [29 August 2024]. Available from: https://progressreport.cancer.gov/detection/colorectal_cancer.

24. Ebell MH, Thai TN, Royalty KJ. Cancer screening recommendations: an international comparison of high income countries. Public Health Rev. 2018;39:7. Epub 20180302. doi: 10.1186/s40985-018-0080-0. PubMed PMID: 29507820; PubMed Central PMCID: PMC5833039.

25. Vaccarella S, Li M, Bray F, Kvale R, Serraino D, Lorenzoni V, et al. Prostate cancer incidence and mortality in Europe and implications for screening activities: population based study. BMJ. 2024;386. doi: 10.1136/bmj-2023-077738.

26. Hamilton A, Coleman H. Shifting tides: the rising tide of early-onset cancers demands attention - PubMed. BMJ oncology. 2023;2(1). doi: 10.1136/bmjonc-2023-000106.

