## Supplemental Tables and Figures for "Trends in cancer incidence in younger and older adults: an international comparative analysis"

Supplemental Material: Table of Contents

| |
|---|
| Appendix Figures 1-13: Age-standardised incidence rates (ASR) per 100,000 for each cancer by age |
| Appendix Figure 14-26: Average annual percent change (AAPC) in cancer incidence (2003-2017) for each cancer by age, country & UN region |
| Appendix Table 1: Source of cancer incidence data by country in GLOBOCAN |
| Appendix Table 2: ICD-10 codes for cancers based on Globocan |
| Appendix Table 3: AAPC comparisons by age (Bayes False Discovery Probability) |
| Appendix Tables 4-16: Segment specific annual percentage change (APC) for cancer incidence trends from 2003 to 2017 by country |
| Appendix Tables 17-19: Summary statistics by cancer and UN region |
| Appendix Tables 20-21: Summary statistics by cancer and sex |

**Appendix Figure 1: Age-standardised incidence rates (ASR) per 100,000**

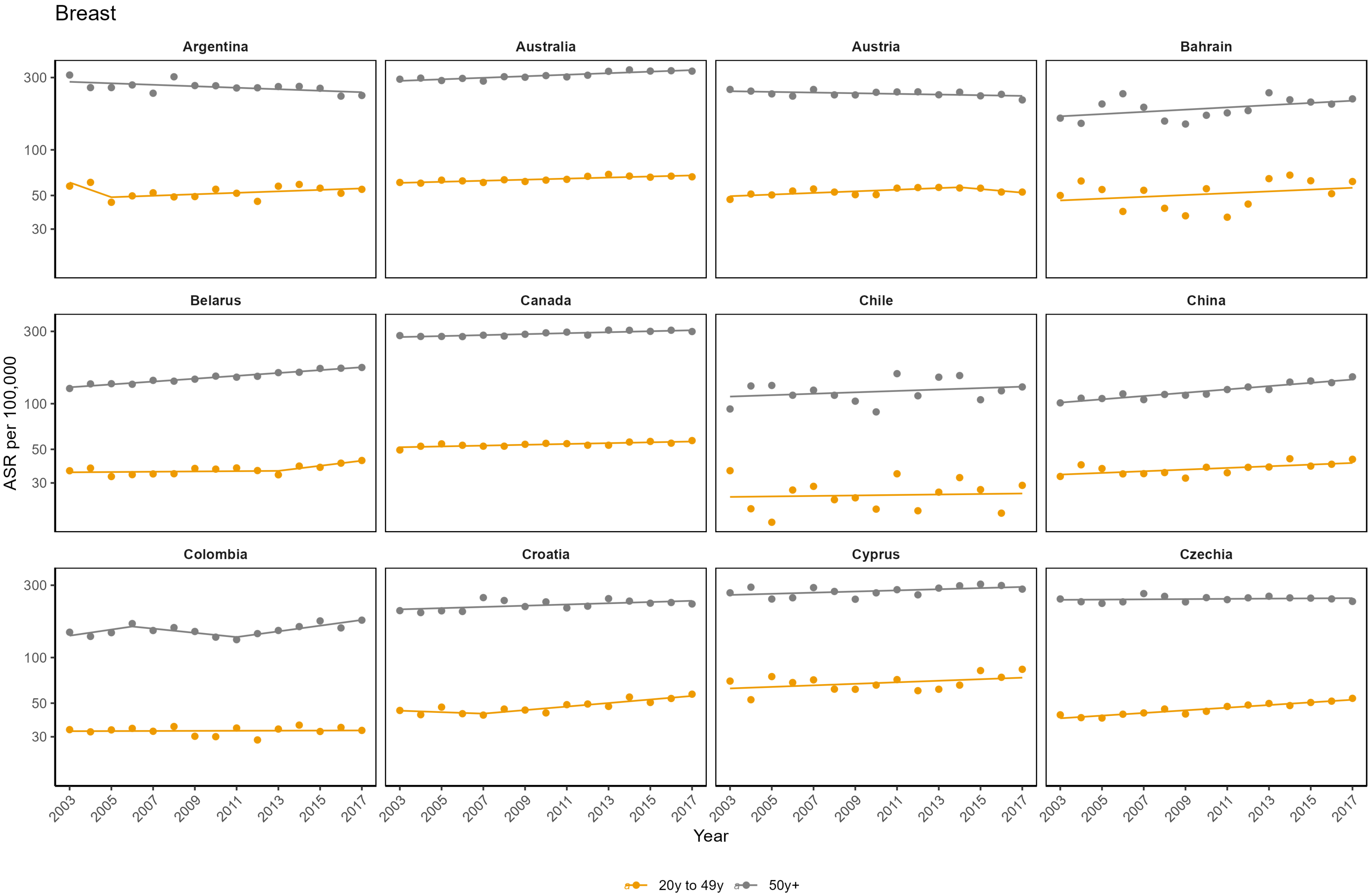

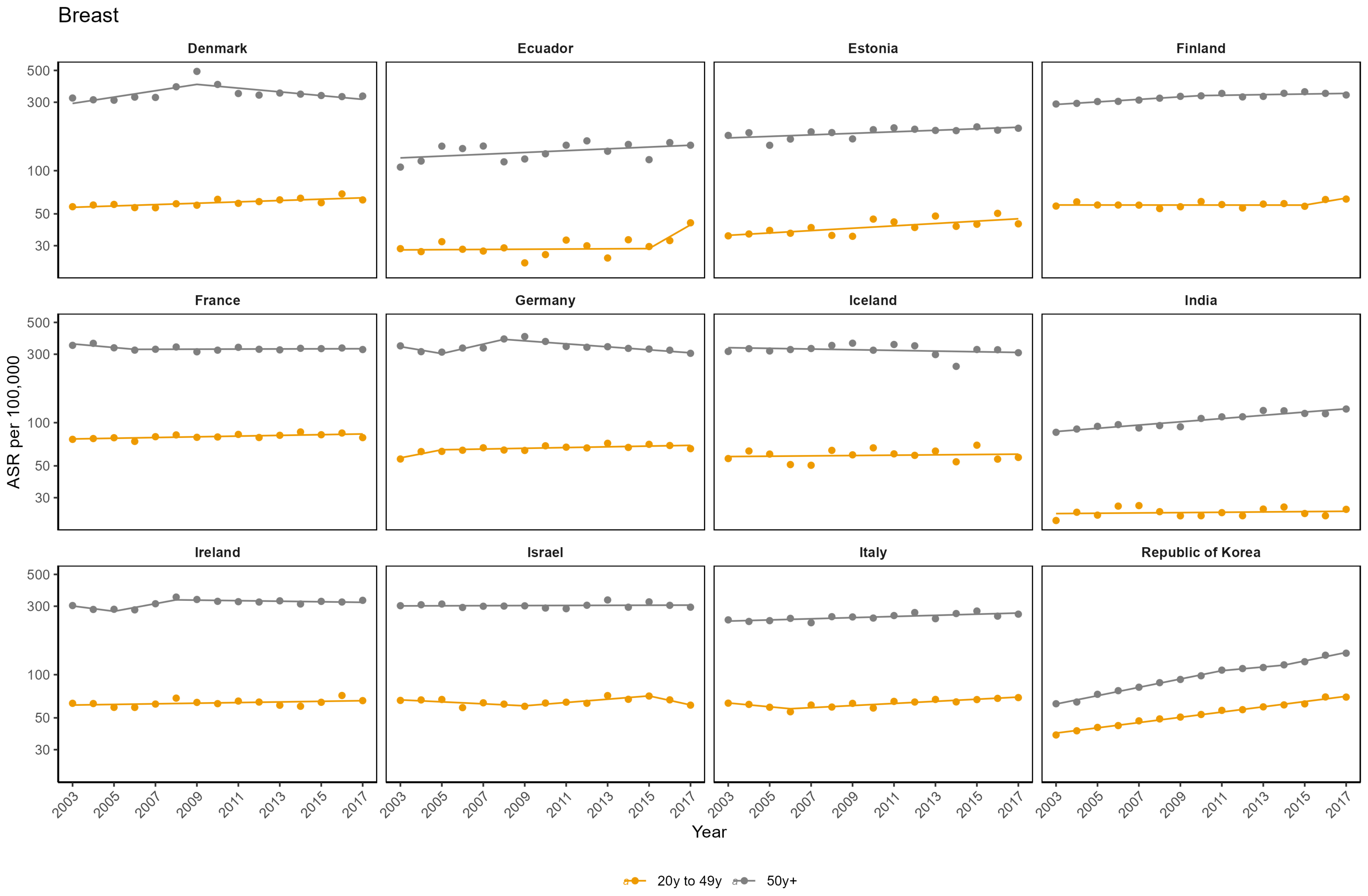

Breast
Denmark
Ecuador
Estonia
Finland
France
Germany
Iceland
India
Ireland
Israel
Italy
Republic of Korea
ASR per 100,000
500
300
100
50
30
2003
2005
2007
2009
2011
2013
2015
2017
Year
20y to 49y
50y+

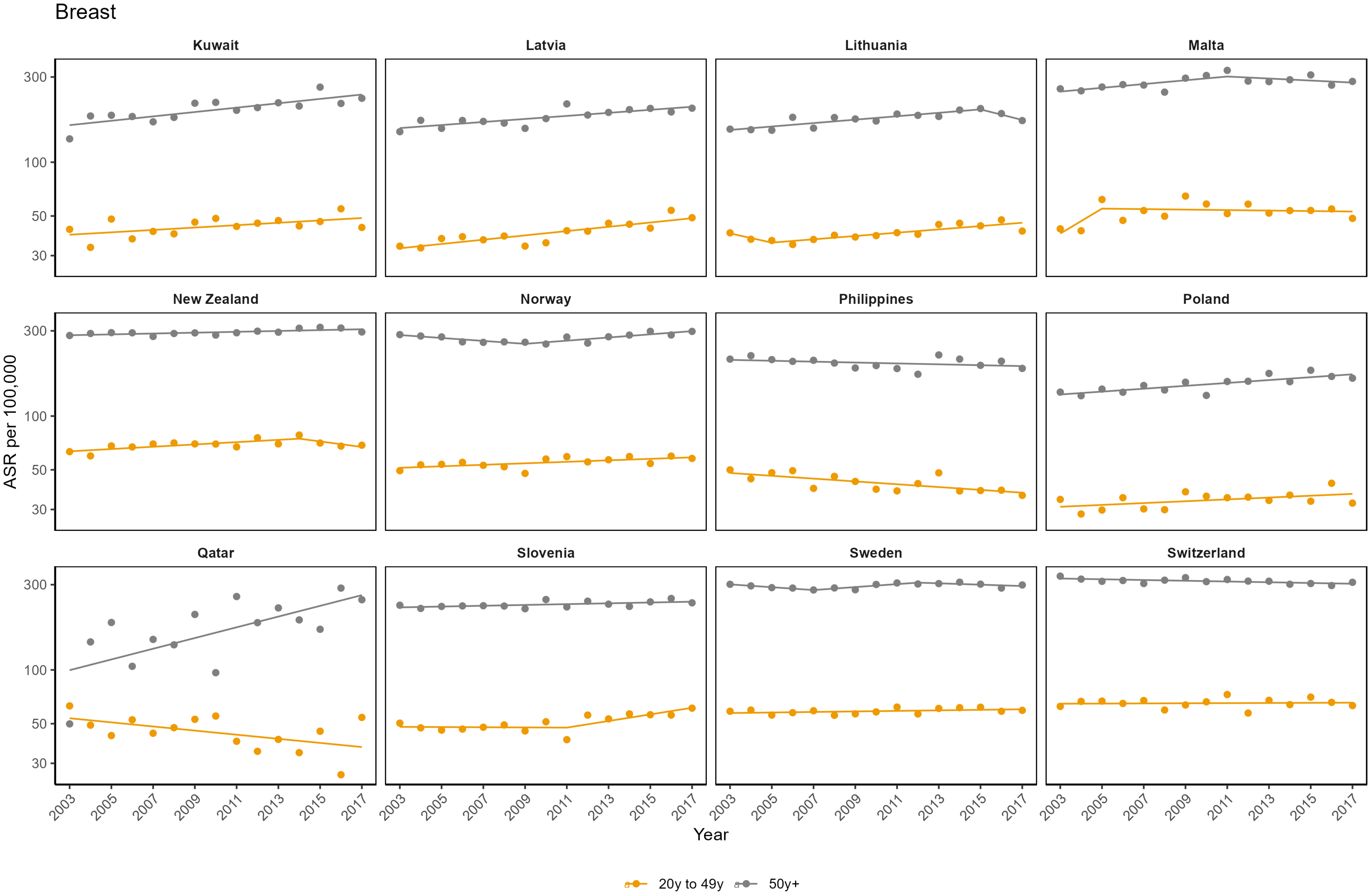

Breast
Kuwait
Latvia
Lithuania
Malta
New Zealand
Norway
Philippines
Poland
Qatar
Slovenia
Sweden
Switzerland
ASR per 100,000
300
100
50
30
2003
2005
2007
2009
2011
2013
2015
2017
Year
20y to 49y
50y+

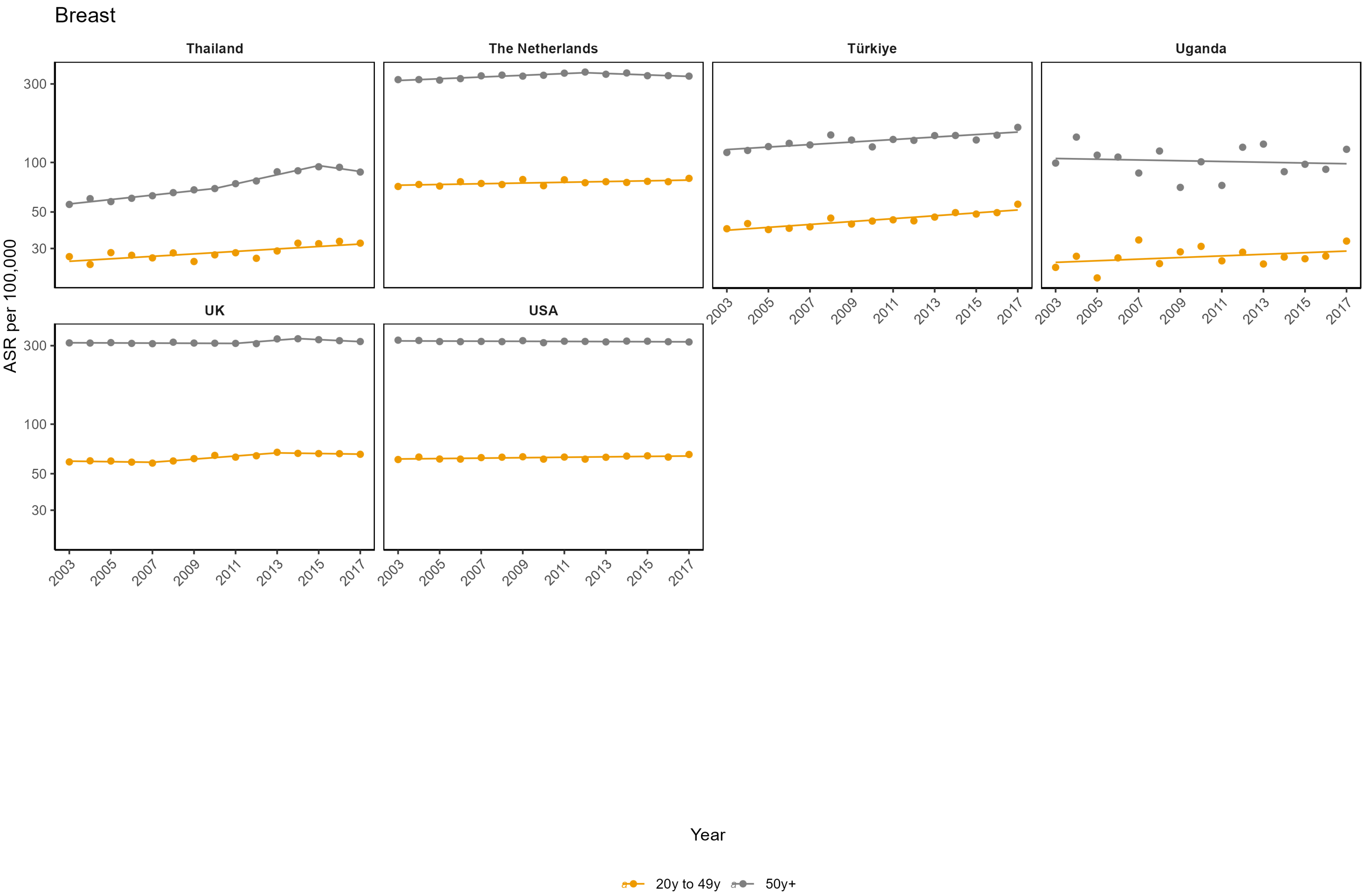
Breast
Thailand
The Netherlands
Türkiye
Uganda
UK
USA
300
100
50
30
ASR per 100,000
2003
2005
2007
2009
2011
2013
2015
2017
Year
20y to 49y
50y+

**Appendix Figure 2: Age-standardised incidence rates (ASR) per 100,000**

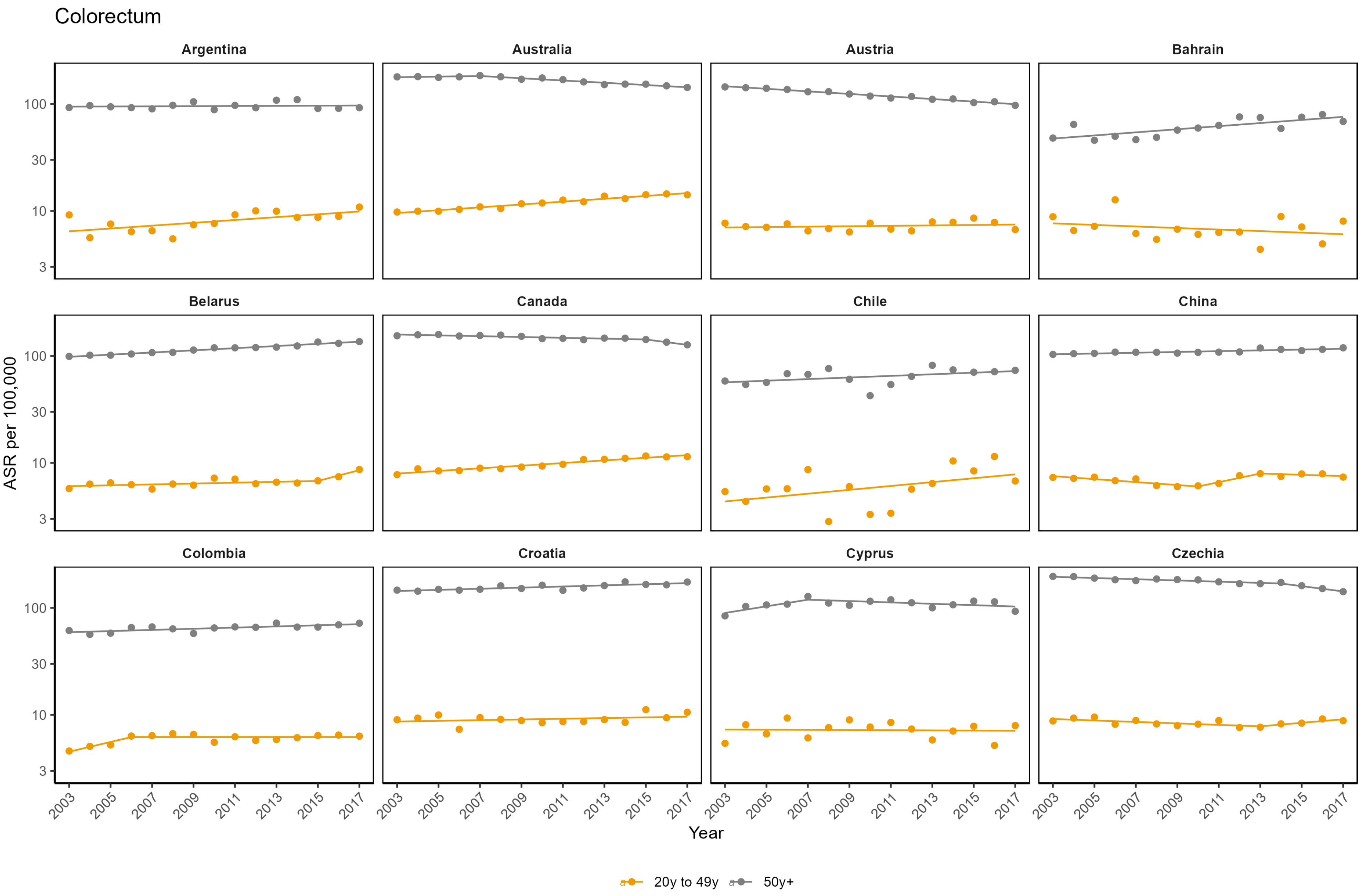

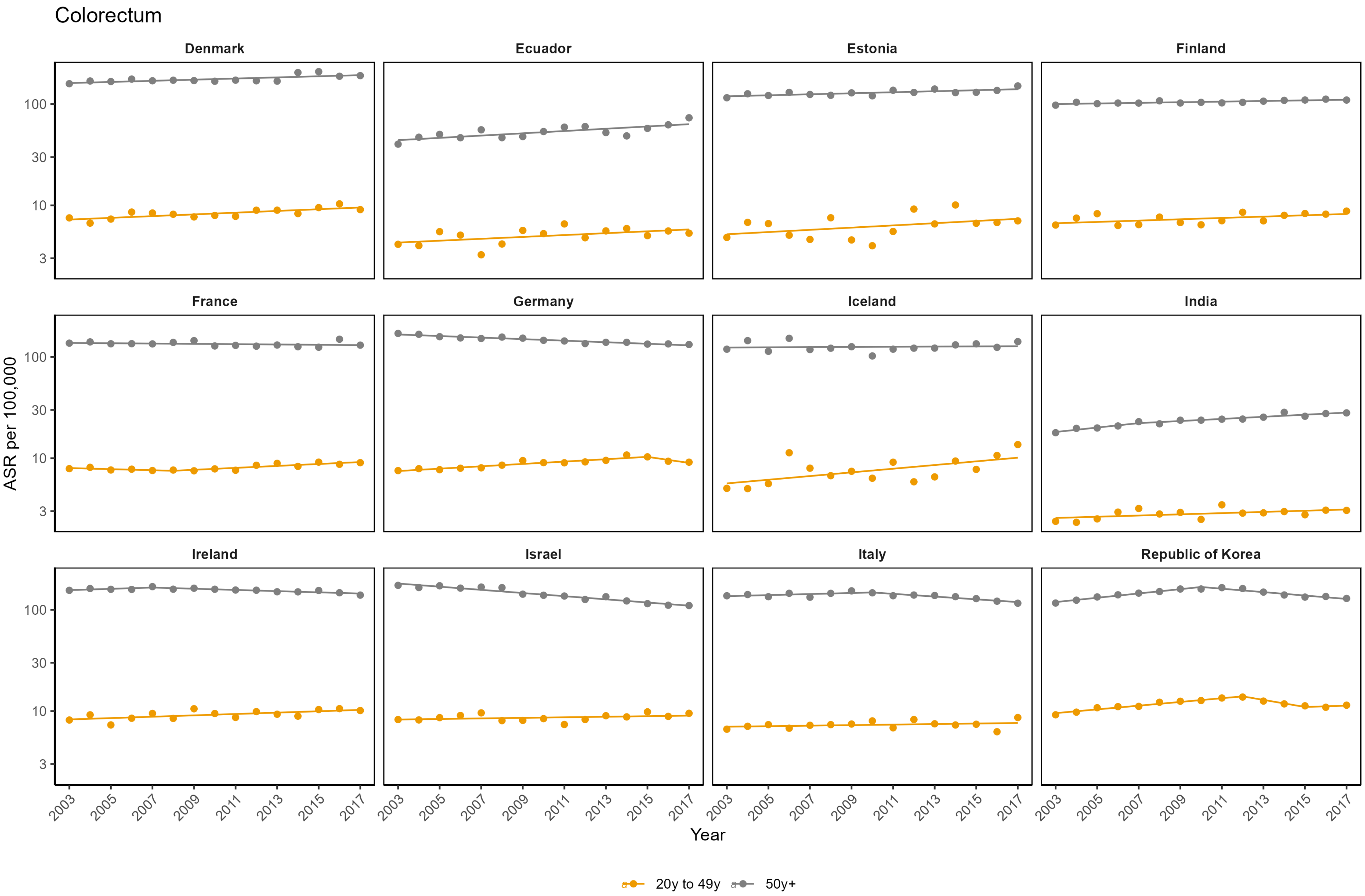
Colorectum
Denmark
Ecuador
Estonia
Finland
France
Germany
Iceland
India
Ireland
Israel
Italy
Republic of Korea
ASR per 100,000
100
30
10
3
2003
2005
2007
2009
2011
2013
2015
2017
Year
20y to 49y
50y+

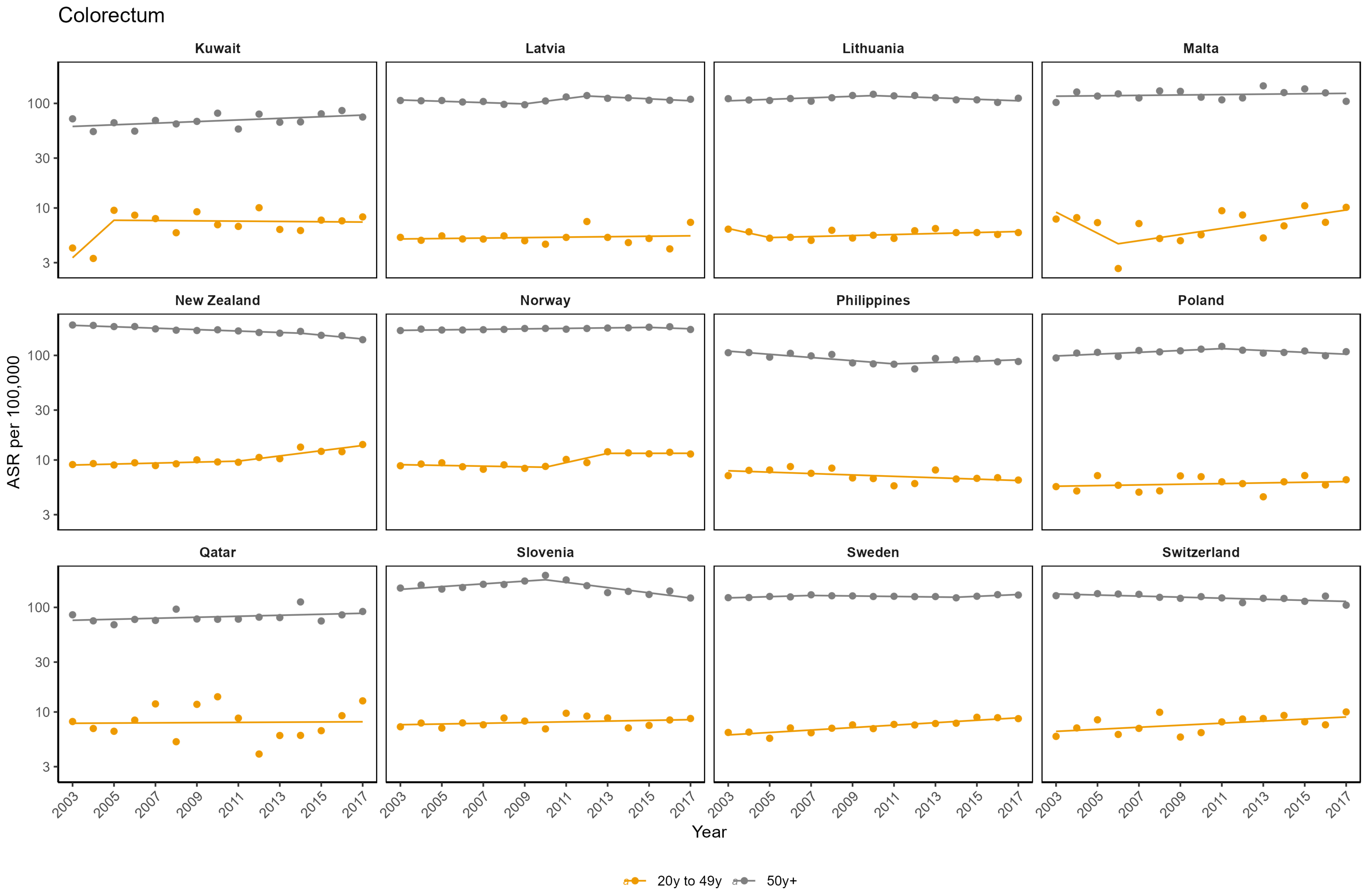

Colorectum
Kuwait
Latvia
Lithuania
Malta
New Zealand
Norway
Philippines
Poland
Qatar
Slovenia
Sweden
Switzerland
ASR per 100,000
100
30
10
3
2003
2005
2007
2009
2011
2013
2015
2017
Year
20y to 49y
50y+

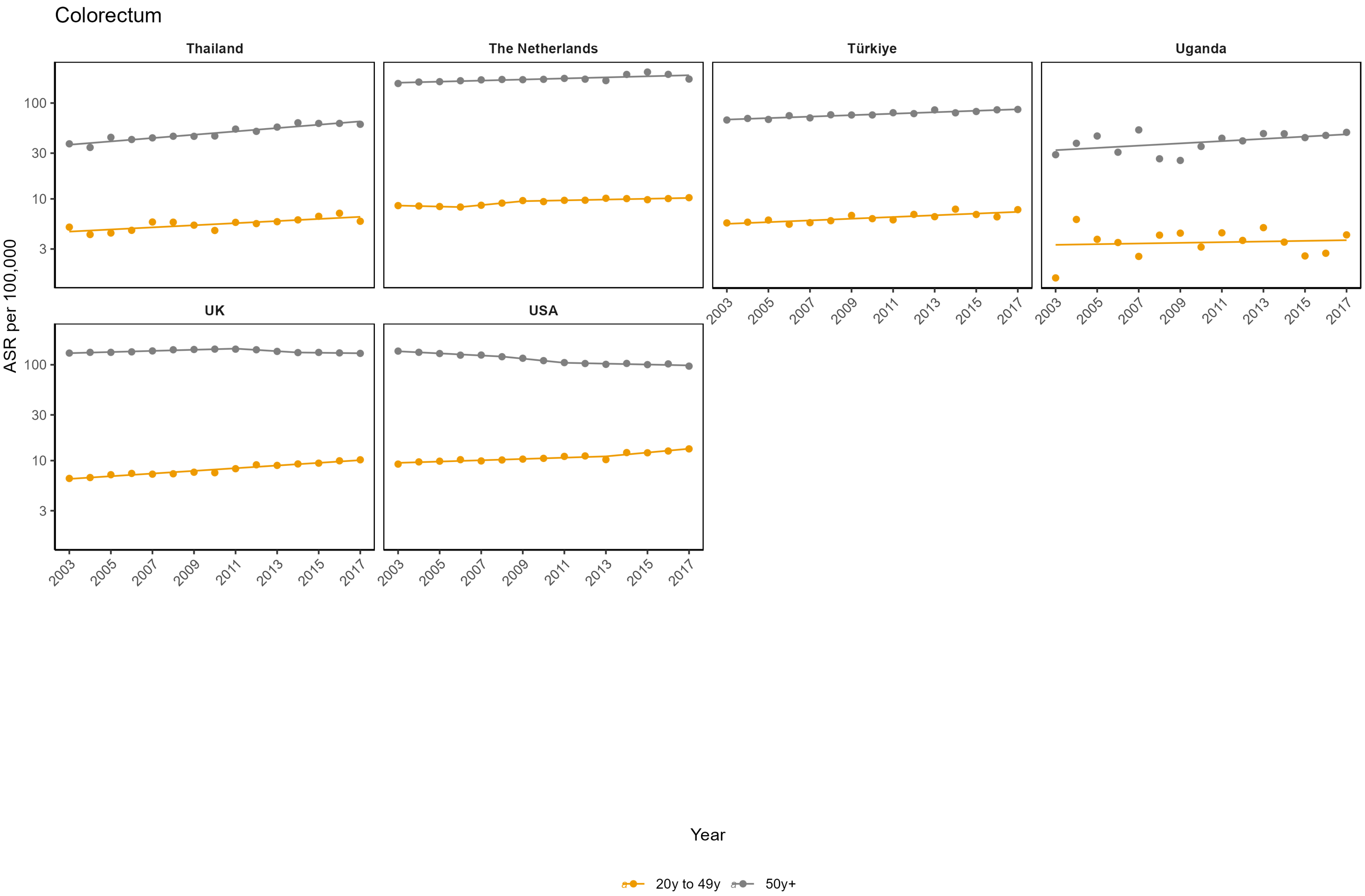

Colorectum
Thailand
The Netherlands
Türkiye
Uganda
UK
USA
ASR per 100,000
100
30
10
3
2003
2005
2007
2009
2011
2013
2015
2017
Year
20y to 49y
50y+

**Appendix Figure 3: Age-standardised incidence rates (ASR) per 100,000**

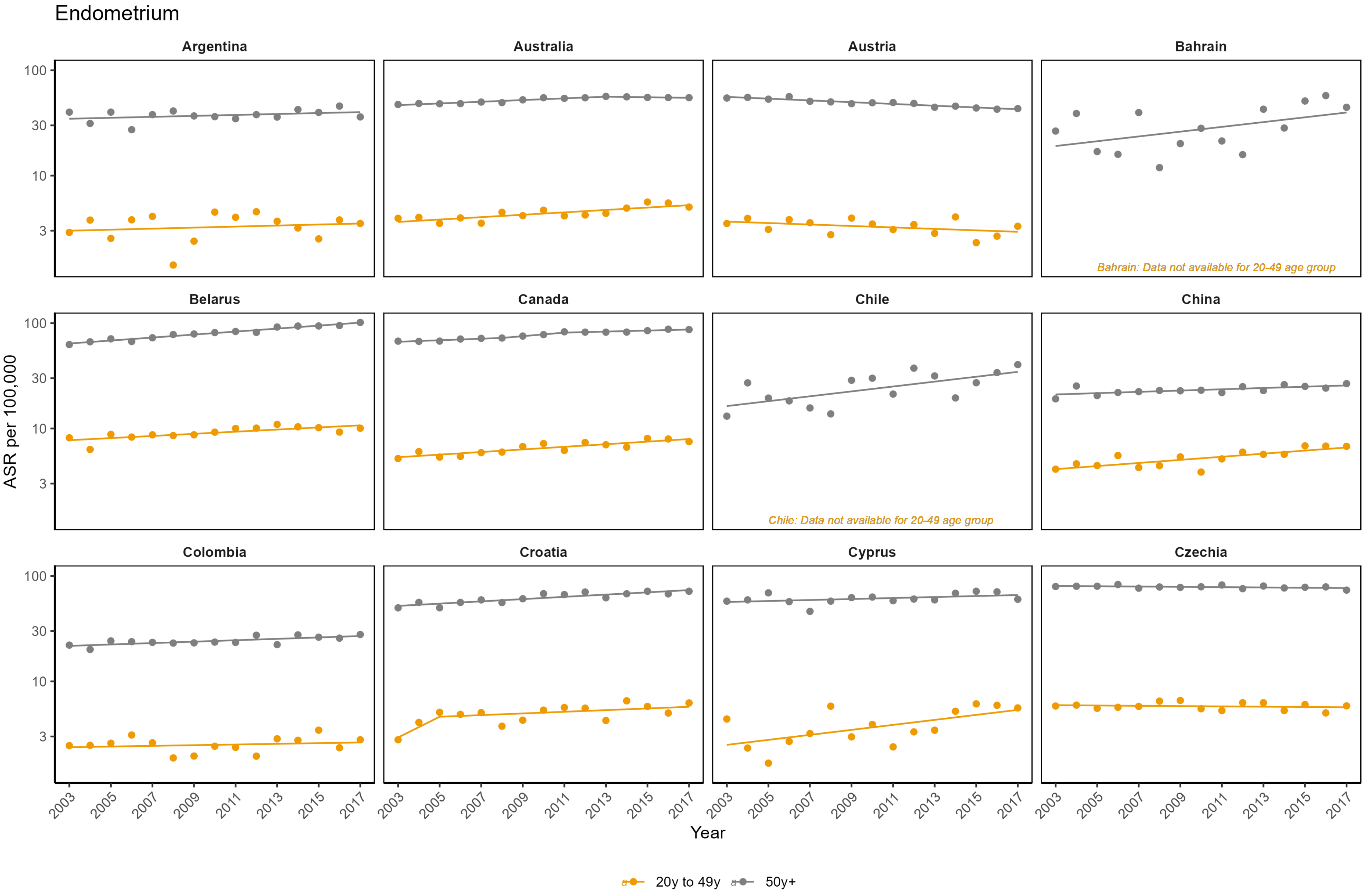

Endometrium

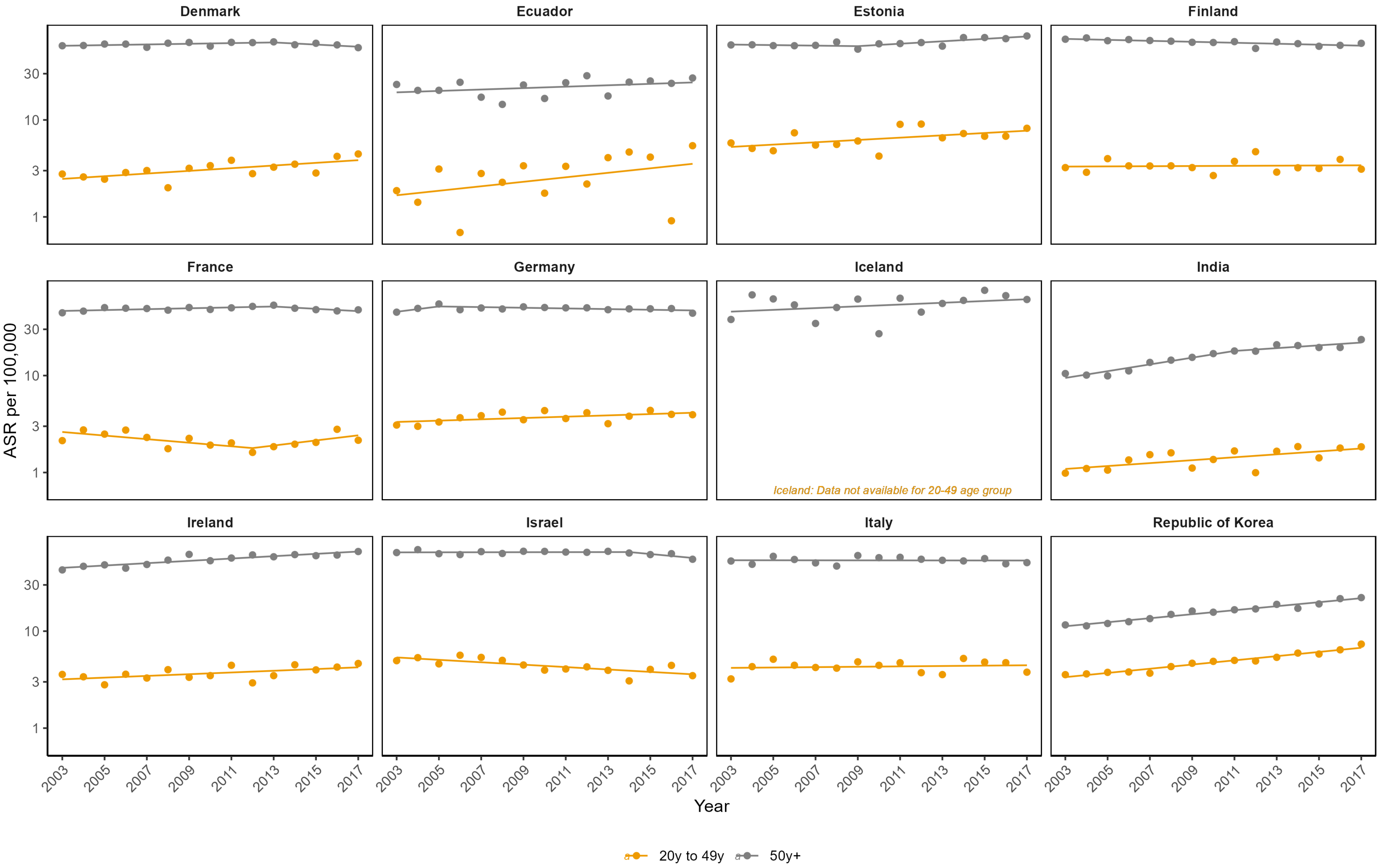

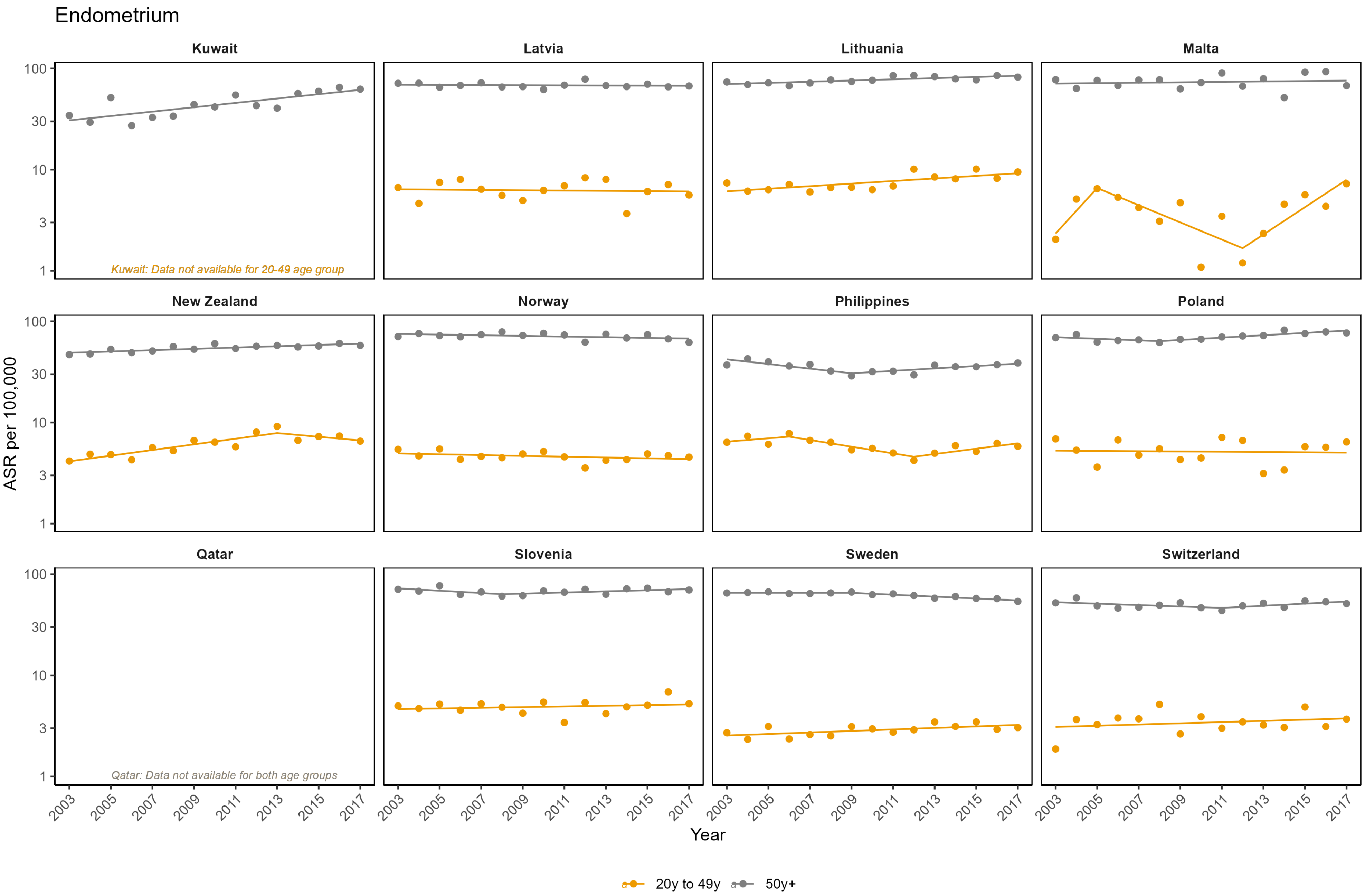

Endometrium
Kuwait
Latvia
Lithuania
Malta
New Zealand
Norway
Philippines
Poland
Qatar
Slovenia
Sweden
Switzerland
Kuwait: Data not available for 20-49 age group
Qatar: Data not available for both age groups
ASR per 100,000
100
30
10
3
1
2003
2005
2007
2009
2011
2013
2015
2017
Year
20y to 49y
50y+

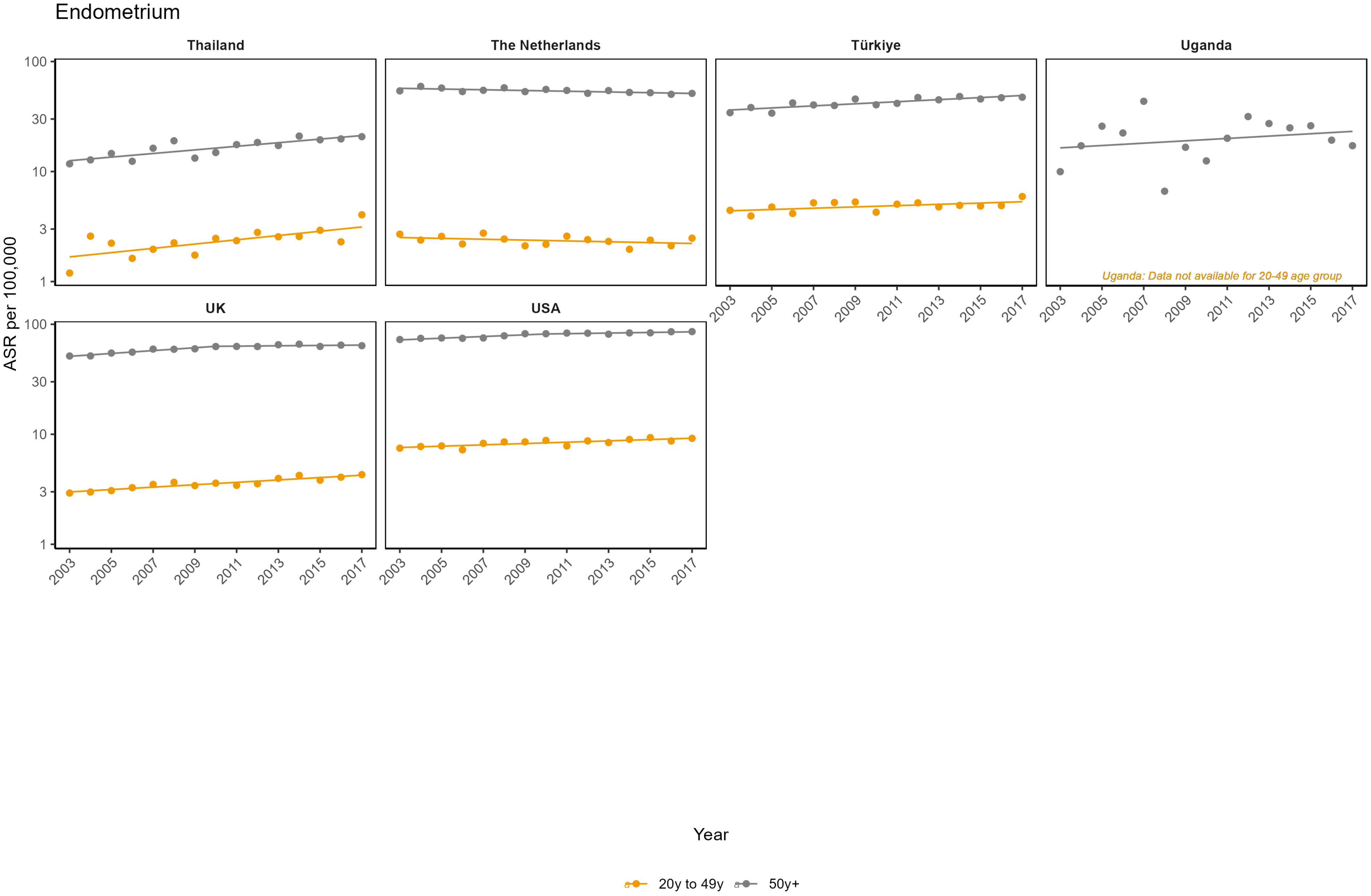
Endometrium
Thailand
The Netherlands
Türkiye
Uganda
UK
USA
Uganda: Data not available for 20-49 age group
ASR per 100,000
100
30
10
3
1
2003
2005
2007
2009
2011
2013
2015
2017
Year
20y to 49y
50y+

**Appendix Figure 4: Age-standardised incidence rates (ASR) per 100,000**

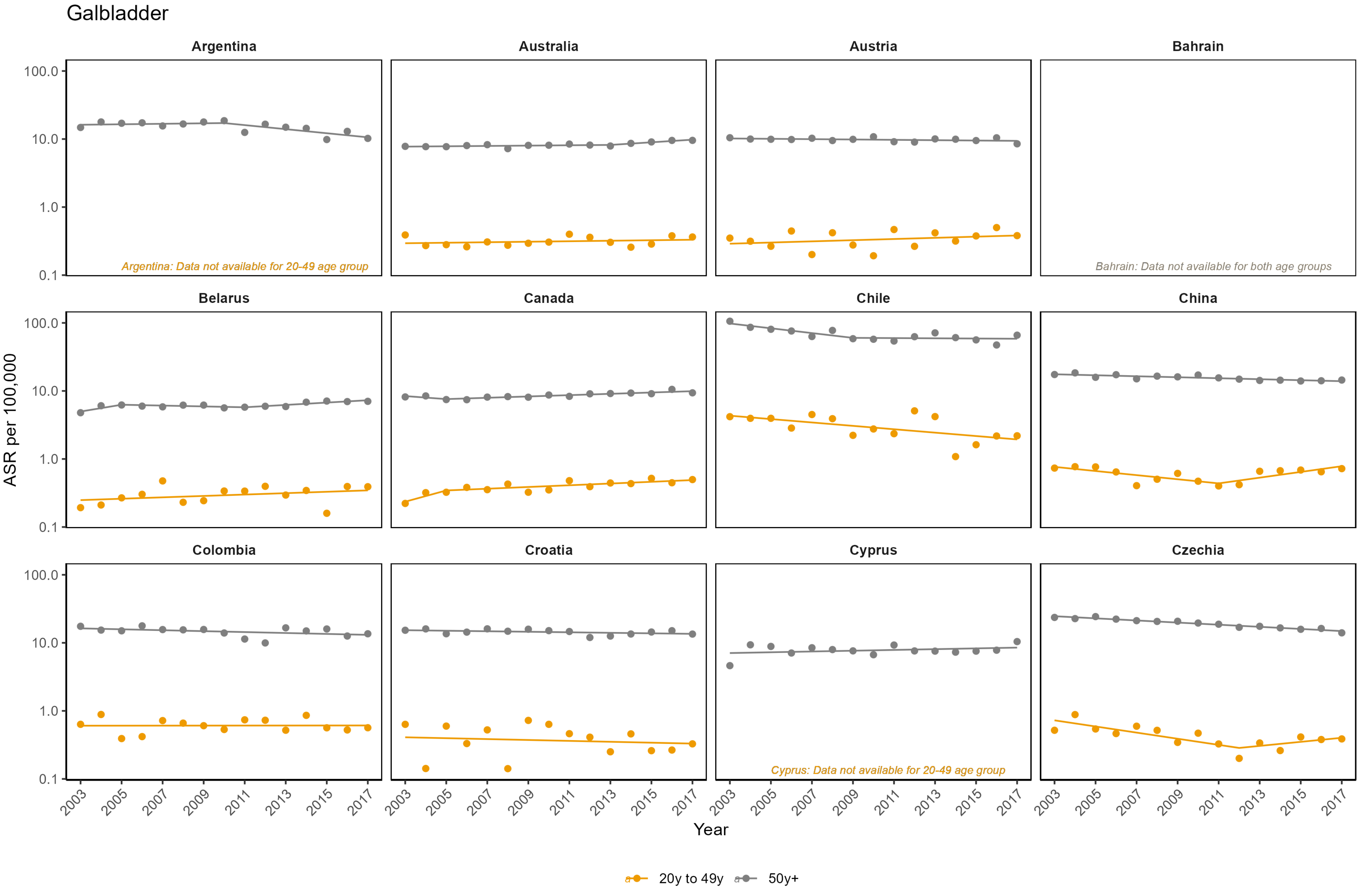

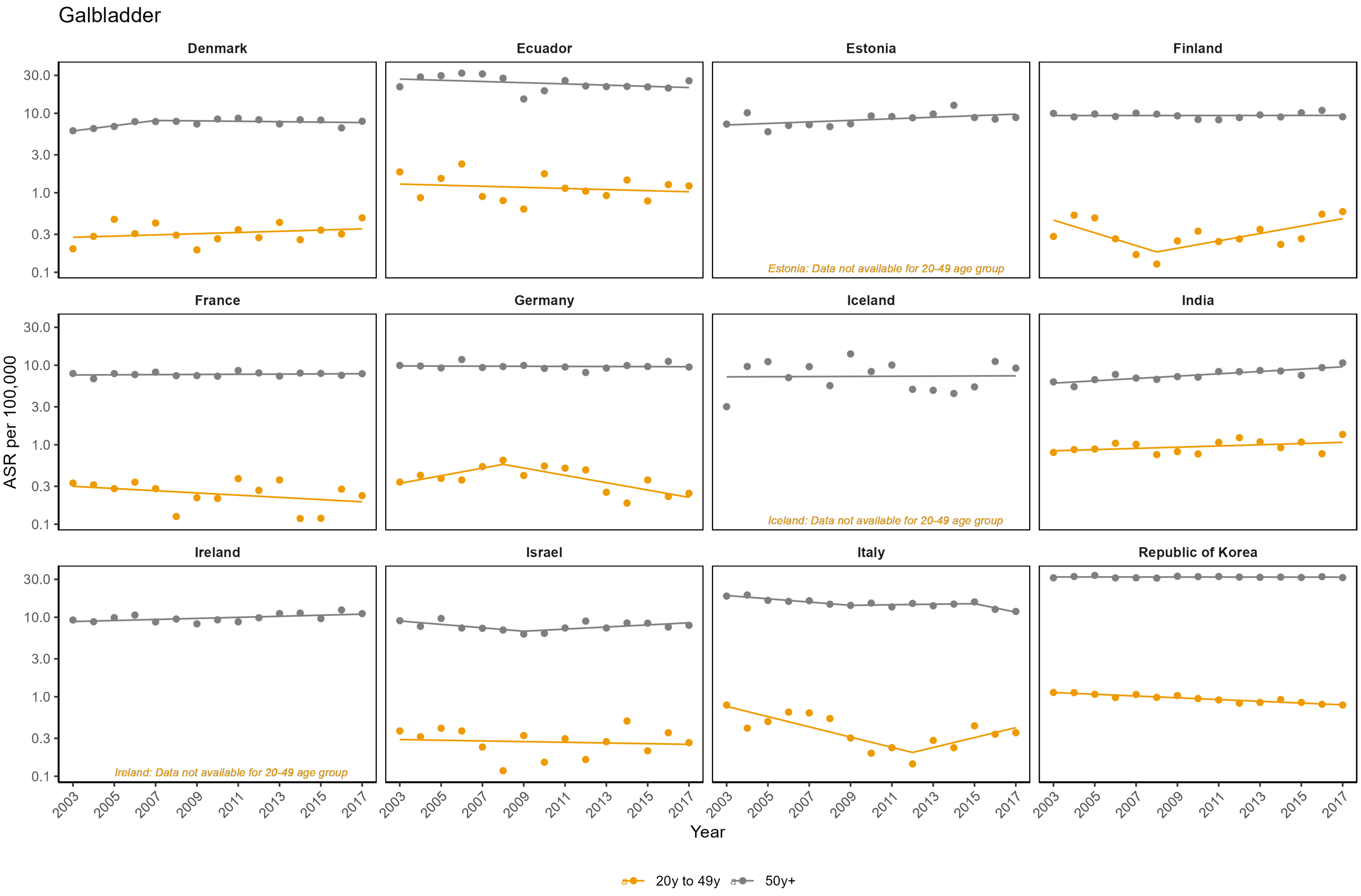

Galbladder
Denmark
Ecuador
Estonia
Finland
France
Germany
Iceland
India
Ireland
Israel
Italy
Republic of Korea
Estonia: Data not available for 20-49 age group
Iceland: Data not available for 20-49 age group
Ireland: Data not available for 20-49 age group
ASR per 100,000
Year
20y to 49y
50y+

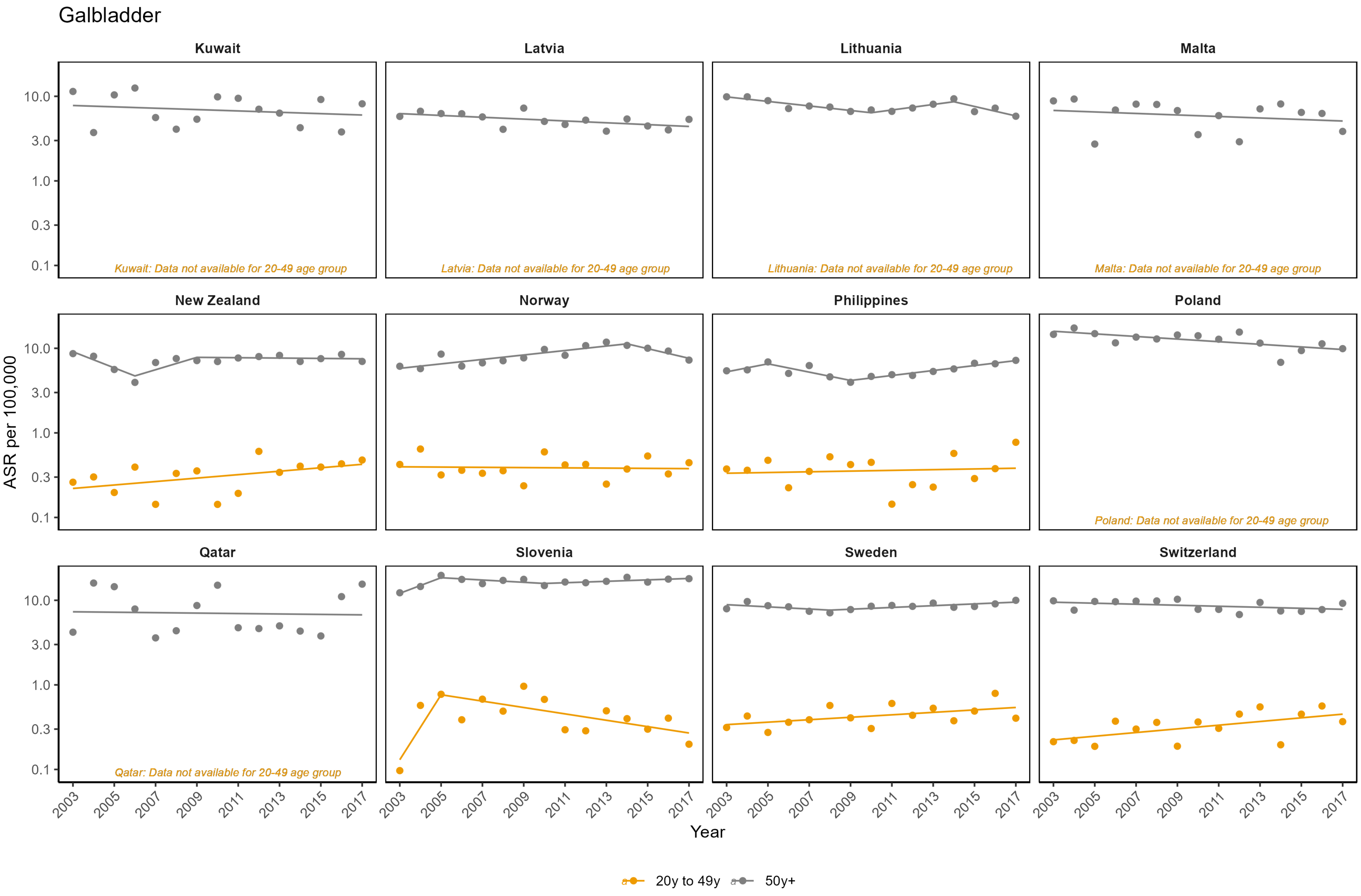
Galbladder
Kuwait
Latvia
Lithuania
Malta
New Zealand
Norway
Philippines
Poland
Qatar
Slovenia
Sweden
Switzerland
Kuwait: Data not available for 20-49 age group
Latvia: Data not available for 20-49 age group
Lithuania: Data not available for 20-49 age group
Malta: Data not available for 20-49 age group
Poland: Data not available for 20-49 age group
Qatar: Data not available for 20-49 age group
ASR per 100,000
10.0
3.0
1.0
0.3
0.1
2003
2005
2007
2009
2011
2013
2015
2017
Year
20y to 49y
50y+

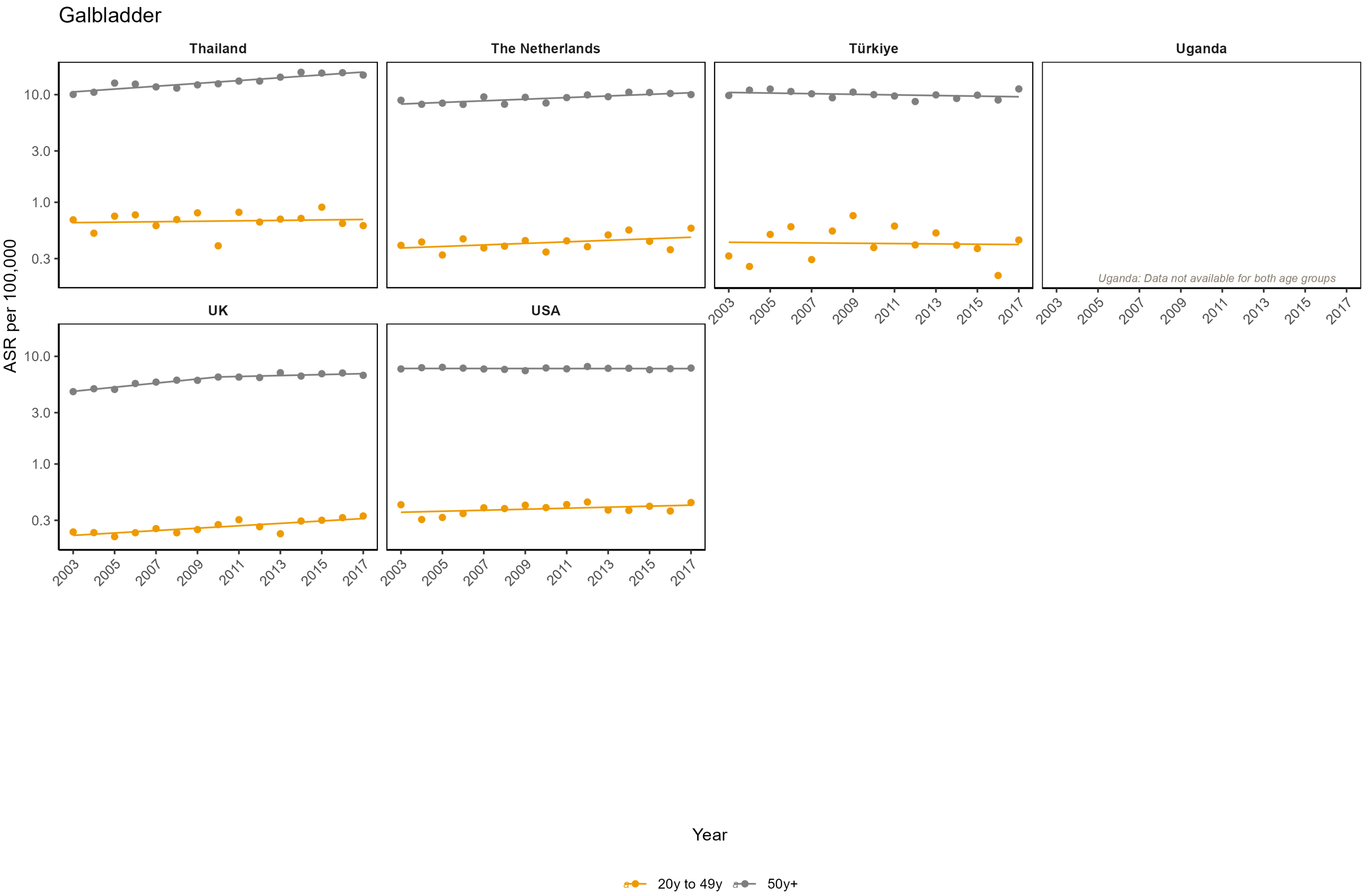

Galbladder
Thailand
The Netherlands
Türkiye
Uganda
UK
USA
Uganda: Data not available for both age groups
ASR per 100,000
10.0
3.0
1.0
0.3
2003
2005
2007
2009
2011
2013
2015
2017
Year
20y to 49y
50y+

**Appendix Figure 5: Age-standardised incidence rates (ASR) per 100,000** [illegible]

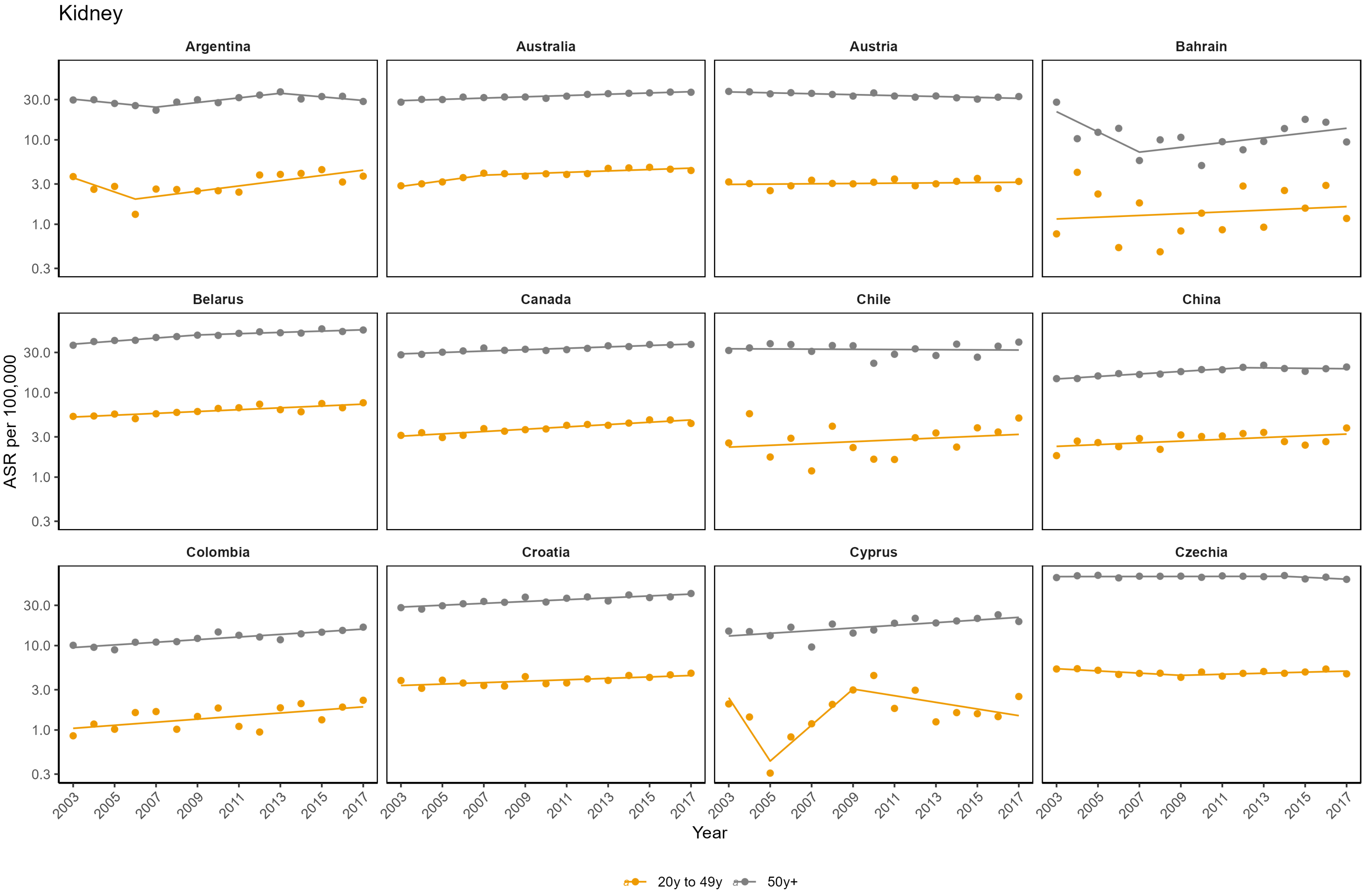

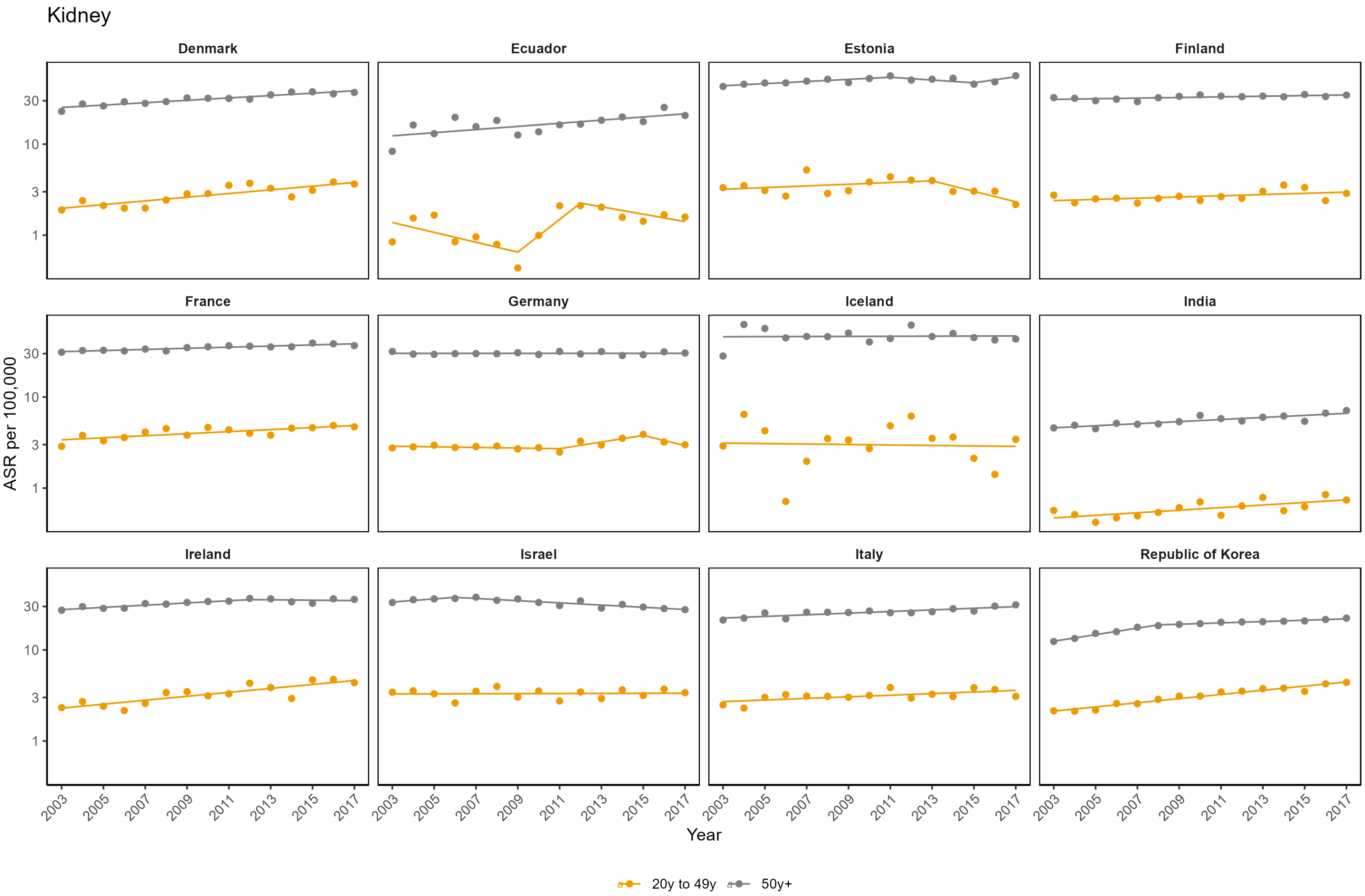
Kidney
Denmark
Ecuador
Estonia
Finland
France
Germany
Iceland
India
Ireland
Israel
Italy
Republic of Korea
ASR per 100,000
30
10
3
1
2003
2005
2007
2009
2011
2013
2015
2017
Year
20y to 49y
50y+

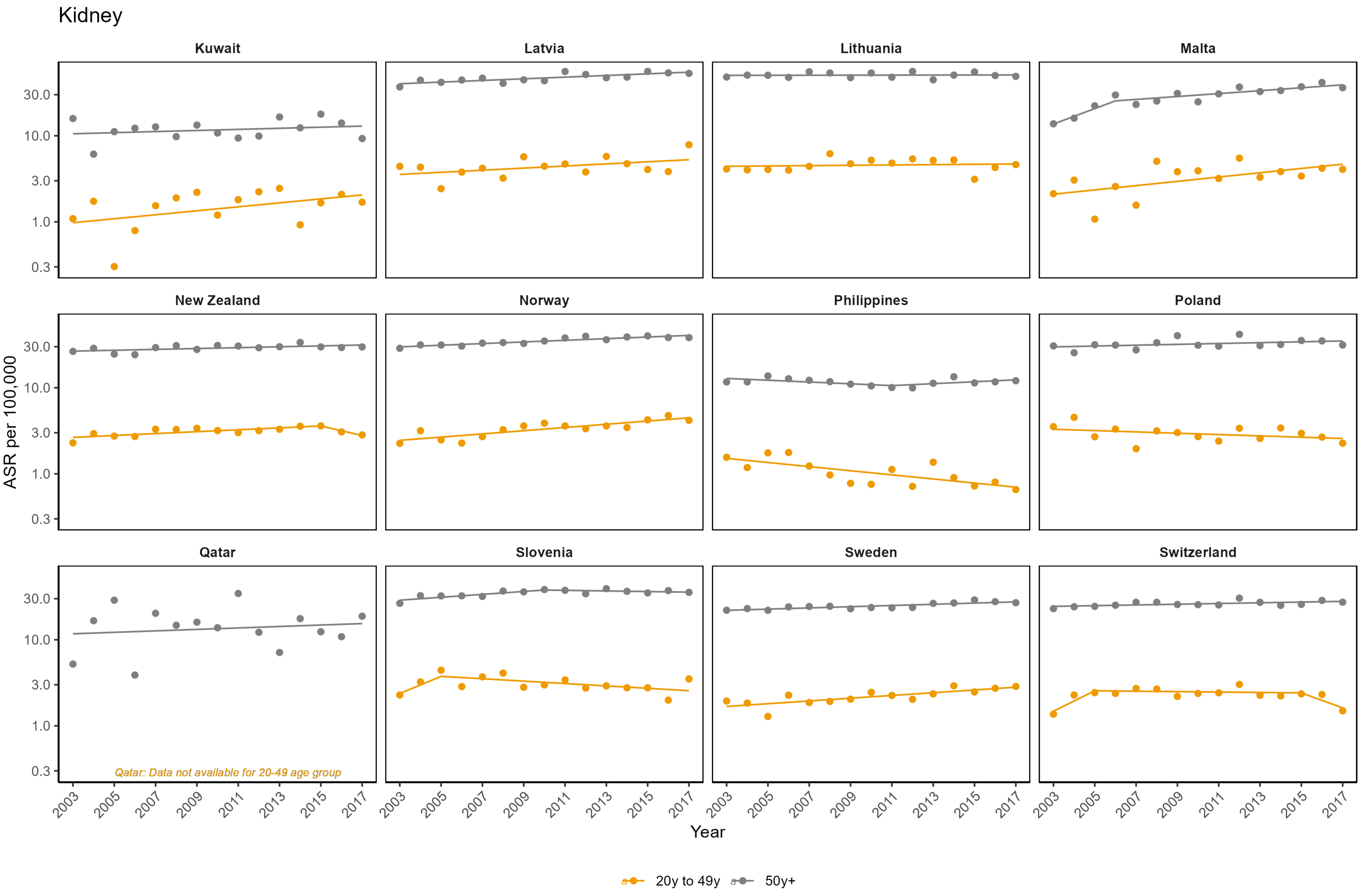

Kidney
Kuwait
Latvia
Lithuania
Malta
New Zealand
Norway
Philippines
Poland
Qatar
Slovenia
Sweden
Switzerland
ASR per 100,000
30.0
10.0
3.0
1.0
0.3
Qatar: Data not available for 20-49 age group
2003
2005
2007
2009
2011
2013
2015
2017
Year
20y to 49y
50y+

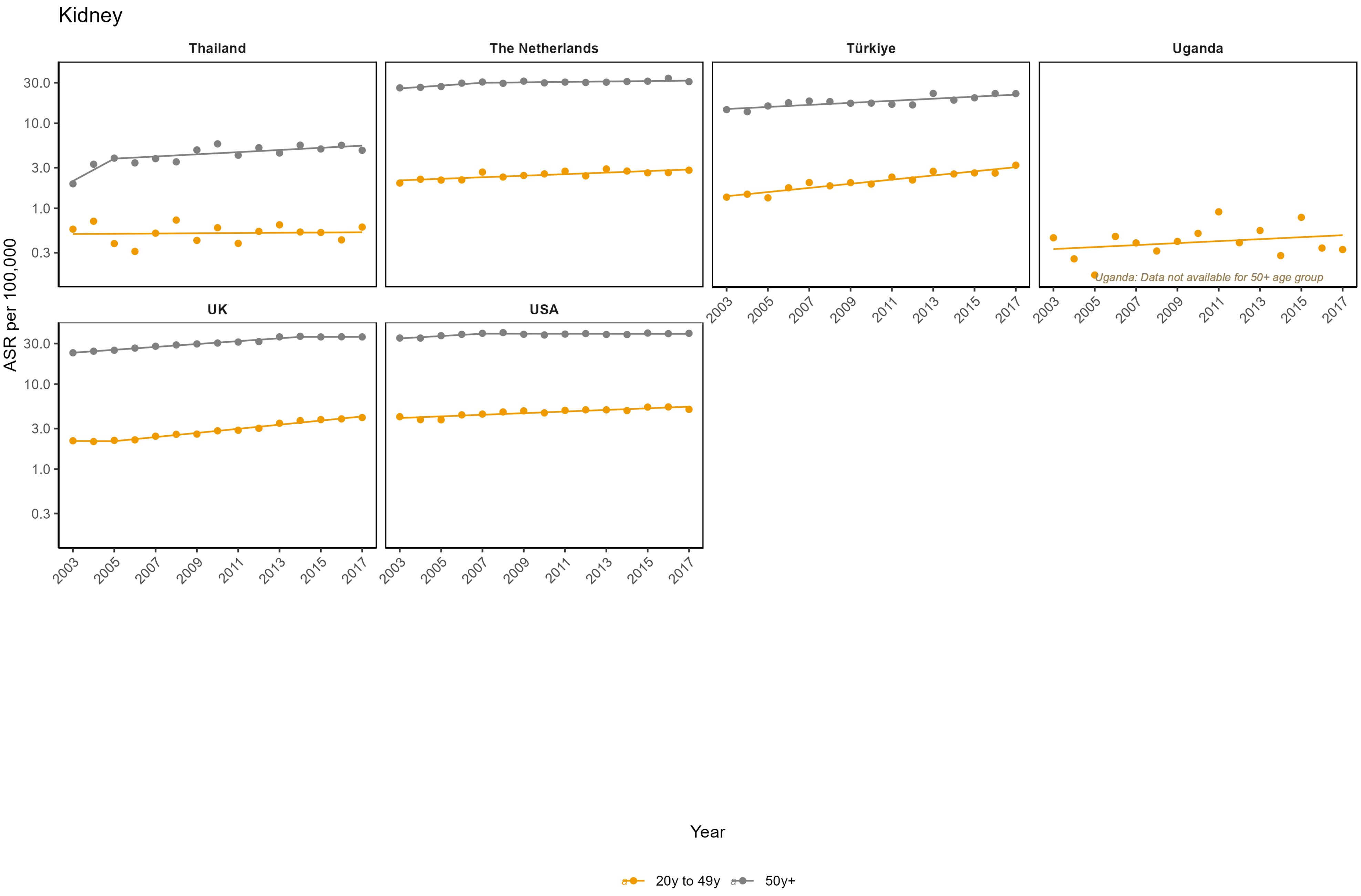
Kidney
Thailand
The Netherlands
Türkiye
Uganda
UK
USA
ASR per 100,000
30.0
10.0
3.0
1.0
0.3
2003
2005
2007
2009
2011
2013
2015
2017
Uganda: Data not available for 50+ age group
Year
20y to 49y
50y+

**Appendix Figure 6: Age-standardised incidence rates (ASR) per 100,000**

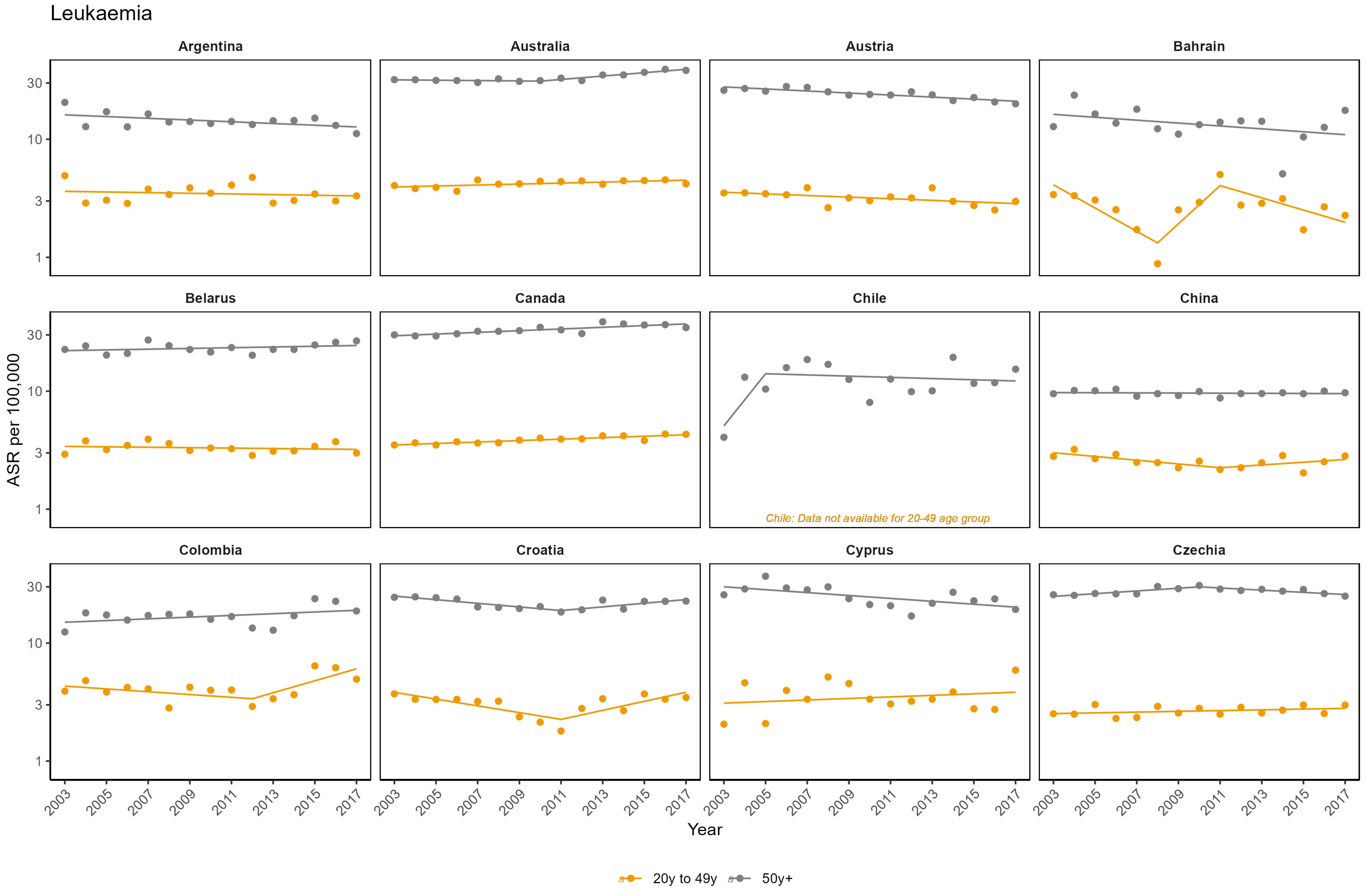

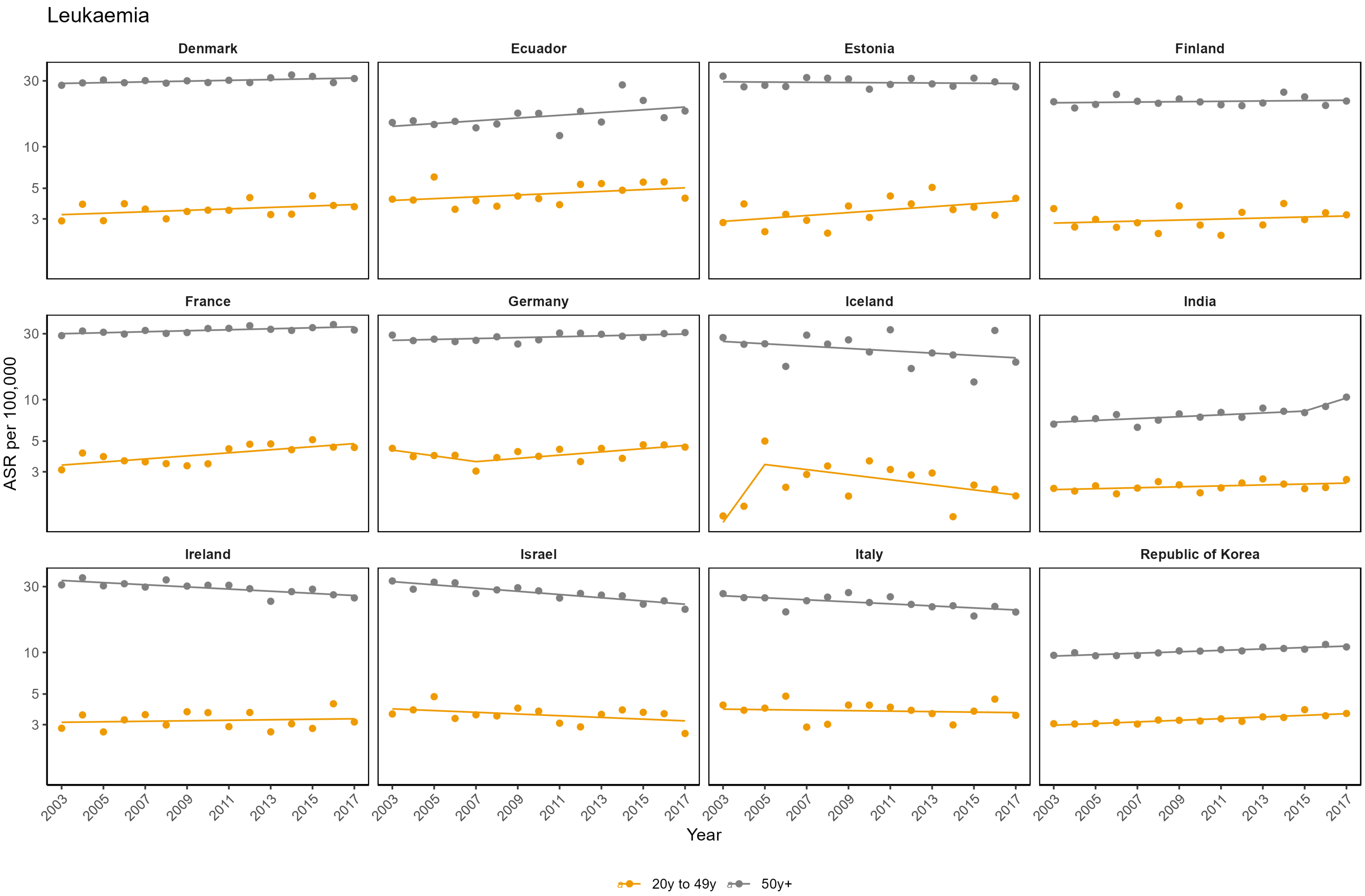
Leukaemia
Denmark
Ecuador
Estonia
Finland
France
Germany
Iceland
India
Ireland
Israel
Italy
Republic of Korea
ASR per 100,000
30
10
5
3
2003
2005
2007
2009
2011
2013
2015
2017
Year
20y to 49y
50y+

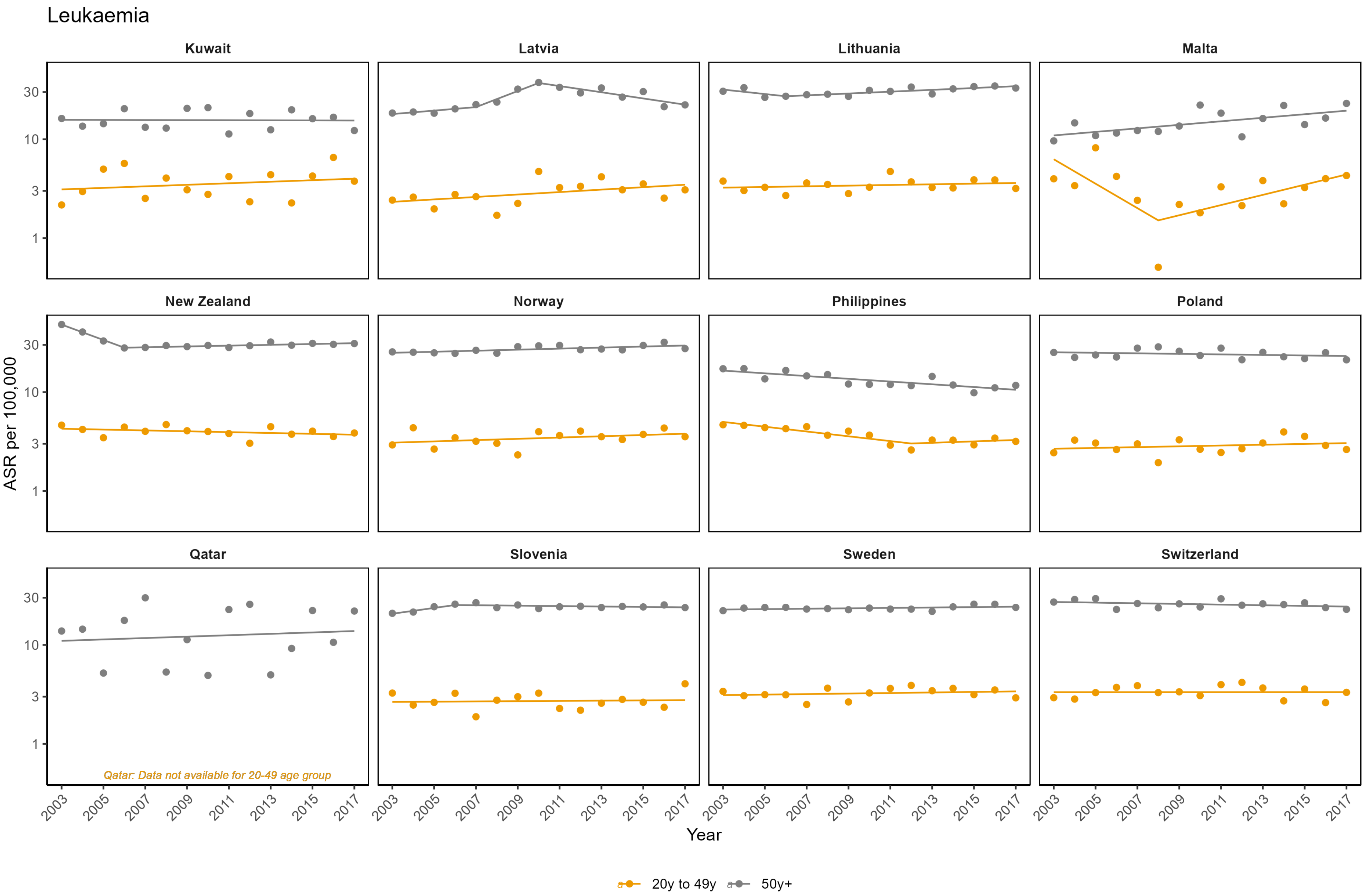

Leukaemia
Kuwait
Latvia
Lithuania
Malta
New Zealand
Norway
Philippines
Poland
Qatar
Slovenia
Sweden
Switzerland
ASR per 100,000
30
10
3
1
Qatar: Data not available for 20-49 age group
2003
2005
2007
2009
2011
2013
2015
2017
Year
20y to 49y
50y+

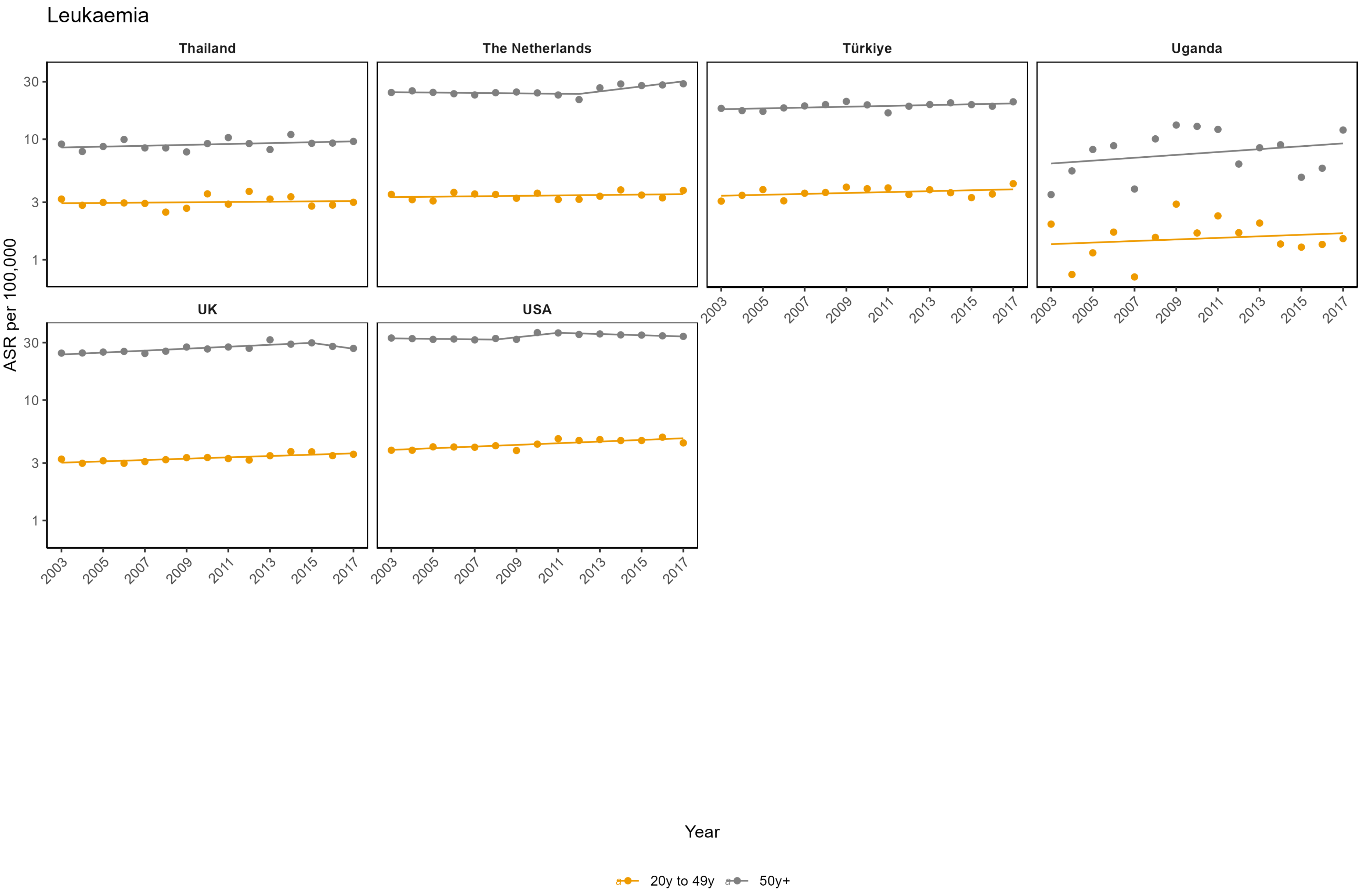

Leukaemia
Thailand
The Netherlands
Türkiye
Uganda
UK
USA
ASR per 100,000
30
10
3
1
2003
2005
2007
2009
2011
2013
2015
2017
Year
20y to 49y
50y+

**Appendix Figure 7: Age-standardised incidence rates (ASR) per 100,000**

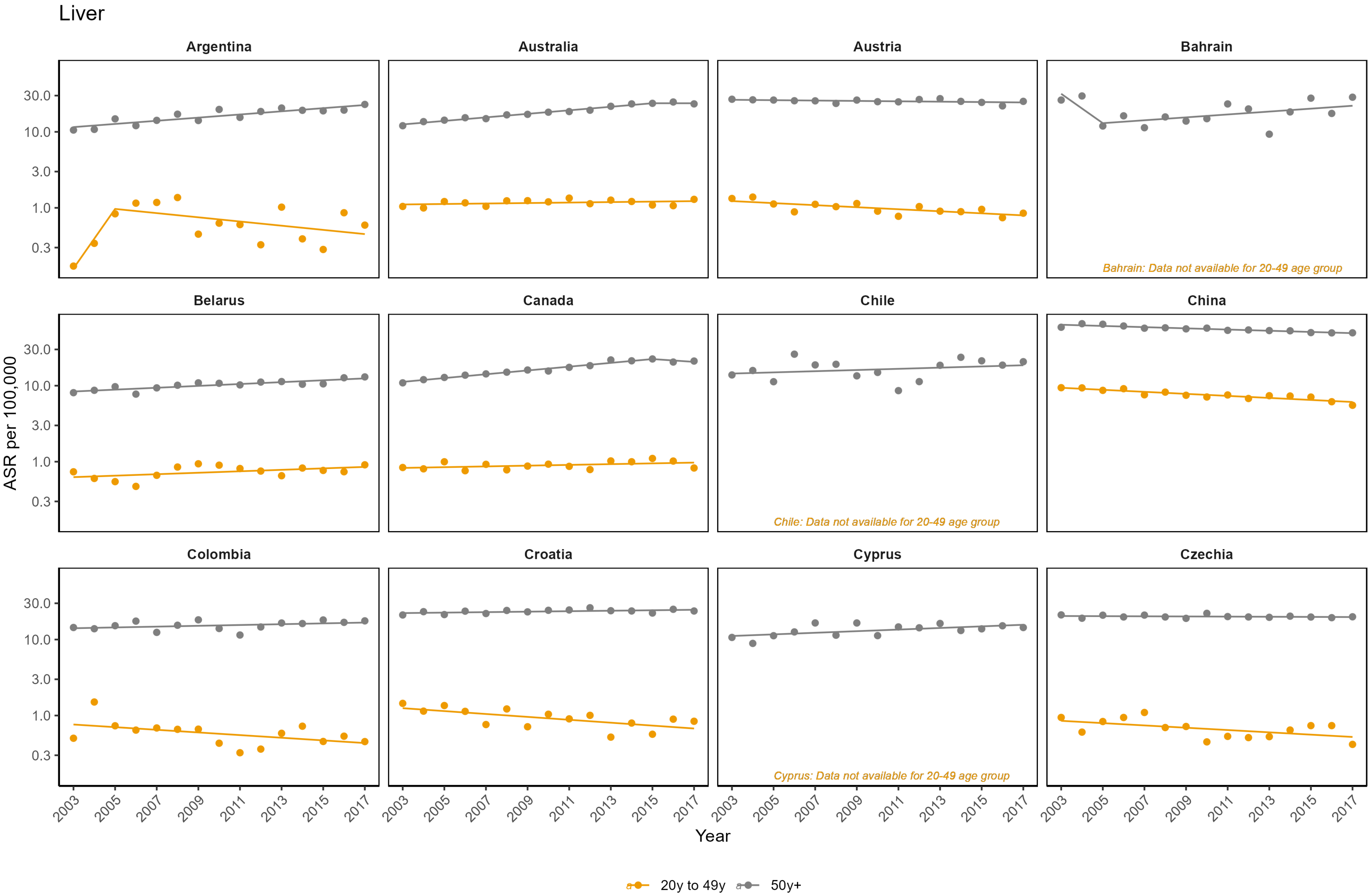

Liver

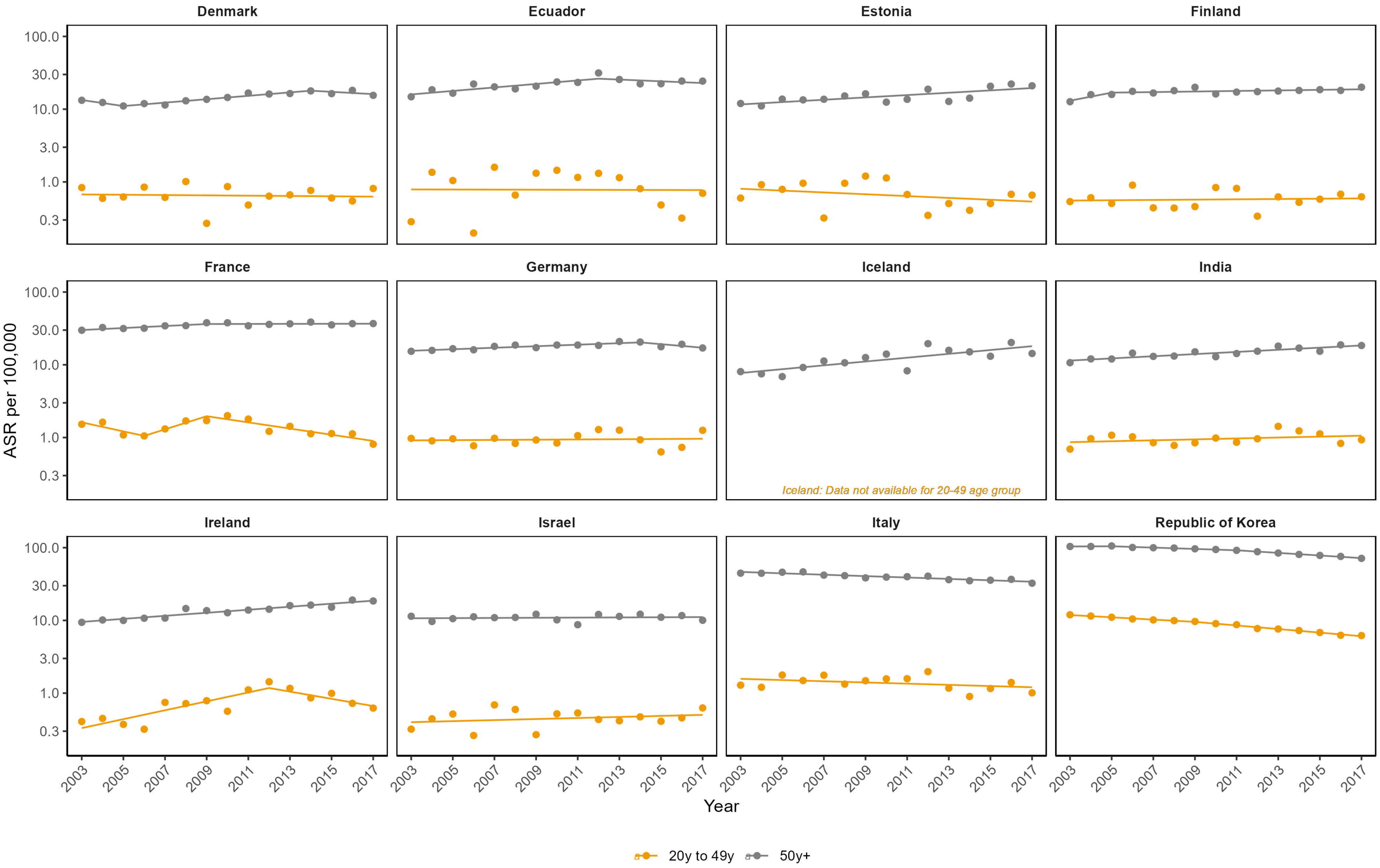

Denmark
Ecuador
Estonia
Finland
France
Germany
Iceland
India
Ireland
Israel
Italy
Republic of Korea
Iceland: Data not available for 20-49 age group
ASR per 100,000
100.0
30.0
10.0
3.0
1.0
0.3
2003
2005
2007
2009
2011
2013
2015
2017
Year
20y to 49y
50y+

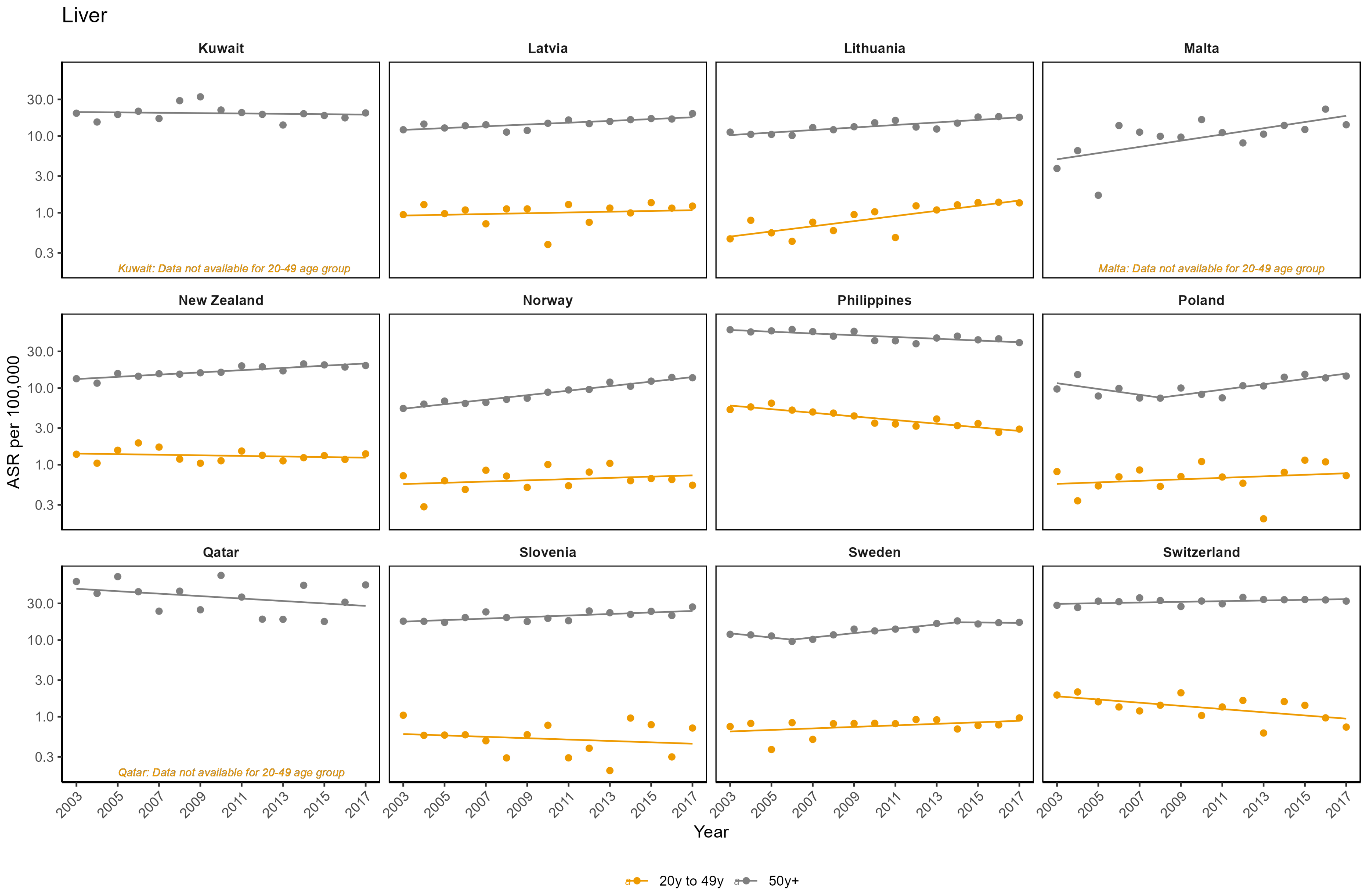

Liver
Kuwait
Latvia
Lithuania
Malta
New Zealand
Norway
Philippines
Poland
Qatar
Slovenia
Sweden
Switzerland
Kuwait: Data not available for 20-49 age group
Malta: Data not available for 20-49 age group
Qatar: Data not available for 20-49 age group
30.0
10.0
3.0
1.0
0.3
ASR per 100,000
2003
2005
2007
2009
2011
2013
2015
2017
Year
20y to 49y
50y+

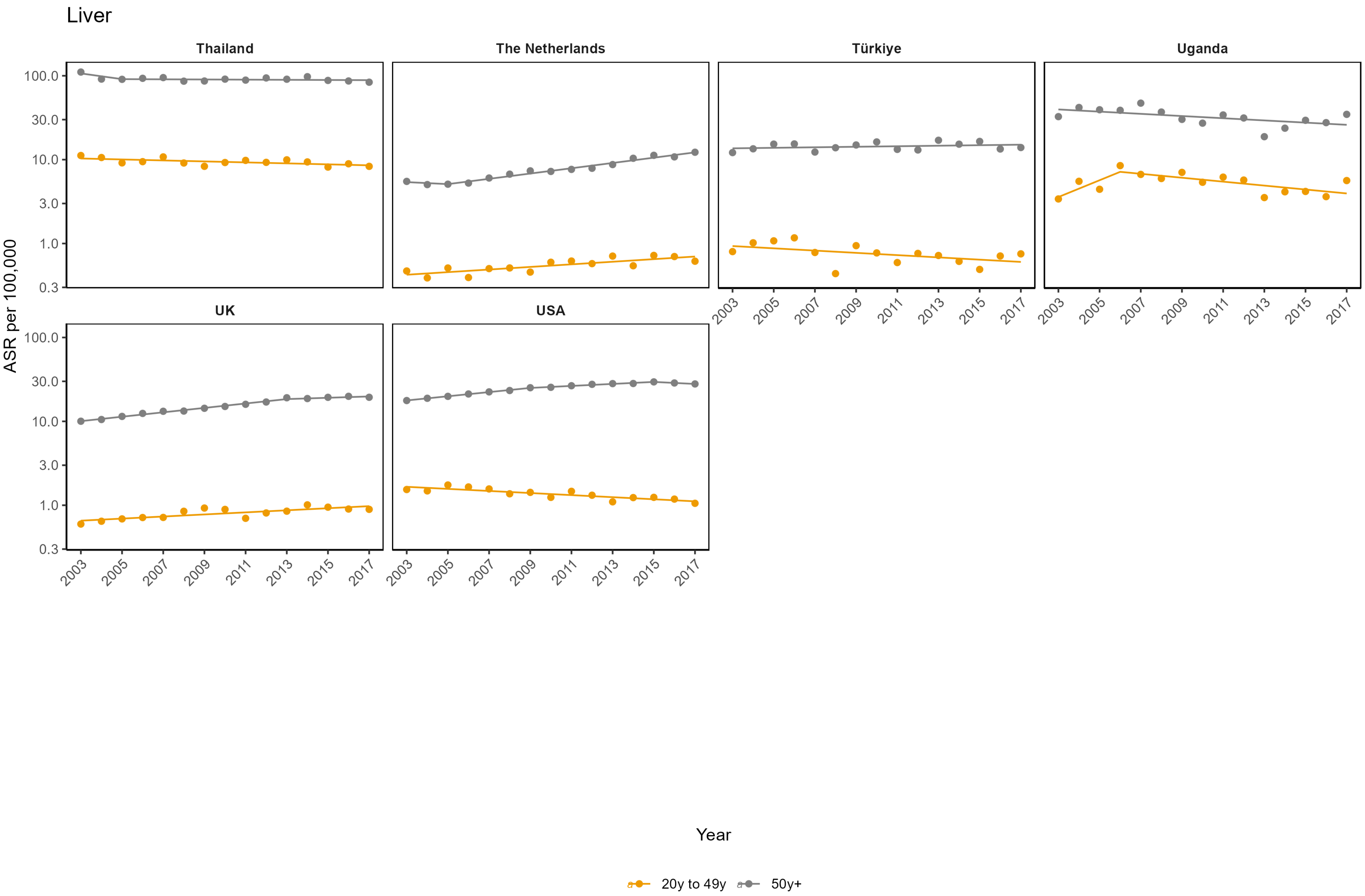

Liver
Thailand
The Netherlands
Türkiye
Uganda
UK
USA
100.0
30.0
10.0
3.0
1.0
0.3
ASR per 100,000
2003
2005
2007
2009
2011
2013
2015
2017
Year
20y to 49y
50y+

**Appendix Figure 8: Age-standardised incidence rates (ASR) per 100,000**

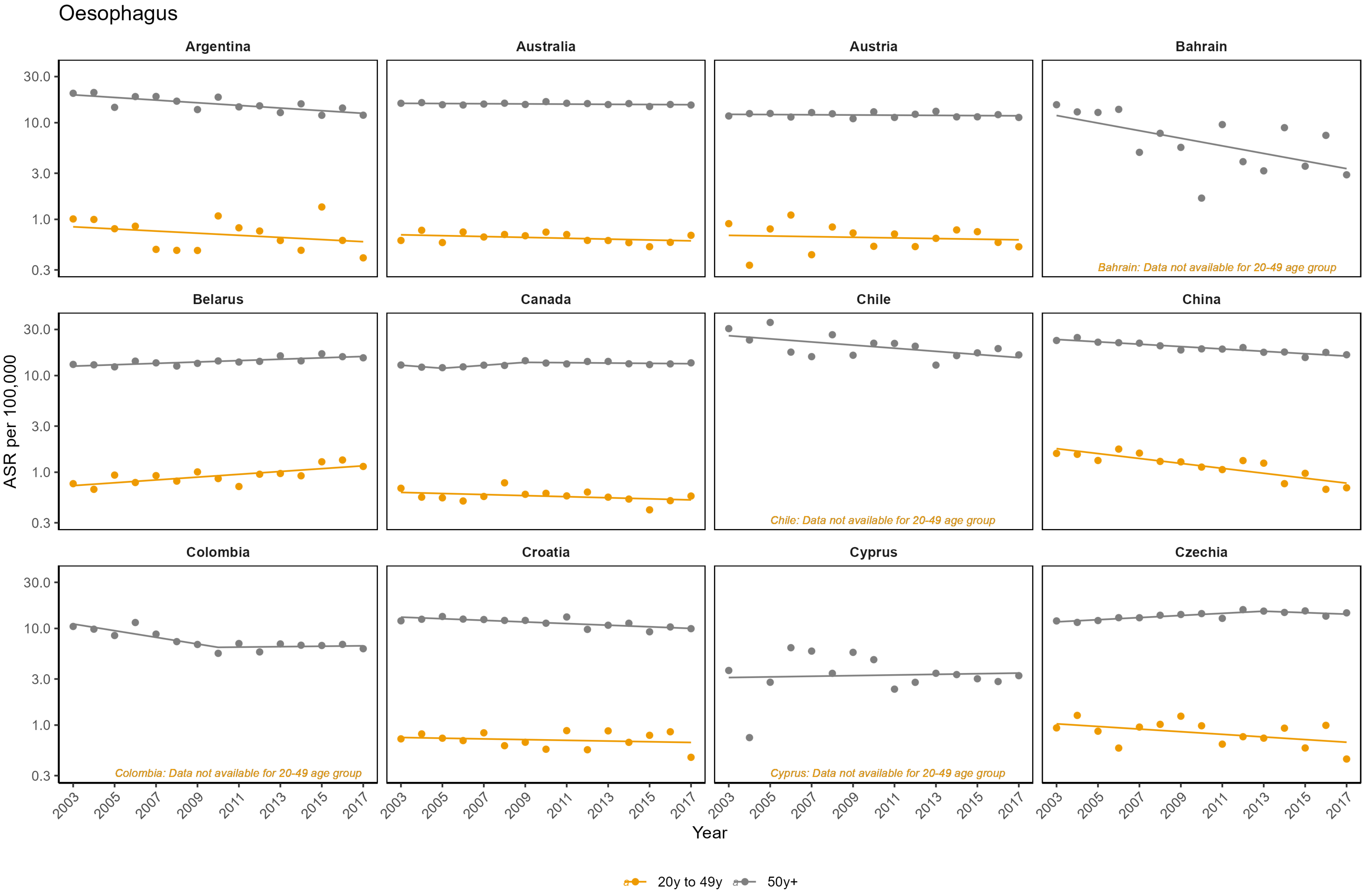

Oesophagus

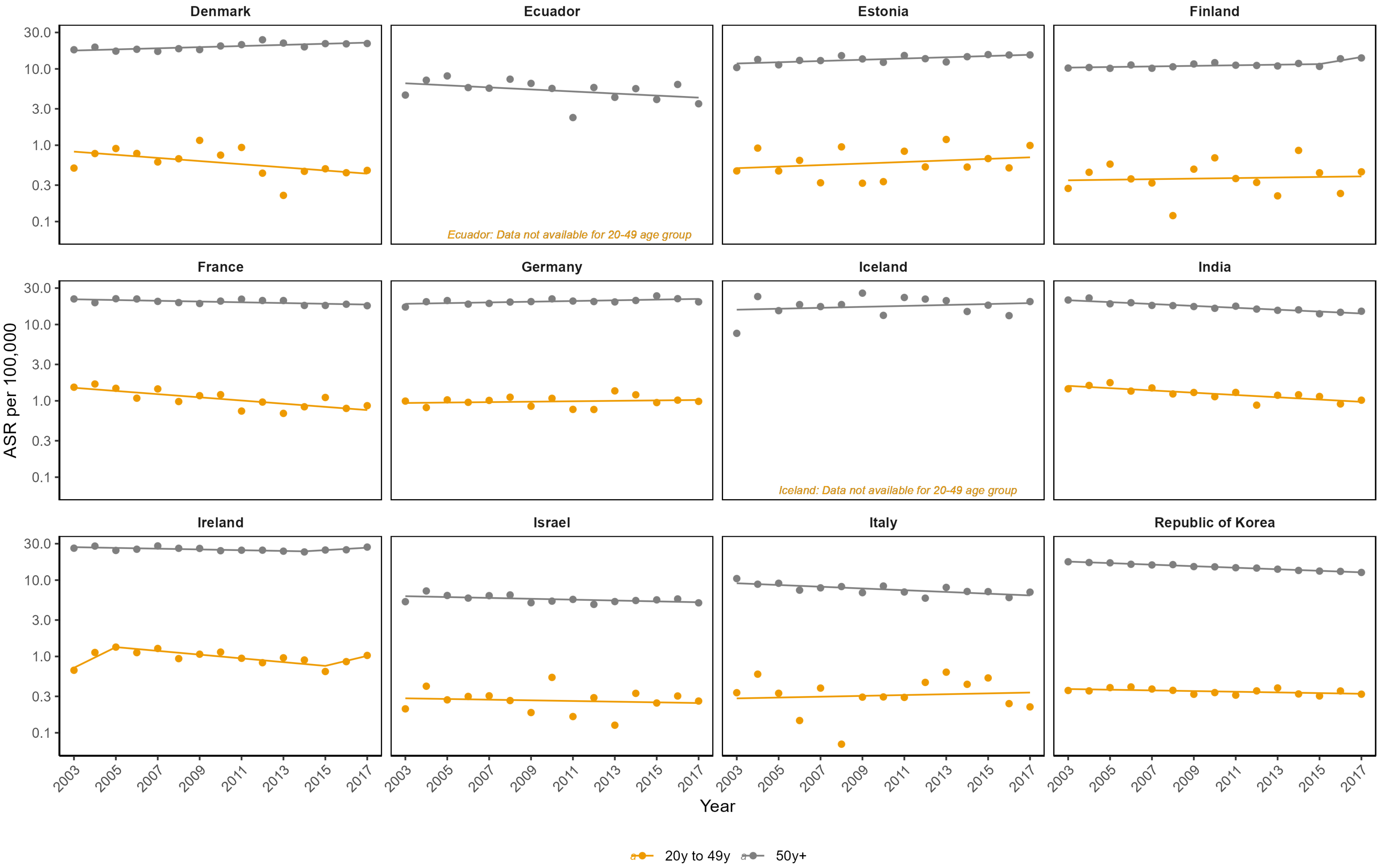
Denmark
Ecuador
Estonia
Finland
France
Germany
Iceland
India
Ireland
Israel
Italy
Republic of Korea
Ecuador: Data not available for 20-49 age group
Iceland: Data not available for 20-49 age group
ASR per 100,000
30.0
10.0
3.0
1.0
0.3
0.1
2003
2005
2007
2009
2011
2013
2015
2017
Year
20y to 49y
50y+

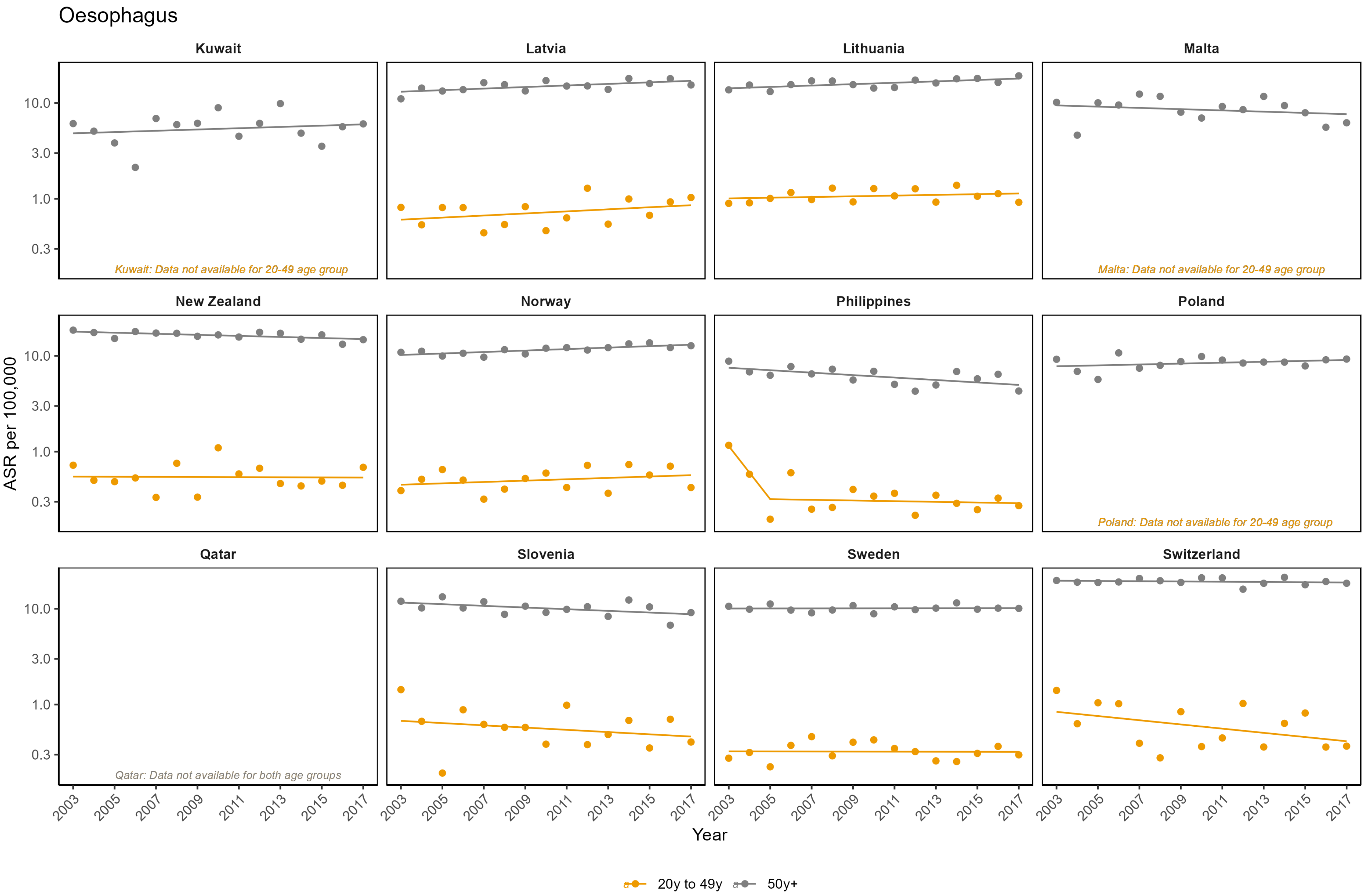
Oesophagus
Kuwait
Latvia
Lithuania
Malta
New Zealand
Norway
Philippines
Poland
Qatar
Slovenia
Sweden
Switzerland
Kuwait: Data not available for 20-49 age group
Malta: Data not available for 20-49 age group
Poland: Data not available for 20-49 age group
Qatar: Data not available for both age groups
ASR per 100,000
10.0
3.0
1.0
0.3
2003
2005
2007
2009
2011
2013
2015
2017
Year
20y to 49y
50y+

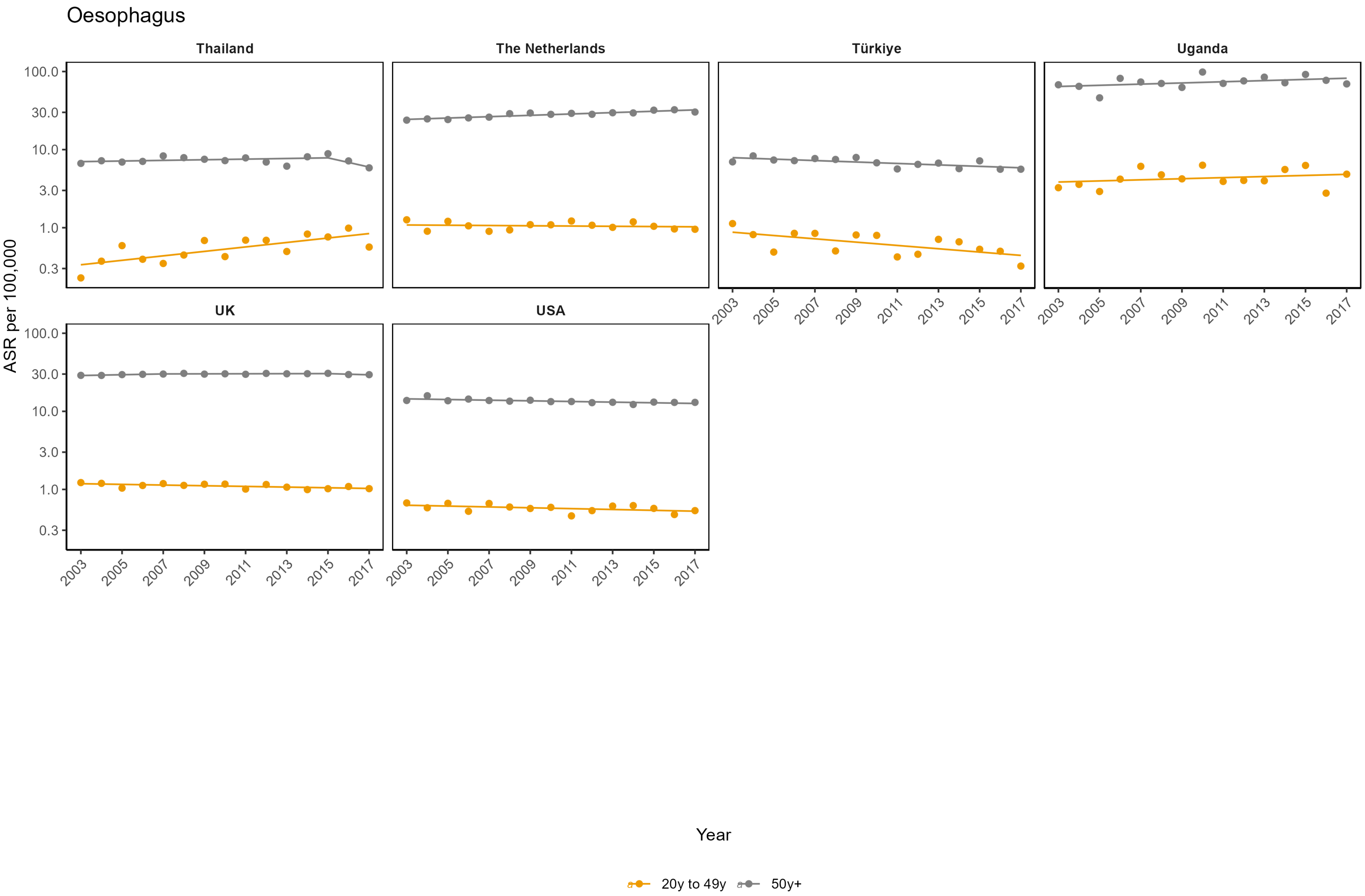

Oesophagus
Thailand
The Netherlands
Türkiye
Uganda
UK
USA
ASR per 100,000
100.0
30.0
10.0
3.0
1.0
0.3
2003
2005
2007
2009
2011
2013
2015
2017
Year
20y to 49y
50y+

**Appendix Figure 9: Age-standardised incidence rates (ASR) per 100,000**

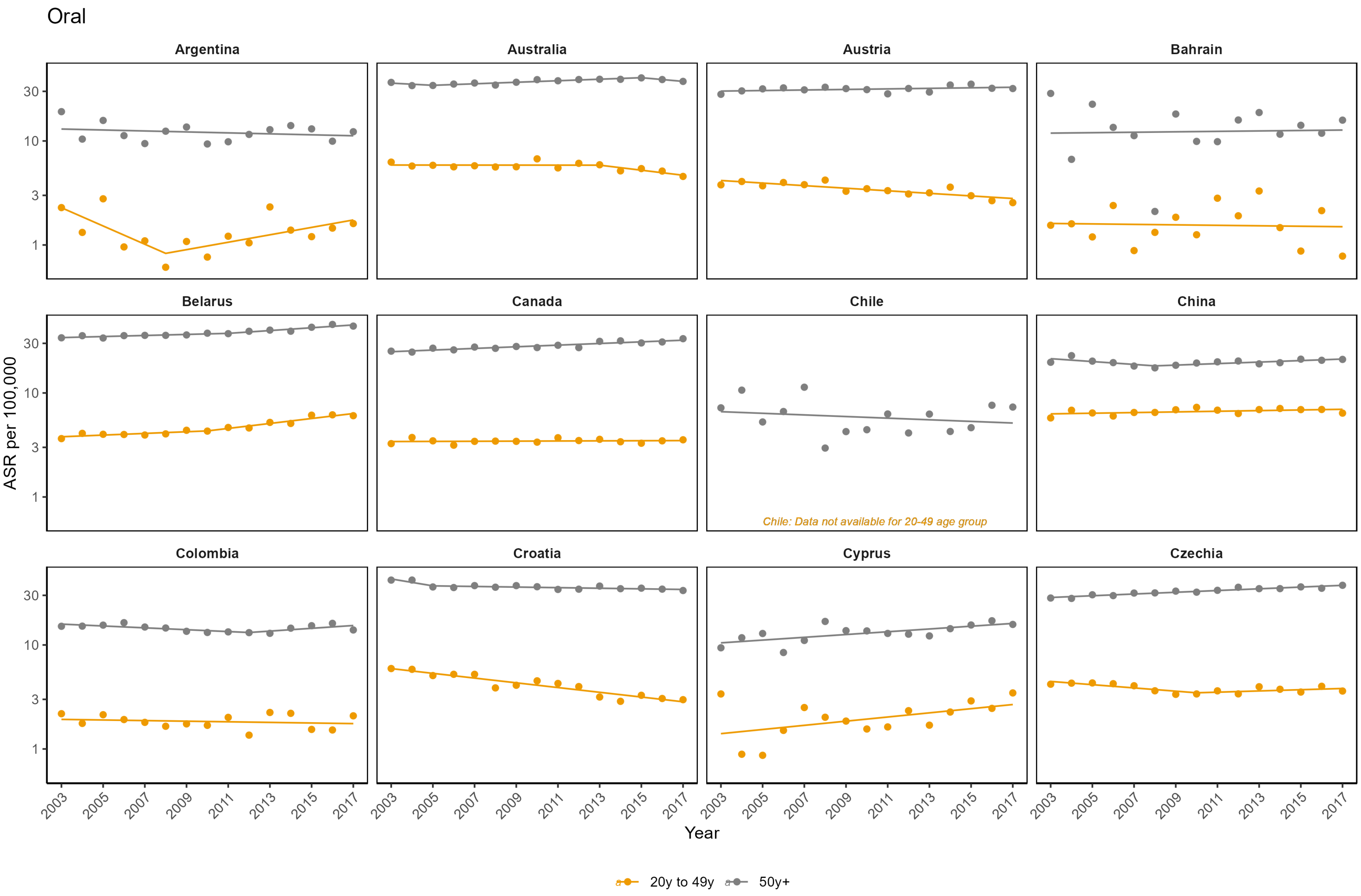

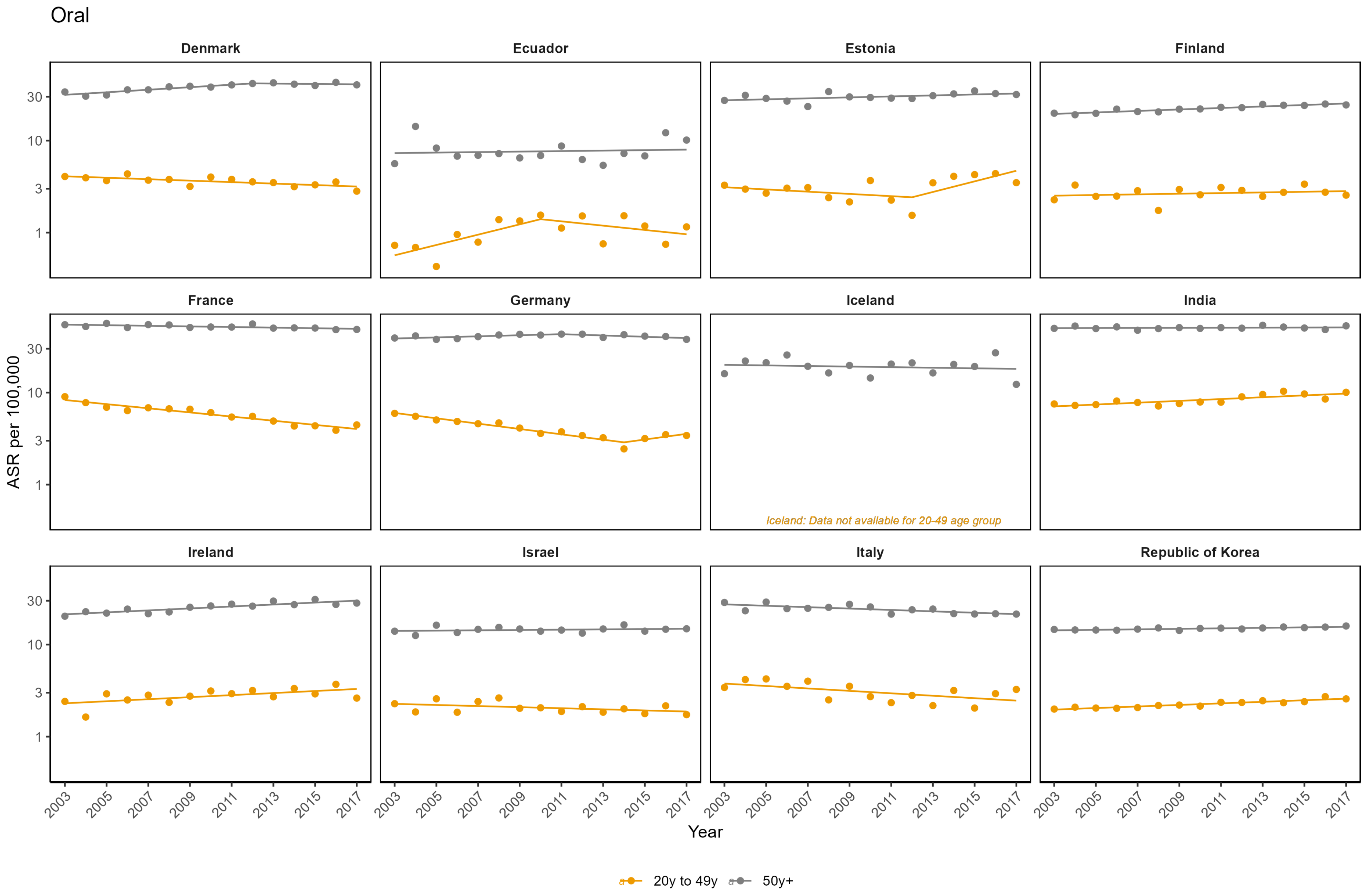

Oral
Denmark
Ecuador
Estonia
Finland
France
Germany
Iceland
India
Ireland
Israel
Italy
Republic of Korea
ASR per 100,000
30
10
3
1
Iceland: Data not available for 20-49 age group
2003
2005
2007
2009
2011
2013
2015
2017
Year
20y to 49y
50y+

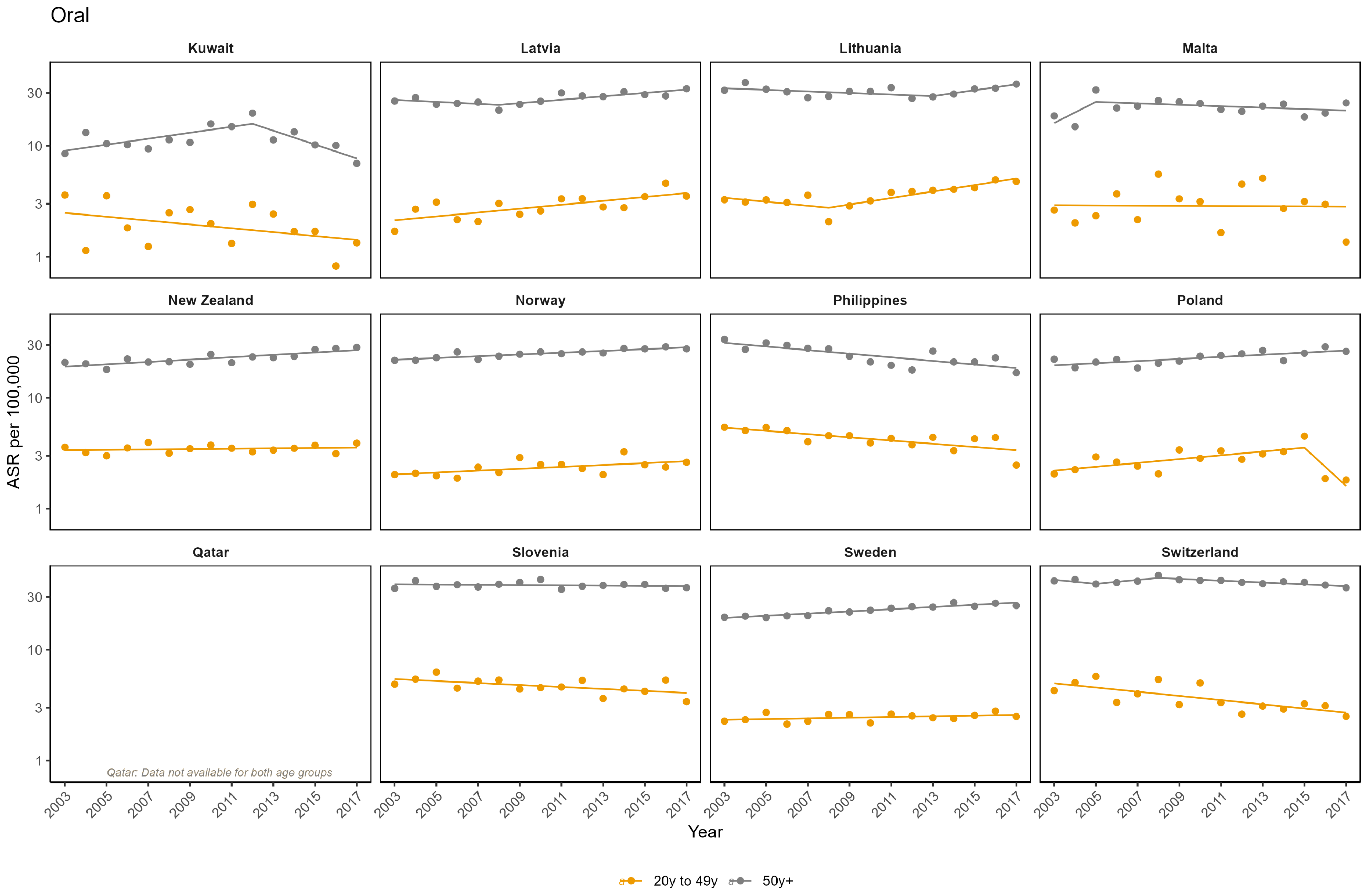
Oral
Kuwait
Latvia
Lithuania
Malta
New Zealand
Norway
Philippines
Poland
Qatar
Slovenia
Sweden
Switzerland
Qatar: Data not available for both age groups
ASR per 100,000
Year
20y to 49y
50y+

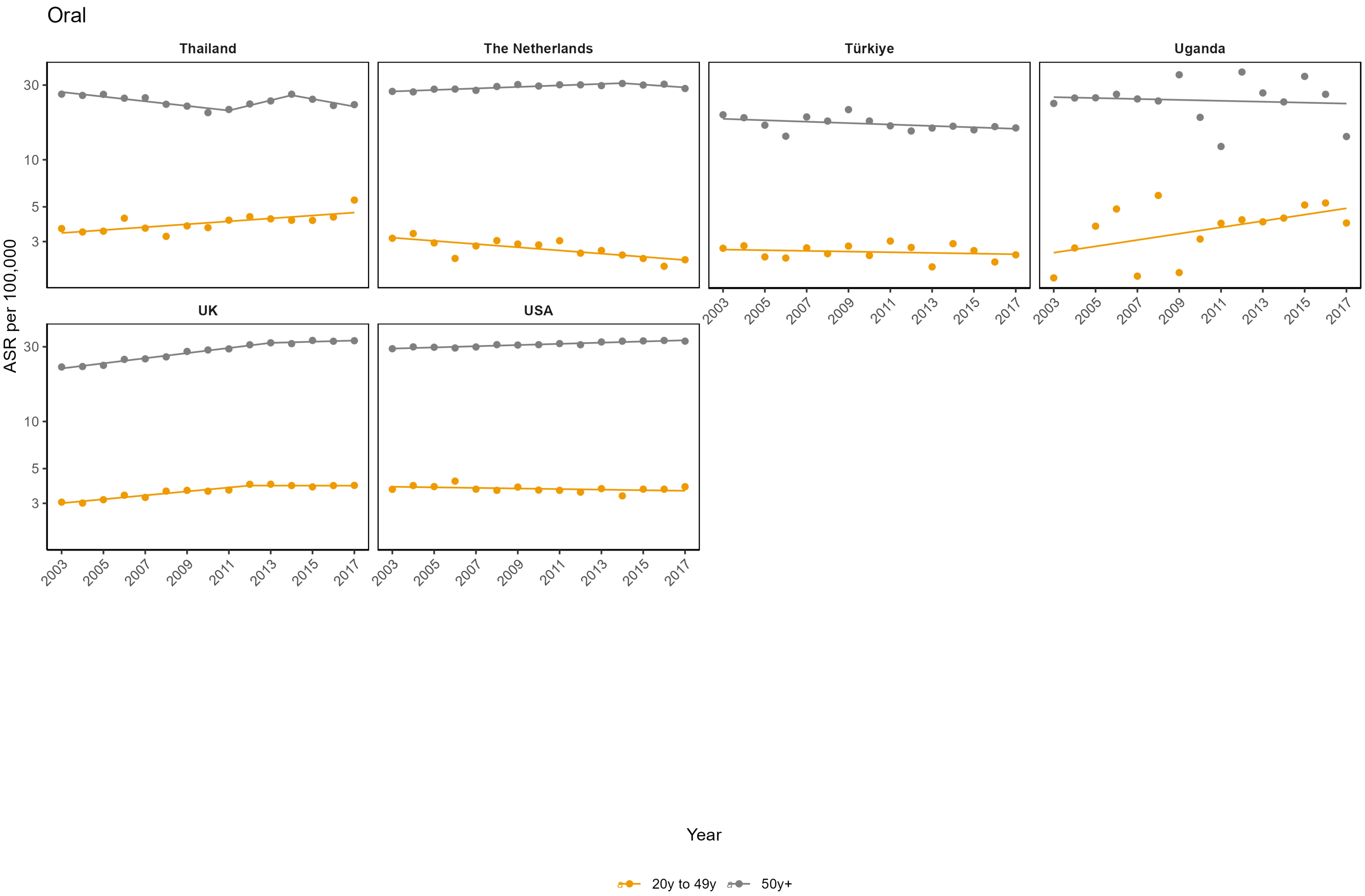
Oral
Thailand
The Netherlands
Türkiye
Uganda
UK
USA
ASR per 100,000
30
10
5
3
2003
2005
2007
2009
2011
2013
2015
2017
Year
20y to 49y
50y+

**Appendix Figure 10: Age-standardised incidence rates (ASR) per 100,000**

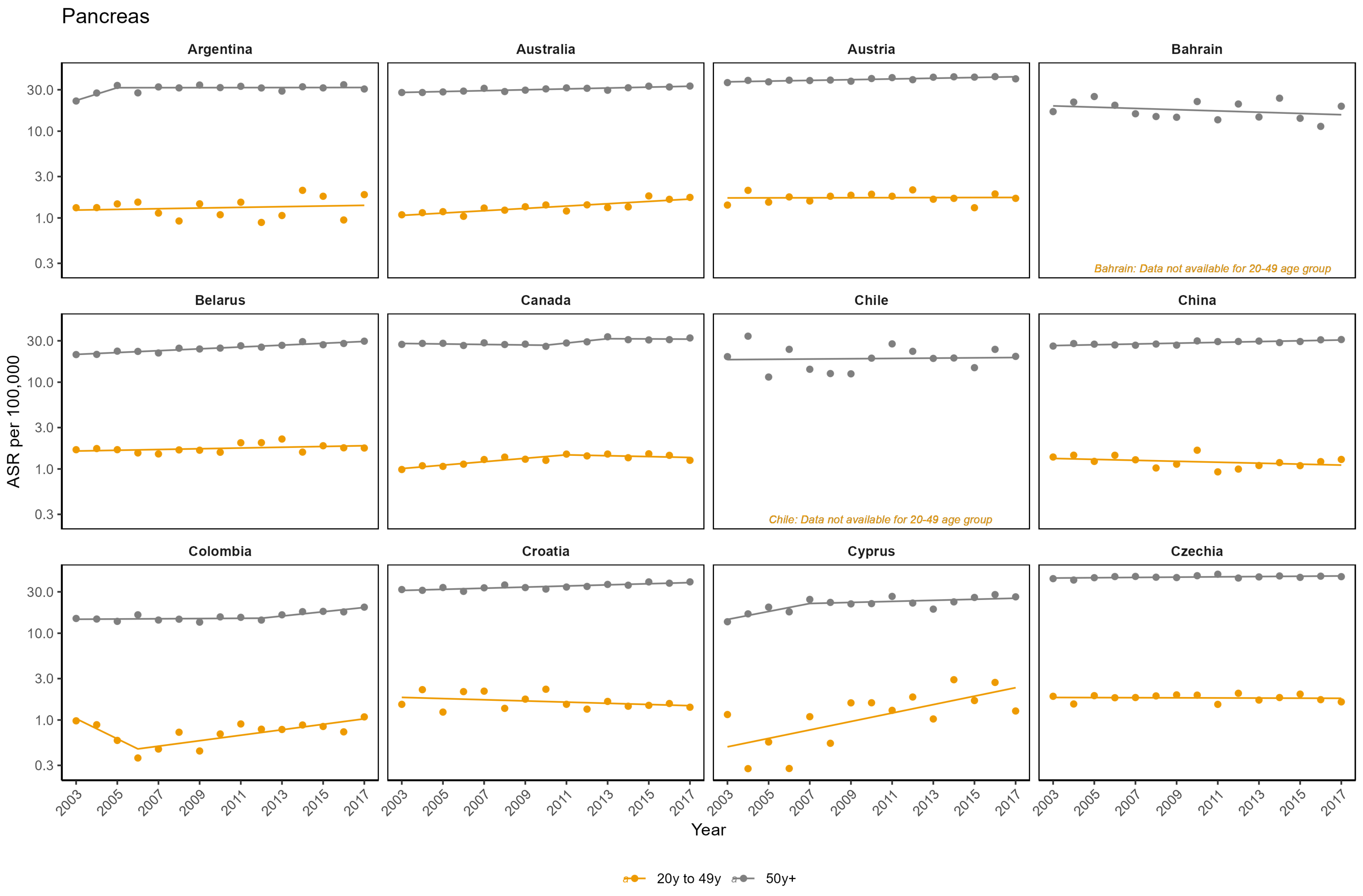

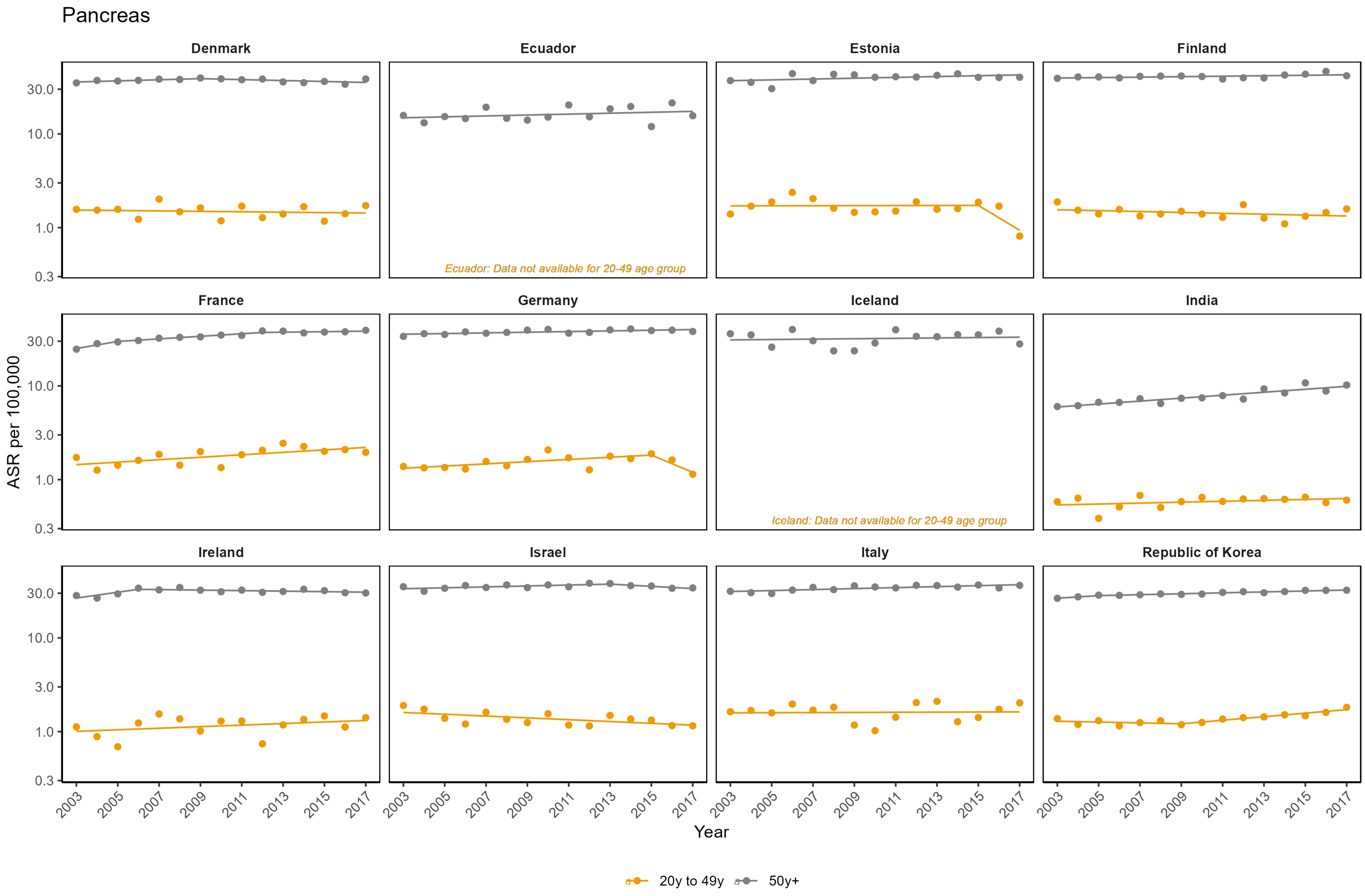
Pancreas
Denmark
Ecuador
Estonia
Finland
France
Germany
Iceland
India
Ireland
Israel
Italy
Republic of Korea
Ecuador: Data not available for 20-49 age group
Iceland: Data not available for 20-49 age group
ASR per 100,000
0.3
1.0
3.0
10.0
30.0
Year
2003
2005
2007
2009
2011
2013
2015
2017
20y to 49y
50y+

### Pancreas

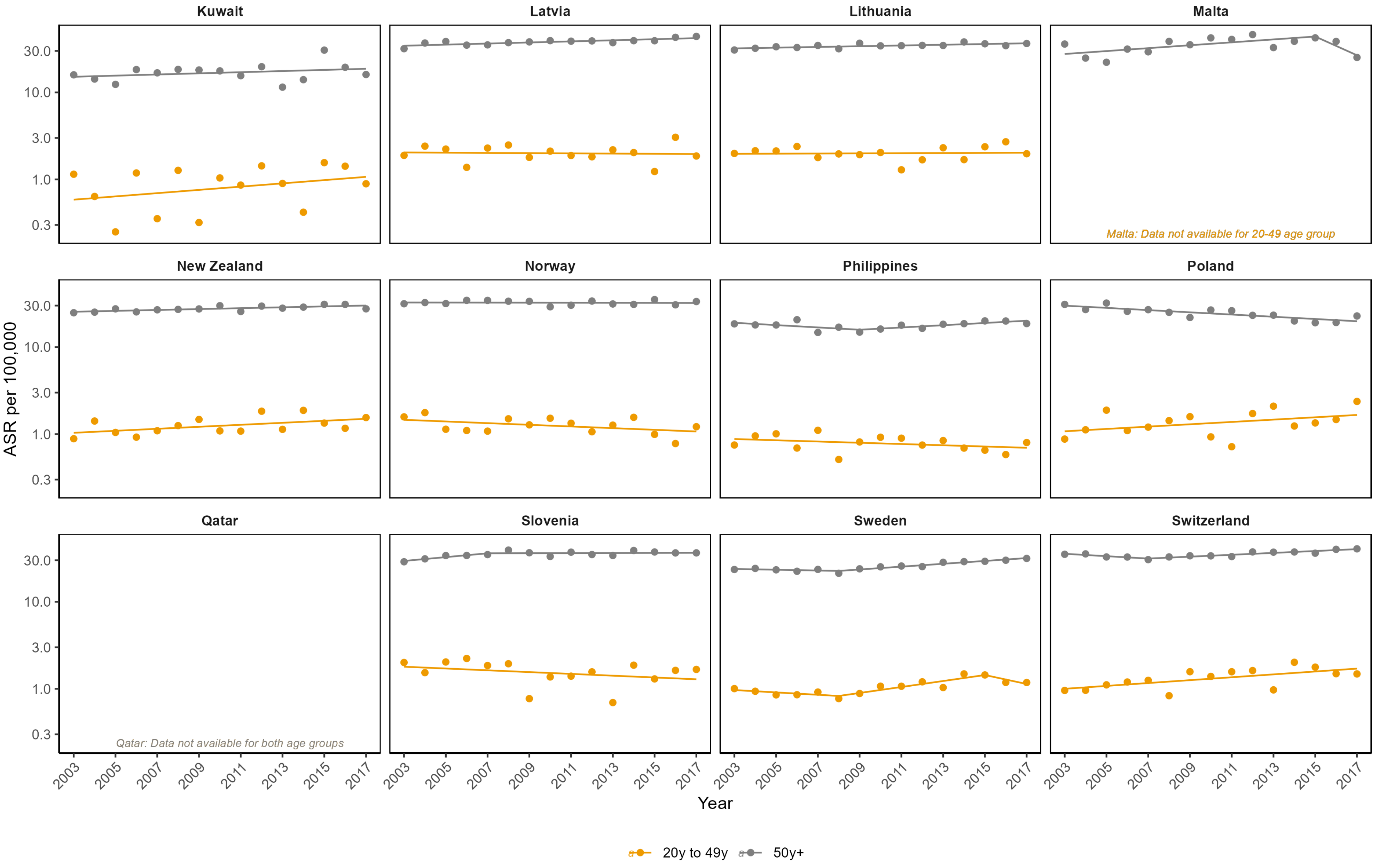
Kuwait
Latvia
Lithuania
Malta
Malta: Data not available for 20-49 age group
New Zealand
Norway
Philippines
Poland
Qatar
Qatar: Data not available for both age groups
Slovenia
Sweden
Switzerland
ASR per 100,000
30.0
10.0
3.0
1.0
0.3
2003
2005
2007
2009
2011
2013
2015
2017
Year
20y to 49y
50y+

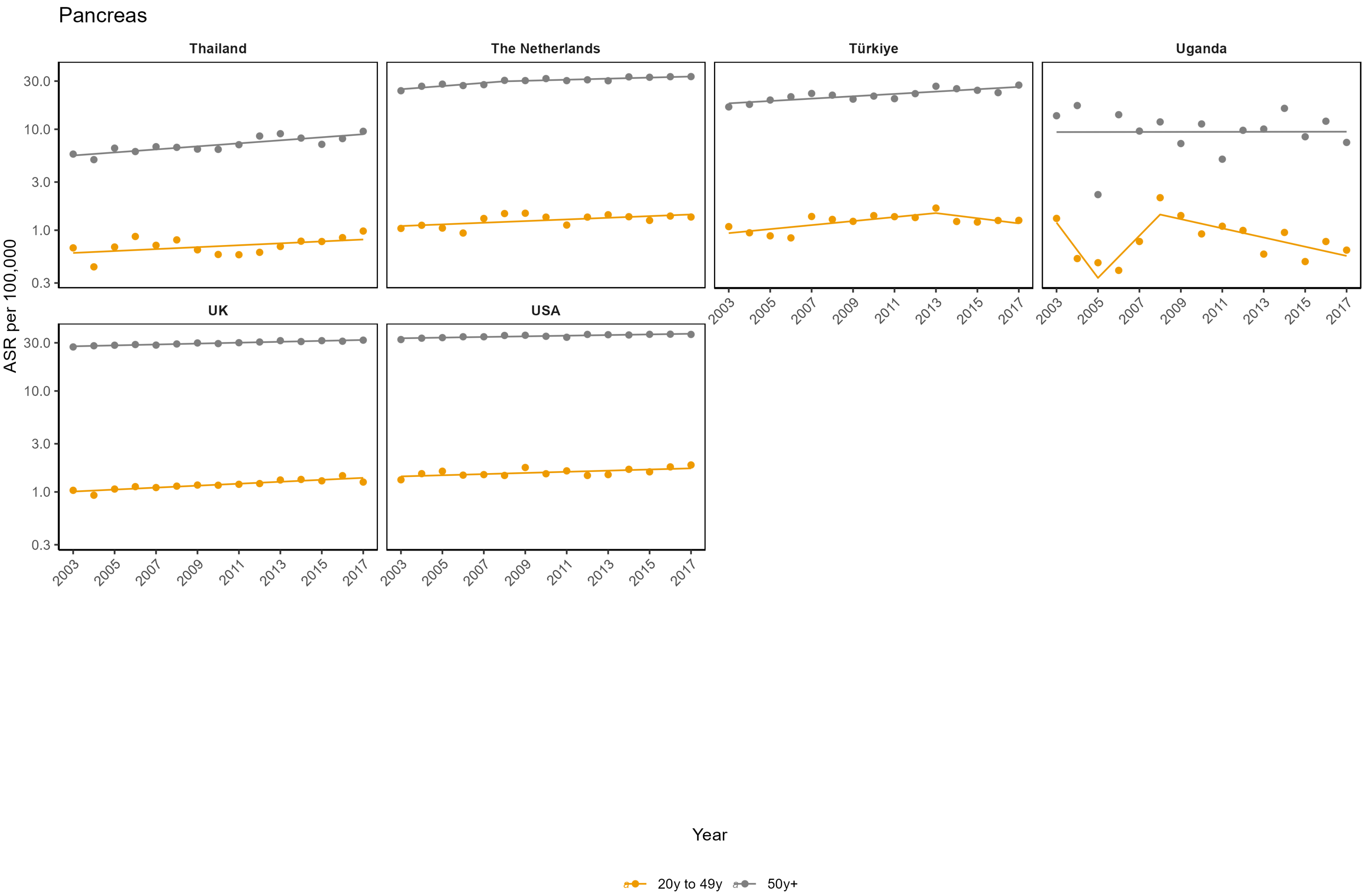

Pancreas
Thailand
The Netherlands
Türkiye
Uganda
UK
USA
ASR per 100,000
30.0
10.0
3.0
1.0
0.3
2003
2005
2007
2009
2011
2013
2015
2017
Year
20y to 49y
50y+

**Appendix Figure 11: Age-standardised incidence rates (ASR) per 100,000**

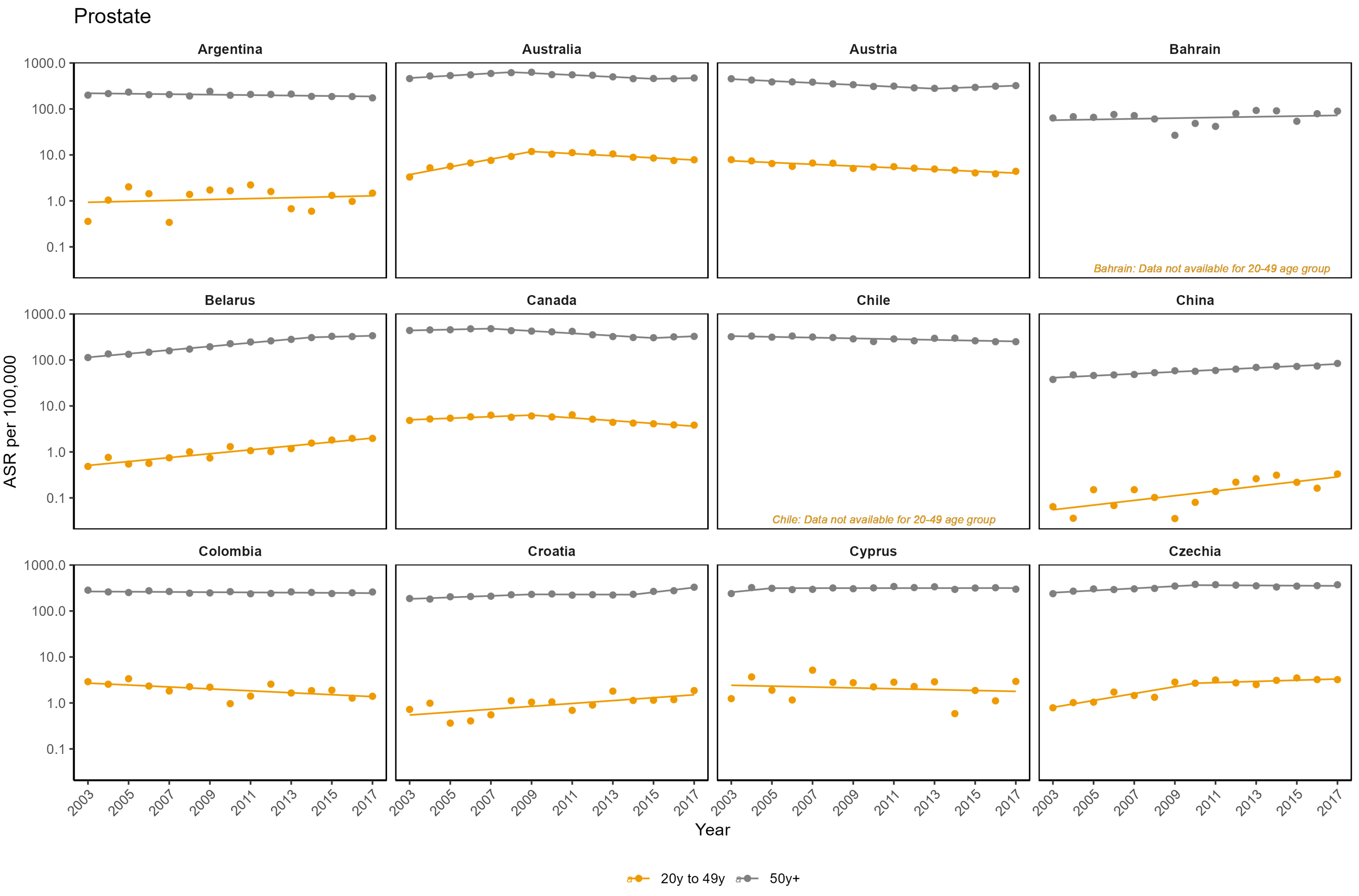

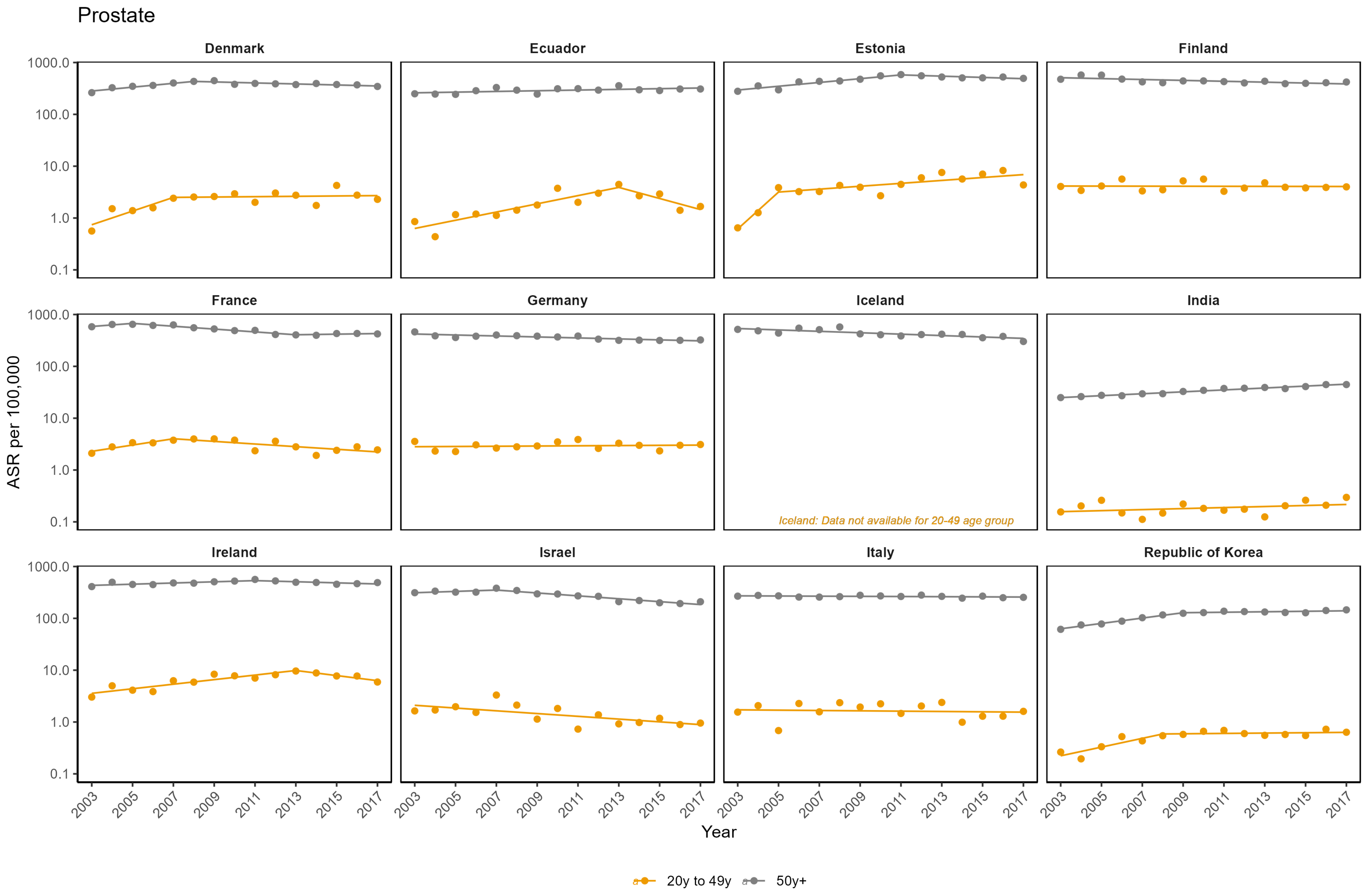

Prostate
Denmark
Ecuador
Estonia
Finland
France
Germany
Iceland
India
Ireland
Israel
Italy
Republic of Korea
Iceland: Data not available for 20-49 age group
ASR per 100,000
1000.0
100.0
10.0
1.0
0.1
2003
2005
2007
2009
2011
2013
2015
2017
Year
20y to 49y
50y+

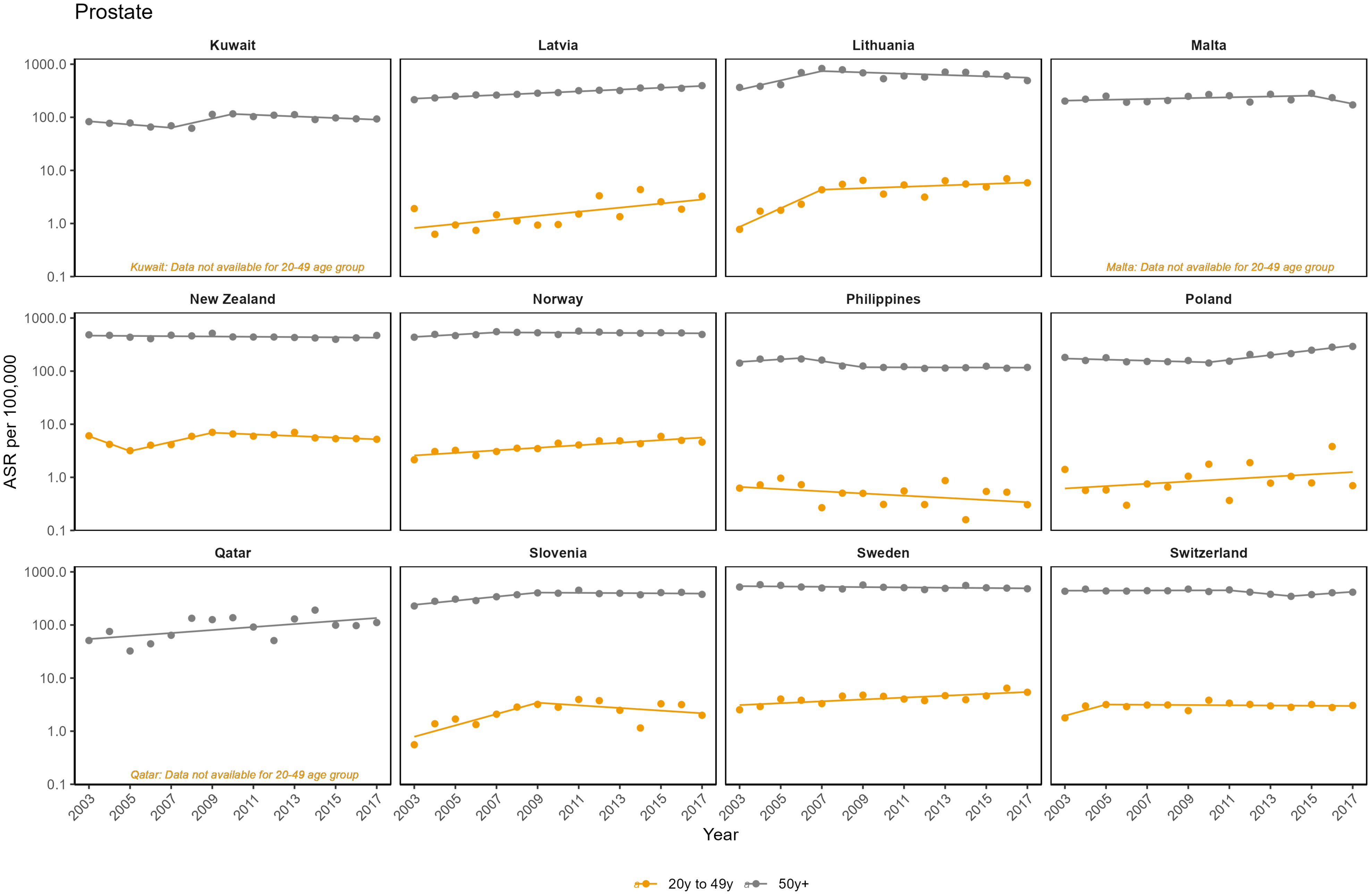

Prostate
Kuwait
Latvia
Lithuania
Malta
New Zealand
Norway
Philippines
Poland
Qatar
Slovenia
Sweden
Switzerland
1000.0
100.0
10.0
1.0
0.1
ASR per 100,000
Kuwait: Data not available for 20-49 age group
Malta: Data not available for 20-49 age group
Qatar: Data not available for 20-49 age group
2003
2005
2007
2009
2011
2013
2015
2017
Year
20y to 49y
50y+

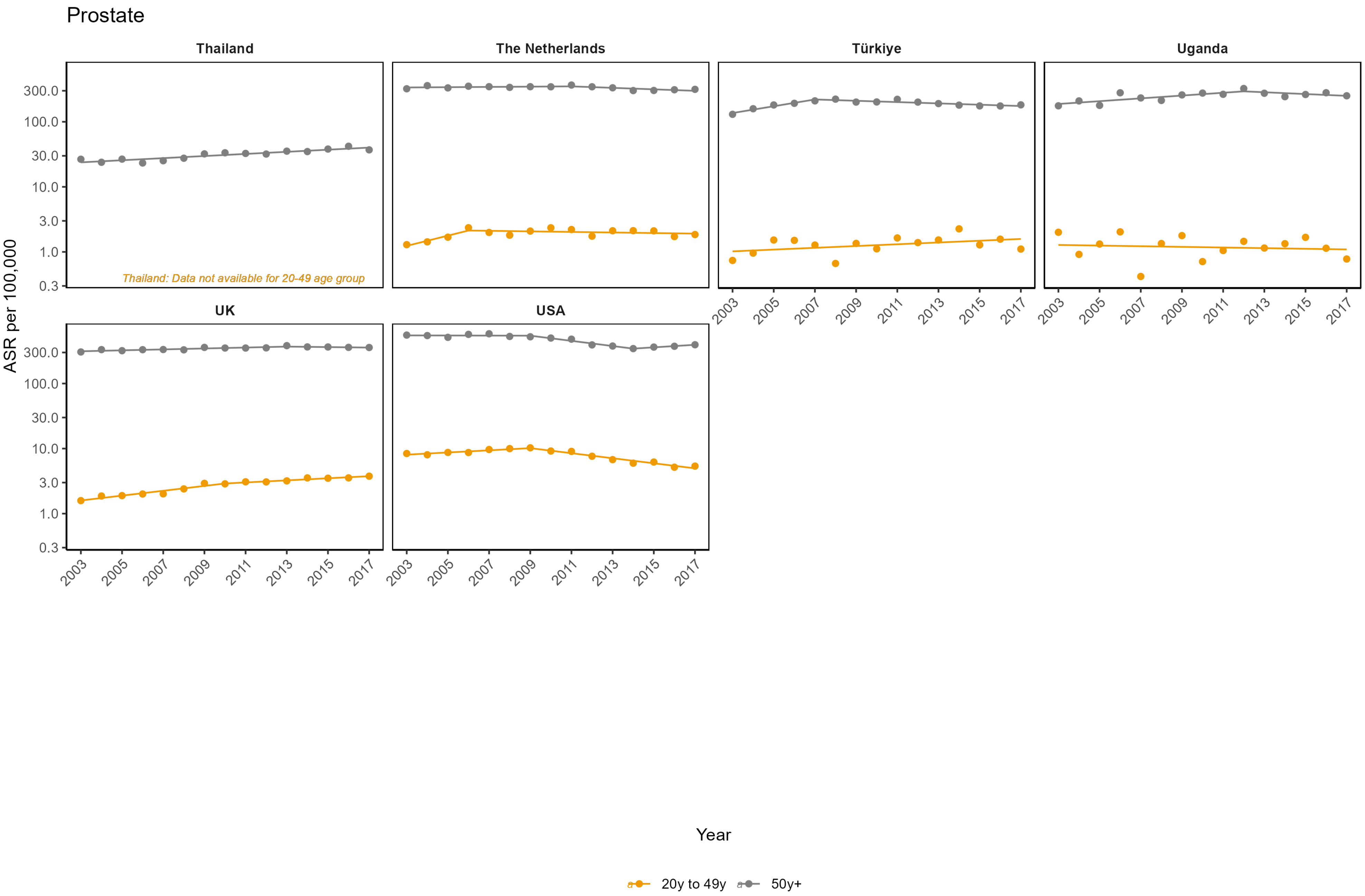
Prostate
Thailand
The Netherlands
Türkiye
Uganda
UK
USA
Thailand: Data not available for 20-49 age group
ASR per 100,000
300.0
100.0
30.0
10.0
3.0
1.0
0.3
2003
2005
2007
2009
2011
2013
2015
2017
Year
20y to 49y
50y+

**Appendix Figure 12: Age-standardised incidence rates (ASR) per 100,000**

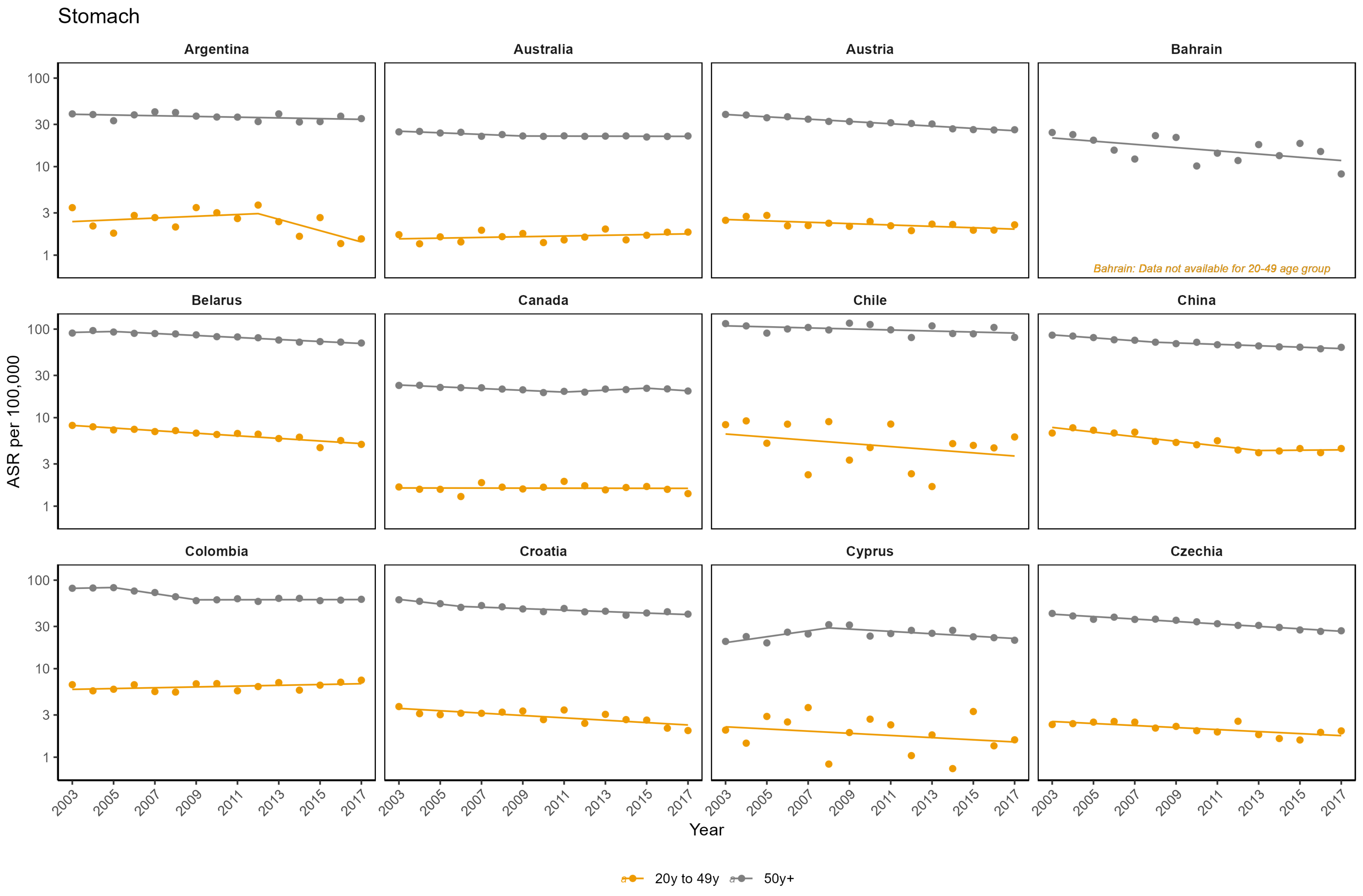

Stomach

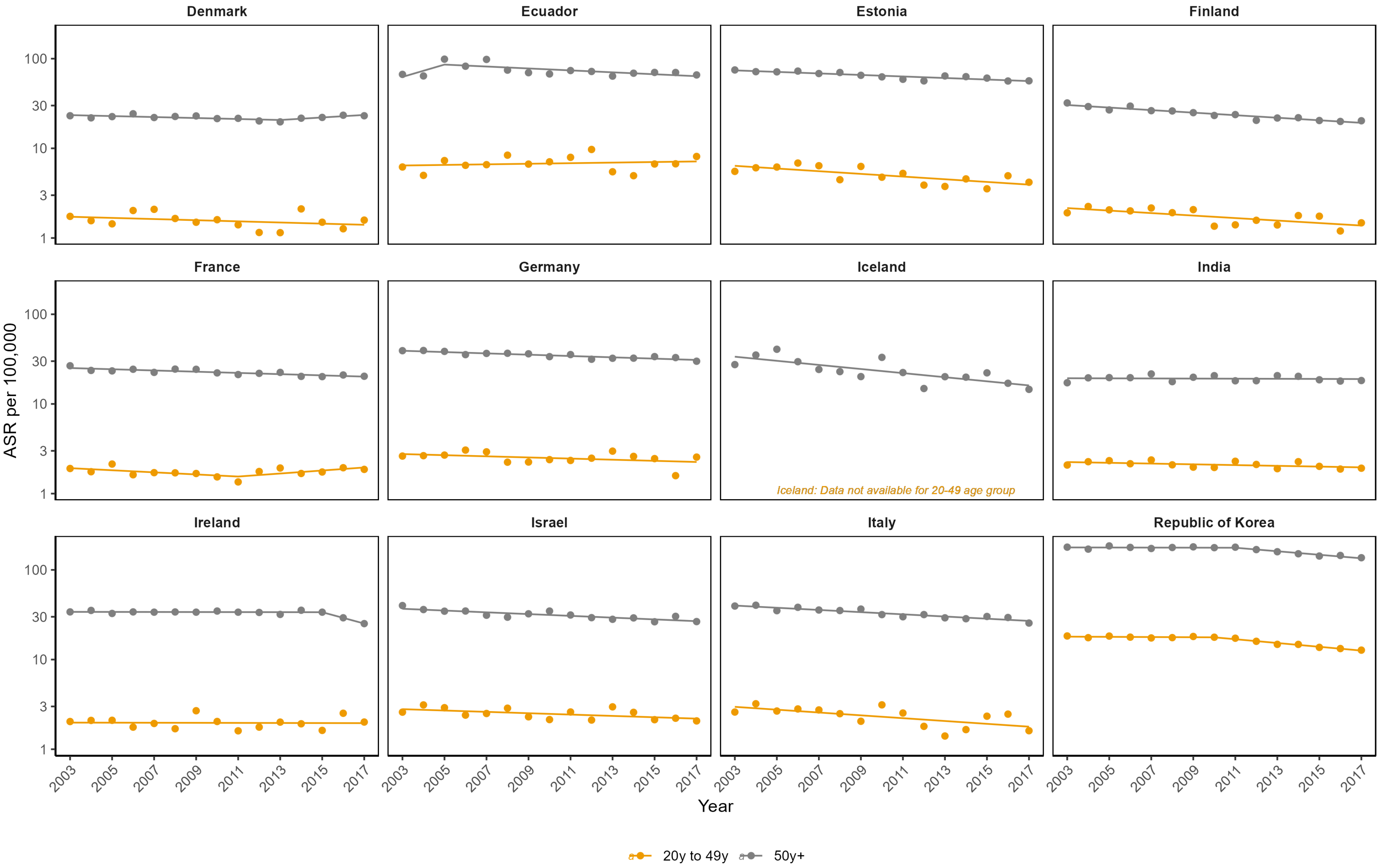
Denmark
Ecuador
Estonia
Finland
France
Germany
Iceland
India
Ireland
Israel
Italy
Republic of Korea
ASR per 100,000
100
30
10
3
1
2003
2005
2007
2009
2011
2013
2015
2017
Year
Iceland: Data not available for 20-49 age group
20y to 49y
50y+

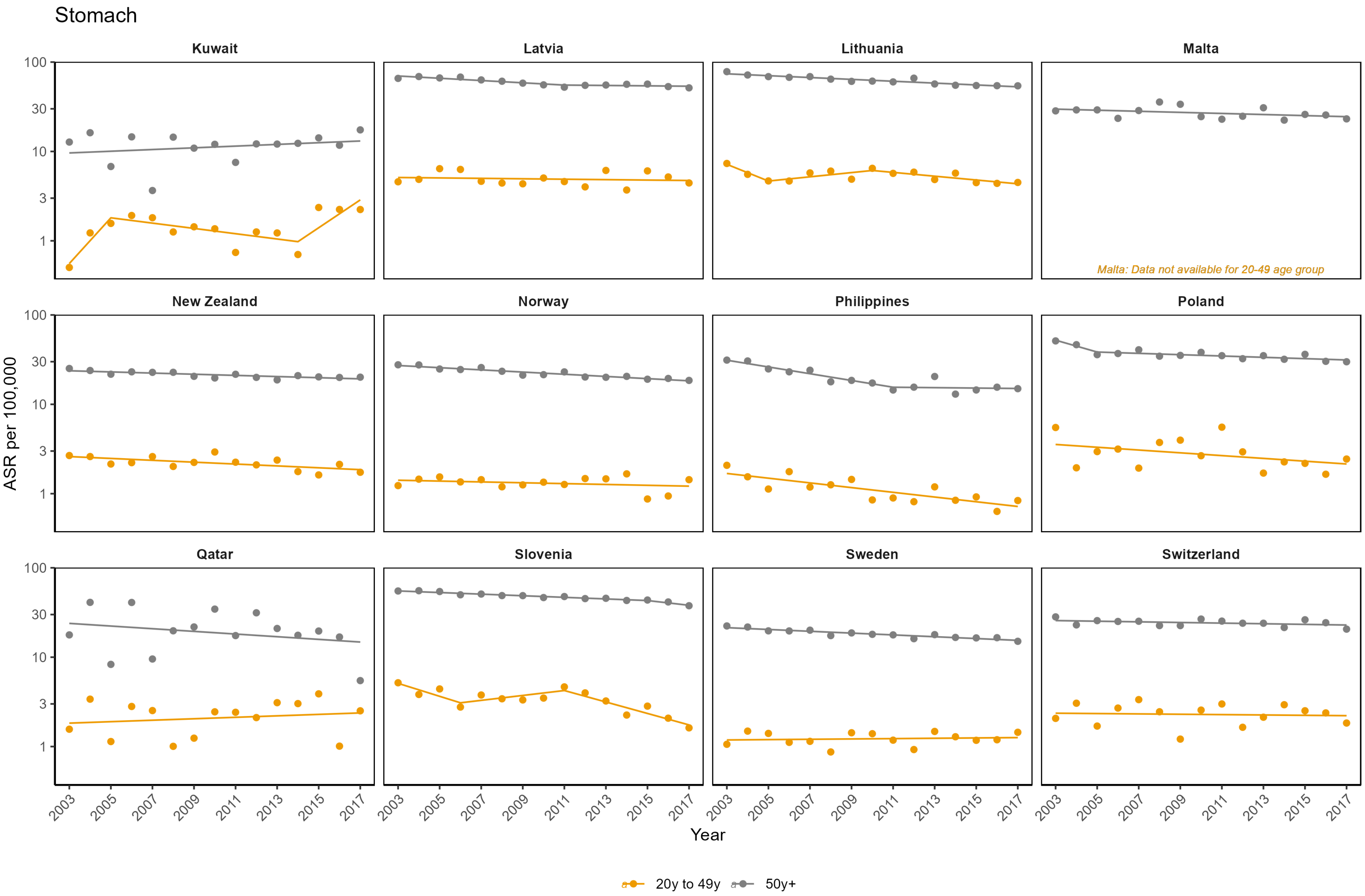
Stomach
Kuwait
Latvia
Lithuania
Malta
New Zealand
Norway
Philippines
Poland
Qatar
Slovenia
Sweden
Switzerland
Malta: Data not available for 20-49 age group
ASR per 100,000
100
30
10
3
1
2003
2005
2007
2009
2011
2013
2015
2017
Year
20y to 49y
50y+

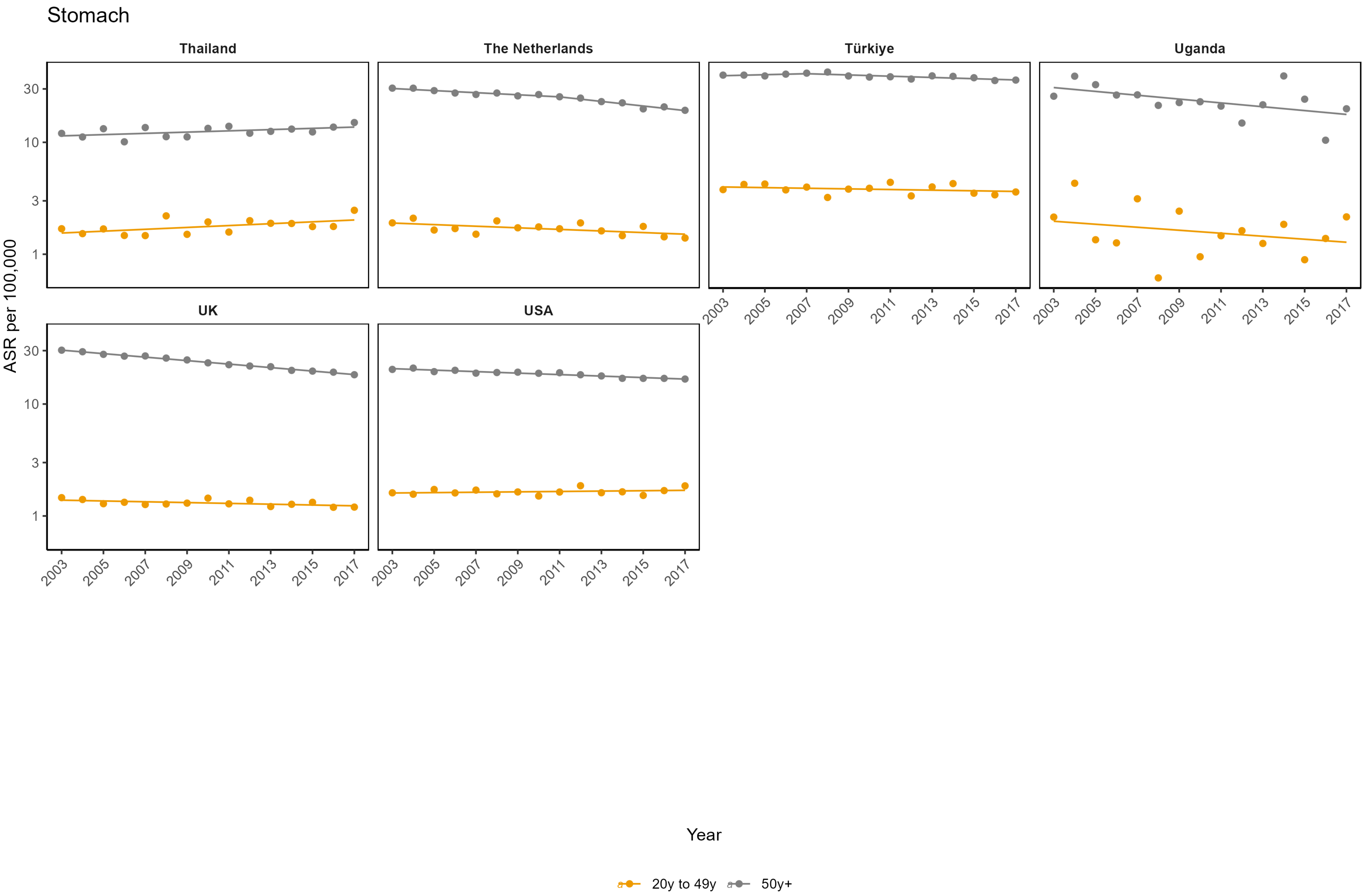
Stomach
Thailand
The Netherlands
Türkiye
Uganda
UK
USA
ASR per 100,000
30
10
3
1
2003
2005
2007
2009
2011
2013
2015
2017
Year
20y to 49y
50y+

**Appendix Figure 13: Age-standardised incidence rates (ASR) per 100,000**

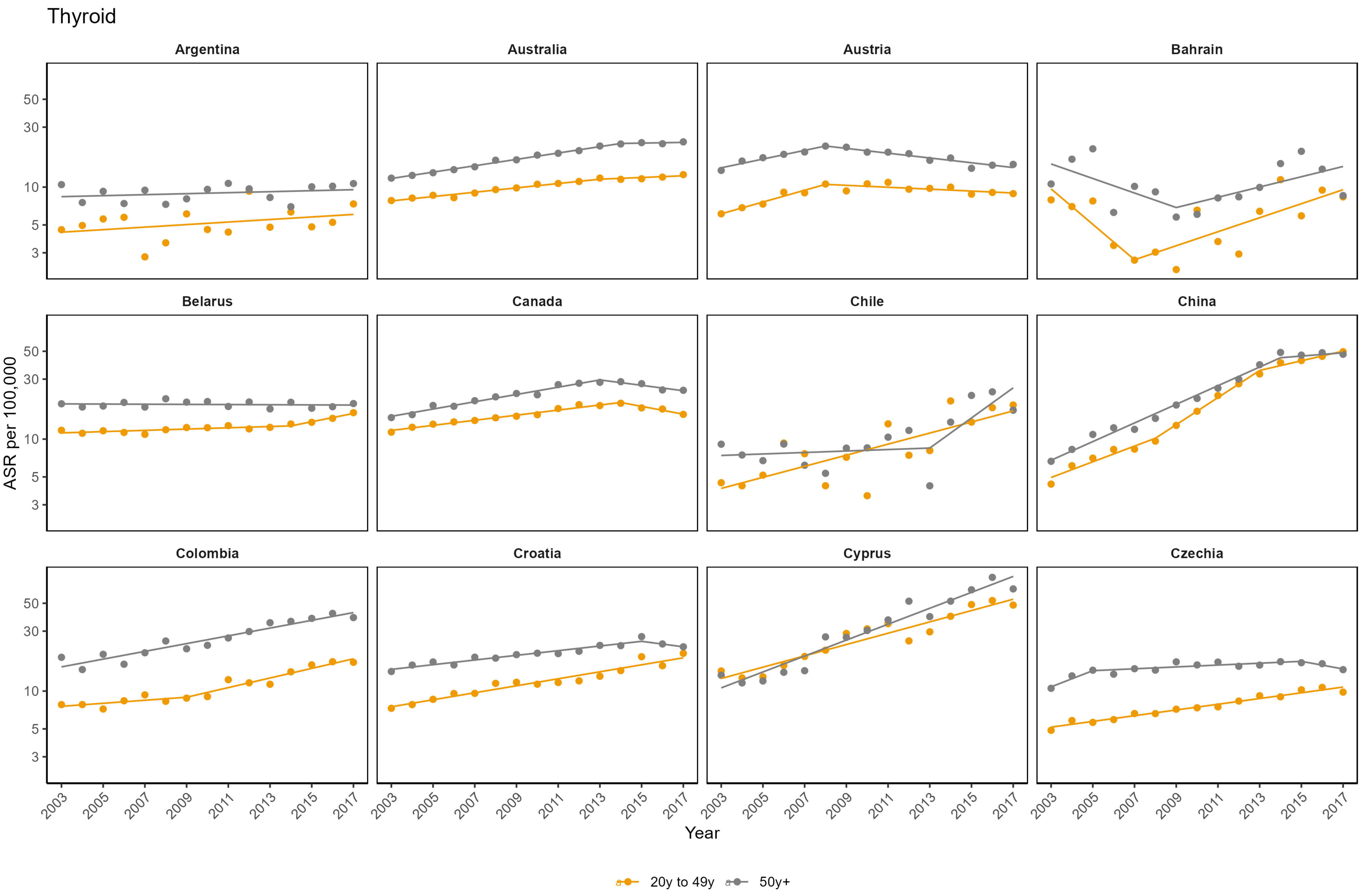

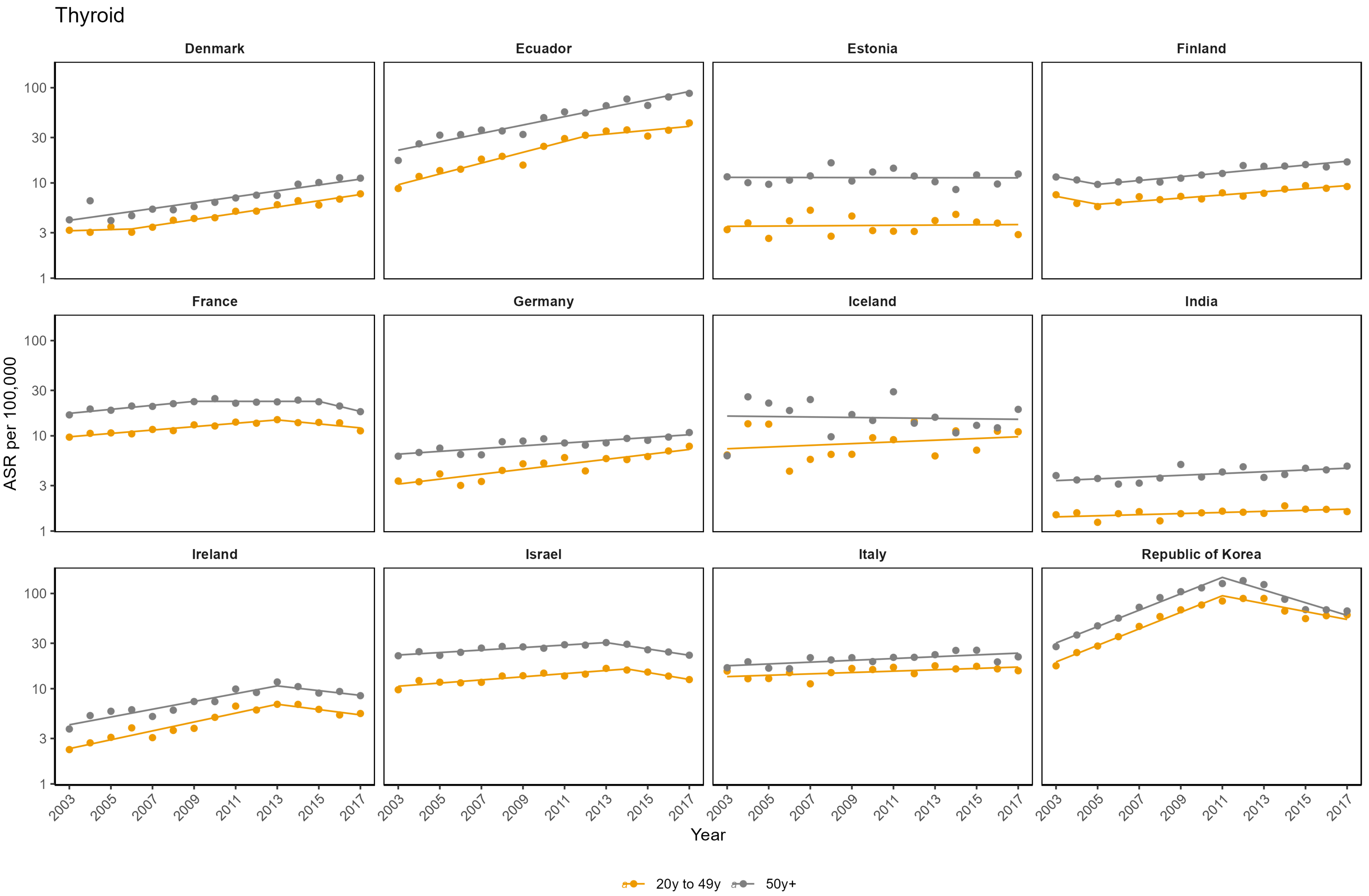
Thyroid
Denmark
Ecuador
Estonia
Finland
France
Germany
Iceland
India
Ireland
Israel
Italy
Republic of Korea
ASR per 100,000
1
3
10
30
100
2003
2005
2007
2009
2011
2013
2015
2017
Year
20y to 49y
50y+

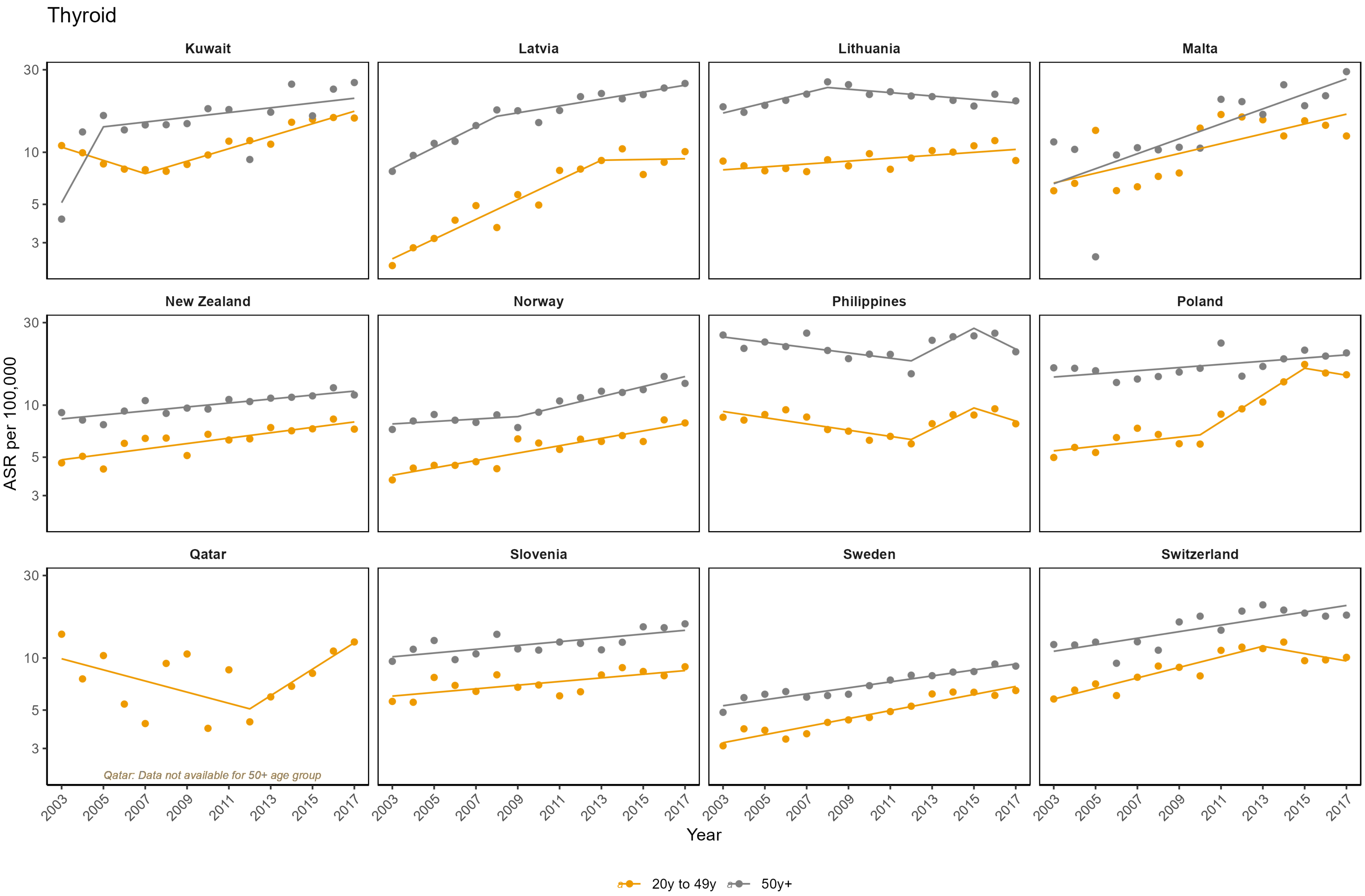
Thyroid
Kuwait
Latvia
Lithuania
Malta
New Zealand
Norway
Philippines
Poland
Qatar
Slovenia
Sweden
Switzerland
ASR per 100,000
30
10
5
3
Qatar: Data not available for 50+ age group
2003
2005
2007
2009
2011
2013
2015
2017
Year
20y to 49y
50y+

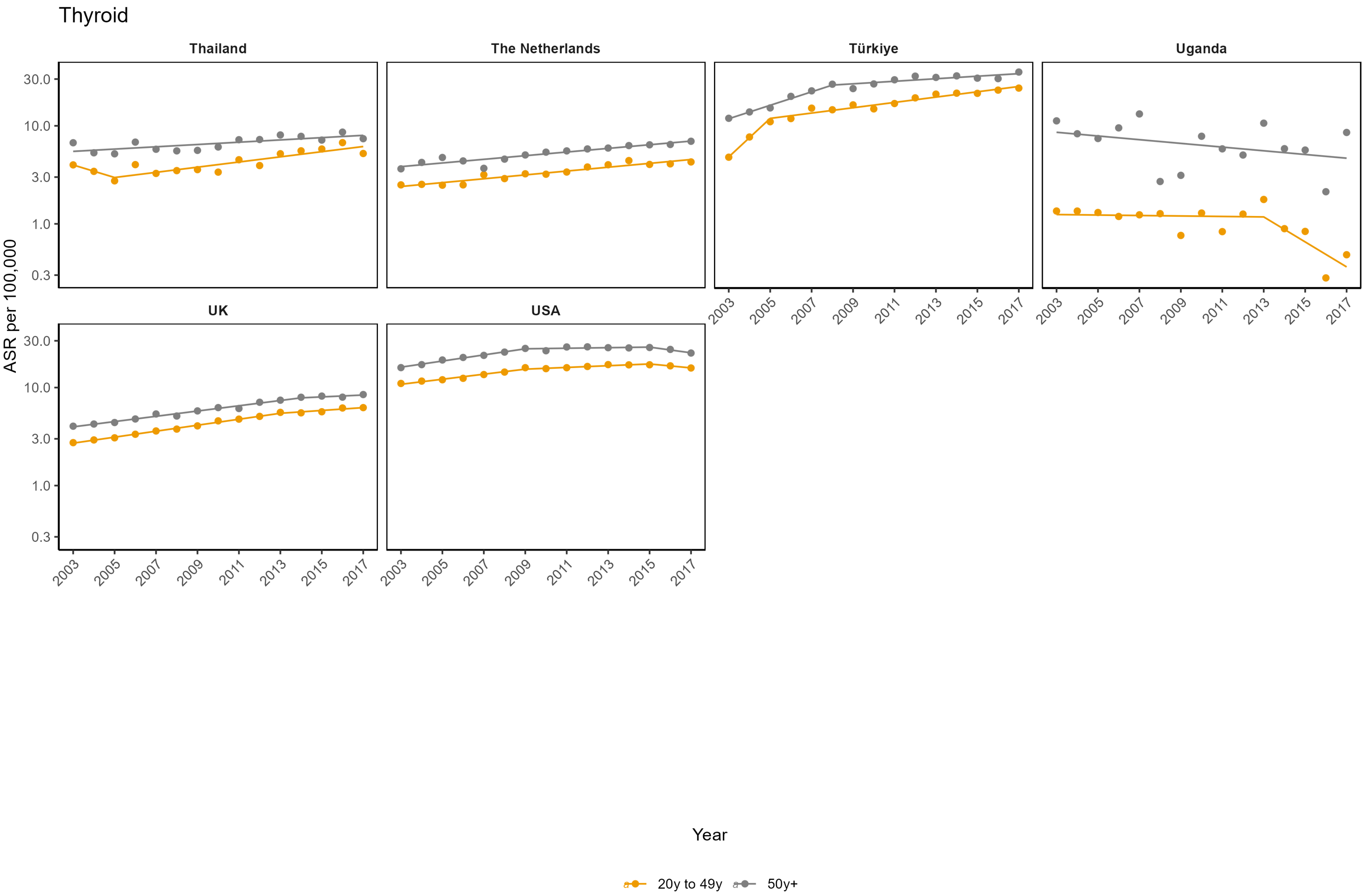

Thyroid
Thailand
The Netherlands
Türkiye
Uganda
UK
USA
ASR per 100,000
30.0
10.0
3.0
1.0
0.3
2003
2005
2007
2009
2011
2013
2015
2017
Year
20y to 49y
50y+

### Appendix Figure 14: Average annual percent change (AAPC) for cancer incidence (2003-2017) by UN region, country and age - Breast Cancer

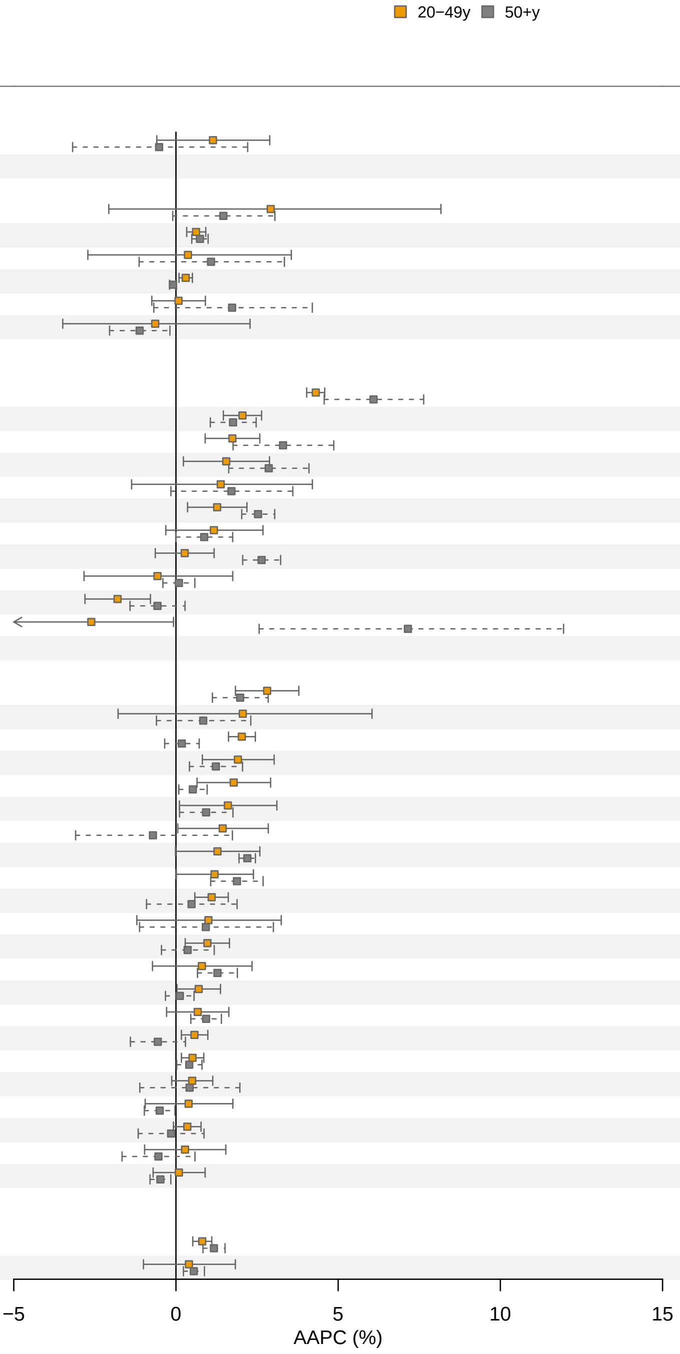

| Country | AAPC (%) 20−49y | AAPC (%) 50+y | p value |
|---|---|---|---|
| **Africa** | | | |
| Uganda | 1.14 | −0.52 | 0.29 |
| **Americas** | | | |
| Ecuador | 2.92 ‡ | 1.46 | 0.59 |
| Canada | 0.62 | 0.74 | 0.54 |
| Chile | 0.37 | 1.08 | 0.71 |
| USA | 0.3 | −0.09 | 0.002 |
| Colombia | 0.08 | 1.73 ‡ | 0.21 |
| Argentina | −0.64 ‡ | −1.12 | 0.76 |
| **Asia** | | | |
| Republic of Korea | 4.31 | 6.09 ‡ | 0.024 |
| Türkiye | 2.05 | 1.76 | 0.52 |
| Thailand | 1.74 | 3.3 ‡ | 0.081 |
| Kuwait | 1.55 | 2.86 | 0.147 |
| Bahrain | 1.38 | 1.71 | 0.84 |
| China | 1.27 | 2.53 | 0.020 |
| Cyprus | 1.17 | 0.87 | 0.72 |
| India | 0.27 | 2.64 | <0.001 |
| Israel | −0.57 ‡ | 0.09 | 0.58 |
| Philippines | −1.8 | −0.57 | 0.065 |
| Qatar | −2.61 | 7.15 | <0.001 |
| **Europe** | | | |
| Latvia | 2.81 | 1.98 | 0.199 |
| Malta | 2.06 ‡ | 0.84 ‡ | 0.57 |
| Czechia | 2.03 | 0.18 | <0.001 |
| Estonia | 1.91 | 1.23 | 0.31 |
| Slovenia | 1.78 ‡ | 0.52 | 0.042 |
| Croatia | 1.6 ‡ | 0.93 | 0.44 |
| Germany | 1.44 ‡ | −0.71 ‡ | 0.134 |
| Belarus | 1.28 ‡ | 2.2 | 0.176 |
| Poland | 1.19 | 1.88 | 0.33 |
| Denmark | 1.1 | 0.48 ‡ | 0.42 |
| Lithuania | 1 ‡ | 0.92 ‡ | 0.96 |
| Norway | 0.97 | 0.36 ‡ | 0.26 |
| Finland | 0.8 ‡ | 1.28 ‡ | 0.57 |
| UK | 0.7 ‡ | 0.12 ‡ | 0.153 |
| Italy | 0.67 ‡ | 0.93 | 0.63 |
| France | 0.57 | −0.56 ‡ | 0.019 |
| The Netherlands | 0.51 | 0.41 ‡ | 0.70 |
| Ireland | 0.5 | 0.42 ‡ | 0.92 |
| Austria | 0.39 ‡ | −0.5 | 0.22 |
| Sweden | 0.35 | −0.15 ‡ | 0.37 |
| Iceland | 0.28 | −0.54 | 0.32 |
| Switzerland | 0.09 | −0.48 | 0.176 |
| **Oceania** | | | |
| Australia | 0.81 | 1.17 | 0.107 |
| New Zealand | 0.4 ‡ | 0.55 | 0.84 |

Data for female sex only.
‡ best fitted Joinpoint model > 0 joins; * p−value for a difference between AAPC 20−49y and AAPC 50+y.

#### Appendix Figure 15: Average annual percent change (AAPC) for cancer incidence (2003-2017) by UN region, country and age - Colorectal Cancer

■ 20−49y ■ 50+y

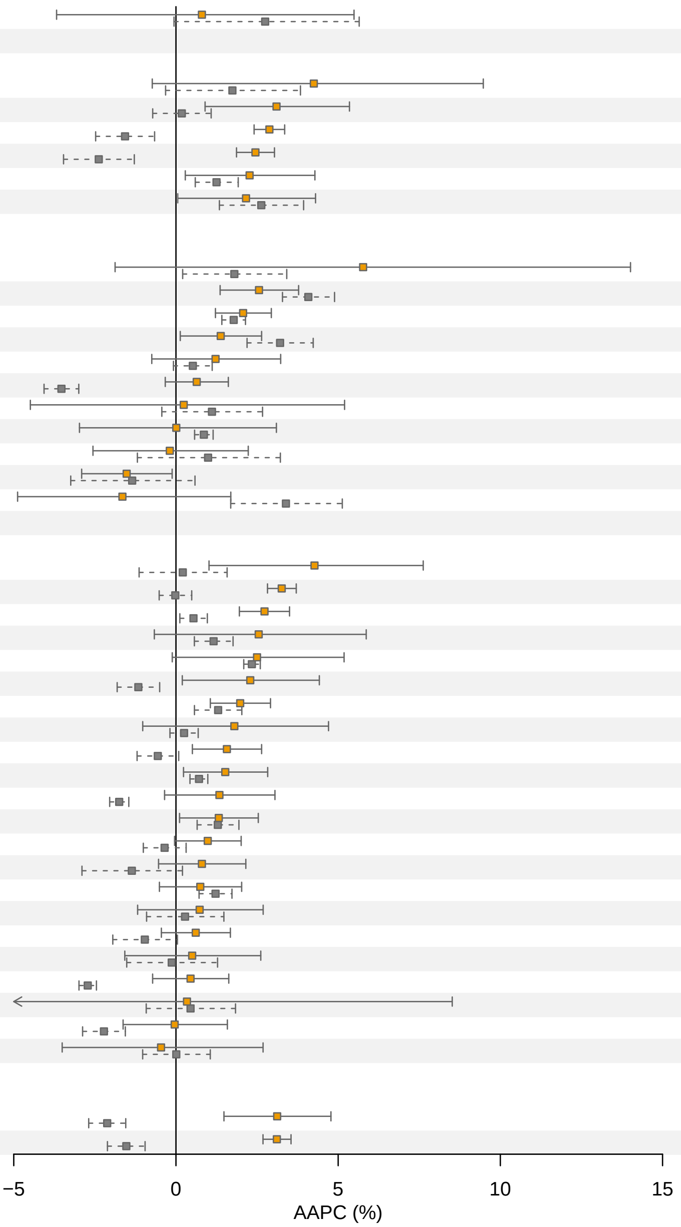

| Country | AAPC (%) 20−49y | AAPC (%) 50+y | p value |
|---|---|---|---|
| **Africa** | | | |
| Uganda | 0.8 | 2.75 | 0.46 |
| **Americas** | | | |
| Chile | 4.25 | 1.74 | 0.34 |
| Argentina | 3.1 | 0.18 | 0.019 |
| Canada | 2.88 | −1.57 ‡ | <0.001 |
| USA | 2.45 ‡ | −2.38 ‡ | <0.001 |
| Colombia | 2.27 ‡ | 1.25 | 0.34 |
| Ecuador | 2.16 | 2.63 | 0.70 |
| **Asia** | | | |
| Kuwait | 5.77 ‡ | 1.8 | 0.33 |
| Thailand | 2.56 | 4.08 | 0.041 |
| Türkiye | 2.07 | 1.78 | 0.51 |
| India | 1.38 | 3.21 ‡ | 0.028 |
| Republic of Korea | 1.22 ‡ | 0.52 ‡ | 0.50 |
| Israel | 0.64 | −3.53 | <0.001 |
| Qatar | 0.24 | 1.11 | 0.72 |
| China | 0.01 ‡ | 0.86 | 0.59 |
| Cyprus | −0.19 | 0.99 ‡ | 0.47 |
| Philippines | −1.52 | −1.35 ‡ | 0.88 |
| Bahrain | −1.65 | 3.39 | 0.011 |
| **Europe** | | | |
| Iceland | 4.27 | 0.21 | 0.026 |
| UK | 3.26 | −0.02 ‡ | <0.001 |
| Sweden | 2.73 | 0.54 ‡ | <0.001 |
| Estonia | 2.55 | 1.16 | 0.38 |
| Belarus | 2.5 ‡ | 2.34 | 0.91 |
| Switzerland | 2.29 | −1.16 | 0.004 |
| Denmark | 1.98 | 1.3 | 0.24 |
| Norway | 1.8 ‡ | 0.25 ‡ | 0.29 |
| Ireland | 1.57 | −0.56 ‡ | 0.002 |
| Finland | 1.52 | 0.71 | 0.21 |
| Germany | 1.34 ‡ | −1.75 | <0.001 |
| The Netherlands | 1.32 ‡ | 1.29 | 0.97 |
| France | 0.98 ‡ | −0.35 | 0.032 |
| Slovenia | 0.8 | −1.36 ‡ | 0.039 |
| Croatia | 0.75 | 1.22 | 0.48 |
| Poland | 0.73 | 0.28 ‡ | 0.68 |
| Italy | 0.61 | −0.96 ‡ | 0.035 |
| Latvia | 0.5 | −0.13 ‡ | 0.61 |
| Austria | 0.45 | −2.72 | <0.001 |
| Malta | 0.34 ‡ | 0.45 | 0.98 |
| Czechia | −0.04 ‡ | −2.22 ‡ | 0.013 |
| Lithuania | −0.46 ‡ | 0.01 ‡ | 0.78 |
| **Oceania** | | | |
| New Zealand | 3.12 ‡ | −2.12 ‡ | <0.001 |
| Australia | 3.11 | −1.53 ‡ | <0.001 |

‡ best fitted Joinpoint model > 0 joins; * p−value for a difference between AAPC 20−49y and AAPC 50+y.

#### Appendix Figure 16: Average annual percent change (AAPC) for cancer incidence (2003-2017) by UN region, country and age - Endometrial Cancer

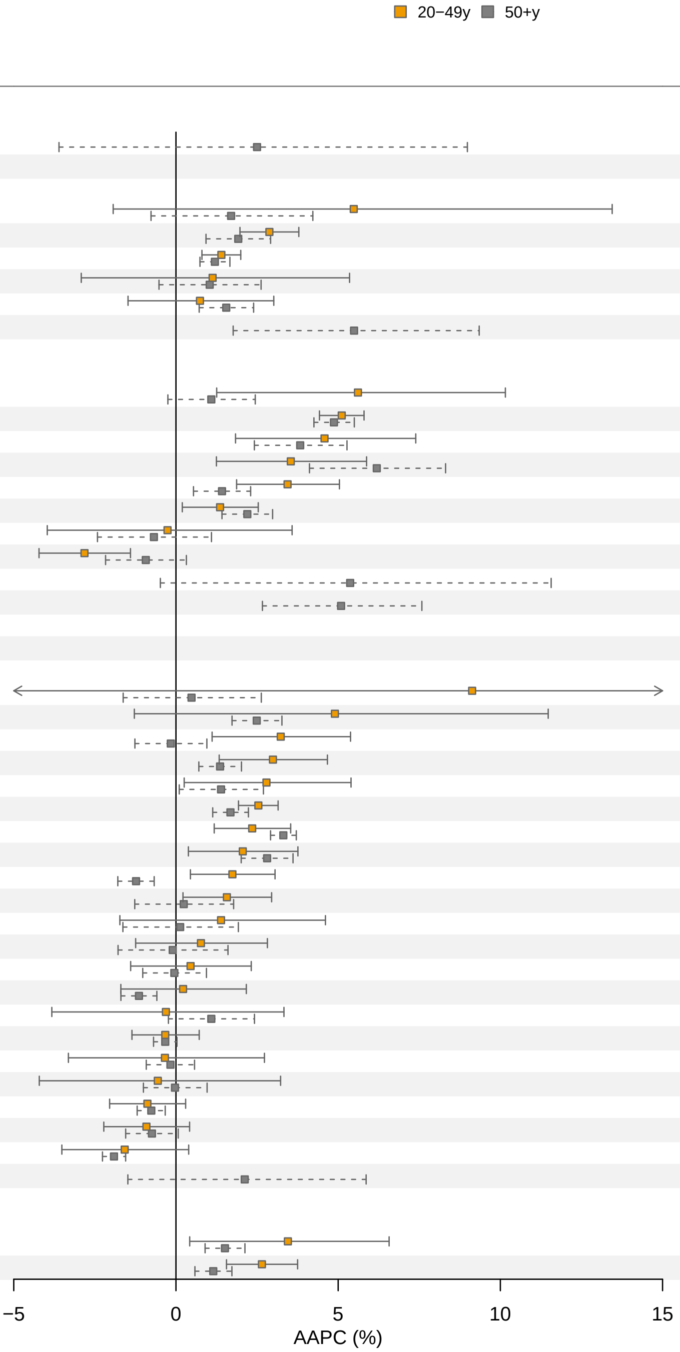

| Country | AAPC (%) 20−49y | AAPC (%) 50+y | p value |
|---|---|---|---|
| **Africa** | | | |
| Uganda | − | 2.5 | − |
| **Americas** | | | |
| Ecuador | 5.48 | 1.7 | 0.33 |
| Canada | 2.88 | 1.92 ‡ | 0.158 |
| USA | 1.4 | 1.2 ‡ | 0.60 |
| Argentina | 1.13 | 1.04 | 0.96 |
| Colombia | 0.74 | 1.55 | 0.49 |
| Chile | − | 5.49 | − |
| **Asia** | | | |
| Cyprus | 5.61 | 1.09 | 0.051 |
| Republic of Korea | 5.11 | 4.87 | 0.60 |
| Thailand | 4.58 | 3.83 | 0.62 |
| India | 3.54 | 6.19 ‡ | 0.093 |
| China | 3.44 | 1.42 | 0.029 |
| Türkiye | 1.36 | 2.2 | 0.23 |
| Philippines | −0.26 ‡ | −0.68 ‡ | 0.85 |
| Israel | −2.82 | −0.93 ‡ | 0.048 |
| Bahrain | − | 5.37 | − |
| Kuwait | − | 5.09 | − |
| Qatar | − | − | − |
| **Europe** | | | |
| Malta | 9.13 ‡ | 0.48 | 0.48 |
| Croatia | 4.9 ‡ | 2.49 | 0.46 |
| Denmark | 3.23 | −0.16 ‡ | 0.008 |
| Lithuania | 2.99 | 1.36 | 0.070 |
| Estonia | 2.79 | 1.39 ‡ | 0.32 |
| UK | 2.54 | 1.68 ‡ | 0.041 |
| Belarus | 2.35 | 3.31 | 0.121 |
| Ireland | 2.06 | 2.81 | 0.41 |
| Sweden | 1.74 | −1.23 ‡ | <0.001 |
| Germany | 1.57 | 0.24 ‡ | 0.197 |
| Switzerland | 1.39 | 0.13 ‡ | 0.48 |
| Slovenia | 0.77 | −0.1 ‡ | 0.51 |
| Italy | 0.45 | −0.05 | 0.63 |
| Finland | 0.22 | −1.14 | 0.167 |
| Poland | −0.31 | 1.09 ‡ | 0.45 |
| Czechia | −0.33 | −0.33 | 0.99 |
| Latvia | −0.34 | −0.17 | 0.91 |
| France | −0.56 ‡ | −0.03 ‡ | 0.78 |
| The Netherlands | −0.88 | −0.76 | 0.85 |
| Norway | −0.91 | −0.74 | 0.82 |
| Austria | −1.58 | −1.91 | 0.73 |
| Iceland | − | 2.12 | − |
| **Oceania** | | | |
| New Zealand | 3.45 ‡ | 1.51 | 0.22 |
| Australia | 2.65 | 1.15 ‡ | 0.019 |

Data for female sex only.
‡ best fitted Joinpoint model > 0 joins; * p−value for a difference between AAPC 20−49y and AAPC 50+y.

#### Appendix Figure 17: Average annual percent change (AAPC) for cancer incidence (2003-2017) by UN region, country and age - Gallbladder Cancer

■ 20−49y ■ 50+y

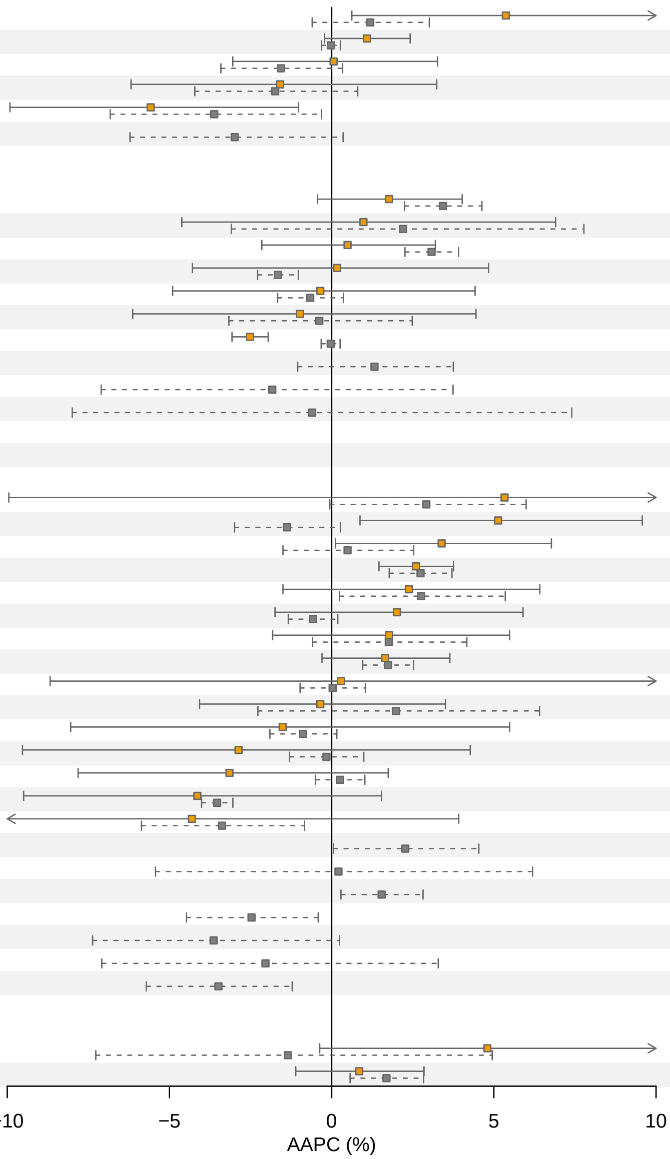

| Country | AAPC (%) 20−49y | AAPC (%) 50+y | p value |
|---|---|---|---|
| **Africa** | | | |
| Uganda | – | – | – |
| **Americas** | | | |
| Canada | 5.37 ‡ | 1.19 ‡ | 0.109 |
| USA | 1.09 | −0.02 | 0.097 |
| Colombia | 0.06 | −1.56 | 0.37 |
| Ecuador | −1.59 | −1.74 | 0.95 |
| Chile | −5.58 | −3.62 ‡ | 0.47 |
| Argentina | – | −2.99 ‡ | – |
| **Asia** | | | |
| India | 1.77 | 3.43 | 0.184 |
| Philippines | 0.98 | 2.2 ‡ | 0.76 |
| Thailand | 0.49 | 3.08 | 0.069 |
| China | 0.17 ‡ | −1.66 | 0.43 |
| Türkiye | −0.35 | −0.66 | 0.89 |
| Israel | −0.98 | −0.38 ‡ | 0.84 |
| Republic of Korea | −2.52 | −0.03 | <0.001 |
| Cyprus | – | 1.32 | – |
| Kuwait | – | −1.83 | – |
| Qatar | – | −0.6 | – |
| Bahrain | – | – | – |
| **Europe** | | | |
| Slovenia | 5.33 ‡ | 2.92 ‡ | 0.78 |
| Switzerland | 5.13 | −1.38 | 0.008 |
| Sweden | 3.39 | 0.49 ‡ | 0.133 |
| UK | 2.6 | 2.74 ‡ | 0.86 |
| Belarus | 2.38 | 2.76 ‡ | 0.87 |
| Austria | 2.01 | −0.58 | 0.172 |
| Denmark | 1.77 | 1.76 ‡ | 0.99 |
| The Netherlands | 1.65 | 1.74 | 0.93 |
| Finland | 0.29 ‡ | 0.03 | 0.96 |
| Norway | −0.35 | 1.98 ‡ | 0.42 |
| Croatia | −1.51 | −0.88 | 0.85 |
| Germany | −2.87 ‡ | −0.16 | 0.45 |
| France | −3.15 | 0.26 | 0.158 |
| Czechia | −4.14 ‡ | −3.53 | 0.83 |
| Italy | −4.31 ‡ | −3.38 ‡ | 0.83 |
| Estonia | – | 2.27 | – |
| Iceland | – | 0.21 | – |
| Ireland | – | 1.54 | – |
| Latvia | – | −2.47 | – |
| Lithuania | – | −3.64 ‡ | – |
| Malta | – | −2.04 | – |
| Poland | – | −3.49 | – |
| **Oceania** | | | |
| New Zealand | 4.8 | −1.35 ‡ | 0.134 |
| Australia | 0.85 | 1.69 ‡ | 0.45 |

‡ best fitted Joinpoint model > 0 joins; * p−value for a difference between AAPC 20−49y and AAPC 50+y.

#### Appendix Figure 18: Average annual percent change (AAPC) for cancer incidence (2003-2017) by UN region, country and age - Kidney Cancer

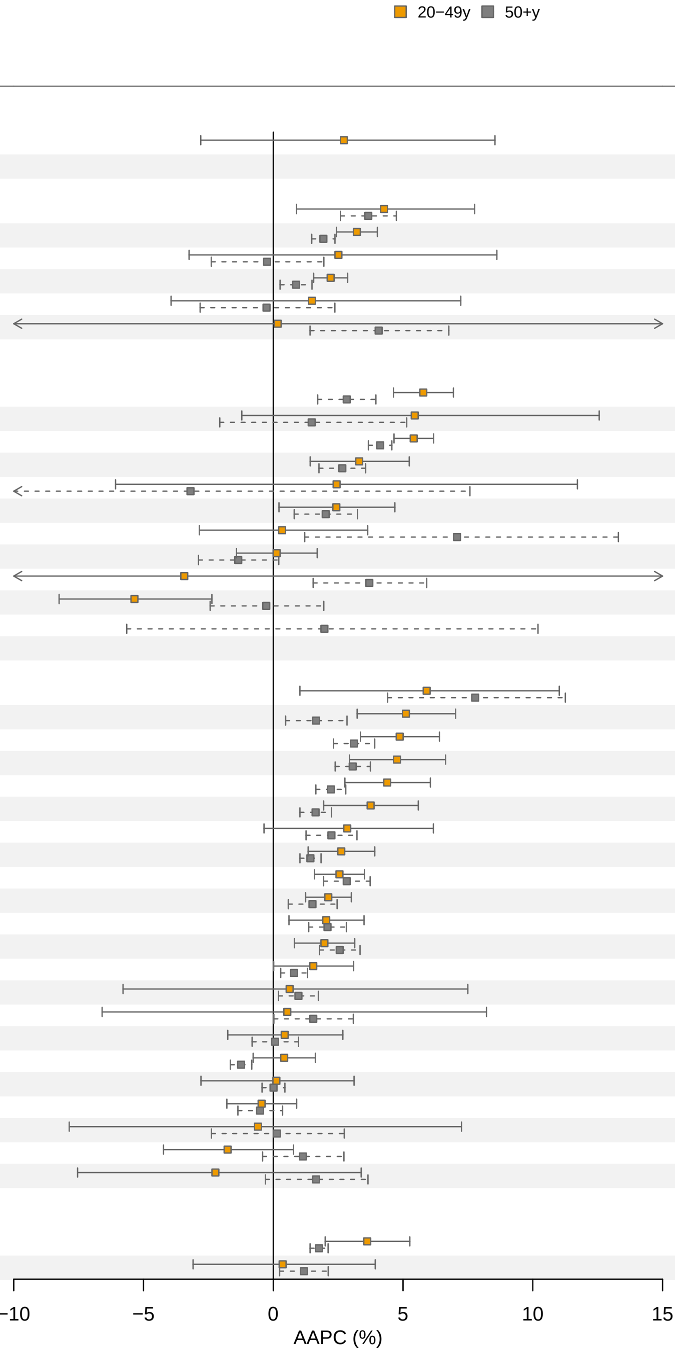

| Country | AAPC (%) 20−49y | AAPC (%) 50+y | p value |
|---|---|---|---|
| **Africa** | | | |
| Uganda | 2.72 | – | – |
| **Americas** | | | |
| Colombia | 4.27 | 3.66 | 0.72 |
| Canada | 3.22 | 1.93 | 0.007 |
| Chile | 2.51 | −0.24 | 0.36 |
| USA | 2.21 | 0.88 ‡ | 0.005 |
| Argentina | 1.49 ‡ | −0.26 ‡ | 0.58 |
| Ecuador | 0.17 ‡ | 4.06 | 0.68 |
| **Asia** | | | |
| Türkiye | 5.78 | 2.83 | <0.001 |
| Kuwait | 5.45 | 1.48 | 0.29 |
| Republic of Korea | 5.41 | 4.12 ‡ | 0.006 |
| India | 3.31 | 2.66 | 0.52 |
| Bahrain | 2.44 | −3.19 ‡ | 0.41 |
| China | 2.43 | 2.02 ‡ | 0.74 |
| Thailand | 0.34 | 7.08 ‡ | 0.049 |
| Israel | 0.13 | −1.35 ‡ | 0.182 |
| Cyprus | −3.43 ‡ | 3.7 | 0.53 |
| Philippines | −5.35 | −0.27 ‡ | 0.009 |
| Qatar | – | 1.97 | – |
| **Europe** | | | |
| Malta | 5.91 | 7.78 ‡ | 0.53 |
| Ireland | 5.11 | 1.65 ‡ | 0.004 |
| UK | 4.87 ‡ | 3.11 ‡ | 0.044 |
| Denmark | 4.77 | 3.06 | 0.082 |
| Norway | 4.39 | 2.22 | 0.017 |
| Sweden | 3.75 | 1.63 | 0.031 |
| Latvia | 2.85 | 2.24 | 0.70 |
| France | 2.62 | 1.43 | 0.078 |
| Belarus | 2.55 | 2.83 ‡ | 0.66 |
| The Netherlands | 2.12 | 1.51 ‡ | 0.35 |
| Italy | 2.04 | 2.09 | 0.95 |
| Croatia | 1.97 | 2.56 | 0.39 |
| Finland | 1.54 | 0.8 | 0.34 |
| Switzerland | 0.63 ‡ | 0.97 | 0.92 |
| Slovenia | 0.54 ‡ | 1.54 ‡ | 0.80 |
| Lithuania | 0.44 | 0.07 | 0.75 |
| Austria | 0.42 | −1.24 | 0.013 |
| Germany | 0.12 ‡ | 0.01 | 0.94 |
| Czechia | −0.45 ‡ | −0.51 ‡ | 0.95 |
| Iceland | −0.59 | 0.14 | 0.85 |
| Poland | −1.76 | 1.14 | 0.054 |
| Estonia | −2.23 ‡ | 1.65 ‡ | 0.197 |
| **Oceania** | | | |
| Australia | 3.62 ‡ | 1.76 | 0.028 |
| New Zealand | 0.36 ‡ | 1.18 | 0.66 |

‡ best fitted Joinpoint model > 0 joins; * p−value for a difference between AAPC 20−49y and AAPC 50+y.

### Appendix Figure 19: Average annual percent change (AAPC) for cancer incidence (2003-2017) by UN region, country and age - Leukaemia

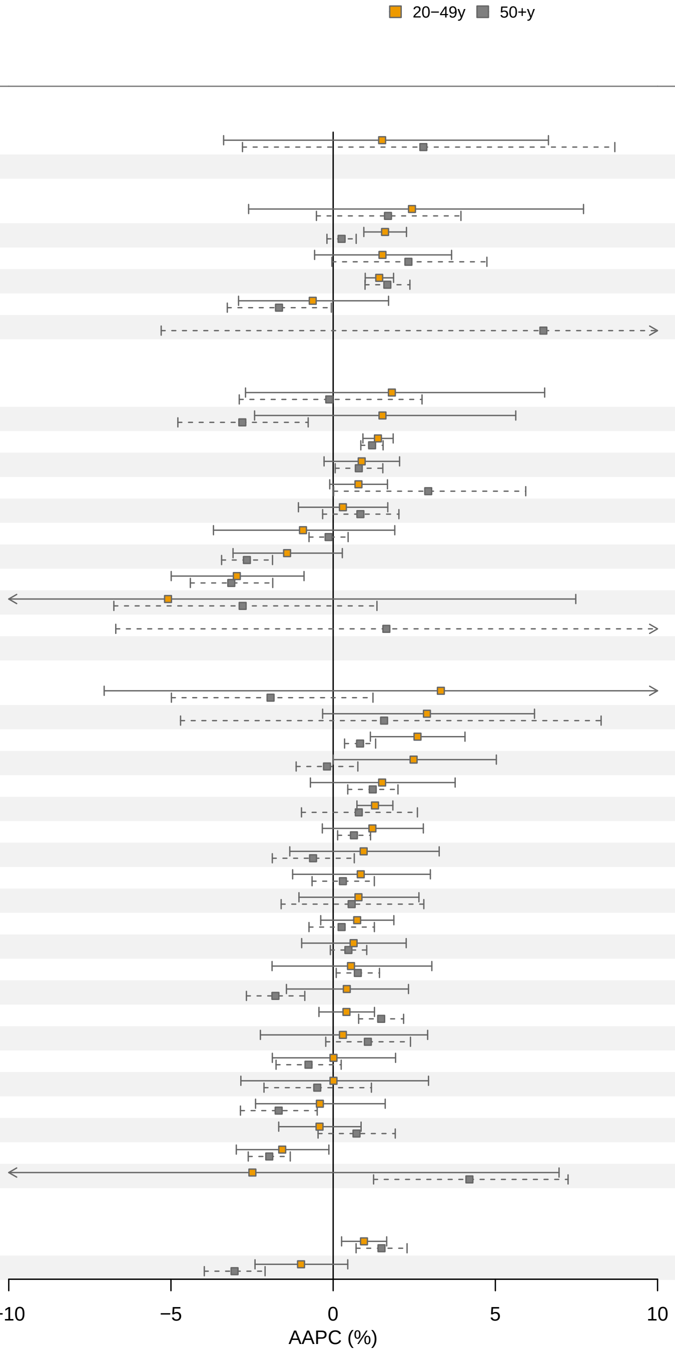

| Country | AAPC (%) 20−49y | AAPC (%) 50+y | p value |
|---|---|---|---|
| **Africa** | | | |
| Uganda | 1.51 | 2.78 | 0.73 |
| **Americas** | | | |
| Colombia | 2.43 ‡ | 1.69 | 0.80 |
| USA | 1.6 | 0.26 ‡ | 0.002 |
| Ecuador | 1.52 | 2.32 | 0.60 |
| Canada | 1.42 | 1.67 | 0.53 |
| Argentina | −0.63 | −1.67 | 0.45 |
| Chile | – | 6.48 ‡ | – |
| **Asia** | | | |
| Kuwait | 1.81 | −0.12 | 0.46 |
| Cyprus | 1.52 | −2.8 | 0.055 |
| Republic of Korea | 1.38 | 1.2 | 0.51 |
| Türkiye | 0.88 | 0.79 | 0.90 |
| India | 0.78 | 2.93 ‡ | 0.168 |
| Thailand | 0.3 | 0.84 | 0.53 |
| China | −0.93 ‡ | −0.14 | 0.59 |
| Israel | −1.42 | −2.66 | 0.174 |
| Philippines | −2.97 ‡ | −3.14 | 0.89 |
| Bahrain | −5.09 ‡ | −2.79 | 0.72 |
| Qatar | – | 1.64 | – |
| **Europe** | | | |
| Iceland | 3.32 ‡ | −1.93 | 0.35 |
| Latvia | 2.89 | 1.57 ‡ | 0.72 |
| France | 2.6 | 0.83 | 0.026 |
| Estonia | 2.48 | −0.19 | 0.049 |
| Norway | 1.51 | 1.22 | 0.80 |
| UK | 1.29 | 0.79 ‡ | 0.61 |
| Denmark | 1.21 | 0.64 | 0.47 |
| Poland | 0.94 | −0.62 | 0.22 |
| Finland | 0.85 | 0.3 | 0.63 |
| Lithuania | 0.78 | 0.57 ‡ | 0.89 |
| Czechia | 0.74 | 0.26 ‡ | 0.52 |
| Sweden | 0.63 | 0.47 | 0.85 |
| Germany | 0.55 ‡ | 0.76 | 0.87 |
| Ireland | 0.42 | −1.78 | 0.038 |
| The Netherlands | 0.41 | 1.48 ‡ | 0.058 |
| Slovenia | 0.3 | 1.07 ‡ | 0.59 |
| Switzerland | 0.01 | −0.76 | 0.46 |
| Croatia | 0.01 ‡ | −0.49 ‡ | 0.77 |
| Italy | −0.41 | −1.68 | 0.26 |
| Belarus | −0.42 | 0.72 | 0.188 |
| Austria | −1.57 | −1.97 | 0.59 |
| Malta | −2.49 ‡ | 4.2 | 0.179 |
| **Oceania** | | | |
| Australia | 0.95 | 1.49 ‡ | 0.31 |
| New Zealand | −0.99 | −3.04 ‡ | 0.020 |

‡ best fitted Joinpoint model > 0 joins; * p−value for a difference between AAPC 20−49y and AAPC 50+y.

#### Appendix Figure 20: Average annual percent change (AAPC) for cancer incidence (2003-2017) by UN region, country and age - Liver Cancer

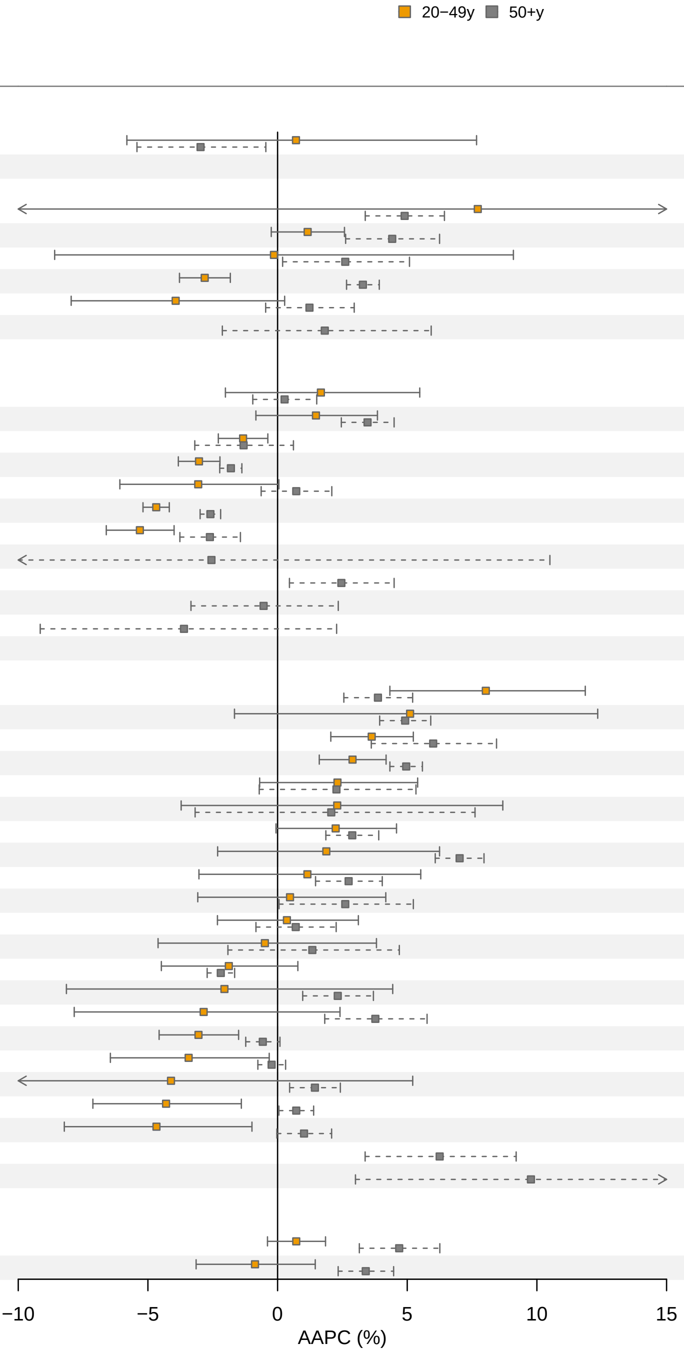

| Country | AAPC (%) 20−49y | AAPC (%) 50+y | p value |
|---|---|---|---|
| **Africa** | | | |
| Uganda | 0.71 ‡ | −2.97 | 0.31 |
| **Americas** | | | |
| Argentina | 7.72 ‡ | 4.9 | 0.80 |
| Canada | 1.16 | 4.42 ‡ | 0.006 |
| Ecuador | −0.14 | 2.61 ‡ | 0.54 |
| USA | −2.81 | 3.29 ‡ | <0.001 |
| Colombia | −3.93 | 1.23 | 0.028 |
| Chile | − | 1.82 | − |
| **Asia** | | | |
| Israel | 1.67 | 0.27 | 0.46 |
| India | 1.48 | 3.47 | 0.120 |
| Thailand | −1.33 | −1.31 ‡ | 0.98 |
| China | −3.03 | −1.8 | 0.011 |
| Türkiye | −3.06 | 0.72 | 0.031 |
| Republic of Korea | −4.68 ‡ | −2.59 ‡ | <0.001 |
| Philippines | −5.31 | −2.61 | 0.004 |
| Bahrain | − | −2.55 ‡ | − |
| Cyprus | − | 2.46 | − |
| Kuwait | − | −0.54 | − |
| Qatar | − | −3.61 | − |
| **Europe** | | | |
| Lithuania | 8.03 | 3.87 | 0.039 |
| Ireland | 5.11 ‡ | 4.92 | 0.96 |
| The Netherlands | 3.63 | 6 ‡ | 0.105 |
| UK | 2.89 | 4.96 ‡ | 0.007 |
| Sweden | 2.31 | 2.27 ‡ | 0.99 |
| Poland | 2.3 | 2.07 ‡ | 0.96 |
| Belarus | 2.24 | 2.88 | 0.60 |
| Norway | 1.88 | 7.02 | 0.027 |
| Latvia | 1.15 | 2.74 | 0.46 |
| Finland | 0.48 | 2.61 ‡ | 0.34 |
| Germany | 0.36 | 0.7 ‡ | 0.82 |
| Denmark | −0.49 | 1.34 ‡ | 0.49 |
| Italy | −1.88 | −2.19 | 0.81 |
| Slovenia | −2.05 | 2.32 | 0.174 |
| Estonia | −2.85 | 3.77 | 0.024 |
| Austria | −3.05 | −0.57 | 0.006 |
| Czechia | −3.43 | −0.23 | 0.047 |
| France | −4.11 ‡ | 1.44 ‡ | 0.24 |
| Croatia | −4.3 | 0.72 | 0.003 |
| Switzerland | −4.67 | 1.02 | 0.007 |
| Iceland | − | 6.25 | − |
| Malta | − | 9.77 | − |
| **Oceania** | | | |
| Australia | 0.72 | 4.69 ‡ | <0.001 |
| New Zealand | −0.87 | 3.4 | 0.003 |

‡ best fitted Joinpoint model > 0 joins; * p−value for a difference between AAPC 20−49y and AAPC 50+y.

#### Appendix Figure 21: Average annual percent change (AAPC) for cancer incidence (2003-2017) by UN region, country and age - Oesophagal Cancer

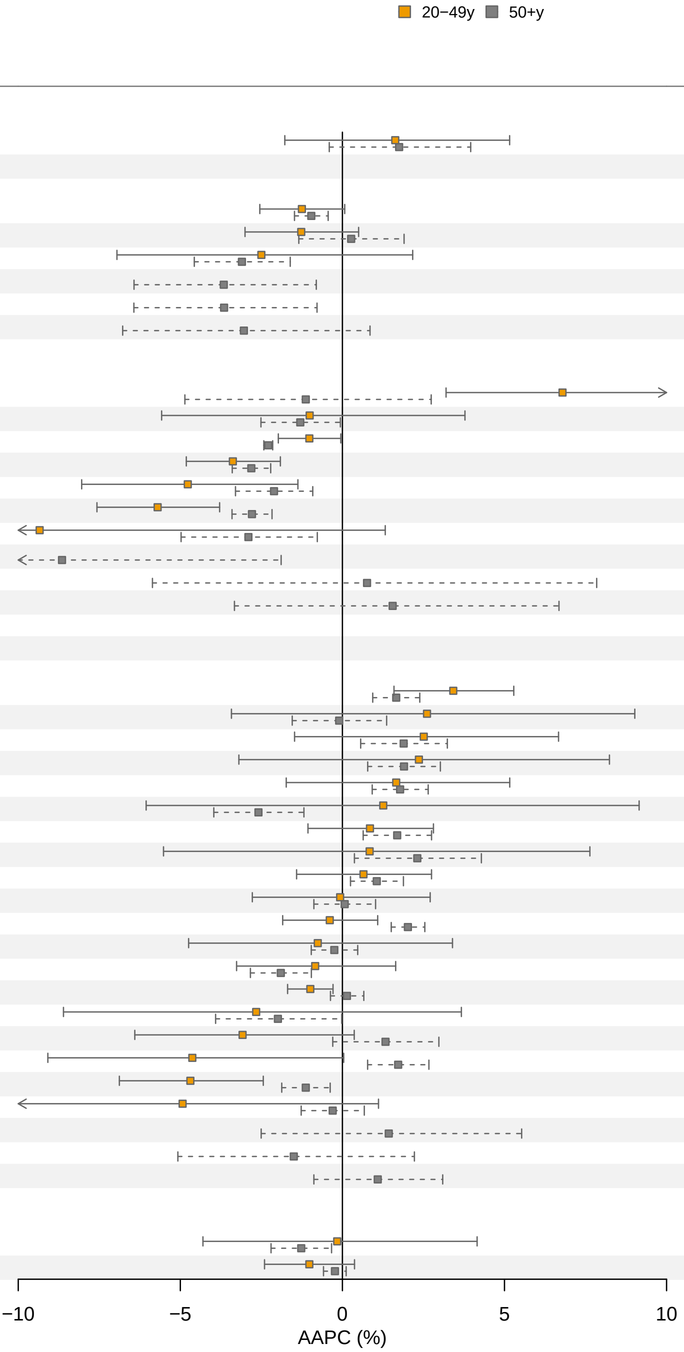

| Country | AAPC (%) 20−49y | AAPC (%) 50+y | p value |
|---|---|---|---|
| **Africa** | | | |
| Uganda | 1.63 | 1.75 | 0.95 |
| **Americas** | | | |
| USA | −1.25 | −0.96 | 0.67 |
| Canada | −1.27 | 0.27 ‡ | 0.20 |
| Argentina | −2.5 | −3.1 | 0.79 |
| Chile | – | −3.66 | – |
| Colombia | – | −3.65 ‡ | – |
| Ecuador | – | −3.04 | – |
| **Asia** | | | |
| Thailand | 6.79 | −1.13 ‡ | 0.004 |
| Israel | −1.01 | −1.3 | 0.90 |
| Republic of Korea | −1.02 | −2.29 | 0.014 |
| India | −3.38 | −2.81 | 0.45 |
| Türkiye | −4.77 | −2.11 | 0.135 |
| China | −5.7 | −2.79 | 0.008 |
| Philippines | −9.34 ‡ | −2.9 | 0.23 |
| Bahrain | – | −8.65 | – |
| Cyprus | – | 0.76 | – |
| Kuwait | – | 1.55 | – |
| Qatar | – | – | – |
| **Europe** | | | |
| Belarus | 3.42 | 1.66 | 0.075 |
| Ireland | 2.61 ‡ | −0.1 ‡ | 0.40 |
| Latvia | 2.51 | 1.89 | 0.76 |
| Estonia | 2.36 | 1.9 | 0.86 |
| Norway | 1.66 | 1.78 | 0.94 |
| Italy | 1.26 | −2.59 | 0.29 |
| Lithuania | 0.85 | 1.69 | 0.44 |
| Finland | 0.84 | 2.31 ‡ | 0.66 |
| Germany | 0.65 | 1.06 | 0.70 |
| Sweden | −0.07 | 0.07 | 0.92 |
| The Netherlands | −0.39 | 2.02 | 0.005 |
| Austria | −0.76 | −0.25 | 0.79 |
| Croatia | −0.84 | −1.9 | 0.40 |
| UK | −0.99 | 0.14 ‡ | 0.012 |
| Slovenia | −2.66 | −1.99 | 0.83 |
| Czechia | −3.08 | 1.33 ‡ | 0.026 |
| Denmark | −4.63 | 1.72 | 0.013 |
| France | −4.69 | −1.13 | 0.006 |
| Switzerland | −4.93 | −0.3 | 0.124 |
| Iceland | – | 1.43 | – |
| Malta | – | −1.5 | – |
| Poland | – | 1.09 | – |
| **Oceania** | | | |
| New Zealand | −0.16 | −1.27 | 0.59 |
| Australia | −1.02 | −0.23 | 0.26 |

‡ best fitted Joinpoint model > 0 joins; * p−value for a difference between AAPC 20−49y and AAPC 50+y.

#### Appendix Figure 22: Average annual percent change (AAPC) for cancer incidence (2003-2017) by UN region, country and age - Oral Cancer

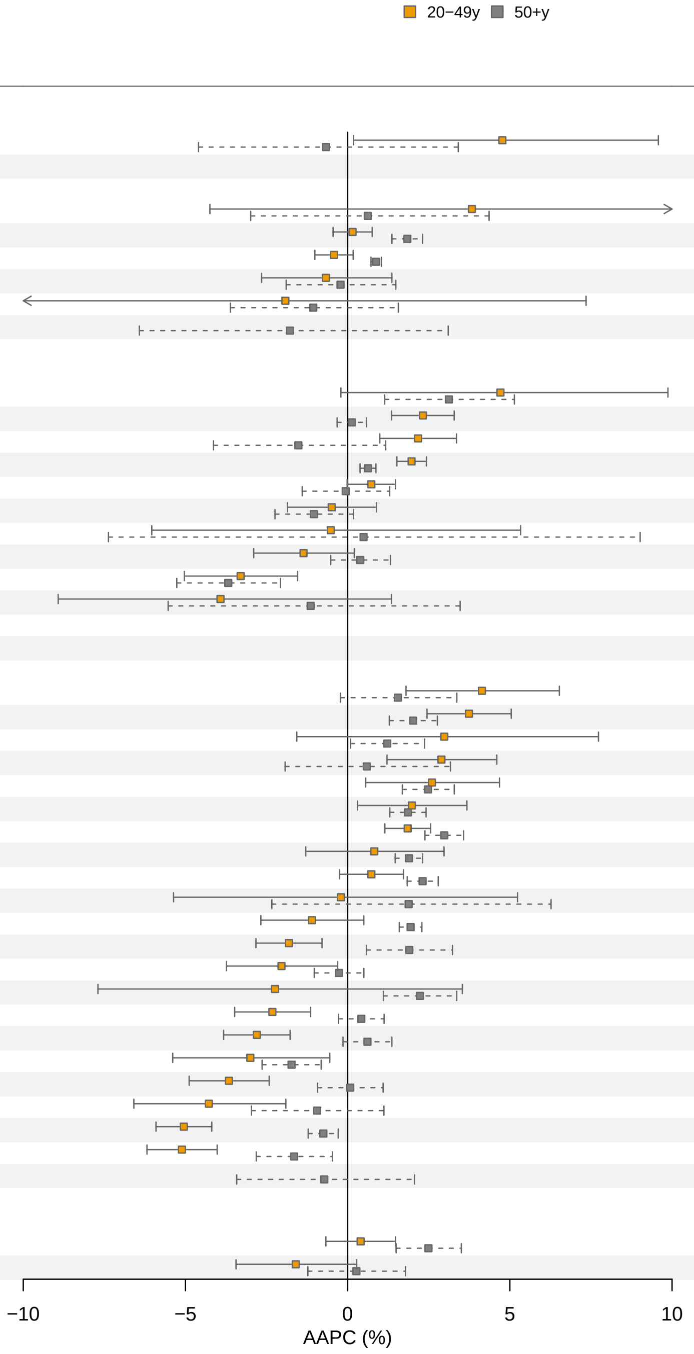

| Country | AAPC (%) 20−49y | AAPC (%) 50+y | p value |
|---|---|---|---|
| **Africa** | | | |
| Uganda | 4.77 | −0.67 | 0.078 |
| **Americas** | | | |
| Ecuador | 3.83 ‡ | 0.62 | 0.49 |
| Canada | 0.15 | 1.84 | <0.001 |
| USA | −0.42 | 0.88 | <0.001 |
| Colombia | −0.67 | −0.22 ‡ | 0.73 |
| Argentina | −1.92 ‡ | −1.06 | 0.86 |
| Chile | – | −1.78 | – |
| **Asia** | | | |
| Cyprus | 4.71 | 3.12 | 0.54 |
| India | 2.32 | 0.13 | <0.001 |
| Thailand | 2.17 | −1.52 ‡ | 0.014 |
| Republic of Korea | 1.97 | 0.63 | <0.001 |
| China | 0.73 | −0.06 ‡ | 0.31 |
| Türkiye | −0.49 | −1.04 | 0.54 |
| Bahrain | −0.52 | 0.49 | 0.84 |
| Israel | −1.36 | 0.39 | 0.056 |
| Philippines | −3.3 | −3.68 | 0.74 |
| Kuwait | −3.92 | −1.14 ‡ | 0.41 |
| Qatar | – | – | – |
| **Europe** | | | |
| Latvia | 4.14 | 1.55 ‡ | 0.084 |
| Belarus | 3.74 ‡ | 2.02 ‡ | 0.024 |
| Estonia | 2.98 ‡ | 1.22 | 0.47 |
| Lithuania | 2.89 ‡ | 0.59 ‡ | 0.142 |
| Ireland | 2.6 | 2.48 | 0.91 |
| Norway | 1.98 | 1.86 | 0.89 |
| UK | 1.85 ‡ | 2.98 ‡ | 0.017 |
| Finland | 0.82 | 1.89 | 0.31 |
| Sweden | 0.73 | 2.31 | 0.007 |
| Malta | −0.21 | 1.88 ‡ | 0.54 |
| Czechia | −1.1 ‡ | 1.94 | <0.001 |
| Denmark | −1.81 | 1.9 ‡ | <0.001 |
| Slovenia | −2.04 | −0.27 | 0.064 |
| Poland | −2.24 ‡ | 2.23 | 0.134 |
| The Netherlands | −2.32 | 0.42 ‡ | <0.001 |
| Austria | −2.8 | 0.61 | <0.001 |
| Italy | −3 | −1.73 | 0.31 |
| Germany | −3.66 ‡ | 0.08 ‡ | <0.001 |
| Switzerland | −4.28 | −0.94 ‡ | 0.036 |
| France | −5.05 | −0.75 | <0.001 |
| Croatia | −5.11 | −1.65 ‡ | <0.001 |
| Iceland | – | −0.72 | – |
| **Oceania** | | | |
| New Zealand | 0.4 | 2.49 | 0.007 |
| Australia | −1.6 ‡ | 0.27 ‡ | 0.127 |

‡ best fitted Joinpoint model > 0 joins; * p−value for a difference between AAPC 20−49y and AAPC 50+y.

#### Appendix Figure 23: Average annual percent change (AAPC) for cancer incidence (2003-2017) by UN region, country and age - Pancreatic Cancer

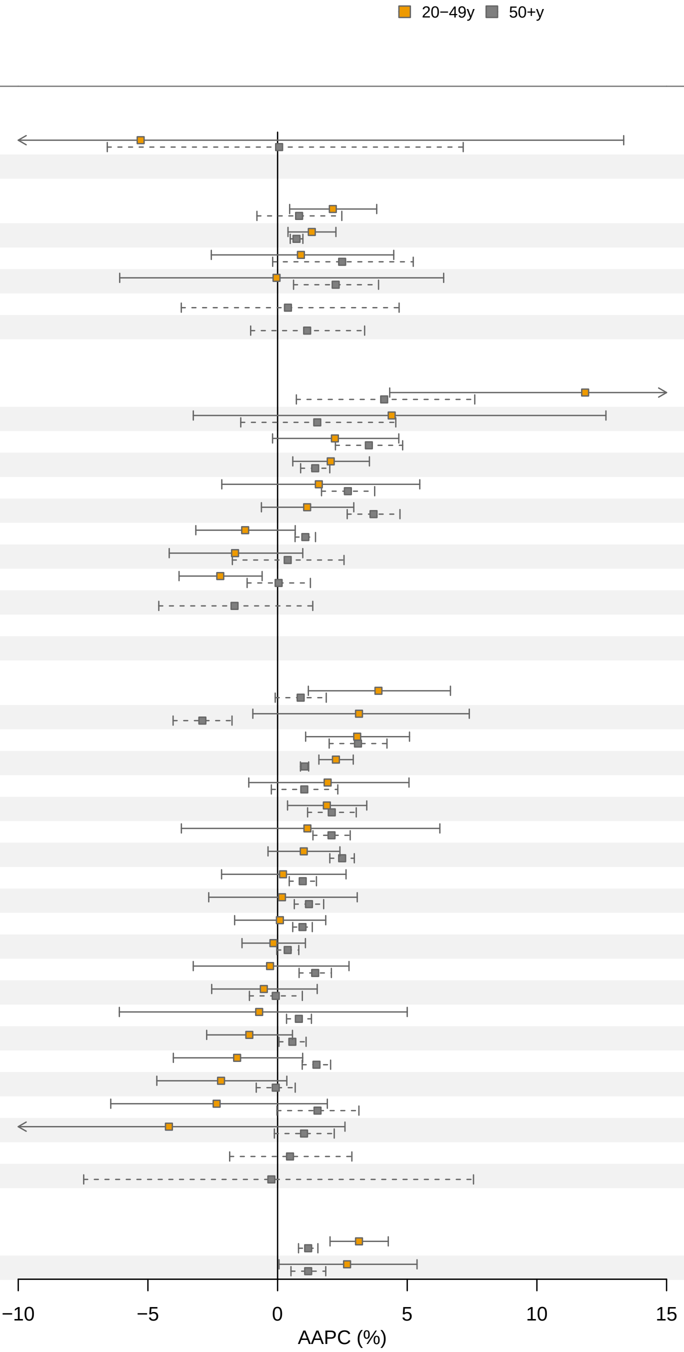

| Country | AAPC (%) 20−49y | AAPC (%) 50+y | p value |
|---|---|---|---|
| **Africa** | | | |
| Uganda | −5.28 ‡ | 0.06 | 0.57 |
| **Americas** | | | |
| Canada | 2.13 ‡ | 0.83 ‡ | 0.28 |
| USA | 1.32 | 0.73 | 0.21 |
| Argentina | 0.9 | 2.49 ‡ | 0.47 |
| Colombia | −0.04 ‡ | 2.24 ‡ | 0.49 |
| Chile | – | 0.4 | – |
| Ecuador | – | 1.14 | – |
| **Asia** | | | |
| Cyprus | 11.86 | 4.11 ‡ | 0.066 |
| Kuwait | 4.4 | 1.53 | 0.48 |
| Thailand | 2.21 | 3.52 | 0.33 |
| Republic of Korea | 2.05 ‡ | 1.45 ‡ | 0.45 |
| Türkiye | 1.59 ‡ | 2.71 | 0.58 |
| India | 1.14 | 3.7 | 0.018 |
| China | −1.25 | 1.07 | 0.024 |
| Philippines | −1.64 | 0.39 ‡ | 0.23 |
| Israel | −2.21 | 0.04 ‡ | 0.030 |
| Bahrain | – | −1.66 | – |
| Qatar | – | – | – |
| **Europe** | | | |
| Switzerland | 3.89 | 0.89 ‡ | 0.042 |
| Poland | 3.14 | −2.9 | 0.008 |
| France | 3.07 | 3.1 ‡ | 0.98 |
| UK | 2.25 | 1.04 | 0.002 |
| Ireland | 1.93 | 1.03 ‡ | 0.58 |
| The Netherlands | 1.9 | 2.09 ‡ | 0.83 |
| Sweden | 1.15 ‡ | 2.08 ‡ | 0.72 |
| Belarus | 1.01 | 2.49 | 0.049 |
| Lithuania | 0.21 | 0.97 | 0.52 |
| Italy | 0.17 | 1.21 | 0.46 |
| Austria | 0.09 | 0.96 | 0.32 |
| Czechia | −0.16 | 0.39 | 0.38 |
| Latvia | −0.29 | 1.45 | 0.25 |
| Denmark | −0.53 | −0.07 ‡ | 0.68 |
| Germany | −0.71 ‡ | 0.82 | 0.59 |
| Finland | −1.09 | 0.57 | 0.059 |
| Croatia | −1.56 | 1.5 | 0.024 |
| Norway | −2.18 | −0.07 | 0.108 |
| Slovenia | −2.35 | 1.54 ‡ | 0.087 |
| Estonia | −4.19 ‡ | 1.02 | 0.134 |
| Iceland | – | 0.48 | – |
| Malta | – | −0.24 ‡ | – |
| **Oceania** | | | |
| Australia | 3.14 | 1.18 | 0.003 |
| New Zealand | 2.68 | 1.18 | 0.26 |

‡ best fitted Joinpoint model > 0 joins; * p−value for a difference between AAPC 20−49y and AAPC 50+y.

#### Appendix Figure 24: Average annual percent change (AAPC) for cancer incidence (2003-2017) by UN region, country and age - Prostate Cancer

■ 20−49y ■ 50+y

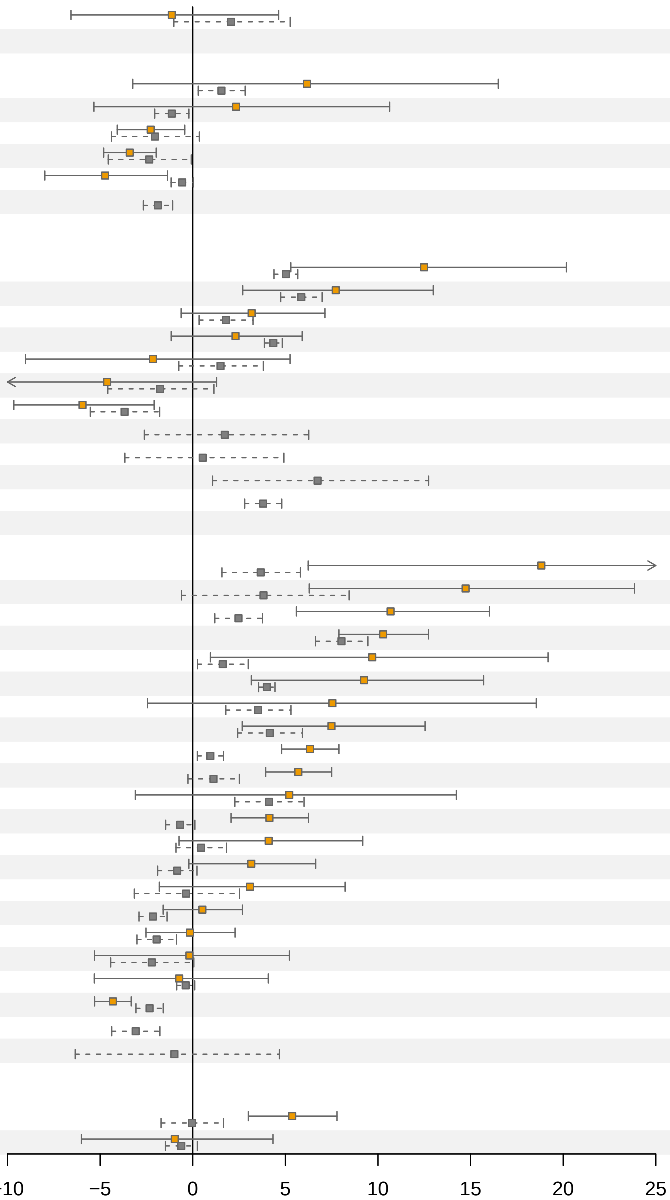

| Country | AAPC (%) 20−49y | AAPC (%) 50+y | p value |
|---|---|---|---|
| **Africa** | | | |
| Uganda | −1.13 | 2.07 ‡ | 0.32 |
| **Americas** | | | |
| Ecuador | 6.17 ‡ | 1.55 | 0.35 |
| Argentina | 2.34 | −1.13 | 0.36 |
| Canada | −2.27 ‡ | −2.04 ‡ | 0.88 |
| USA | −3.4 ‡ | −2.35 ‡ | 0.43 |
| Colombia | −4.73 | −0.57 | 0.021 |
| Chile | − | −1.88 | − |
| **Asia** | | | |
| China | 12.49 | 5.03 | 0.044 |
| Republic of Korea | 7.72 ‡ | 5.86 ‡ | 0.49 |
| Türkiye | 3.18 | 1.79 ‡ | 0.48 |
| India | 2.31 | 4.35 | 0.24 |
| Cyprus | −2.15 | 1.5 ‡ | 0.32 |
| Philippines | −4.62 | −1.76 ‡ | 0.37 |
| Israel | −5.95 | −3.68 ‡ | 0.28 |
| Bahrain | − | 1.73 | − |
| Kuwait | − | 0.54 ‡ | − |
| Qatar | − | 6.74 | − |
| Thailand | − | 3.8 | − |
| **Europe** | | | |
| Estonia | 18.82 ‡ | 3.67 ‡ | 0.019 |
| Lithuania | 14.73 ‡ | 3.82 ‡ | 0.026 |
| Czechia | 10.68 ‡ | 2.47 ‡ | 0.002 |
| Belarus | 10.28 | 8.03 ‡ | 0.107 |
| Denmark | 9.69 ‡ | 1.62 ‡ | 0.075 |
| Latvia | 9.25 | 4 | 0.087 |
| Slovenia | 7.54 ‡ | 3.53 ‡ | 0.45 |
| Croatia | 7.49 | 4.16 ‡ | 0.190 |
| UK | 6.33 ‡ | 0.95 ‡ | <0.001 |
| Norway | 5.7 | 1.12 ‡ | <0.001 |
| Poland | 5.21 | 4.12 ‡ | 0.80 |
| Sweden | 4.14 | −0.68 | <0.001 |
| Ireland | 4.1 ‡ | 0.45 ‡ | 0.158 |
| The Netherlands | 3.16 ‡ | −0.84 ‡ | 0.026 |
| Switzerland | 3.09 ‡ | −0.36 ‡ | 0.24 |
| Germany | 0.52 | −2.15 | 0.023 |
| Finland | −0.15 | −1.95 | 0.163 |
| France | −0.18 ‡ | −2.21 ‡ | 0.48 |
| Italy | −0.73 | −0.38 | 0.88 |
| Austria | −4.31 | −2.33 ‡ | 0.003 |
| Iceland | − | −3.08 | − |
| Malta | − | −0.99 ‡ | − |
| **Oceania** | | | |
| Australia | 5.37 ‡ | −0.04 ‡ | <0.001 |
| New Zealand | −0.97 ‡ | −0.62 | 0.89 |

Data for male sex only.
‡ best fitted Joinpoint model > 0 joins; * p−value for a difference between AAPC 20−49y and AAPC 50+y.

#### Appendix Figure 25: Average annual percent change (AAPC) for cancer incidence (2003-2017) by UN region, country and age - Stomach Cancer

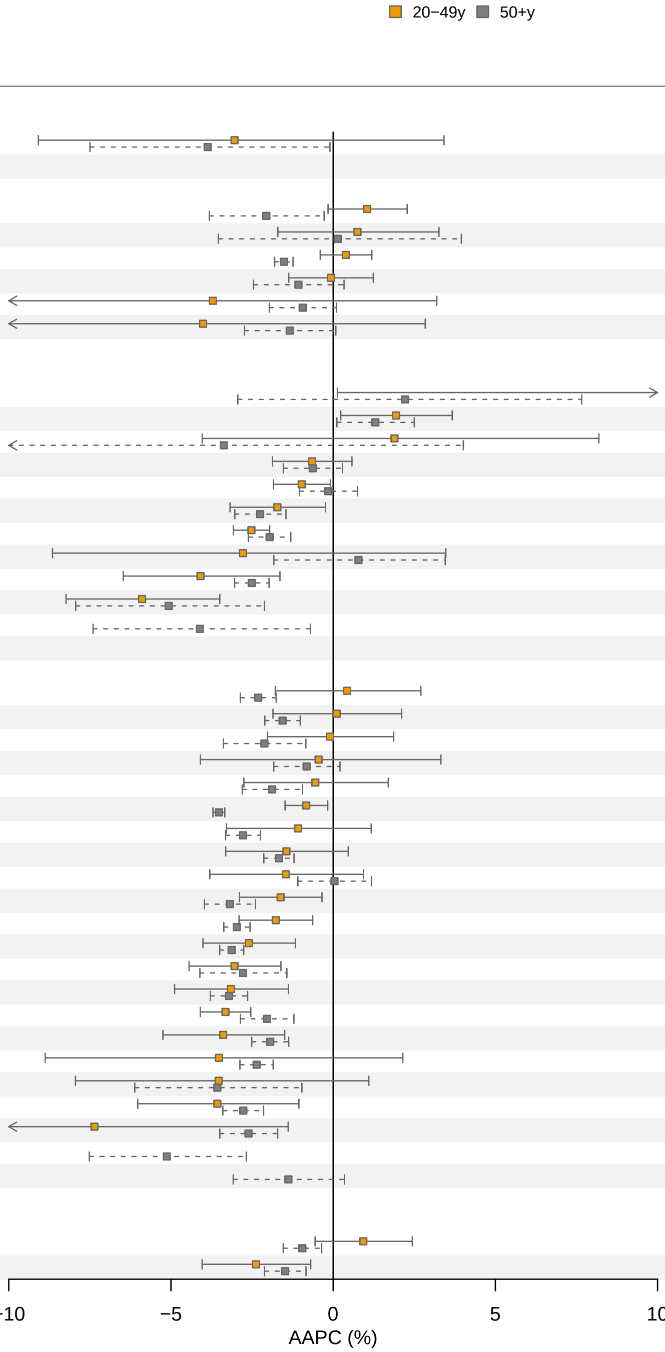

| Country | AAPC (%) 20−49y | AAPC (%) 50+y | p value |
|---|---|---|---|
| **Africa** | | | |
| Uganda | −3.04 | −3.87 | 0.81 |
| **Americas** | | | |
| Colombia | 1.05 | −2.06 ‡ | 0.005 |
| Ecuador | 0.75 | 0.14 ‡ | 0.79 |
| USA | 0.39 | −1.52 | <0.001 |
| Canada | −0.07 | −1.07 ‡ | 0.30 |
| Argentina | −3.71 ‡ | −0.94 | 0.43 |
| Chile | −4.01 | −1.34 | 0.42 |
| **Asia** | | | |
| Kuwait | 12.44 ‡ | 2.22 | 0.140 |
| Thailand | 1.94 | 1.3 | 0.53 |
| Qatar | 1.89 | −3.37 | 0.26 |
| Türkiye | −0.65 | −0.63 ‡ | 0.97 |
| India | −0.97 | −0.15 | 0.187 |
| Israel | −1.72 | −2.25 | 0.51 |
| Republic of Korea | −2.52 ‡ | −1.96 ‡ | 0.21 |
| Cyprus | −2.78 | 0.78 ‡ | 0.28 |
| China | −4.09 ‡ | −2.51 ‡ | 0.21 |
| Philippines | −5.89 | −5.07 ‡ | 0.66 |
| Bahrain | − | −4.11 | − |
| **Europe** | | | |
| Sweden | 0.43 | −2.31 | 0.022 |
| France | 0.11 ‡ | −1.56 | 0.108 |
| Ireland | −0.1 | −2.12 ‡ | 0.082 |
| Switzerland | −0.45 | −0.82 | 0.84 |
| Latvia | −0.55 | −1.88 ‡ | 0.26 |
| UK | −0.83 | −3.52 | <0.001 |
| Norway | −1.08 | −2.78 | 0.132 |
| Germany | −1.44 | −1.67 | 0.80 |
| Denmark | −1.46 | 0.04 ‡ | 0.25 |
| The Netherlands | −1.62 | −3.18 ‡ | 0.041 |
| Austria | −1.77 | −2.97 | 0.050 |
| Czechia | −2.6 | −3.13 | 0.45 |
| Croatia | −3.04 | −2.78 ‡ | 0.79 |
| Finland | −3.15 | −3.21 | 0.94 |
| Belarus | −3.32 | −2.04 ‡ | 0.027 |
| Estonia | −3.39 | −1.94 | 0.138 |
| Lithuania | −3.52 ‡ | −2.36 | 0.68 |
| Poland | −3.53 | −3.57 ‡ | 0.99 |
| Italy | −3.57 | −2.77 | 0.52 |
| Slovenia | −7.36 ‡ | −2.61 ‡ | 0.120 |
| Iceland | − | −5.13 | − |
| Malta | − | −1.38 | − |
| **Oceania** | | | |
| Australia | 0.93 | −0.95 ‡ | 0.024 |
| New Zealand | −2.38 | −1.48 | 0.30 |

‡ best fitted Joinpoint model > 0 joins; * p−value for a difference between AAPC 20−49y and AAPC 50+y.

#### Appendix Figure 26: Average annual percent change (AAPC) for cancer incidence (2003-2017) by UN region, country and age - Thyroid Cancer

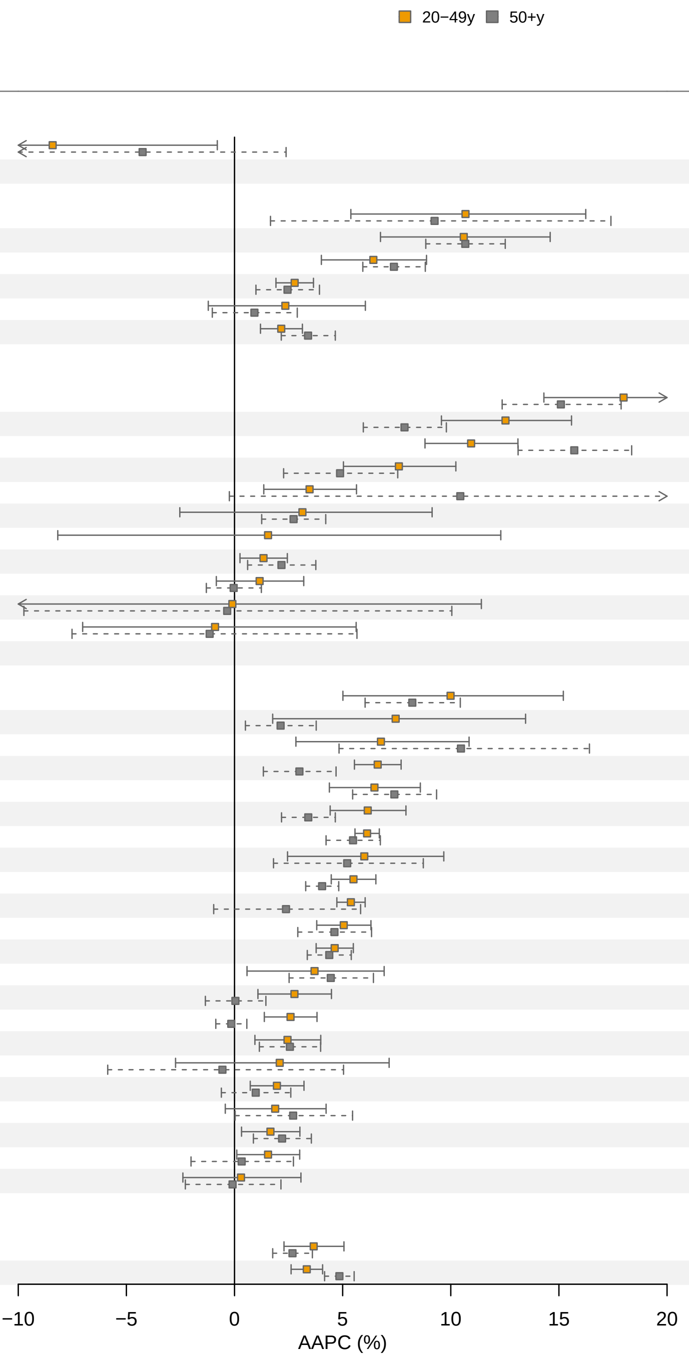

| Country | AAPC (%) 20−49y | AAPC (%) 50+y | p value |
|---|---|---|---|
| **Africa** | | | |
| Uganda | −8.41 ‡ | −4.25 | 0.40 |
| **Americas** | | | |
| Chile | 10.68 | 9.25 ‡ | 0.77 |
| Ecuador | 10.6 ‡ | 10.67 | 0.98 |
| Colombia | 6.42 ‡ | 7.37 | 0.51 |
| USA | 2.78 ‡ | 2.45 ‡ | 0.70 |
| Argentina | 2.35 | 0.92 | 0.47 |
| Canada | 2.16 ‡ | 3.4 ‡ | 0.124 |
| **Asia** | | | |
| China | 17.99 ‡ | 15.09 ‡ | 0.22 |
| Türkiye | 12.53 ‡ | 7.86 ‡ | 0.010 |
| Cyprus | 10.94 | 15.71 | 0.008 |
| Republic of Korea | 7.6 ‡ | 4.88 ‡ | 0.149 |
| Kuwait | 3.47 ‡ | 10.44 ‡ | 0.22 |
| Thailand | 3.14 ‡ | 2.73 | 0.89 |
| Qatar | 1.55 ‡ | – | – |
| India | 1.34 | 2.17 | 0.38 |
| Israel | 1.16 ‡ | −0.04 ‡ | 0.32 |
| Bahrain | −0.1 ‡ | −0.34 ‡ | 0.98 |
| Philippines | −0.9 ‡ | −1.15 ‡ | 0.96 |
| **Europe** | | | |
| Latvia | 9.99 ‡ | 8.22 ‡ | 0.53 |
| Poland | 7.45 ‡ | 2.13 | 0.079 |
| Malta | 6.77 | 10.47 | 0.28 |
| Croatia | 6.62 | 3 ‡ | <0.001 |
| Denmark | 6.47 ‡ | 7.39 | 0.52 |
| Germany | 6.16 | 3.41 | 0.014 |
| UK | 6.13 ‡ | 5.48 ‡ | 0.35 |
| Ireland | 6 ‡ | 5.21 ‡ | 0.76 |
| Sweden | 5.5 | 4.05 | 0.028 |
| Czechia | 5.38 | 2.38 ‡ | 0.093 |
| Norway | 5.05 | 4.62 ‡ | 0.69 |
| The Netherlands | 4.63 | 4.38 | 0.69 |
| Switzerland | 3.7 ‡ | 4.45 | 0.69 |
| Austria | 2.77 ‡ | 0.04 ‡ | 0.015 |
| Belarus | 2.59 ‡ | −0.15 | <0.001 |
| Slovenia | 2.45 | 2.56 | 0.92 |
| Iceland | 2.09 | −0.56 | 0.46 |
| Lithuania | 1.96 | 0.98 ‡ | 0.34 |
| Finland | 1.88 ‡ | 2.71 ‡ | 0.65 |
| Italy | 1.66 | 2.2 | 0.56 |
| France | 1.55 ‡ | 0.33 ‡ | 0.39 |
| Estonia | 0.3 | −0.09 | 0.82 |
| **Oceania** | | | |
| New Zealand | 3.66 | 2.68 | 0.23 |
| Australia | 3.34 ‡ | 4.85 ‡ | 0.003 |

‡ best fitted Joinpoint model > 0 joins; * p−value for a difference between AAPC 20−49y and AAPC 50+y.

Appendix Table 1: Source of cancer incidence data by country in GLOBOCAN

| **Country** | **Data provider** | **Method*** |
|---|---|---|
| Argentina | Mendoza cancer registry | 3c |
| Australia | National cancer registry | 1 |
| Austria | National cancer registry | 1 |
| Bahrain | Bahrain: Bahraini cancer registry | 3c |
| Belarus | National cancer registry | 1 |
| Canada | Canada (Excl Nova Scotia, Northwest Territories, Nunavut, Quebec and Yukon) cancer registry | 1 |
| Chile | Valdivia cancer registry | 3c |
| China | Shanghai City, Jiashan County, Zhongshan City, Nangang District, Harbin City cancer registries | 2b |
| Colombia | Cali, Bucaramanga, Manizales, Pasto cancer registries | 3c |
| Croatia | National cancer registry | 1 |
| Cyprus | Cyprus cancer registry | 1 |
| Czechia | National cancer registry | 1 |
| Denmark | National cancer registry (Nordcan database) | 1 |
| Ecuador | Quito cancer registry | 3c |
| Estonia | National cancer registry | 1 |
| Finland | National cancer registry (Nordcan database) | 1 |
| France (metropolitan) | Calvados, Doubs, Haut-Rhin, Isère, Somme, Hérault, Loire-Atlantique, Manche, Vendée cancer registries | 3a |
| Germany | Hamburg, Bremen, Schleswig-Holstein, Saarland cancer registries | 3a |
| Iceland | National cancer registry (Nordcan database) | 1 |
| India | Mumbai, Chennai, Barshi, Dindigul Ambilikkai cancer registries | 2b |
| Ireland | National cancer registry | 1 |
| Israel | National cancer registry | 1 |
| Italy | Trento, Syracuse, Palermo, South Tyrol cancer registries | 3a |
| Kuwait | Kuwait: Kuwaiti cancer registry | 1 |
| Latvia | National cancer registry | 1 |
| Lithuania | National cancer registry | 1 |
| Malta | National cancer registry | 1 |
| New Zealand | National cancer registry | 1 |
| Norway | National cancer registry (Nordcan database) | 1 |

| | | |
|---|---|---|
| Phillippines | Manila cancer registry | 2b |
| Poland | Kielce cancer registry | 1 |
| Qatar | Qatar: Qatari cancer registry | 3a |
| Republic of Korea | National cancer registry | 1 |
| Slovenia | National cancer registry | 1 |
| Sweden | National cancer registry (Nordcan database) | 1 |
| Switzerland | Geneva, Vaud, Valais, Ticino, Graubünden and Glarus cancer registries | 3a |
| Thailand | Chiang Mai, Khon Kaen, Songkhla, Lampang cancer registries | 2b |
| The Netherlands | National cancer registry | 1 |
| Türkiye | Izmir, Antalya cancer registries | 2b |
| Uganda | Kyadondo County cancer registry | 2b |
| UK | Combined data England cancer registry, Wales cancer registry and Scotland cancer registry | 1 |
| USA | SEER 9 (San Francisco-Oakland and Los Angeles Cancer Registries (California), Connecticut Tumor Registry, Atlanta Tumor Registry (Georgia), Hawaii Tumor Registry, State Health Registry of Iowa, Detroit Cancer Registry (Michigan), New Mexico Tumor Registry, Utah Cancer Registry, Seattle-Puget Sound Tumor Registry (Washington State) | 1 |

*Method for estimating cancer incidence:

1 National (or sub-national with coverage greater than 50%) rates

2b Weighted/simple average of the most recent sub-national rates

3a Estimated from national mortality estimates by modelling, using mortality:incidence ratios derived from country-specific cancer registry data

3c Estimated from national mortality estimates by modelling, using mortality:incidence ratios derived from survival estimation

Appendix Table 2: ICD-10 codes for cancers based on Globocan

| **Cancer** | **ICD-10 code** |
|---|---|
| Bladder | C67 |
| Brain and CNS | C70-C72 |
| Breast | C50 |
| Cervix | C53 |
| Colorectum | C18-C21 |
| Endometrium | C54 |
| Gallbladder | C23-C24 |
| Hodgkin lymphoma | C81 |
| Kidney | C64 |
| Larynx | C32 |
| Leukaemia | C91-95 |
| Liver | C22 |
| Lung | C33-C34 |
| Melanoma of skin | C43 |
| Multiple myeloma | C88,C90 |
| Non-Hodkin lymphoma | C82-C86, C96 |
| Oesophagus | C15 |
| Oral | C00-C14 |
| Ovary | C56 |
| Pancreas | C25 |
| Prostate | C61 |
| Stomach | C16 |
| Testis | C62 |
| Thyroid | C73 |

**Appendix Table 3:** Number and percentage of countries where the average annual percentage change (AAPC) in cancer incidence is significantly greater in younger than older adults (and increasing AAPC in younger adults). Results show the number of countries with a statistically significant (P<0.05) AAPC difference, and the subset of those countries where the Bayes false discovery probability (BFDP) is <0.8[1], using a range of prior effect sizes for the expected magnitude of the AAPC difference. The prior probability of a true difference in AAPC was set to 0.05. BFDPs were calculated using three prior variances, corresponding to the prior beliefs that increases in AAPC from 0 to AAPCs greater than 1%, 2.5%, and 5% occur with a probability of 2.5%

| | | **Countries with significant (P<0.05) AAPC differences between age groups** *2* | | **Countries with significant AAPC differences between age groups and BFDP <0.8** | | | | | |
|---|---|---|---|---|---|---|---|---|---|
| | | | | **AAPC used in prior variance** | | | | | |
| | | | | 1%[3] | | 2.5%[3] | | 5%[3] | |
| Cancer | Total number of countries | n | % | n | % | n | % | n | % |
| Breast | 42 | 4 | 10% | 2 | 5% | 2 | 5% | 2 | 5% |
| Colorectum | 42 | 16 | 38% | 8 | 19% | 11 | 26% | 11 | 26% |
| Endometrium | 36 | 5 | 14% | 1 | 3% | 1 | 3% | 2 | 6% |
| Gallbladder | 29 | 1 | 3% | 0 | 0% | 0 | 0% | 0 | 0% |
| Kidney | 40 | 10 | 25% | 2 | 5% | 6 | 15% | 5 | 12% |
| Leukaemia | 40 | 4 | 10% | 1 | 2% | 1 | 2% | 1 | 2% |
| Liver | 35 | 1 | 3% | 0 | 0% | 0 | 0% | 0 | 0% |
| Oesophagus | 32 | 1 | 3% | 0 | 0% | 0 | 0% | 1 | 3% |
| Oral | 39 | 4 | 10% | 2 | 5% | 2 | 5% | 2 | 5% |
| Pancreas | 36 | 4 | 11% | 1 | 3% | 2 | 6% | 2 | 6% |
| Prostate | 35 | 10 | 29% | 1 | 3% | 4 | 11% | 5 | 14% |
| Stomach | 39 | 4 | 10% | 1 | 3% | 2 | 5% | 2 | 5% |
| Thyroid | 41 | 6 | 15% | 1 | 2% | 2 | 5% | 5 | 12% |

*1* BFDP < 0.8 was used to identify "noteworthy findings" among those with p < 0.05.

*2* Based on countries where the AAPC for younger people is greater than zero (increasing trends) and larger in magnitude than the AAPC for older people.

*3*Prior effect sizes correspond to 97.5% quantile AAPC increases of 1%, 2.5% and 5% from an AAPC of 0, used to derive the prior variances for BFDP calculations.

**Appendix Table 4: Segment specific annual percentage change (APC) for cancer incidence trends from 2003 to 2017 by country: breast cancer**

| | Joinpoint trend 1 | | | Joinpoint trend 2 | | | Joinpoint trend 3 | | | |
|---|---|---|---|---|---|---|---|---|---|---|
| **Country** | **Years** | **APC** | **95% CI** | **Years** | **APC** | **95% CI** | **Years** | **APC** | **95% CI** | **AAPC** |
| Argentina | | | | | | | | | | |
| 20-49y | 2003-2005 | -10.59 | -28.1, 11.18 | 2005-2017 | 1.12 | -0.18, 2.43 | - | - | - | -0.64 |
| 50+y | 2003-2017 | -1.12 | -2.05, -0.19 | - | - | - | - | - | - | -1.12 |
| Australia | | | | | | | | | | |
| 20-49y | 2003-2017 | 0.81 | 0.52, 1.1 | - | - | - | - | - | - | 0.81 |
| 50+y | 2003-2017 | 1.17 | 0.83, 1.51 | - | - | - | - | - | - | 1.17 |
| Austria | | | | | | | | | | |
| 20-49y | 2003-2014 | 1.25 | 0.38, 2.12 | 2014-2017 | -2.67 | -8.71, 3.76 | - | - | - | 0.39 |
| 50+y | 2003-2017 | -0.50 | -0.97, -0.03 | - | - | - | - | - | - | -0.50 |
| Bahrain | | | | | | | | | | |
| 20-49y | 2003-2017 | 1.38 | -1.37, 4.2 | - | - | - | - | - | - | 1.38 |
| 50+y | 2003-2017 | 1.71 | -0.16, 3.6 | - | - | - | - | - | - | 1.71 |
| Belarus | | | | | | | | | | |
| 20-49y | 2003-2013 | 0.23 | -0.84, 1.31 | 2013-2017 | 3.94 | -0.49, 8.56 | - | - | - | 1.28 |
| 50+y | 2003-2017 | 2.20 | 1.94, 2.45 | - | - | - | - | - | - | 2.20 |
| Canada | | | | | | | | | | |
| 20-49y | 2003-2017 | 0.62 | 0.33, 0.92 | - | - | - | - | - | - | 0.62 |
| 50+y | 2003-2017 | 0.74 | 0.49, 0.99 | - | - | - | - | - | - | 0.74 |
| Chile | | | | | | | | | | |
| 20-49y | 2003-2017 | 0.37 | -2.72, 3.56 | - | - | - | - | - | - | 0.37 |
| 50+y | 2003-2017 | 1.08 | -1.13, 3.34 | - | - | - | - | - | - | 1.08 |
| China | | | | | | | | | | |
| 20-49y | 2003-2017 | 1.27 | 0.36, 2.19 | - | - | - | - | - | - | 1.27 |
| 50+y | 2003-2017 | 2.53 | 2.03, 3.04 | - | - | - | - | - | - | 2.53 |
| Colombia | | | | | | | | | | |
| 20-49y | 2003-2017 | 0.08 | -0.74, 0.91 | - | - | - | - | - | - | 0.08 |
| 50+y | 2003-2006 | 4.81 | -3.67, 14.03 | 2006-2011 | -3.16 | -8.19, 2.15 | 2011-2017 | 4.43 | 1.49, 7.45 | 1.73 |
| Croatia | | | | | | | | | | |
| 20-49y | 2003-2007 | -1.15 | -5.98, 3.92 | 2007-2017 | 2.72 | 1.46, 3.99 | - | - | - | 1.60 |
| 50+y | 2003-2017 | 0.93 | 0.11, 1.76 | - | - | - | - | - | - | 0.93 |
| Cyprus | | | | | | | | | | |
| 20-49y | 2003-2017 | 1.17 | -0.31, 2.68 | - | - | - | - | - | - | 1.17 |
| 50+y | 2003-2017 | 0.87 | 0, 1.75 | - | - | - | - | - | - | 0.87 |
| Czechia | | | | | | | | | | |
| 20-49y | 2003-2017 | 2.03 | 1.62, 2.45 | - | - | - | - | - | - | 2.03 |
| 50+y | 2003-2017 | 0.18 | -0.35, 0.72 | - | - | - | - | - | - | 0.18 |
| Denmark | | | | | | | | | | |
| 20-49y | 2003-2017 | 1.10 | 0.58, 1.61 | - | - | - | - | - | - | 1.10 |
| 50+y | 2003-2009 | 5.25 | 2.35, 8.23 | 2009-2017 | -2.96 | -4.69, -1.19 | - | - | - | 0.48 |
| Ecuador | | | | | | | | | | |
| 20-49y | 2003-2015 | 0.18 | -2, 2.41 | 2015-2017 | 21.02 | -16.65, 75.71 | - | - | - | 2.92 |
| 50+y | 2003-2017 | 1.46 | -0.1, 3.05 | - | - | - | - | - | - | 1.46 |
| Estonia | | | | | | | | | | |
| 20-49y | 2003-2017 | 1.91 | 0.81, 3.03 | - | - | - | - | - | - | 1.91 |

**Appendix Table 4: Segment specific annual percentage change (APC) for cancer incidence trends from 2003 to 2017 by country: breast cancer**

| | Joinpoint trend 1 | | | Joinpoint trend 2 | | | Joinpoint trend 3 | | | |
|---|---|---|---|---|---|---|---|---|---|---|
| Country | Years | APC | 95% CI | Years | APC | 95% CI | Years | APC | 95% CI | AAPC |
| 50+y | 2003-2017 | 1.23 | 0.42, 2.05 | - | - | - | - | - | - | 1.23 |
| Finland | | | | | | | | | | |
| 20-49y | 2003-2015 | -0.01 | -0.68, 0.67 | 2015-2017 | 5.77 | -5.64, 18.57 | - | - | - | 0.80 |
| 50+y | 2003-2010 | 2.06 | 1.07, 3.06 | 2010-2017 | 0.50 | -0.48, 1.49 | - | - | - | 1.28 |
| France | | | | | | | | | | |
| 20-49y | 2003-2017 | 0.57 | 0.17, 0.98 | - | - | - | - | - | - | 0.57 |
| 50+y | 2003-2006 | -2.90 | -6.76, 1.12 | 2006-2017 | 0.09 | -0.45, 0.64 | - | - | - | -0.56 |
| Germany | | | | | | | | | | |
| 20-49y | 2003-2005 | 6.69 | -3.77, 18.28 | 2005-2017 | 0.59 | -0.02, 1.21 | - | - | - | 1.44 |
| 50+y | 2003-2005 | -5.48 | -15.42, 5.64 | 2005-2008 | 7.96 | -3.4, 20.65 | 2008-2017 | -2.37 | -3.36, -1.38 | -0.71 |
| Iceland | | | | | | | | | | |
| 20-49y | 2003-2017 | 0.28 | -0.97, 1.54 | - | - | - | - | - | - | 0.28 |
| 50+y | 2003-2017 | -0.54 | -1.66, 0.59 | - | - | - | - | - | - | -0.54 |
| India | | | | | | | | | | |
| 20-49y | 2003-2017 | 0.27 | -0.64, 1.18 | - | - | - | - | - | - | 0.27 |
| 50+y | 2003-2017 | 2.64 | 2.06, 3.23 | - | - | - | - | - | - | 2.64 |
| Ireland | | | | | | | | | | |
| 20-49y | 2003-2017 | 0.50 | -0.13, 1.13 | - | - | - | - | - | - | 0.50 |
| 50+y | 2003-2005 | -4.20 | -10.69, 2.77 | 2005-2008 | 6.31 | -0.9, 14.03 | 2008-2017 | -0.44 | -1.07, 0.2 | 0.42 |
| Israel | | | | | | | | | | |
| 20-49y | 2003-2009 | -1.55 | -4, 0.96 | 2009-2015 | 2.69 | -0.68, 6.16 | 2015-2017 | -7.00 | -19.87, 7.94 | -0.57 |
| 50+y | 2003-2017 | 0.09 | -0.4, 0.58 | - | - | - | - | - | - | 0.09 |
| Italy | | | | | | | | | | |
| 20-49y | 2003-2006 | -3.07 | -7.36, 1.43 | 2006-2017 | 1.71 | 1.09, 2.33 | - | - | - | 0.67 |
| 50+y | 2003-2017 | 0.93 | 0.46, 1.4 | - | - | - | - | - | - | 0.93 |
| Kuwait | | | | | | | | | | |
| 20-49y | 2003-2017 | 1.55 | 0.23, 2.88 | - | - | - | - | - | - | 1.55 |
| 50+y | 2003-2017 | 2.86 | 1.63, 4.1 | - | - | - | - | - | - | 2.86 |
| Latvia | | | | | | | | | | |
| 20-49y | 2003-2017 | 2.81 | 1.83, 3.79 | - | - | - | - | - | - | 2.81 |
| 50+y | 2003-2017 | 1.98 | 1.12, 2.84 | - | - | - | - | - | - | 1.98 |
| Lithuania | | | | | | | | | | |
| 20-49y | 2003-2005 | -5.69 | -20.05, 11.26 | 2005-2017 | 2.15 | 1.16, 3.16 | - | - | - | 1.00 |
| 50+y | 2003-2015 | 2.23 | 1.3, 3.16 | 2015-2017 | -6.58 | -19.86, 8.89 | - | - | - | 0.92 |
| Malta | | | | | | | | | | |
| 20-49y | 2003-2005 | 17.48 | -11.87, 56.62 | 2005-2017 | -0.31 | -1.99, 1.4 | - | - | - | 2.06 |
| 50+y | 2003-2011 | 2.47 | 0.57, 4.41 | 2011-2017 | -1.29 | -4.11, 1.61 | - | - | - | 0.84 |
| New Zealand | | | | | | | | | | |
| 20-49y | 2003-2014 | 1.49 | 0.58, 2.41 | 2014-2017 | -3.48 | -9.74, 3.22 | - | - | - | 0.40 |
| 50+y | 2003-2017 | 0.55 | 0.23, 0.88 | - | - | - | - | - | - | 0.55 |
| Norway | | | | | | | | | | |
| 20-49y | 2003-2017 | 0.97 | 0.29, 1.65 | - | - | - | - | - | - | 0.97 |
| 50+y | 2003-2009 | -1.86 | -3.45, -0.25 | 2009-2017 | 2.07 | 1, 3.14 | - | - | - | 0.36 |
| Philippines | | | | | | | | | | |
| 20-49y | 2003-2017 | -1.80 | -2.8, -0.79 | - | - | - | - | - | - | -1.80 |

**Appendix Table 4: Segment specific annual percentage change (APC) for cancer incidence trends from 2003 to 2017 by country: breast cancer**

| | Joinpoint trend 1 | | | Joinpoint trend 2 | | | Joinpoint trend 3 | | | |
|---|---|---|---|---|---|---|---|---|---|---|
| Country | Years | APC | 95% CI | Years | APC | 95% CI | Years | APC | 95% CI | AAPC |
| 50+y | 2003-2017 | -0.57 | -1.42, 0.28 | - | - | - | - | - | - | -0.57 |
| Poland | | | | | | | | | | |
| 20-49y | 2003-2017 | 1.19 | 0.01, 2.39 | - | - | - | - | - | - | 1.19 |
| 50+y | 2003-2017 | 1.88 | 1.07, 2.69 | - | - | - | - | - | - | 1.88 |
| Qatar | | | | | | | | | | |
| 20-49y | 2003-2017 | -2.61 | -5.09, -0.07 | - | - | - | - | - | - | -2.61 |
| 50+y | 2003-2017 | 7.15 | 2.56, 11.95 | - | - | - | - | - | - | 7.15 |
| Republic of Korea | | | | | | | | | | |
| 20-49y | 2003-2017 | 4.31 | 4.03, 4.59 | - | - | - | - | - | - | 4.31 |
| 50+y | 2003-2011 | 6.94 | 6.12, 7.77 | 2011-2014 | 2.97 | -4.04, 10.48 | 2014-2017 | 7.01 | 3.31, 10.85 | 6.09 |
| Slovenia | | | | | | | | | | |
| 20-49y | 2003-2011 | -0.11 | -1.55, 1.34 | 2011-2017 | 4.36 | 2.04, 6.72 | - | - | - | 1.78 |
| 50+y | 2003-2017 | 0.52 | 0.09, 0.96 | - | - | - | - | - | - | 0.52 |
| Sweden | | | | | | | | | | |
| 20-49y | 2003-2017 | 0.35 | -0.08, 0.77 | - | - | - | - | - | - | 0.35 |
| 50+y | 2003-2007 | -1.82 | -4.09, 0.52 | 2007-2012 | 1.93 | -0.44, 4.35 | 2012-2017 | -0.87 | -2.51, 0.78 | -0.15 |
| Switzerland | | | | | | | | | | |
| 20-49y | 2003-2017 | 0.09 | -0.7, 0.9 | - | - | - | - | - | - | 0.09 |
| 50+y | 2003-2017 | -0.48 | -0.8, -0.16 | - | - | - | - | - | - | -0.48 |
| Thailand | | | | | | | | | | |
| 20-49y | 2003-2017 | 1.74 | 0.9, 2.58 | - | - | - | - | - | - | 1.74 |
| 50+y | 2003-2010 | 3.15 | 1.87, 4.44 | 2010-2015 | 6.62 | 3.52, 9.82 | 2015-2017 | -4.05 | -12.61, 5.34 | 3.30 |
| The Netherlands | | | | | | | | | | |
| 20-49y | 2003-2017 | 0.51 | 0.17, 0.86 | - | - | - | - | - | - | 0.51 |
| 50+y | 2003-2012 | 1.22 | 0.81, 1.63 | 2012-2017 | -1.03 | -2, -0.04 | - | - | - | 0.41 |
| Türkiye | | | | | | | | | | |
| 20-49y | 2003-2017 | 2.05 | 1.46, 2.64 | - | - | - | - | - | - | 2.05 |
| 50+y | 2003-2017 | 1.76 | 1.06, 2.47 | - | - | - | - | - | - | 1.76 |
| UK | | | | | | | | | | |
| 20-49y | 2003-2007 | -0.36 | -1.93, 1.24 | 2007-2013 | 2.18 | 1.04, 3.34 | 2013-2017 | -0.44 | -2.01, 1.16 | 0.70 |
| 50+y | 2003-2011 | -0.12 | -0.36, 0.11 | 2011-2014 | 2.32 | 0.15, 4.54 | 2014-2017 | -1.41 | -2.46, -0.35 | 0.12 |
| USA | | | | | | | | | | |
| 20-49y | 2003-2017 | 0.30 | 0.09, 0.51 | - | - | - | - | - | - | 0.30 |
| 50+y | 2003-2017 | -0.09 | -0.2, 0.02 | - | - | - | - | - | - | -0.09 |
| Uganda | | | | | | | | | | |
| 20-49y | 2003-2017 | 1.14 | -0.59, 2.89 | - | - | - | - | - | - | 1.14 |
| 50+y | 2003-2017 | -0.52 | -3.19, 2.21 | - | - | - | - | - | - | -0.52 |

**Appendix Table 5: Segment specific annual percentage change (APC) for cancer incidence trends from 2003 to 2017 by country: colorectum cancer**

| | Joinpoint trend 1 | | | Joinpoint trend 2 | | | Joinpoint trend 3 | | | |
|---|---|---|---|---|---|---|---|---|---|---|
| **Country** | **Years** | **APC** | **95% CI** | **Years** | **APC** | **95% CI** | **Years** | **APC** | **95% CI** | **AAPC** |
| Argentina | | | | | | | | | | |
| 20-49y | 2003-2017 | 3.10 | 0.9, 5.35 | - | - | - | - | - | - | 3.10 |
| 50+y | 2003-2017 | 0.18 | -0.72, 1.08 | - | - | - | - | - | - | 0.18 |
| Australia | | | | | | | | | | |
| 20-49y | 2003-2017 | 3.11 | 2.68, 3.55 | - | - | - | - | - | - | 3.11 |
| 50+y | 2003-2007 | 0.65 | -1.34, 2.68 | 2007-2017 | -2.39 | -2.87, -1.91 | - | - | - | -1.53 |
| Austria | | | | | | | | | | |
| 20-49y | 2003-2017 | 0.45 | -0.72, 1.63 | - | - | - | - | - | - | 0.45 |
| 50+y | 2003-2017 | -2.72 | -2.99, -2.45 | - | - | - | - | - | - | -2.72 |
| Bahrain | | | | | | | | | | |
| 20-49y | 2003-2017 | -1.65 | -4.88, 1.69 | - | - | - | - | - | - | -1.65 |
| 50+y | 2003-2017 | 3.39 | 1.69, 5.13 | - | - | - | - | - | - | 3.39 |
| Belarus | | | | | | | | | | |
| 20-49y | 2003-2015 | 0.95 | -0.2, 2.11 | 2015-2017 | 12.34 | -7.45, 36.36 | - | - | - | 2.50 |
| 50+y | 2003-2017 | 2.34 | 2.09, 2.6 | - | - | - | - | - | - | 2.34 |
| Canada | | | | | | | | | | |
| 20-49y | 2003-2017 | 2.88 | 2.41, 3.35 | - | - | - | - | - | - | 2.88 |
| 50+y | 2003-2015 | -0.87 | -1.28, -0.47 | 2015-2017 | -5.66 | -11.97, 1.1 | - | - | - | -1.57 |
| Chile | | | | | | | | | | |
| 20-49y | 2003-2017 | 4.25 | -0.73, 9.47 | - | - | - | - | - | - | 4.25 |
| 50+y | 2003-2017 | 1.74 | -0.32, 3.84 | - | - | - | - | - | - | 1.74 |
| China | | | | | | | | | | |
| 20-49y | 2003-2010 | -3.14 | -5.08, -1.16 | 2010-2013 | 9.80 | -5.62, 27.73 | 2013-2017 | -1.37 | -5.98, 3.46 | 0.01 |
| 50+y | 2003-2017 | 0.86 | 0.58, 1.15 | - | - | - | - | - | - | 0.86 |
| Colombia | | | | | | | | | | |
| 20-49y | 2003-2006 | 11.04 | 1.19, 21.85 | 2006-2017 | 0.00 | -1.25, 1.26 | - | - | - | 2.27 |
| 50+y | 2003-2017 | 1.25 | 0.59, 1.92 | - | - | - | - | - | - | 1.25 |
| Croatia | | | | | | | | | | |
| 20-49y | 2003-2017 | 0.75 | -0.51, 2.03 | - | - | - | - | - | - | 0.75 |
| 50+y | 2003-2017 | 1.22 | 0.71, 1.73 | - | - | - | - | - | - | 1.22 |
| Cyprus | | | | | | | | | | |
| 20-49y | 2003-2017 | -0.19 | -2.56, 2.23 | - | - | - | - | - | - | -0.19 |
| 50+y | 2003-2007 | 7.38 | -0.27, 15.62 | 2007-2017 | -1.46 | -3.24, 0.35 | - | - | - | 0.99 |
| Czechia | | | | | | | | | | |
| 20-49y | 2003-2013 | -1.56 | -2.87, -0.23 | 2013-2017 | 3.88 | -1.63, 9.69 | - | - | - | -0.04 |
| 50+y | 2003-2014 | -1.30 | -1.72, -0.87 | 2014-2017 | -5.53 | -8.51, -2.45 | - | - | - | -2.22 |
| Denmark | | | | | | | | | | |
| 20-49y | 2003-2017 | 1.98 | 1.06, 2.92 | - | - | - | - | - | - | 1.98 |
| 50+y | 2003-2017 | 1.30 | 0.57, 2.03 | - | - | - | - | - | - | 1.30 |
| Ecuador | | | | | | | | | | |
| 20-49y | 2003-2017 | 2.16 | 0.05, 4.3 | - | - | - | - | - | - | 2.16 |
| 50+y | 2003-2017 | 2.63 | 1.34, 3.94 | - | - | - | - | - | - | 2.63 |
| Estonia | | | | | | | | | | |
| 20-49y | 2003-2017 | 2.55 | -0.67, 5.86 | - | - | - | - | - | - | 2.55 |

#### Appendix Table 5: Segment specific annual percentage change (APC) for cancer incidence trends from 2003 to 2017 by country: colorectum cancer

| | Joinpoint trend 1 | | | Joinpoint trend 2 | | | Joinpoint trend 3 | | | |
|---|---|---|---|---|---|---|---|---|---|---|
| **Country** | **Years** | **APC** | **95% CI** | **Years** | **APC** | **95% CI** | **Years** | **APC** | **95% CI** | **AAPC** |
| 50+y | 2003-2017 | 1.16 | 0.57, 1.76 | - | - | - | - | - | - | 1.16 |
| Finland | | | | | | | | | | |
| 20-49y | 2003-2017 | 1.52 | 0.23, 2.83 | - | - | - | - | - | - | 1.52 |
| 50+y | 2003-2017 | 0.71 | 0.43, 0.98 | - | - | - | - | - | - | 0.71 |
| France | | | | | | | | | | |
| 20-49y | 2003-2008 | -1.23 | -3.77, 1.38 | 2008-2017 | 2.23 | 1.14, 3.32 | - | - | - | 0.98 |
| 50+y | 2003-2017 | -0.35 | -1.01, 0.31 | - | - | - | - | - | - | -0.35 |
| Germany | | | | | | | | | | |
| 20-49y | 2003-2015 | 2.73 | 1.97, 3.5 | 2015-2017 | -6.66 | -17.7, 5.87 | - | - | - | 1.34 |
| 50+y | 2003-2017 | -1.75 | -2.04, -1.45 | - | - | - | - | - | - | -1.75 |
| Iceland | | | | | | | | | | |
| 20-49y | 2003-2017 | 4.27 | 1.02, 7.62 | - | - | - | - | - | - | 4.27 |
| 50+y | 2003-2017 | 0.21 | -1.14, 1.58 | - | - | - | - | - | - | 0.21 |
| India | | | | | | | | | | |
| 20-49y | 2003-2017 | 1.38 | 0.13, 2.64 | - | - | - | - | - | - | 1.38 |
| 50+y | 2003-2007 | 5.18 | 1.72, 8.76 | 2007-2017 | 2.43 | 1.59, 3.28 | - | - | - | 3.21 |
| Ireland | | | | | | | | | | |
| 20-49y | 2003-2017 | 1.57 | 0.51, 2.64 | - | - | - | - | - | - | 1.57 |
| 50+y | 2003-2007 | 1.46 | -0.73, 3.71 | 2007-2017 | -1.36 | -1.89, -0.82 | - | - | - | -0.56 |
| Israel | | | | | | | | | | |
| 20-49y | 2003-2017 | 0.64 | -0.33, 1.61 | - | - | - | - | - | - | 0.64 |
| 50+y | 2003-2017 | -3.53 | -4.07, -3 | - | - | - | - | - | - | -3.53 |
| Italy | | | | | | | | | | |
| 20-49y | 2003-2017 | 0.61 | -0.45, 1.68 | - | - | - | - | - | - | 0.61 |
| 50+y | 2003-2010 | 1.21 | -0.42, 2.86 | 2010-2017 | -3.07 | -4.63, -1.49 | - | - | - | -0.96 |
| Kuwait | | | | | | | | | | |
| 20-49y | 2003-2005 | 51.05 | -13.96, 165.17 | 2005-2017 | -0.33 | -3.59, 3.04 | - | - | - | 5.77 |
| 50+y | 2003-2017 | 1.80 | 0.21, 3.42 | - | - | - | - | - | - | 1.80 |
| Latvia | | | | | | | | | | |
| 20-49y | 2003-2017 | 0.50 | -1.58, 2.61 | - | - | - | - | - | - | 0.50 |
| 50+y | 2003-2009 | -1.45 | -2.62, -0.27 | 2009-2012 | 5.98 | -1.23, 13.73 | 2012-2017 | -2.07 | -3.6, -0.51 | -0.13 |
| Lithuania | | | | | | | | | | |
| 20-49y | 2003-2005 | -9.41 | -28.26, 14.38 | 2005-2017 | 1.12 | -0.27, 2.52 | - | - | - | -0.46 |
| 50+y | 2003-2010 | 1.67 | -0.02, 3.39 | 2010-2017 | -1.62 | -3.26, 0.04 | - | - | - | 0.01 |
| Malta | | | | | | | | | | |
| 20-49y | 2003-2006 | -20.69 | -45.35, 15.1 | 2006-2017 | 6.99 | 1.75, 12.5 | - | - | - | 0.34 |
| 50+y | 2003-2017 | 0.45 | -0.92, 1.84 | - | - | - | - | - | - | 0.45 |
| New Zealand | | | | | | | | | | |
| 20-49y | 2003-2011 | 1.06 | -1.01, 3.18 | 2011-2017 | 5.92 | 2.57, 9.38 | - | - | - | 3.12 |
| 50+y | 2003-2014 | -1.56 | -1.93, -1.19 | 2014-2017 | -4.15 | -6.78, -1.46 | - | - | - | -2.12 |
| Norway | | | | | | | | | | |
| 20-49y | 2003-2010 | -0.80 | -2.64, 1.08 | 2010-2013 | 10.69 | -3.79, 27.36 | 2013-2017 | 0.03 | -4.31, 4.57 | 1.80 |
| 50+y | 2003-2015 | 0.55 | 0.36, 0.74 | 2015-2017 | -1.53 | -4.68, 1.73 | - | - | - | 0.25 |
| Philippines | | | | | | | | | | |
| 20-49y | 2003-2017 | -1.52 | -2.91, -0.12 | - | - | - | - | - | - | -1.52 |

**Appendix Table 5: Segment specific annual percentage change (APC) for cancer incidence trends from 2003 to 2017 by country: colorectum cancer**

| | Joinpoint trend 1 | | | Joinpoint trend 2 | | | Joinpoint trend 3 | | | |
|---|---|---|---|---|---|---|---|---|---|---|
| Country | Years | APC | 95% CI | Years | APC | 95% CI | Years | APC | 95% CI | AAPC |
| 50+y | 2003-2011 | -3.43 | -5.83, -0.97 | 2011-2017 | 1.50 | -2.39, 5.54 | - | - | - | -1.35 |
| Poland | | | | | | | | | | |
| 20-49y | 2003-2017 | 0.73 | -1.18, 2.69 | - | - | - | - | - | - | 0.73 |
| 50+y | 2003-2011 | 2.03 | 0.47, 3.62 | 2011-2017 | -2.01 | -4.32, 0.36 | - | - | - | 0.28 |
| Qatar | | | | | | | | | | |
| 20-49y | 2003-2017 | 0.24 | -4.49, 5.2 | - | - | - | - | - | - | 0.24 |
| 50+y | 2003-2017 | 1.11 | -0.44, 2.67 | - | - | - | - | - | - | 1.11 |
| Republic of Korea | | | | | | | | | | |
| 20-49y | 2003-2012 | 4.32 | 3.47, 5.18 | 2012-2015 | -7.71 | -15.62, 0.94 | 2015-2017 | 1.50 | -7.2, 11.02 | 1.22 |
| 50+y | 2003-2010 | 5.04 | 4.04, 6.04 | 2010-2017 | -3.81 | -4.72, -2.88 | - | - | - | 0.52 |
| Slovenia | | | | | | | | | | |
| 20-49y | 2003-2017 | 0.80 | -0.54, 2.15 | - | - | - | - | - | - | 0.80 |
| 50+y | 2003-2010 | 3.05 | 0.48, 5.69 | 2010-2017 | -5.58 | -7.94, -3.17 | - | - | - | -1.36 |
| Sweden | | | | | | | | | | |
| 20-49y | 2003-2017 | 2.73 | 1.95, 3.5 | - | - | - | - | - | - | 2.73 |
| 50+y | 2003-2007 | 1.38 | 0.39, 2.38 | 2007-2014 | -0.55 | -1.07, -0.02 | 2014-2017 | 1.99 | 0.42, 3.59 | 0.54 |
| Switzerland | | | | | | | | | | |
| 20-49y | 2003-2017 | 2.29 | 0.2, 4.42 | - | - | - | - | - | - | 2.29 |
| 50+y | 2003-2017 | -1.16 | -1.81, -0.5 | - | - | - | - | - | - | -1.16 |
| Thailand | | | | | | | | | | |
| 20-49y | 2003-2017 | 2.56 | 1.36, 3.78 | - | - | - | - | - | - | 2.56 |
| 50+y | 2003-2017 | 4.08 | 3.28, 4.89 | - | - | - | - | - | - | 4.08 |
| The Netherlands | | | | | | | | | | |
| 20-49y | 2003-2006 | -1.23 | -4.07, 1.7 | 2006-2009 | 4.90 | -1.05, 11.2 | 2009-2017 | 0.96 | 0.32, 1.61 | 1.32 |
| 50+y | 2003-2017 | 1.29 | 0.65, 1.94 | - | - | - | - | - | - | 1.29 |
| Türkiye | | | | | | | | | | |
| 20-49y | 2003-2017 | 2.07 | 1.22, 2.94 | - | - | - | - | - | - | 2.07 |
| 50+y | 2003-2017 | 1.78 | 1.41, 2.14 | - | - | - | - | - | - | 1.78 |
| UK | | | | | | | | | | |
| 20-49y | 2003-2017 | 3.26 | 2.82, 3.71 | - | - | - | - | - | - | 3.26 |
| 50+y | 2003-2011 | 1.35 | 1.08, 1.62 | 2011-2014 | -3.00 | -5.34, -0.6 | 2014-2017 | -0.61 | -1.82, 0.61 | -0.02 |
| USA | | | | | | | | | | |
| 20-49y | 2003-2013 | 1.57 | 1.09, 2.06 | 2013-2017 | 4.68 | 2.67, 6.72 | - | - | - | 2.45 |
| 50+y | 2003-2008 | -2.48 | -3.7, -1.25 | 2008-2011 | -4.85 | -10.05, 0.65 | 2011-2017 | -1.04 | -1.97, -0.09 | -2.38 |
| Uganda | | | | | | | | | | |
| 20-49y | 2003-2017 | 0.80 | -3.68, 5.49 | - | - | - | - | - | - | 0.80 |
| 50+y | 2003-2017 | 2.75 | -0.06, 5.65 | - | - | - | - | - | - | 2.75 |

**Appendix Table 6: Segment specific annual percentage change (APC) for cancer incidence trends from 2003 to 2017 by country: endometrium cancer**

| | Joinpoint trend 1 | | | Joinpoint trend 2 | | | Joinpoint trend 3 | | | |
|---|---|---|---|---|---|---|---|---|---|---|
| Country | Years | APC | 95% CI | Years | APC | 95% CI | Years | APC | 95% CI | AAPC |
| Argentina | | | | | | | | | | |
| 20-49y | 2003-2017 | 1.13 | -2.92, 5.35 | - | - | - | - | - | - | 1.13 |
| 50+y | 2003-2017 | 1.04 | -0.52, 2.62 | - | - | - | - | - | - | 1.04 |
| Australia | | | | | | | | | | |
| 20-49y | 2003-2017 | 2.65 | 1.56, 3.75 | - | - | - | - | - | - | 2.65 |
| 50+y | 2003-2013 | 1.87 | 1.4, 2.35 | 2013-2017 | -0.63 | -2.51, 1.29 | - | - | - | 1.15 |
| Austria | | | | | | | | | | |
| 20-49y | 2003-2017 | -1.58 | -3.52, 0.39 | - | - | - | - | - | - | -1.58 |
| 50+y | 2003-2017 | -1.91 | -2.26, -1.55 | - | - | - | - | - | - | -1.91 |
| Bahrain | | | | | | | | | | |
| 20-49y | - | - | - | - | - | - | - | - | - | - |
| 50+y | 2003-2017 | 5.37 | -0.48, 11.57 | - | - | - | - | - | - | 5.37 |
| Belarus | | | | | | | | | | |
| 20-49y | 2003-2017 | 2.35 | 1.18, 3.54 | - | - | - | - | - | - | 2.35 |
| 50+y | 2003-2017 | 3.31 | 2.92, 3.71 | - | - | - | - | - | - | 3.31 |
| Canada | | | | | | | | | | |
| 20-49y | 2003-2017 | 2.88 | 1.97, 3.79 | - | - | - | - | - | - | 2.88 |
| 50+y | 2003-2008 | 1.68 | 0.57, 2.8 | 2008-2011 | 3.93 | -1.06, 9.16 | 2011-2017 | 1.12 | 0.29, 1.97 | 1.92 |
| Chile | | | | | | | | | | |
| 20-49y | - | - | - | - | - | - | - | - | - | - |
| 50+y | 2003-2017 | 5.49 | 1.76, 9.35 | - | - | - | - | - | - | 5.49 |
| China | | | | | | | | | | |
| 20-49y | 2003-2017 | 3.44 | 1.87, 5.04 | - | - | - | - | - | - | 3.44 |
| 50+y | 2003-2017 | 1.42 | 0.54, 2.3 | - | - | - | - | - | - | 1.42 |
| Colombia | | | | | | | | | | |
| 20-49y | 2003-2017 | 0.74 | -1.48, 3.02 | - | - | - | - | - | - | 0.74 |
| 50+y | 2003-2017 | 1.55 | 0.72, 2.39 | - | - | - | - | - | - | 1.55 |
| Croatia | | | | | | | | | | |
| 20-49y | 2003-2005 | 25.37 | -20.52, 97.76 | 2005-2017 | 1.83 | -0.88, 4.61 | - | - | - | 4.90 |
| 50+y | 2003-2017 | 2.49 | 1.73, 3.27 | - | - | - | - | - | - | 2.49 |
| Cyprus | | | | | | | | | | |
| 20-49y | 2003-2017 | 5.61 | 1.26, 10.15 | - | - | - | - | - | - | 5.61 |
| 50+y | 2003-2017 | 1.09 | -0.25, 2.44 | - | - | - | - | - | - | 1.09 |
| Czechia | | | | | | | | | | |
| 20-49y | 2003-2017 | -0.33 | -1.36, 0.72 | - | - | - | - | - | - | -0.33 |
| 50+y | 2003-2017 | -0.33 | -0.69, 0.03 | - | - | - | - | - | - | -0.33 |
| Denmark | | | | | | | | | | |
| 20-49y | 2003-2017 | 3.23 | 1.11, 5.38 | - | - | - | - | - | - | 3.23 |
| 50+y | 2003-2013 | 0.79 | -0.14, 1.73 | 2013-2017 | -2.50 | -6.1, 1.24 | - | - | - | -0.16 |
| Ecuador | | | | | | | | | | |
| 20-49y | 2003-2017 | 5.48 | -1.94, 13.45 | - | - | - | - | - | - | 5.48 |
| 50+y | 2003-2017 | 1.70 | -0.77, 4.22 | - | - | - | - | - | - | 1.70 |
| Estonia | | | | | | | | | | |
| 20-49y | 2003-2017 | 2.79 | 0.25, 5.4 | - | - | - | - | - | - | 2.79 |

**Appendix Table 6: Segment specific annual percentage change (APC) for cancer incidence trends from 2003 to 2017 by country: endometrium cancer**

| | Joinpoint trend 1 | | | Joinpoint trend 2 | | | Joinpoint trend 3 | | | |
|---|---|---|---|---|---|---|---|---|---|---|
| Country | Years | APC | 95% CI | Years | APC | 95% CI | Years | APC | 95% CI | AAPC |
| 50+y | 2003-2009 | -0.61 | -3.14, 1.98 | 2009-2017 | 2.92 | 1.22, 4.64 | - | - | - | 1.39 |
| Finland | | | | | | | | | | |
| 20-49y | 2003-2017 | 0.22 | -1.7, 2.17 | - | - | - | - | - | - | 0.22 |
| 50+y | 2003-2017 | -1.14 | -1.7, -0.59 | - | - | - | - | - | - | -1.14 |
| France | | | | | | | | | | |
| 20-49y | 2003-2012 | -4.16 | -7.84, -0.33 | 2012-2017 | 6.25 | -3.47, 16.94 | - | - | - | -0.56 |
| 50+y | 2003-2013 | 1.07 | 0.25, 1.9 | 2013-2017 | -2.71 | -5.9, 0.58 | - | - | - | -0.03 |
| Germany | | | | | | | | | | |
| 20-49y | 2003-2017 | 1.57 | 0.22, 2.95 | - | - | - | - | - | - | 1.57 |
| 50+y | 2003-2005 | 6.62 | -4.87, 19.5 | 2005-2017 | -0.78 | -1.45, -0.11 | - | - | - | 0.24 |
| Iceland | | | | | | | | | | |
| 20-49y | - | - | - | - | - | - | - | - | - | - |
| 50+y | 2003-2017 | 2.12 | -1.48, 5.86 | - | - | - | - | - | - | 2.12 |
| India | | | | | | | | | | |
| 20-49y | 2003-2017 | 3.54 | 1.25, 5.88 | - | - | - | - | - | - | 3.54 |
| 50+y | 2003-2011 | 8.36 | 5.62, 11.18 | 2011-2017 | 3.37 | -0.66, 7.56 | - | - | - | 6.19 |
| Ireland | | | | | | | | | | |
| 20-49y | 2003-2017 | 2.06 | 0.38, 3.76 | - | - | - | - | - | - | 2.06 |
| 50+y | 2003-2017 | 2.81 | 2.01, 3.61 | - | - | - | - | - | - | 2.81 |
| Israel | | | | | | | | | | |
| 20-49y | 2003-2017 | -2.82 | -4.22, -1.4 | - | - | - | - | - | - | -2.82 |
| 50+y | 2003-2014 | 0.08 | -0.73, 0.89 | 2014-2017 | -4.55 | -10.09, 1.33 | - | - | - | -0.93 |
| Italy | | | | | | | | | | |
| 20-49y | 2003-2017 | 0.45 | -1.4, 2.32 | - | - | - | - | - | - | 0.45 |
| 50+y | 2003-2017 | -0.05 | -1.02, 0.94 | - | - | - | - | - | - | -0.05 |
| Kuwait | | | | | | | | | | |
| 20-49y | - | - | - | - | - | - | - | - | - | - |
| 50+y | 2003-2017 | 5.09 | 2.67, 7.58 | - | - | - | - | - | - | 5.09 |
| Latvia | | | | | | | | | | |
| 20-49y | 2003-2017 | -0.34 | -3.32, 2.73 | - | - | - | - | - | - | -0.34 |
| 50+y | 2003-2017 | -0.17 | -0.92, 0.57 | - | - | - | - | - | - | -0.17 |
| Lithuania | | | | | | | | | | |
| 20-49y | 2003-2017 | 2.99 | 1.33, 4.67 | - | - | - | - | - | - | 2.99 |
| 50+y | 2003-2017 | 1.36 | 0.71, 2.02 | - | - | - | - | - | - | 1.36 |
| Malta | | | | | | | | | | |
| 20-49y | 2003-2005 | 67.83 | -62.33, 647.63 | 2005-2012 | -17.78 | -36.13, 5.84 | 2012-2017 | 36.55 | -2.23, 90.71 | 9.13 |
| 50+y | 2003-2017 | 0.48 | -1.63, 2.63 | - | - | - | - | - | - | 0.48 |
| New Zealand | | | | | | | | | | |
| 20-49y | 2003-2013 | 6.65 | 4.04, 9.32 | 2013-2017 | -4.12 | -13.31, 6.03 | - | - | - | 3.45 |
| 50+y | 2003-2017 | 1.51 | 0.9, 2.13 | - | - | - | - | - | - | 1.51 |
| Norway | | | | | | | | | | |
| 20-49y | 2003-2017 | -0.91 | -2.22, 0.42 | - | - | - | - | - | - | -0.91 |
| 50+y | 2003-2017 | -0.74 | -1.55, 0.07 | - | - | - | - | - | - | -0.74 |
| Philippines | | | | | | | | | | |
| 20-49y | 2003-2006 | 3.89 | -9.57, 19.35 | 2006-2012 | -7.33 | -12.91, -1.4 | 2012-2017 | 6.30 | -0.1, 13.11 | -0.26 |

**Appendix Table 6: Segment specific annual percentage change (APC) for cancer incidence trends from 2003 to 2017 by country: endometrium cancer**

| | Joinpoint trend 1 | | | Joinpoint trend 2 | | | Joinpoint trend 3 | | | |
|---|---|---|---|---|---|---|---|---|---|---|
| Country | Years | APC | 95% CI | Years | APC | 95% CI | Years | APC | 95% CI | AAPC |
| 50+y | 2003-2009 | -5.14 | -8.46, -1.71 | 2009-2017 | 2.81 | 0.47, 5.2 | - | - | - | -0.68 |
| Poland | | | | | | | | | | |
| 20-49y | 2003-2017 | -0.31 | -3.83, 3.33 | - | - | - | - | - | - | -0.31 |
| 50+y | 2003-2008 | -1.79 | -5.04, 1.58 | 2008-2017 | 2.72 | 1.32, 4.14 | - | - | - | 1.09 |
| Qatar | | | | | | | | | | |
| 20-49y | - | - | - | - | - | - | - | - | - | - |
| 50+y | - | - | - | - | - | - | - | - | - | - |
| Republic of Korea | | | | | | | | | | |
| 20-49y | 2003-2017 | 5.11 | 4.42, 5.8 | - | - | - | - | - | - | 5.11 |
| 50+y | 2003-2017 | 4.87 | 4.25, 5.5 | - | - | - | - | - | - | 4.87 |
| Slovenia | | | | | | | | | | |
| 20-49y | 2003-2017 | 0.77 | -1.24, 2.82 | - | - | - | - | - | - | 0.77 |
| 50+y | 2003-2008 | -2.59 | -6.73, 1.74 | 2008-2017 | 1.30 | -0.48, 3.12 | - | - | - | -0.10 |
| Sweden | | | | | | | | | | |
| 20-49y | 2003-2017 | 1.74 | 0.45, 3.05 | - | - | - | - | - | - | 1.74 |
| 50+y | 2003-2009 | -0.02 | -1.15, 1.13 | 2009-2017 | -2.14 | -2.85, -1.41 | - | - | - | -1.23 |
| Switzerland | | | | | | | | | | |
| 20-49y | 2003-2017 | 1.39 | -1.73, 4.61 | - | - | - | - | - | - | 1.39 |
| 50+y | 2003-2011 | -1.59 | -3.84, 0.7 | 2011-2017 | 2.47 | -1.13, 6.2 | - | - | - | 0.13 |
| Thailand | | | | | | | | | | |
| 20-49y | 2003-2017 | 4.58 | 1.84, 7.39 | - | - | - | - | - | - | 4.58 |
| 50+y | 2003-2017 | 3.83 | 2.42, 5.27 | - | - | - | - | - | - | 3.83 |
| The Netherlands | | | | | | | | | | |
| 20-49y | 2003-2017 | -0.88 | -2.05, 0.3 | - | - | - | - | - | - | -0.88 |
| 50+y | 2003-2017 | -0.76 | -1.2, -0.33 | - | - | - | - | - | - | -0.76 |
| Türkiye | | | | | | | | | | |
| 20-49y | 2003-2017 | 1.36 | 0.19, 2.54 | - | - | - | - | - | - | 1.36 |
| 50+y | 2003-2017 | 2.20 | 1.42, 2.98 | - | - | - | - | - | - | 2.20 |
| UK | | | | | | | | | | |
| 20-49y | 2003-2017 | 2.54 | 1.93, 3.15 | - | - | - | - | - | - | 2.54 |
| 50+y | 2003-2010 | 3.01 | 2.12, 3.91 | 2010-2017 | 0.37 | -0.5, 1.24 | - | - | - | 1.68 |
| USA | | | | | | | | | | |
| 20-49y | 2003-2017 | 1.40 | 0.8, 2 | - | - | - | - | - | - | 1.40 |
| 50+y | 2003-2010 | 1.77 | 1.02, 2.52 | 2010-2017 | 0.64 | -0.1, 1.38 | - | - | - | 1.20 |
| Uganda | | | | | | | | | | |
| 20-49y | - | - | - | - | - | - | - | - | - | - |
| 50+y | 2003-2017 | 2.50 | -3.61, 8.98 | - | - | - | - | - | - | 2.50 |

**Appendix Table 7: Segment specific annual percentage change (APC) for cancer incidence trends from 2003 to 2017 by country: gallbladder cancer**

| | Joinpoint trend 1 | | | Joinpoint trend 2 | | | Joinpoint trend 3 | | | |
|---|---|---|---|---|---|---|---|---|---|---|
| **Country** | **Years** | **APC** | **95% CI** | **Years** | **APC** | **95% CI** | **Years** | **APC** | **95% CI** | **AAPC** |
| Argentina | | | | | | | | | | |
| 20-49y | - | - | - | - | - | - | - | - | - | - |
| 50+y | 2003-2010 | 0.82 | -4.52, 6.46 | 2010-2017 | -6.65 | -11.6, -1.43 | - | - | - | -2.99 |
| Australia | | | | | | | | | | |
| 20-49y | 2003-2017 | 0.85 | -1.11, 2.85 | - | - | - | - | - | - | 0.85 |
| 50+y | 2003-2013 | 0.57 | -0.36, 1.51 | 2013-2017 | 4.55 | 0.68, 8.57 | - | - | - | 1.69 |
| Austria | | | | | | | | | | |
| 20-49y | 2003-2017 | 2.01 | -1.75, 5.9 | - | - | - | - | - | - | 2.01 |
| 50+y | 2003-2017 | -0.58 | -1.34, 0.19 | - | - | - | - | - | - | -0.58 |
| Bahrain | | | | | | | | | | |
| 20-49y | - | - | - | - | - | - | - | - | - | - |
| 50+y | - | - | - | - | - | - | - | - | - | - |
| Belarus | | | | | | | | | | |
| 20-49y | 2003-2017 | 2.38 | -1.5, 6.42 | - | - | - | - | - | - | 2.38 |
| 50+y | 2003-2005 | 12.15 | -4.51, 31.71 | 2005-2011 | -1.42 | -4.9, 2.19 | 2011-2017 | 4.05 | 1.26, 6.92 | 2.76 |
| Canada | | | | | | | | | | |
| 20-49y | 2003-2005 | 20.98 | -14.37, 70.93 | 2005-2017 | 2.97 | 0.89, 5.1 | - | - | - | 5.37 |
| 50+y | 2003-2005 | -4.98 | -16.88, 8.63 | 2005-2017 | 2.26 | 1.45, 3.07 | - | - | - | 1.19 |
| Chile | | | | | | | | | | |
| 20-49y | 2003-2017 | -5.58 | -9.92, -1.03 | - | - | - | - | - | - | -5.58 |
| 50+y | 2003-2009 | -7.80 | -13.85, -1.32 | 2009-2017 | -0.37 | -4.64, 4.09 | - | - | - | -3.62 |
| China | | | | | | | | | | |
| 20-49y | 2003-2011 | -6.73 | -12.08, -1.05 | 2011-2017 | 10.17 | 0.52, 20.74 | - | - | - | 0.17 |
| 50+y | 2003-2017 | -1.66 | -2.28, -1.03 | - | - | - | - | - | - | -1.66 |
| Colombia | | | | | | | | | | |
| 20-49y | 2003-2017 | 0.06 | -3.05, 3.26 | - | - | - | - | - | - | 0.06 |
| 50+y | 2003-2017 | -1.56 | -3.41, 0.34 | - | - | - | - | - | - | -1.56 |
| Croatia | | | | | | | | | | |
| 20-49y | 2003-2017 | -1.51 | -8.05, 5.49 | - | - | - | - | - | - | -1.51 |
| 50+y | 2003-2017 | -0.88 | -1.9, 0.16 | - | - | - | - | - | - | -0.88 |
| Cyprus | | | | | | | | | | |
| 20-49y | - | - | - | - | - | - | - | - | - | - |
| 50+y | 2003-2017 | 1.32 | -1.05, 3.75 | - | - | - | - | - | - | 1.32 |
| Czechia | | | | | | | | | | |
| 20-49y | 2003-2012 | -9.87 | -15.14, -4.27 | 2012-2017 | 7.11 | -7.58, 24.14 | - | - | - | -4.14 |
| 50+y | 2003-2017 | -3.53 | -4.01, -3.04 | - | - | - | - | - | - | -3.53 |
| Denmark | | | | | | | | | | |
| 20-49y | 2003-2017 | 1.77 | -1.82, 5.49 | - | - | - | - | - | - | 1.77 |
| 50+y | 2003-2007 | 7.91 | -0.3, 16.8 | 2007-2017 | -0.60 | -2.52, 1.36 | - | - | - | 1.76 |
| Ecuador | | | | | | | | | | |
| 20-49y | 2003-2017 | -1.59 | -6.18, 3.24 | - | - | - | - | - | - | -1.59 |
| 50+y | 2003-2017 | -1.74 | -4.22, 0.8 | - | - | - | - | - | - | -1.74 |
| Estonia | | | | | | | | | | |
| 20-49y | - | - | - | - | - | - | - | - | - | - |

**Appendix Table 7: Segment specific annual percentage change (APC) for cancer incidence trends from 2003 to 2017 by country: gallbladder cancer**

| | Joinpoint trend 1 | | | Joinpoint trend 2 | | | Joinpoint trend 3 | | | |
|---|---|---|---|---|---|---|---|---|---|---|
| Country | Years | APC | 95% CI | Years | APC | 95% CI | Years | APC | 95% CI | AAPC |
| 50+y | 2003-2017 | 2.27 | 0.05, 4.54 | - | - | - | - | - | - | 2.27 |
| Finland | | | | | | | | | | |
| 20-49y | 2003-2008 | -16.87 | -34.62, 5.7 | 2008-2017 | 11.30 | 0.91, 22.77 | - | - | - | 0.29 |
| 50+y | 2003-2017 | 0.03 | -0.97, 1.05 | - | - | - | - | - | - | 0.03 |
| France | | | | | | | | | | |
| 20-49y | 2003-2017 | -3.15 | -7.82, 1.74 | - | - | - | - | - | - | -3.15 |
| 50+y | 2003-2017 | 0.26 | -0.5, 1.02 | - | - | - | - | - | - | 0.26 |
| Germany | | | | | | | | | | |
| 20-49y | 2003-2008 | 11.61 | -6.97, 33.9 | 2008-2017 | -10.09 | -16.53, -3.15 | - | - | - | -2.87 |
| 50+y | 2003-2017 | -0.16 | -1.3, 0.99 | - | - | - | - | - | - | -0.16 |
| Iceland | | | | | | | | | | |
| 20-49y | - | - | - | - | - | - | - | - | - | - |
| 50+y | 2003-2017 | 0.21 | -5.43, 6.19 | - | - | - | - | - | - | 0.21 |
| India | | | | | | | | | | |
| 20-49y | 2003-2017 | 1.77 | -0.44, 4.03 | - | - | - | - | - | - | 1.77 |
| 50+y | 2003-2017 | 3.43 | 2.25, 4.63 | - | - | - | - | - | - | 3.43 |
| Ireland | | | | | | | | | | |
| 20-49y | - | - | - | - | - | - | - | - | - | - |
| 50+y | 2003-2017 | 1.54 | 0.29, 2.82 | - | - | - | - | - | - | 1.54 |
| Israel | | | | | | | | | | |
| 20-49y | 2003-2017 | -0.98 | -6.13, 4.45 | - | - | - | - | - | - | -0.98 |
| 50+y | 2003-2009 | -4.82 | -10.1, 0.76 | 2009-2017 | 3.09 | -0.64, 6.95 | - | - | - | -0.38 |
| Italy | | | | | | | | | | |
| 20-49y | 2003-2012 | -13.80 | -20.93, -6.02 | 2012-2017 | 15.47 | -6.56, 42.68 | - | - | - | -4.31 |
| 50+y | 2003-2009 | -4.64 | -7.31, -1.89 | 2009-2015 | 0.83 | -2.89, 4.69 | 2015-2017 | -11.59 | -25.26, 4.59 | -3.38 |
| Kuwait | | | | | | | | | | |
| 20-49y | - | - | - | - | - | - | - | - | - | - |
| 50+y | 2003-2017 | -1.83 | -7.1, 3.74 | - | - | - | - | - | - | -1.83 |
| Latvia | | | | | | | | | | |
| 20-49y | - | - | - | - | - | - | - | - | - | - |
| 50+y | 2003-2017 | -2.47 | -4.47, -0.41 | - | - | - | - | - | - | -2.47 |
| Lithuania | | | | | | | | | | |
| 20-49y | - | - | - | - | - | - | - | - | - | - |
| 50+y | 2003-2010 | -6.00 | -9.09, -2.81 | 2010-2014 | 7.81 | -4.85, 22.16 | 2014-2017 | -12.08 | -22.41, -0.38 | -3.64 |
| Malta | | | | | | | | | | |
| 20-49y | - | - | - | - | - | - | - | - | - | - |
| 50+y | 2003-2017 | -2.04 | -7.08, 3.29 | - | - | - | - | - | - | -2.04 |
| New Zealand | | | | | | | | | | |
| 20-49y | 2003-2017 | 4.80 | -0.37, 10.25 | - | - | - | - | - | - | 4.80 |
| 50+y | 2003-2006 | -19.66 | -30.9, -6.58 | 2006-2009 | 18.35 | -12.47, 60.02 | 2009-2017 | -0.49 | -3.71, 2.84 | -1.35 |
| Norway | | | | | | | | | | |
| 20-49y | 2003-2017 | -0.35 | -4.07, 3.51 | - | - | - | - | - | - | -0.35 |
| 50+y | 2003-2014 | 6.26 | 3.4, 9.2 | 2014-2017 | -12.30 | -28.37, 7.37 | - | - | - | 1.98 |
| Philippines | | | | | | | | | | |
| 20-49y | 2003-2017 | 0.98 | -4.62, 6.91 | - | - | - | - | - | - | 0.98 |

#### Appendix Table 7: Segment specific annual percentage change (APC) for cancer incidence trends from 2003 to 2017 by country: gallbladder cancer

| | Joinpoint trend 1 | | | Joinpoint trend 2 | | | Joinpoint trend 3 | | | |
|---|---|---|---|---|---|---|---|---|---|---|
| Country | Years | APC | 95% CI | Years | APC | 95% CI | Years | APC | 95% CI | AAPC |
| 50+y | 2003-2005 | 11.45 | -17.71, 50.94 | 2005-2009 | -10.70 | -23.27, 3.92 | 2009-2017 | 6.99 | 3.5, 10.59 | 2.20 |
| Poland | | | | | | | | | | |
| 20-49y | - | - | - | - | - | - | - | - | - | - |
| 50+y | 2003-2017 | -3.49 | -5.71, -1.22 | - | - | - | - | - | - | -3.49 |
| Qatar | | | | | | | | | | |
| 20-49y | - | - | - | - | - | - | - | - | - | - |
| 50+y | 2003-2017 | -0.60 | -8, 7.4 | - | - | - | - | - | - | -0.60 |
| Republic of Korea | | | | | | | | | | |
| 20-49y | 2003-2017 | -2.52 | -3.07, -1.96 | - | - | - | - | - | - | -2.52 |
| 50+y | 2003-2017 | -0.03 | -0.32, 0.26 | - | - | - | - | - | - | -0.03 |
| Slovenia | | | | | | | | | | |
| 20-49y | 2003-2005 | 142.03 | -25.29, 684.09 | 2005-2017 | -8.31 | -14.47, -1.71 | - | - | - | 5.33 |
| 50+y | 2003-2005 | 23.67 | 3.01, 48.48 | 2005-2010 | -3.10 | -8.54, 2.67 | 2010-2017 | 1.96 | -0.5, 4.48 | 2.92 |
| Sweden | | | | | | | | | | |
| 20-49y | 2003-2017 | 3.39 | 0.12, 6.77 | - | - | - | - | - | - | 3.39 |
| 50+y | 2003-2008 | -2.93 | -7.79, 2.19 | 2008-2017 | 2.45 | 0.32, 4.62 | - | - | - | 0.49 |
| Switzerland | | | | | | | | | | |
| 20-49y | 2003-2017 | 5.13 | 0.87, 9.57 | - | - | - | - | - | - | 5.13 |
| 50+y | 2003-2017 | -1.38 | -2.99, 0.27 | - | - | - | - | - | - | -1.38 |
| Thailand | | | | | | | | | | |
| 20-49y | 2003-2017 | 0.49 | -2.15, 3.2 | - | - | - | - | - | - | 0.49 |
| 50+y | 2003-2017 | 3.08 | 2.26, 3.91 | - | - | - | - | - | - | 3.08 |
| The Netherlands | | | | | | | | | | |
| 20-49y | 2003-2017 | 1.65 | -0.3, 3.64 | - | - | - | - | - | - | 1.65 |
| 50+y | 2003-2017 | 1.74 | 0.96, 2.53 | - | - | - | - | - | - | 1.74 |
| Türkiye | | | | | | | | | | |
| 20-49y | 2003-2017 | -0.35 | -4.9, 4.42 | - | - | - | - | - | - | -0.35 |
| 50+y | 2003-2017 | -0.66 | -1.67, 0.36 | - | - | - | - | - | - | -0.66 |
| UK | | | | | | | | | | |
| 20-49y | 2003-2017 | 2.60 | 1.46, 3.76 | - | - | - | - | - | - | 2.60 |
| 50+y | 2003-2010 | 4.50 | 2.93, 6.09 | 2010-2017 | 1.01 | -0.51, 2.55 | - | - | - | 2.74 |
| USA | | | | | | | | | | |
| 20-49y | 2003-2017 | 1.09 | -0.22, 2.42 | - | - | - | - | - | - | 1.09 |
| 50+y | 2003-2017 | -0.02 | -0.31, 0.27 | - | - | - | - | - | - | -0.02 |
| Uganda | | | | | | | | | | |
| 20-49y | - | - | - | - | - | - | - | - | - | - |
| 50+y | - | - | - | - | - | - | - | - | - | - |

**Appendix Table 8: Segment specific annual percentage change (APC) for cancer incidence trends from 2003 to 2017 by country: kidney cancer**

| | Joinpoint trend 1 | | | Joinpoint trend 2 | | | Joinpoint trend 3 | | | |
|---|---|---|---|---|---|---|---|---|---|---|
| Country | Years | APC | 95% CI | Years | APC | 95% CI | Years | APC | 95% CI | AAPC |
| Argentina | | | | | | | | | | |
| 20-49y | 2003-2006 | -17.63 | -36.58, 6.98 | 2006-2017 | 7.44 | 3.71, 11.29 | - | - | - | 1.49 |
| 50+y | 2003-2007 | -5.33 | -11.04, 0.74 | 2007-2013 | 6.54 | 1.96, 11.33 | 2013-2017 | -4.80 | -10.54, 1.3 | -0.26 |
| Australia | | | | | | | | | | |
| 20-49y | 2003-2007 | 7.95 | 2.35, 13.87 | 2007-2017 | 1.93 | 0.6, 3.28 | - | - | - | 3.62 |
| 50+y | 2003-2017 | 1.76 | 1.42, 2.11 | - | - | - | - | - | - | 1.76 |
| Austria | | | | | | | | | | |
| 20-49y | 2003-2017 | 0.42 | -0.78, 1.62 | - | - | - | - | - | - | 0.42 |
| 50+y | 2003-2017 | -1.24 | -1.66, -0.83 | - | - | - | - | - | - | -1.24 |
| Bahrain | | | | | | | | | | |
| 20-49y | 2003-2017 | 2.44 | -6.08, 11.72 | - | - | - | - | - | - | 2.44 |
| 50+y | 2003-2007 | -24.13 | -46.93, 8.47 | 2007-2017 | 6.72 | -2.27, 16.54 | - | - | - | -3.19 |
| Belarus | | | | | | | | | | |
| 20-49y | 2003-2017 | 2.55 | 1.59, 3.52 | - | - | - | - | - | - | 2.55 |
| 50+y | 2003-2009 | 4.25 | 2.44, 6.1 | 2009-2017 | 1.78 | 0.63, 2.94 | - | - | - | 2.83 |
| Canada | | | | | | | | | | |
| 20-49y | 2003-2017 | 3.22 | 2.43, 4.01 | - | - | - | - | - | - | 3.22 |
| 50+y | 2003-2017 | 1.93 | 1.48, 2.38 | - | - | - | - | - | - | 1.93 |
| Chile | | | | | | | | | | |
| 20-49y | 2003-2017 | 2.51 | -3.25, 8.62 | - | - | - | - | - | - | 2.51 |
| 50+y | 2003-2017 | -0.24 | -2.39, 1.95 | - | - | - | - | - | - | -0.24 |
| China | | | | | | | | | | |
| 20-49y | 2003-2017 | 2.43 | 0.22, 4.69 | - | - | - | - | - | - | 2.43 |
| 50+y | 2003-2012 | 3.48 | 2.19, 4.78 | 2012-2017 | -0.56 | -3.57, 2.53 | - | - | - | 2.02 |
| Colombia | | | | | | | | | | |
| 20-49y | 2003-2017 | 4.27 | 0.9, 7.76 | - | - | - | - | - | - | 4.27 |
| 50+y | 2003-2017 | 3.66 | 2.59, 4.74 | - | - | - | - | - | - | 3.66 |
| Croatia | | | | | | | | | | |
| 20-49y | 2003-2017 | 1.97 | 0.81, 3.14 | - | - | - | - | - | - | 1.97 |
| 50+y | 2003-2017 | 2.56 | 1.78, 3.34 | - | - | - | - | - | - | 2.56 |
| Cyprus | | | | | | | | | | |
| 20-49y | 2003-2005 | -57.82 | -88.17, 50.38 | 2005-2009 | 63.35 | -13.49, 208.45 | 2009-2017 | -8.66 | -20.49, 4.93 | -3.43 |
| 50+y | 2003-2017 | 3.70 | 1.54, 5.91 | - | - | - | - | - | - | 3.70 |
| Czechia | | | | | | | | | | |
| 20-49y | 2003-2009 | -2.95 | -5.55, -0.27 | 2009-2017 | 1.46 | -0.3, 3.25 | - | - | - | -0.45 |
| 50+y | 2003-2014 | 0.06 | -0.49, 0.62 | 2014-2017 | -2.55 | -6.48, 1.54 | - | - | - | -0.51 |
| Denmark | | | | | | | | | | |
| 20-49y | 2003-2017 | 4.77 | 2.94, 6.64 | - | - | - | - | - | - | 4.77 |
| 50+y | 2003-2017 | 3.06 | 2.38, 3.74 | - | - | - | - | - | - | 3.06 |
| Ecuador | | | | | | | | | | |
| 20-49y | 2003-2009 | -11.73 | -24.15, 2.74 | 2009-2012 | 51.27 | -38.35, 271.16 | 2012-2017 | -8.95 | -25.51, 11.28 | 0.17 |
| 50+y | 2003-2017 | 4.06 | 1.42, 6.76 | - | - | - | - | - | - | 4.06 |
| Estonia | | | | | | | | | | |
| 20-49y | 2003-2013 | 2.16 | -2.49, 7.03 | 2013-2017 | -12.40 | -27.5, 5.85 | - | - | - | -2.23 |

**Appendix Table 8: Segment specific annual percentage change (APC) for cancer incidence trends from 2003 to 2017 by country: kidney cancer**

| | Joinpoint trend 1 | | | Joinpoint trend 2 | | | Joinpoint trend 3 | | | |
|---|---|---|---|---|---|---|---|---|---|---|
| Country | Years | APC | 95% CI | Years | APC | 95% CI | Years | APC | 95% CI | AAPC |
| 50+y | 2003-2011 | 2.75 | 1.52, 4.01 | 2011-2015 | -3.46 | -8.67, 2.05 | 2015-2017 | 7.95 | -3.39, 20.61 | 1.65 |
| Finland | | | | | | | | | | |
| 20-49y | 2003-2017 | 1.54 | 0.02, 3.1 | - | - | - | - | - | - | 1.54 |
| 50+y | 2003-2017 | 0.80 | 0.29, 1.32 | - | - | - | - | - | - | 0.80 |
| France | | | | | | | | | | |
| 20-49y | 2003-2017 | 2.62 | 1.34, 3.91 | - | - | - | - | - | - | 2.62 |
| 50+y | 2003-2017 | 1.43 | 1.03, 1.84 | - | - | - | - | - | - | 1.43 |
| Germany | | | | | | | | | | |
| 20-49y | 2003-2011 | -0.77 | -2.57, 1.07 | 2011-2015 | 8.78 | 0.01, 18.31 | 2015-2017 | -12.10 | -25.7, 3.99 | 0.12 |
| 50+y | 2003-2017 | 0.01 | -0.43, 0.45 | - | - | - | - | - | - | 0.01 |
| Iceland | | | | | | | | | | |
| 20-49y | 2003-2017 | -0.59 | -7.86, 7.25 | - | - | - | - | - | - | -0.59 |
| 50+y | 2003-2017 | 0.14 | -2.38, 2.73 | - | - | - | - | - | - | 0.14 |
| India | | | | | | | | | | |
| 20-49y | 2003-2017 | 3.31 | 1.42, 5.24 | - | - | - | - | - | - | 3.31 |
| 50+y | 2003-2017 | 2.66 | 1.76, 3.56 | - | - | - | - | - | - | 2.66 |
| Ireland | | | | | | | | | | |
| 20-49y | 2003-2017 | 5.11 | 3.23, 7.03 | - | - | - | - | - | - | 5.11 |
| 50+y | 2003-2012 | 2.86 | 1.61, 4.12 | 2012-2017 | -0.49 | -3.41, 2.53 | - | - | - | 1.65 |
| Israel | | | | | | | | | | |
| 20-49y | 2003-2017 | 0.13 | -1.42, 1.69 | - | - | - | - | - | - | 0.13 |
| 50+y | 2003-2006 | 3.96 | -3.51, 12.01 | 2006-2017 | -2.75 | -3.72, -1.76 | - | - | - | -1.35 |
| Italy | | | | | | | | | | |
| 20-49y | 2003-2017 | 2.04 | 0.6, 3.5 | - | - | - | - | - | - | 2.04 |
| 50+y | 2003-2017 | 2.09 | 1.37, 2.82 | - | - | - | - | - | - | 2.09 |
| Kuwait | | | | | | | | | | |
| 20-49y | 2003-2017 | 5.45 | -1.21, 12.56 | - | - | - | - | - | - | 5.45 |
| 50+y | 2003-2017 | 1.48 | -2.06, 5.14 | - | - | - | - | - | - | 1.48 |
| Latvia | | | | | | | | | | |
| 20-49y | 2003-2017 | 2.85 | -0.36, 6.17 | - | - | - | - | - | - | 2.85 |
| 50+y | 2003-2017 | 2.24 | 1.26, 3.22 | - | - | - | - | - | - | 2.24 |
| Lithuania | | | | | | | | | | |
| 20-49y | 2003-2017 | 0.44 | -1.75, 2.68 | - | - | - | - | - | - | 0.44 |
| 50+y | 2003-2017 | 0.07 | -0.82, 0.97 | - | - | - | - | - | - | 0.07 |
| Malta | | | | | | | | | | |
| 20-49y | 2003-2017 | 5.91 | 1.03, 11.02 | - | - | - | - | - | - | 5.91 |
| 50+y | 2003-2006 | 22.96 | 5.72, 43 | 2006-2017 | 3.97 | 1.88, 6.11 | - | - | - | 7.78 |
| New Zealand | | | | | | | | | | |
| 20-49y | 2003-2015 | 2.57 | 0.99, 4.17 | 2015-2017 | -11.93 | -32.25, 14.47 | - | - | - | 0.36 |
| 50+y | 2003-2017 | 1.18 | 0.24, 2.12 | - | - | - | - | - | - | 1.18 |
| Norway | | | | | | | | | | |
| 20-49y | 2003-2017 | 4.39 | 2.76, 6.05 | - | - | - | - | - | - | 4.39 |
| 50+y | 2003-2017 | 2.22 | 1.64, 2.79 | - | - | - | - | - | - | 2.22 |
| Philippines | | | | | | | | | | |
| 20-49y | 2003-2017 | -5.35 | -8.25, -2.37 | - | - | - | - | - | - | -5.35 |

**Appendix Table 8: Segment specific annual percentage change (APC) for cancer incidence trends from 2003 to 2017 by country: kidney cancer**

| | Joinpoint trend 1 | | | Joinpoint trend 2 | | | Joinpoint trend 3 | | | |
|---|---|---|---|---|---|---|---|---|---|---|
| Country | Years | APC | 95% CI | Years | APC | 95% CI | Years | APC | 95% CI | AAPC |
| 50+y | 2003-2011 | -2.38 | -5.12, 0.44 | 2011-2017 | 2.62 | -1.81, 7.25 | - | - | - | -0.27 |
| Poland | | | | | | | | | | |
| 20-49y | 2003-2017 | -1.76 | -4.23, 0.78 | - | - | - | - | - | - | -1.76 |
| 50+y | 2003-2017 | 1.14 | -0.41, 2.72 | - | - | - | - | - | - | 1.14 |
| Qatar | | | | | | | | | | |
| 20-49y | - | - | - | - | - | - | - | - | - | - |
| 50+y | 2003-2017 | 1.97 | -5.65, 10.2 | - | - | - | - | - | - | 1.97 |
| Republic of Korea | | | | | | | | | | |
| 20-49y | 2003-2017 | 5.41 | 4.65, 6.18 | - | - | - | - | - | - | 5.41 |
| 50+y | 2003-2008 | 8.60 | 7.39, 9.81 | 2008-2017 | 1.71 | 1.24, 2.17 | - | - | - | 4.12 |
| Slovenia | | | | | | | | | | |
| 20-49y | 2003-2005 | 25.79 | -27.56, 118.44 | 2005-2017 | -3.15 | -6.26, 0.06 | - | - | - | 0.54 |
| 50+y | 2003-2010 | 3.95 | 1.46, 6.5 | 2010-2017 | -0.81 | -3.19, 1.62 | - | - | - | 1.54 |
| Sweden | | | | | | | | | | |
| 20-49y | 2003-2017 | 3.75 | 1.94, 5.59 | - | - | - | - | - | - | 3.75 |
| 50+y | 2003-2017 | 1.63 | 1.03, 2.24 | - | - | - | - | - | - | 1.63 |
| Switzerland | | | | | | | | | | |
| 20-49y | 2003-2005 | 31.40 | -9.68, 91.17 | 2005-2015 | -0.57 | -3.91, 2.89 | 2015-2017 | -18.16 | -43.74, 19.07 | 0.63 |
| 50+y | 2003-2017 | 0.97 | 0.2, 1.74 | - | - | - | - | - | - | 0.97 |
| Thailand | | | | | | | | | | |
| 20-49y | 2003-2017 | 0.34 | -2.85, 3.64 | - | - | - | - | - | - | 0.34 |
| 50+y | 2003-2005 | 35.40 | -11.3, 106.7 | 2005-2017 | 2.98 | 0.43, 5.59 | - | - | - | 7.08 |
| The Netherlands | | | | | | | | | | |
| 20-49y | 2003-2017 | 2.12 | 1.25, 3.01 | - | - | - | - | - | - | 2.12 |
| 50+y | 2003-2007 | 3.88 | 0.67, 7.2 | 2007-2017 | 0.58 | -0.19, 1.36 | - | - | - | 1.51 |
| Türkiye | | | | | | | | | | |
| 20-49y | 2003-2017 | 5.78 | 4.63, 6.94 | - | - | - | - | - | - | 5.78 |
| 50+y | 2003-2017 | 2.83 | 1.71, 3.95 | - | - | - | - | - | - | 2.83 |
| UK | | | | | | | | | | |
| 20-49y | 2003-2005 | -0.24 | -10.53, 11.22 | 2005-2017 | 5.75 | 5.07, 6.43 | - | - | - | 4.87 |
| 50+y | 2003-2014 | 3.98 | 3.46, 4.49 | 2014-2017 | 0.00 | -3.6, 3.73 | - | - | - | 3.11 |
| USA | | | | | | | | | | |
| 20-49y | 2003-2017 | 2.21 | 1.56, 2.87 | - | - | - | - | - | - | 2.21 |
| 50+y | 2003-2007 | 3.23 | 1.12, 5.38 | 2007-2017 | -0.05 | -0.56, 0.46 | - | - | - | 0.88 |
| Uganda | | | | | | | | | | |
| 20-49y | 2003-2017 | 2.72 | -2.79, 8.54 | - | - | - | - | - | - | 2.72 |
| 50+y | - | - | - | - | - | - | - | - | - | - |

#### Appendix Table 9: Segment specific annual percentage change (APC) for cancer incidence trends from 2003 to 2017 by country: leukaemia cancer

| | Joinpoint trend 1 | | | Joinpoint trend 2 | | | Joinpoint trend 3 | | | |
|---|---|---|---|---|---|---|---|---|---|---|
| Country | Years | APC | 95% CI | Years | APC | 95% CI | Years | APC | 95% CI | AAPC |
| Argentina | | | | | | | | | | |
| 20-49y | 2003-2017 | -0.63 | -2.92, 1.71 | - | - | - | - | - | - | -0.63 |
| 50+y | 2003-2017 | -1.67 | -3.26, -0.06 | - | - | - | - | - | - | -1.67 |
| Australia | | | | | | | | | | |
| 20-49y | 2003-2017 | 0.95 | 0.26, 1.65 | - | - | - | - | - | - | 0.95 |
| 50+y | 2003-2010 | -0.33 | -1.57, 0.92 | 2010-2017 | 3.34 | 2.07, 4.64 | - | - | - | 1.49 |
| Austria | | | | | | | | | | |
| 20-49y | 2003-2017 | -1.57 | -2.99, -0.13 | - | - | - | - | - | - | -1.57 |
| 50+y | 2003-2017 | -1.97 | -2.62, -1.32 | - | - | - | - | - | - | -1.97 |
| Bahrain | | | | | | | | | | |
| 20-49y | 2003-2008 | -20.31 | -30.71, -8.34 | 2008-2011 | 45.27 | -22.28, 171.51 | 2011-2017 | -11.26 | -20.16, -1.36 | -5.09 |
| 50+y | 2003-2017 | -2.79 | -6.76, 1.35 | - | - | - | - | - | - | -2.79 |
| Belarus | | | | | | | | | | |
| 20-49y | 2003-2017 | -0.42 | -1.68, 0.86 | - | - | - | - | - | - | -0.42 |
| 50+y | 2003-2017 | 0.72 | -0.46, 1.91 | - | - | - | - | - | - | 0.72 |
| Canada | | | | | | | | | | |
| 20-49y | 2003-2017 | 1.42 | 0.99, 1.86 | - | - | - | - | - | - | 1.42 |
| 50+y | 2003-2017 | 1.67 | 0.98, 2.37 | - | - | - | - | - | - | 1.67 |
| Chile | | | | | | | | | | |
| 20-49y | - | - | - | - | - | - | - | - | - | - |
| 50+y | 2003-2005 | 66.57 | -30.86, 301.29 | 2005-2017 | -1.17 | -6.18, 4.1 | - | - | - | 6.48 |
| China | | | | | | | | | | |
| 20-49y | 2003-2011 | -3.59 | -7.05, 0.01 | 2011-2017 | 2.71 | -2.95, 8.71 | - | - | - | -0.93 |
| 50+y | 2003-2017 | -0.14 | -0.74, 0.46 | - | - | - | - | - | - | -0.14 |
| Colombia | | | | | | | | | | |
| 20-49y | 2003-2012 | -2.72 | -7.72, 2.55 | 2012-2017 | 12.38 | -1.24, 27.88 | - | - | - | 2.43 |
| 50+y | 2003-2017 | 1.69 | -0.52, 3.94 | - | - | - | - | - | - | 1.69 |
| Croatia | | | | | | | | | | |
| 20-49y | 2003-2011 | -6.39 | -9.84, -2.81 | 2011-2017 | 9.22 | 3.05, 15.76 | - | - | - | 0.01 |
| 50+y | 2003-2011 | -3.48 | -5.54, -1.37 | 2011-2017 | 3.64 | 0.23, 7.16 | - | - | - | -0.49 |
| Cyprus | | | | | | | | | | |
| 20-49y | 2003-2017 | 1.52 | -2.42, 5.63 | - | - | - | - | - | - | 1.52 |
| 50+y | 2003-2017 | -2.80 | -4.79, -0.77 | - | - | - | - | - | - | -2.80 |
| Czechia | | | | | | | | | | |
| 20-49y | 2003-2017 | 0.74 | -0.38, 1.87 | - | - | - | - | - | - | 0.74 |
| 50+y | 2003-2010 | 2.69 | 1.04, 4.36 | 2010-2017 | -2.11 | -3.68, -0.52 | - | - | - | 0.26 |
| Denmark | | | | | | | | | | |
| 20-49y | 2003-2017 | 1.21 | -0.33, 2.78 | - | - | - | - | - | - | 1.21 |
| 50+y | 2003-2017 | 0.64 | 0.14, 1.15 | - | - | - | - | - | - | 0.64 |
| Ecuador | | | | | | | | | | |
| 20-49y | 2003-2017 | 1.52 | -0.57, 3.65 | - | - | - | - | - | - | 1.52 |
| 50+y | 2003-2017 | 2.32 | -0.04, 4.74 | - | - | - | - | - | - | 2.32 |
| Estonia | | | | | | | | | | |
| 20-49y | 2003-2017 | 2.48 | -0.01, 5.03 | - | - | - | - | - | - | 2.48 |

**Appendix Table 9: Segment specific annual percentage change (APC) for cancer incidence trends from 2003 to 2017 by country: leukaemia cancer**

| | Joinpoint trend 1 | | | Joinpoint trend 2 | | | Joinpoint trend 3 | | | |
|---|---|---|---|---|---|---|---|---|---|---|
| Country | Years | APC | 95% CI | Years | APC | 95% CI | Years | APC | 95% CI | AAPC |
| 50+y | 2003-2017 | -0.19 | -1.14, 0.76 | - | - | - | - | - | - | -0.19 |
| Finland | | | | | | | | | | |
| 20-49y | 2003-2017 | 0.85 | -1.25, 3 | - | - | - | - | - | - | 0.85 |
| 50+y | 2003-2017 | 0.30 | -0.65, 1.27 | - | - | - | - | - | - | 0.30 |
| France | | | | | | | | | | |
| 20-49y | 2003-2017 | 2.60 | 1.15, 4.06 | - | - | - | - | - | - | 2.60 |
| 50+y | 2003-2017 | 0.83 | 0.35, 1.31 | - | - | - | - | - | - | 0.83 |
| Germany | | | | | | | | | | |
| 20-49y | 2003-2007 | -4.74 | -12.32, 3.5 | 2007-2017 | 2.75 | 0.67, 4.87 | - | - | - | 0.55 |
| 50+y | 2003-2017 | 0.76 | 0.09, 1.43 | - | - | - | - | - | - | 0.76 |
| Iceland | | | | | | | | | | |
| 20-49y | 2003-2005 | 62.02 | -26.73, 258.29 | 2005-2017 | -4.15 | -8.54, 0.46 | - | - | - | 3.32 |
| 50+y | 2003-2017 | -1.93 | -4.99, 1.23 | - | - | - | - | - | - | -1.93 |
| India | | | | | | | | | | |
| 20-49y | 2003-2017 | 0.78 | -0.11, 1.67 | - | - | - | - | - | - | 0.78 |
| 50+y | 2003-2015 | 1.56 | 0.28, 2.87 | 2015-2017 | 11.54 | -10.1, 38.39 | - | - | - | 2.93 |
| Ireland | | | | | | | | | | |
| 20-49y | 2003-2017 | 0.42 | -1.44, 2.32 | - | - | - | - | - | - | 0.42 |
| 50+y | 2003-2017 | -1.78 | -2.67, -0.87 | - | - | - | - | - | - | -1.78 |
| Israel | | | | | | | | | | |
| 20-49y | 2003-2017 | -1.42 | -3.09, 0.28 | - | - | - | - | - | - | -1.42 |
| 50+y | 2003-2017 | -2.66 | -3.44, -1.87 | - | - | - | - | - | - | -2.66 |
| Italy | | | | | | | | | | |
| 20-49y | 2003-2017 | -0.41 | -2.39, 1.6 | - | - | - | - | - | - | -0.41 |
| 50+y | 2003-2017 | -1.68 | -2.86, -0.49 | - | - | - | - | - | - | -1.68 |
| Kuwait | | | | | | | | | | |
| 20-49y | 2003-2017 | 1.81 | -2.7, 6.52 | - | - | - | - | - | - | 1.81 |
| 50+y | 2003-2017 | -0.12 | -2.89, 2.74 | - | - | - | - | - | - | -0.12 |
| Latvia | | | | | | | | | | |
| 20-49y | 2003-2017 | 2.89 | -0.32, 6.21 | - | - | - | - | - | - | 2.89 |
| 50+y | 2003-2007 | 4.12 | -5.84, 15.13 | 2007-2010 | 20.62 | -12.24, 65.77 | 2010-2017 | -6.97 | -10.84, -2.93 | 1.57 |
| Lithuania | | | | | | | | | | |
| 20-49y | 2003-2017 | 0.78 | -1.05, 2.64 | - | - | - | - | - | - | 0.78 |
| 50+y | 2003-2006 | -5.03 | -14.41, 5.37 | 2006-2017 | 2.16 | 0.74, 3.6 | - | - | - | 0.57 |
| Malta | | | | | | | | | | |
| 20-49y | 2003-2008 | -24.74 | -40.64, -4.59 | 2008-2017 | 12.61 | 2.21, 24.06 | - | - | - | -2.49 |
| 50+y | 2003-2017 | 4.20 | 1.25, 7.24 | - | - | - | - | - | - | 4.20 |
| New Zealand | | | | | | | | | | |
| 20-49y | 2003-2017 | -0.99 | -2.41, 0.45 | - | - | - | - | - | - | -0.99 |
| 50+y | 2003-2006 | -16.39 | -20.15, -12.46 | 2006-2017 | 0.96 | 0.33, 1.58 | - | - | - | -3.04 |
| Norway | | | | | | | | | | |
| 20-49y | 2003-2017 | 1.51 | -0.7, 3.76 | - | - | - | - | - | - | 1.51 |
| 50+y | 2003-2017 | 1.22 | 0.45, 2 | - | - | - | - | - | - | 1.22 |
| Philippines | | | | | | | | | | |
| 20-49y | 2003-2012 | -5.44 | -7.51, -3.33 | 2012-2017 | 1.65 | -3.71, 7.3 | - | - | - | -2.97 |

**Appendix Table 9: Segment specific annual percentage change (APC) for cancer incidence trends from 2003 to 2017 by country: leukaemia cancer**

| | Joinpoint trend 1 | | | Joinpoint trend 2 | | | Joinpoint trend 3 | | | |
|---|---|---|---|---|---|---|---|---|---|---|
| Country | Years | APC | 95% CI | Years | APC | 95% CI | Years | APC | 95% CI | AAPC |
| 50+y | 2003-2017 | -3.14 | -4.4, -1.86 | - | - | - | - | - | - | -3.14 |
| Poland | | | | | | | | | | |
| 20-49y | 2003-2017 | 0.94 | -1.34, 3.27 | - | - | - | - | - | - | 0.94 |
| 50+y | 2003-2017 | -0.62 | -1.88, 0.65 | - | - | - | - | - | - | -0.62 |
| Qatar | | | | | | | | | | |
| 20-49y | - | - | - | - | - | - | - | - | - | - |
| 50+y | 2003-2017 | 1.64 | -6.7, 10.73 | - | - | - | - | - | - | 1.64 |
| Republic of Korea | | | | | | | | | | |
| 20-49y | 2003-2017 | 1.38 | 0.92, 1.85 | - | - | - | - | - | - | 1.38 |
| 50+y | 2003-2017 | 1.20 | 0.85, 1.54 | - | - | - | - | - | - | 1.20 |
| Slovenia | | | | | | | | | | |
| 20-49y | 2003-2017 | 0.30 | -2.24, 2.91 | - | - | - | - | - | - | 0.30 |
| 50+y | 2003-2006 | 7.00 | 0.63, 13.78 | 2006-2017 | -0.49 | -1.31, 0.34 | - | - | - | 1.07 |
| Sweden | | | | | | | | | | |
| 20-49y | 2003-2017 | 0.63 | -0.97, 2.25 | - | - | - | - | - | - | 0.63 |
| 50+y | 2003-2017 | 0.47 | -0.09, 1.04 | - | - | - | - | - | - | 0.47 |
| Switzerland | | | | | | | | | | |
| 20-49y | 2003-2017 | 0.01 | -1.88, 1.92 | - | - | - | - | - | - | 0.01 |
| 50+y | 2003-2017 | -0.76 | -1.76, 0.25 | - | - | - | - | - | - | -0.76 |
| Thailand | | | | | | | | | | |
| 20-49y | 2003-2017 | 0.30 | -1.07, 1.68 | - | - | - | - | - | - | 0.30 |
| 50+y | 2003-2017 | 0.84 | -0.32, 2.03 | - | - | - | - | - | - | 0.84 |
| The Netherlands | | | | | | | | | | |
| 20-49y | 2003-2017 | 0.41 | -0.44, 1.27 | - | - | - | - | - | - | 0.41 |
| 50+y | 2003-2012 | -0.39 | -1.1, 0.32 | 2012-2017 | 4.93 | 3.11, 6.79 | - | - | - | 1.48 |
| Türkiye | | | | | | | | | | |
| 20-49y | 2003-2017 | 0.88 | -0.28, 2.05 | - | - | - | - | - | - | 0.88 |
| 50+y | 2003-2017 | 0.79 | 0.06, 1.53 | - | - | - | - | - | - | 0.79 |
| UK | | | | | | | | | | |
| 20-49y | 2003-2017 | 1.29 | 0.73, 1.84 | - | - | - | - | - | - | 1.29 |
| 50+y | 2003-2015 | 1.88 | 1.08, 2.68 | 2015-2017 | -5.47 | -17.24, 7.97 | - | - | - | 0.79 |
| USA | | | | | | | | | | |
| 20-49y | 2003-2017 | 1.60 | 0.95, 2.26 | - | - | - | - | - | - | 1.60 |
| 50+y | 2003-2008 | -0.46 | -0.96, 0.05 | 2008-2011 | 4.41 | 2.08, 6.8 | 2011-2017 | -1.17 | -1.55, -0.79 | 0.26 |
| Uganda | | | | | | | | | | |
| 20-49y | 2003-2017 | 1.51 | -3.38, 6.64 | - | - | - | - | - | - | 1.51 |
| 50+y | 2003-2017 | 2.78 | -2.79, 8.68 | - | - | - | - | - | - | 2.78 |

**Appendix Table 10: Segment specific annual percentage change (APC) for cancer incidence trends from 2003 to 2017 by country: liver cancer**

| | Joinpoint trend 1 | | | Joinpoint trend 2 | | | Joinpoint trend 3 | | | |
|---|---|---|---|---|---|---|---|---|---|---|
| Country | Years | APC | 95% CI | Years | APC | 95% CI | Years | APC | 95% CI | AAPC |
| Argentina | | | | | | | | | | |
| 20-49y | 2003-2005 | 146.33 | -45.98, 1023.14 | 2005-2017 | -6.15 | -14.2, 2.66 | - | - | - | 7.72 |
| 50+y | 2003-2017 | 4.90 | 3.38, 6.43 | - | - | - | - | - | - | 4.90 |
| Australia | | | | | | | | | | |
| 20-49y | 2003-2017 | 0.72 | -0.39, 1.85 | - | - | - | - | - | - | 0.72 |
| 50+y | 2003-2015 | 5.50 | 4.81, 6.2 | 2015-2017 | -0.04 | -10.55, 11.72 | - | - | - | 4.69 |
| Austria | | | | | | | | | | |
| 20-49y | 2003-2017 | -3.05 | -4.57, -1.5 | - | - | - | - | - | - | -3.05 |
| 50+y | 2003-2017 | -0.57 | -1.23, 0.09 | - | - | - | - | - | - | -0.57 |
| Bahrain | | | | | | | | | | |
| 20-49y | - | - | - | - | - | - | - | - | - | - |
| 50+y | 2003-2005 | -35.89 | -75.03, 64.56 | 2005-2017 | 4.49 | -1.17, 10.48 | - | - | - | -2.55 |
| Belarus | | | | | | | | | | |
| 20-49y | 2003-2017 | 2.24 | -0.06, 4.59 | - | - | - | - | - | - | 2.24 |
| 50+y | 2003-2017 | 2.88 | 1.86, 3.9 | - | - | - | - | - | - | 2.88 |
| Canada | | | | | | | | | | |
| 20-49y | 2003-2017 | 1.16 | -0.24, 2.58 | - | - | - | - | - | - | 1.16 |
| 50+y | 2003-2015 | 5.92 | 5.11, 6.74 | 2015-2017 | -4.16 | -15.85, 9.15 | - | - | - | 4.42 |
| Chile | | | | | | | | | | |
| 20-49y | - | - | - | - | - | - | - | - | - | - |
| 50+y | 2003-2017 | 1.82 | -2.13, 5.92 | - | - | - | - | - | - | 1.82 |
| China | | | | | | | | | | |
| 20-49y | 2003-2017 | -3.03 | -3.83, -2.22 | - | - | - | - | - | - | -3.03 |
| 50+y | 2003-2017 | -1.80 | -2.23, -1.38 | - | - | - | - | - | - | -1.80 |
| Colombia | | | | | | | | | | |
| 20-49y | 2003-2017 | -3.93 | -7.96, 0.27 | - | - | - | - | - | - | -3.93 |
| 50+y | 2003-2017 | 1.23 | -0.46, 2.96 | - | - | - | - | - | - | 1.23 |
| Croatia | | | | | | | | | | |
| 20-49y | 2003-2017 | -4.30 | -7.12, -1.4 | - | - | - | - | - | - | -4.30 |
| 50+y | 2003-2017 | 0.72 | 0.05, 1.39 | - | - | - | - | - | - | 0.72 |
| Cyprus | | | | | | | | | | |
| 20-49y | - | - | - | - | - | - | - | - | - | - |
| 50+y | 2003-2017 | 2.46 | 0.46, 4.49 | - | - | - | - | - | - | 2.46 |
| Czechia | | | | | | | | | | |
| 20-49y | 2003-2017 | -3.43 | -6.45, -0.32 | - | - | - | - | - | - | -3.43 |
| 50+y | 2003-2017 | -0.23 | -0.76, 0.31 | - | - | - | - | - | - | -0.23 |
| Denmark | | | | | | | | | | |
| 20-49y | 2003-2017 | -0.49 | -4.61, 3.81 | - | - | - | - | - | - | -0.49 |
| 50+y | 2003-2005 | -9.08 | -25.94, 11.61 | 2005-2014 | 5.54 | 3.2, 7.92 | 2014-2017 | -3.56 | -12.96, 6.86 | 1.34 |
| Ecuador | | | | | | | | | | |
| 20-49y | 2003-2017 | -0.14 | -8.6, 9.09 | - | - | - | - | - | - | -0.14 |
| 50+y | 2003-2012 | 5.69 | 3.09, 8.36 | 2012-2017 | -2.71 | -8.48, 3.42 | - | - | - | 2.61 |
| Estonia | | | | | | | | | | |
| 20-49y | 2003-2017 | -2.85 | -7.84, 2.41 | - | - | - | - | - | - | -2.85 |

**Appendix Table 10: Segment specific annual percentage change (APC) for cancer incidence trends from 2003 to 2017 by country: liver cancer**

| | Joinpoint trend 1 | | | Joinpoint trend 2 | | | Joinpoint trend 3 | | | |
|---|---|---|---|---|---|---|---|---|---|---|
| **Country** | **Years** | **APC** | **95% CI** | **Years** | **APC** | **95% CI** | **Years** | **APC** | **95% CI** | **AAPC** |
| 50+y | 2003-2017 | 3.77 | 1.82, 5.76 | - | - | - | - | - | - | 3.77 |
| Finland | | | | | | | | | | |
| 20-49y | 2003-2017 | 0.48 | -3.08, 4.17 | - | - | - | - | - | - | 0.48 |
| 50+y | 2003-2005 | 13.62 | -5.97, 37.3 | 2005-2017 | 0.89 | -0.24, 2.02 | - | - | - | 2.61 |
| France | | | | | | | | | | |
| 20-49y | 2003-2006 | -13.31 | -30.85, 8.69 | 2006-2009 | 23.12 | -21.67, 93.52 | 2009-2017 | -9.33 | -13.69, -4.74 | -4.11 |
| 50+y | 2003-2009 | 3.23 | 1.25, 5.25 | 2009-2017 | 0.11 | -1.13, 1.38 | - | - | - | 1.44 |
| Germany | | | | | | | | | | |
| 20-49y | 2003-2017 | 0.36 | -2.32, 3.12 | - | - | - | - | - | - | 0.36 |
| 50+y | 2003-2014 | 2.44 | 1.44, 3.45 | 2014-2017 | -5.42 | -12.09, 1.74 | - | - | - | 0.70 |
| Iceland | | | | | | | | | | |
| 20-49y | - | - | - | - | - | - | - | - | - | - |
| 50+y | 2003-2017 | 6.25 | 3.38, 9.2 | - | - | - | - | - | - | 6.25 |
| India | | | | | | | | | | |
| 20-49y | 2003-2017 | 1.48 | -0.84, 3.85 | - | - | - | - | - | - | 1.48 |
| 50+y | 2003-2017 | 3.47 | 2.46, 4.49 | - | - | - | - | - | - | 3.47 |
| Ireland | | | | | | | | | | |
| 20-49y | 2003-2012 | 15.13 | 7.38, 23.45 | 2012-2017 | -10.79 | -24.79, 5.82 | - | - | - | 5.11 |
| 50+y | 2003-2017 | 4.92 | 3.94, 5.91 | - | - | - | - | - | - | 4.92 |
| Israel | | | | | | | | | | |
| 20-49y | 2003-2017 | 1.67 | -2.01, 5.48 | - | - | - | - | - | - | 1.67 |
| 50+y | 2003-2017 | 0.27 | -0.96, 1.51 | - | - | - | - | - | - | 0.27 |
| Italy | | | | | | | | | | |
| 20-49y | 2003-2017 | -1.88 | -4.48, 0.78 | - | - | - | - | - | - | -1.88 |
| 50+y | 2003-2017 | -2.19 | -2.72, -1.66 | - | - | - | - | - | - | -2.19 |
| Kuwait | | | | | | | | | | |
| 20-49y | - | - | - | - | - | - | - | - | - | - |
| 50+y | 2003-2017 | -0.54 | -3.34, 2.34 | - | - | - | - | - | - | -0.54 |
| Latvia | | | | | | | | | | |
| 20-49y | 2003-2017 | 1.15 | -3.03, 5.52 | - | - | - | - | - | - | 1.15 |
| 50+y | 2003-2017 | 2.74 | 1.46, 4.04 | - | - | - | - | - | - | 2.74 |
| Lithuania | | | | | | | | | | |
| 20-49y | 2003-2017 | 8.03 | 4.33, 11.87 | - | - | - | - | - | - | 8.03 |
| 50+y | 2003-2017 | 3.87 | 2.56, 5.21 | - | - | - | - | - | - | 3.87 |
| Malta | | | | | | | | | | |
| 20-49y | - | - | - | - | - | - | - | - | - | - |
| 50+y | 2003-2017 | 9.77 | 3, 16.98 | - | - | - | - | - | - | 9.77 |
| New Zealand | | | | | | | | | | |
| 20-49y | 2003-2017 | -0.87 | -3.14, 1.45 | - | - | - | - | - | - | -0.87 |
| 50+y | 2003-2017 | 3.40 | 2.34, 4.48 | - | - | - | - | - | - | 3.40 |
| Norway | | | | | | | | | | |
| 20-49y | 2003-2017 | 1.88 | -2.31, 6.25 | - | - | - | - | - | - | 1.88 |
| 50+y | 2003-2017 | 7.02 | 6.08, 7.96 | - | - | - | - | - | - | 7.02 |
| Philippines | | | | | | | | | | |
| 20-49y | 2003-2017 | -5.31 | -6.61, -3.99 | - | - | - | - | - | - | -5.31 |

**Appendix Table 10: Segment specific annual percentage change (APC) for cancer incidence trends from 2003 to 2017 by country: liver cancer**

| | Joinpoint trend 1 | | | Joinpoint trend 2 | | | Joinpoint trend 3 | | | |
|---|---|---|---|---|---|---|---|---|---|---|
| **Country** | **Years** | **APC** | **95% CI** | **Years** | **APC** | **95% CI** | **Years** | **APC** | **95% CI** | **AAPC** |
| 50+y | 2003-2017 | -2.61 | -3.77, -1.43 | - | - | - | - | - | - | -2.61 |
| Poland | | | | | | | | | | |
| 20-49y | 2003-2017 | 2.30 | -3.72, 8.69 | - | - | - | - | - | - | 2.30 |
| 50+y | 2003-2008 | -8.32 | -19.94, 4.99 | 2008-2017 | 8.35 | 2.52, 14.52 | - | - | - | 2.07 |
| Qatar | | | | | | | | | | |
| 20-49y | - | - | - | - | - | - | - | - | - | - |
| 50+y | 2003-2017 | -3.61 | -9.15, 2.28 | - | - | - | - | - | - | -3.61 |
| Republic of Korea | | | | | | | | | | |
| 20-49y | 2003-2009 | -3.57 | -4.6, -2.54 | 2009-2017 | -5.51 | -6.16, -4.86 | - | - | - | -4.68 |
| 50+y | 2003-2005 | 0.39 | -2.21, 3.06 | 2005-2011 | -2.11 | -2.68, -1.53 | 2011-2017 | -4.05 | -4.47, -3.62 | -2.59 |
| Slovenia | | | | | | | | | | |
| 20-49y | 2003-2017 | -2.05 | -8.14, 4.44 | - | - | - | - | - | - | -2.05 |
| 50+y | 2003-2017 | 2.32 | 0.97, 3.7 | - | - | - | - | - | - | 2.32 |
| Sweden | | | | | | | | | | |
| 20-49y | 2003-2017 | 2.31 | -0.69, 5.4 | - | - | - | - | - | - | 2.31 |
| 50+y | 2003-2006 | -6.13 | -15.49, 4.26 | 2006-2014 | 6.76 | 3.8, 9.8 | 2014-2017 | -0.63 | -10.54, 10.37 | 2.27 |
| Switzerland | | | | | | | | | | |
| 20-49y | 2003-2017 | -4.67 | -8.22, -0.99 | - | - | - | - | - | - | -4.67 |
| 50+y | 2003-2017 | 1.02 | -0.03, 2.08 | - | - | - | - | - | - | 1.02 |
| Thailand | | | | | | | | | | |
| 20-49y | 2003-2017 | -1.33 | -2.28, -0.38 | - | - | - | - | - | - | -1.33 |
| 50+y | 2003-2005 | -7.53 | -19.98, 6.85 | 2005-2017 | -0.23 | -1.08, 0.63 | - | - | - | -1.31 |
| The Netherlands | | | | | | | | | | |
| 20-49y | 2003-2017 | 3.63 | 2.05, 5.23 | - | - | - | - | - | - | 3.63 |
| 50+y | 2003-2005 | -2.72 | -17.99, 15.41 | 2005-2017 | 7.53 | 6.45, 8.62 | - | - | - | 6.00 |
| Türkiye | | | | | | | | | | |
| 20-49y | 2003-2017 | -3.06 | -6.08, 0.05 | - | - | - | - | - | - | -3.06 |
| 50+y | 2003-2017 | 0.72 | -0.63, 2.09 | - | - | - | - | - | - | 0.72 |
| UK | | | | | | | | | | |
| 20-49y | 2003-2017 | 2.89 | 1.61, 4.18 | - | - | - | - | - | - | 2.89 |
| 50+y | 2003-2013 | 6.24 | 5.71, 6.77 | 2013-2017 | 1.82 | -0.22, 3.9 | - | - | - | 4.96 |
| USA | | | | | | | | | | |
| 20-49y | 2003-2017 | -2.81 | -3.78, -1.82 | - | - | - | - | - | - | -2.81 |
| 50+y | 2003-2009 | 5.92 | 5.22, 6.63 | 2009-2015 | 2.77 | 1.86, 3.68 | 2015-2017 | -2.76 | -6.52, 1.16 | 3.29 |
| Uganda | | | | | | | | | | |
| 20-49y | 2003-2006 | 25.92 | -8.39, 73.09 | 2006-2017 | -5.25 | -9.23, -1.09 | - | - | - | 0.71 |
| 50+y | 2003-2017 | -2.97 | -5.42, -0.45 | - | - | - | - | - | - | -2.97 |

**Appendix Table 11: Segment specific annual percentage change (APC) for cancer incidence trends from 2003 to 2017 by country: oesophagus cancer**

| | Joinpoint trend 1 | | | Joinpoint trend 2 | | | Joinpoint trend 3 | | | |
|---|---|---|---|---|---|---|---|---|---|---|
| **Country** | **Years** | **APC** | **95% CI** | **Years** | **APC** | **95% CI** | **Years** | **APC** | **95% CI** | **AAPC** |
| Argentina | | | | | | | | | | |
| 20-49y | 2003-2017 | -2.50 | -6.96, 2.17 | - | - | - | - | - | - | -2.50 |
| 50+y | 2003-2017 | -3.10 | -4.57, -1.61 | - | - | - | - | - | - | -3.10 |
| Australia | | | | | | | | | | |
| 20-49y | 2003-2017 | -1.02 | -2.4, 0.37 | - | - | - | - | - | - | -1.02 |
| 50+y | 2003-2017 | -0.23 | -0.58, 0.12 | - | - | - | - | - | - | -0.23 |
| Austria | | | | | | | | | | |
| 20-49y | 2003-2017 | -0.76 | -4.74, 3.4 | - | - | - | - | - | - | -0.76 |
| 50+y | 2003-2017 | -0.25 | -0.96, 0.47 | - | - | - | - | - | - | -0.25 |
| Bahrain | | | | | | | | | | |
| 20-49y | - | - | - | - | - | - | - | - | - | - |
| 50+y | 2003-2017 | -8.65 | -14.94, -1.89 | - | - | - | - | - | - | -8.65 |
| Belarus | | | | | | | | | | |
| 20-49y | 2003-2017 | 3.42 | 1.59, 5.29 | - | - | - | - | - | - | 3.42 |
| 50+y | 2003-2017 | 1.66 | 0.94, 2.39 | - | - | - | - | - | - | 1.66 |
| Canada | | | | | | | | | | |
| 20-49y | 2003-2017 | -1.27 | -3, 0.5 | - | - | - | - | - | - | -1.27 |
| 50+y | 2003-2005 | -3.57 | -12.08, 5.76 | 2005-2009 | 3.61 | -1.06, 8.51 | 2009-2017 | -0.40 | -1.4, 0.61 | 0.27 |
| Chile | | | | | | | | | | |
| 20-49y | - | - | - | - | - | - | - | - | - | - |
| 50+y | 2003-2017 | -3.66 | -6.43, -0.81 | - | - | - | - | - | - | -3.66 |
| China | | | | | | | | | | |
| 20-49y | 2003-2017 | -5.70 | -7.57, -3.79 | - | - | - | - | - | - | -5.70 |
| 50+y | 2003-2017 | -2.79 | -3.41, -2.17 | - | - | - | - | - | - | -2.79 |
| Colombia | | | | | | | | | | |
| 20-49y | - | - | - | - | - | - | - | - | - | - |
| 50+y | 2003-2010 | -7.64 | -11.9, -3.19 | 2010-2017 | 0.52 | -4.11, 5.37 | - | - | - | -3.65 |
| Croatia | | | | | | | | | | |
| 20-49y | 2003-2017 | -0.84 | -3.27, 1.64 | - | - | - | - | - | - | -0.84 |
| 50+y | 2003-2017 | -1.90 | -2.84, -0.96 | - | - | - | - | - | - | -1.90 |
| Cyprus | | | | | | | | | | |
| 20-49y | - | - | - | - | - | - | - | - | - | - |
| 50+y | 2003-2017 | 0.76 | -5.86, 7.84 | - | - | - | - | - | - | 0.76 |
| Czechia | | | | | | | | | | |
| 20-49y | 2003-2017 | -3.08 | -6.4, 0.36 | - | - | - | - | - | - | -3.08 |
| 50+y | 2003-2013 | 2.57 | 1.2, 3.97 | 2013-2017 | -1.73 | -6.96, 3.8 | - | - | - | 1.33 |
| Denmark | | | | | | | | | | |
| 20-49y | 2003-2017 | -4.63 | -9.09, 0.04 | - | - | - | - | - | - | -4.63 |
| 50+y | 2003-2017 | 1.72 | 0.78, 2.67 | - | - | - | - | - | - | 1.72 |
| Ecuador | | | | | | | | | | |
| 20-49y | - | - | - | - | - | - | - | - | - | - |
| 50+y | 2003-2017 | -3.04 | -6.78, 0.85 | - | - | - | - | - | - | -3.04 |
| Estonia | | | | | | | | | | |
| 20-49y | 2003-2017 | 2.36 | -3.19, 8.24 | - | - | - | - | - | - | 2.36 |

**Appendix Table 11: Segment specific annual percentage change (APC) for cancer incidence trends from 2003 to 2017 by country: oesophagus cancer**

| | Joinpoint trend 1 | | | Joinpoint trend 2 | | | Joinpoint trend 3 | | | |
|---|---|---|---|---|---|---|---|---|---|---|
| **Country** | **Years** | **APC** | **95% CI** | **Years** | **APC** | **95% CI** | **Years** | **APC** | **95% CI** | **AAPC** |
| 50+y | 2003-2017 | 1.90 | 0.78, 3.02 | - | - | - | - | - | - | 1.90 |
| Finland | | | | | | | | | | |
| 20-49y | 2003-2017 | 0.84 | -5.52, 7.64 | - | - | - | - | - | - | 0.84 |
| 50+y | 2003-2015 | 0.89 | 0.04, 1.75 | 2015-2017 | 11.25 | -3.63, 28.42 | - | - | - | 2.31 |
| France | | | | | | | | | | |
| 20-49y | 2003-2017 | -4.69 | -6.88, -2.44 | - | - | - | - | - | - | -4.69 |
| 50+y | 2003-2017 | -1.13 | -1.87, -0.38 | - | - | - | - | - | - | -1.13 |
| Germany | | | | | | | | | | |
| 20-49y | 2003-2017 | 0.65 | -1.41, 2.75 | - | - | - | - | - | - | 0.65 |
| 50+y | 2003-2017 | 1.06 | 0.25, 1.88 | - | - | - | - | - | - | 1.06 |
| Iceland | | | | | | | | | | |
| 20-49y | - | - | - | - | - | - | - | - | - | - |
| 50+y | 2003-2017 | 1.43 | -2.51, 5.53 | - | - | - | - | - | - | 1.43 |
| India | | | | | | | | | | |
| 20-49y | 2003-2017 | -3.38 | -4.82, -1.91 | - | - | - | - | - | - | -3.38 |
| 50+y | 2003-2017 | -2.81 | -3.4, -2.21 | - | - | - | - | - | - | -2.81 |
| Ireland | | | | | | | | | | |
| 20-49y | 2003-2005 | 36.57 | -3.21, 92.72 | 2005-2015 | -5.49 | -8.42, -2.48 | 2015-2017 | 16.33 | -17.56, 64.15 | 2.61 |
| 50+y | 2003-2014 | -1.13 | -2.05, -0.2 | 2014-2017 | 3.75 | -3.2, 11.19 | - | - | - | -0.10 |
| Israel | | | | | | | | | | |
| 20-49y | 2003-2017 | -1.01 | -5.58, 3.78 | - | - | - | - | - | - | -1.01 |
| 50+y | 2003-2017 | -1.30 | -2.51, -0.06 | - | - | - | - | - | - | -1.30 |
| Italy | | | | | | | | | | |
| 20-49y | 2003-2017 | 1.26 | -6.05, 9.15 | - | - | - | - | - | - | 1.26 |
| 50+y | 2003-2017 | -2.59 | -3.97, -1.19 | - | - | - | - | - | - | -2.59 |
| Kuwait | | | | | | | | | | |
| 20-49y | - | - | - | - | - | - | - | - | - | - |
| 50+y | 2003-2017 | 1.55 | -3.33, 6.68 | - | - | - | - | - | - | 1.55 |
| Latvia | | | | | | | | | | |
| 20-49y | 2003-2017 | 2.51 | -1.48, 6.67 | - | - | - | - | - | - | 2.51 |
| 50+y | 2003-2017 | 1.89 | 0.56, 3.24 | - | - | - | - | - | - | 1.89 |
| Lithuania | | | | | | | | | | |
| 20-49y | 2003-2017 | 0.85 | -1.06, 2.81 | - | - | - | - | - | - | 0.85 |
| 50+y | 2003-2017 | 1.69 | 0.64, 2.75 | - | - | - | - | - | - | 1.69 |
| Malta | | | | | | | | | | |
| 20-49y | - | - | - | - | - | - | - | - | - | - |
| 50+y | 2003-2017 | -1.50 | -5.08, 2.22 | - | - | - | - | - | - | -1.50 |
| New Zealand | | | | | | | | | | |
| 20-49y | 2003-2017 | -0.16 | -4.3, 4.16 | - | - | - | - | - | - | -0.16 |
| 50+y | 2003-2017 | -1.27 | -2.2, -0.33 | - | - | - | - | - | - | -1.27 |
| Norway | | | | | | | | | | |
| 20-49y | 2003-2017 | 1.66 | -1.73, 5.16 | - | - | - | - | - | - | 1.66 |
| 50+y | 2003-2017 | 1.78 | 0.92, 2.65 | - | - | - | - | - | - | 1.78 |
| Philippines | | | | | | | | | | |
| 20-49y | 2003-2005 | -47.17 | -77.06, 21.66 | 2005-2017 | -0.81 | -5.58, 4.21 | - | - | - | -9.34 |

**Appendix Table 11: Segment specific annual percentage change (APC) for cancer incidence trends from 2003 to 2017 by country: oesophagus cancer**

| | Joinpoint trend 1 | | | Joinpoint trend 2 | | | Joinpoint trend 3 | | | |
|---|---|---|---|---|---|---|---|---|---|---|
| Country | Years | APC | 95% CI | Years | APC | 95% CI | Years | APC | 95% CI | AAPC |
| 50+y | 2003-2017 | -2.90 | -4.98, -0.77 | - | - | - | - | - | - | -2.90 |
| Poland | | | | | | | | | | |
| 20-49y | - | - | - | - | - | - | - | - | - | - |
| 50+y | 2003-2017 | 1.09 | -0.88, 3.09 | - | - | - | - | - | - | 1.09 |
| Qatar | | | | | | | | | | |
| 20-49y | - | - | - | - | - | - | - | - | - | - |
| 50+y | - | - | - | - | - | - | - | - | - | - |
| Republic of Korea | | | | | | | | | | |
| 20-49y | 2003-2017 | -1.02 | -1.98, -0.05 | - | - | - | - | - | - | -1.02 |
| 50+y | 2003-2017 | -2.29 | -2.42, -2.15 | - | - | - | - | - | - | -2.29 |
| Slovenia | | | | | | | | | | |
| 20-49y | 2003-2017 | -2.66 | -8.6, 3.67 | - | - | - | - | - | - | -2.66 |
| 50+y | 2003-2017 | -1.99 | -3.91, -0.02 | - | - | - | - | - | - | -1.99 |
| Sweden | | | | | | | | | | |
| 20-49y | 2003-2017 | -0.07 | -2.78, 2.71 | - | - | - | - | - | - | -0.07 |
| 50+y | 2003-2017 | 0.07 | -0.88, 1.02 | - | - | - | - | - | - | 0.07 |
| Switzerland | | | | | | | | | | |
| 20-49y | 2003-2017 | -4.93 | -10.61, 1.11 | - | - | - | - | - | - | -4.93 |
| 50+y | 2003-2017 | -0.30 | -1.27, 0.68 | - | - | - | - | - | - | -0.30 |
| Thailand | | | | | | | | | | |
| 20-49y | 2003-2017 | 6.79 | 3.2, 10.51 | - | - | - | - | - | - | 6.79 |
| 50+y | 2003-2015 | 0.93 | -0.77, 2.67 | 2015-2017 | -12.68 | -34.54, 16.48 | - | - | - | -1.13 |
| The Netherlands | | | | | | | | | | |
| 20-49y | 2003-2017 | -0.39 | -1.84, 1.09 | - | - | - | - | - | - | -0.39 |
| 50+y | 2003-2017 | 2.02 | 1.51, 2.54 | - | - | - | - | - | - | 2.02 |
| Türkiye | | | | | | | | | | |
| 20-49y | 2003-2017 | -4.77 | -8.04, -1.37 | - | - | - | - | - | - | -4.77 |
| 50+y | 2003-2017 | -2.11 | -3.3, -0.91 | - | - | - | - | - | - | -2.11 |
| UK | | | | | | | | | | |
| 20-49y | 2003-2017 | -0.99 | -1.69, -0.29 | - | - | - | - | - | - | -0.99 |
| 50+y | 2003-2007 | 1.17 | 0.11, 2.24 | 2007-2015 | 0.12 | -0.33, 0.56 | 2015-2017 | -1.76 | -4.99, 1.57 | 0.14 |
| USA | | | | | | | | | | |
| 20-49y | 2003-2017 | -1.25 | -2.55, 0.07 | - | - | - | - | - | - | -1.25 |
| 50+y | 2003-2017 | -0.96 | -1.48, -0.44 | - | - | - | - | - | - | -0.96 |
| Uganda | | | | | | | | | | |
| 20-49y | 2003-2017 | 1.63 | -1.78, 5.16 | - | - | - | - | - | - | 1.63 |
| 50+y | 2003-2017 | 1.75 | -0.41, 3.96 | - | - | - | - | - | - | 1.75 |

**Appendix Table 12: Segment specific annual percentage change (APC) for cancer incidence trends from 2003 to 2017 by country: oral cancer**

| | Joinpoint trend 1 | | | Joinpoint trend 2 | | | Joinpoint trend 3 | | | |
|---|---|---|---|---|---|---|---|---|---|---|
| Country | Years | APC | 95% CI | Years | APC | 95% CI | Years | APC | 95% CI | AAPC |
| Argentina | | | | | | | | | | |
| 20-49y | 2003-2008 | -18.30 | -35.18, 2.99 | 2008-2017 | 8.56 | -1.23, 19.33 | - | - | - | -1.92 |
| 50+y | 2003-2017 | -1.06 | -3.61, 1.56 | - | - | - | - | - | - | -1.06 |
| Australia | | | | | | | | | | |
| 20-49y | 2003-2013 | -0.03 | -1.59, 1.56 | 2013-2017 | -5.42 | -11.29, 0.83 | - | - | - | -1.60 |
| 50+y | 2003-2005 | -2.49 | -10.46, 6.19 | 2005-2015 | 1.71 | 0.92, 2.51 | 2015-2017 | -4.00 | -11.85, 4.54 | 0.27 |
| Austria | | | | | | | | | | |
| 20-49y | 2003-2017 | -2.80 | -3.82, -1.77 | - | - | - | - | - | - | -2.80 |
| 50+y | 2003-2017 | 0.61 | -0.14, 1.36 | - | - | - | - | - | - | 0.61 |
| Bahrain | | | | | | | | | | |
| 20-49y | 2003-2017 | -0.52 | -6.04, 5.33 | - | - | - | - | - | - | -0.52 |
| 50+y | 2003-2017 | 0.49 | -7.37, 9.02 | - | - | - | - | - | - | 0.49 |
| Belarus | | | | | | | | | | |
| 20-49y | 2003-2010 | 1.86 | -0.17, 3.93 | 2010-2017 | 5.65 | 3.55, 7.8 | - | - | - | 3.74 |
| 50+y | 2003-2011 | 1.18 | 0.23, 2.14 | 2011-2017 | 3.16 | 1.66, 4.67 | - | - | - | 2.02 |
| Canada | | | | | | | | | | |
| 20-49y | 2003-2017 | 0.15 | -0.45, 0.76 | - | - | - | - | - | - | 0.15 |
| 50+y | 2003-2017 | 1.84 | 1.37, 2.31 | - | - | - | - | - | - | 1.84 |
| Chile | | | | | | | | | | |
| 20-49y | - | - | - | - | - | - | - | - | - | - |
| 50+y | 2003-2017 | -1.78 | -6.42, 3.1 | - | - | - | - | - | - | -1.78 |
| China | | | | | | | | | | |
| 20-49y | 2003-2017 | 0.73 | -0.02, 1.47 | - | - | - | - | - | - | 0.73 |
| 50+y | 2003-2008 | -3.14 | -6.44, 0.27 | 2008-2017 | 1.69 | 0.26, 3.14 | - | - | - | -0.06 |
| Colombia | | | | | | | | | | |
| 20-49y | 2003-2017 | -0.67 | -2.65, 1.36 | - | - | - | - | - | - | -0.67 |
| 50+y | 2003-2012 | -2.04 | -3.76, -0.29 | 2012-2017 | 3.14 | -1.24, 7.72 | - | - | - | -0.22 |
| Croatia | | | | | | | | | | |
| 20-49y | 2003-2017 | -5.11 | -6.19, -4.02 | - | - | - | - | - | - | -5.11 |
| 50+y | 2003-2005 | -7.40 | -15.33, 1.27 | 2005-2017 | -0.66 | -1.18, -0.13 | - | - | - | -1.65 |
| Cyprus | | | | | | | | | | |
| 20-49y | 2003-2017 | 4.71 | -0.21, 9.87 | - | - | - | - | - | - | 4.71 |
| 50+y | 2003-2017 | 3.12 | 1.14, 5.14 | - | - | - | - | - | - | 3.12 |
| Czechia | | | | | | | | | | |
| 20-49y | 2003-2010 | -3.52 | -5.97, -0.99 | 2010-2017 | 1.37 | -1.21, 4.02 | - | - | - | -1.10 |
| 50+y | 2003-2017 | 1.94 | 1.59, 2.29 | - | - | - | - | - | - | 1.94 |
| Denmark | | | | | | | | | | |
| 20-49y | 2003-2017 | -1.81 | -2.83, -0.79 | - | - | - | - | - | - | -1.81 |
| 50+y | 2003-2012 | 3.23 | 1.83, 4.65 | 2012-2017 | -0.46 | -3.73, 2.92 | - | - | - | 1.90 |
| Ecuador | | | | | | | | | | |
| 20-49y | 2003-2010 | 13.78 | -0.11, 29.6 | 2010-2017 | -5.25 | -16.82, 7.93 | - | - | - | 3.83 |
| 50+y | 2003-2017 | 0.62 | -2.99, 4.36 | - | - | - | - | - | - | 0.62 |
| Estonia | | | | | | | | | | |
| 20-49y | 2003-2012 | -2.78 | -7.27, 1.93 | 2012-2017 | 14.21 | 1.72, 28.23 | - | - | - | 2.98 |

**Appendix Table 12: Segment specific annual percentage change (APC) for cancer incidence trends from 2003 to 2017 by country: oral cancer**

| | Joinpoint trend 1 | | | Joinpoint trend 2 | | | Joinpoint trend 3 | | | |
|---|---|---|---|---|---|---|---|---|---|---|
| Country | Years | APC | 95% CI | Years | APC | 95% CI | Years | APC | 95% CI | AAPC |
| 50+y | 2003-2017 | 1.22 | 0.09, 2.37 | - | - | - | - | - | - | 1.22 |
| Finland | | | | | | | | | | |
| 20-49y | 2003-2017 | 0.82 | -1.29, 2.97 | - | - | - | - | - | - | 0.82 |
| 50+y | 2003-2017 | 1.89 | 1.47, 2.31 | - | - | - | - | - | - | 1.89 |
| France | | | | | | | | | | |
| 20-49y | 2003-2017 | -5.05 | -5.91, -4.19 | - | - | - | - | - | - | -5.05 |
| 50+y | 2003-2017 | -0.75 | -1.22, -0.29 | - | - | - | - | - | - | -0.75 |
| Germany | | | | | | | | | | |
| 20-49y | 2003-2014 | -6.46 | -7.23, -5.69 | 2014-2017 | 7.36 | 1.01, 14.1 | - | - | - | -3.66 |
| 50+y | 2003-2011 | 1.38 | 0.06, 2.72 | 2011-2017 | -1.63 | -3.61, 0.39 | - | - | - | 0.08 |
| Iceland | | | | | | | | | | |
| 20-49y | - | - | - | - | - | - | - | - | - | - |
| 50+y | 2003-2017 | -0.72 | -3.42, 2.06 | - | - | - | - | - | - | -0.72 |
| India | | | | | | | | | | |
| 20-49y | 2003-2017 | 2.32 | 1.36, 3.28 | - | - | - | - | - | - | 2.32 |
| 50+y | 2003-2017 | 0.13 | -0.32, 0.58 | - | - | - | - | - | - | 0.13 |
| Ireland | | | | | | | | | | |
| 20-49y | 2003-2017 | 2.60 | 0.56, 4.68 | - | - | - | - | - | - | 2.60 |
| 50+y | 2003-2017 | 2.48 | 1.69, 3.29 | - | - | - | - | - | - | 2.48 |
| Israel | | | | | | | | | | |
| 20-49y | 2003-2017 | -1.36 | -2.9, 0.2 | - | - | - | - | - | - | -1.36 |
| 50+y | 2003-2017 | 0.39 | -0.52, 1.32 | - | - | - | - | - | - | 0.39 |
| Italy | | | | | | | | | | |
| 20-49y | 2003-2017 | -3.00 | -5.39, -0.55 | - | - | - | - | - | - | -3.00 |
| 50+y | 2003-2017 | -1.73 | -2.64, -0.82 | - | - | - | - | - | - | -1.73 |
| Kuwait | | | | | | | | | | |
| 20-49y | 2003-2017 | -3.92 | -8.92, 1.35 | - | - | - | - | - | - | -3.92 |
| 50+y | 2003-2012 | 6.43 | 1.48, 11.63 | 2012-2017 | -13.43 | -22.97, -2.71 | - | - | - | -1.14 |
| Latvia | | | | | | | | | | |
| 20-49y | 2003-2017 | 4.14 | 1.8, 6.53 | - | - | - | - | - | - | 4.14 |
| 50+y | 2003-2008 | -2.04 | -6.38, 2.5 | 2008-2017 | 3.61 | 1.71, 5.54 | - | - | - | 1.55 |
| Lithuania | | | | | | | | | | |
| 20-49y | 2003-2008 | -4.06 | -8.02, 0.08 | 2008-2017 | 6.97 | 5.14, 8.83 | - | - | - | 2.89 |
| 50+y | 2003-2013 | -1.61 | -3.67, 0.49 | 2013-2017 | 6.30 | -2.44, 15.82 | - | - | - | 0.59 |
| Malta | | | | | | | | | | |
| 20-49y | 2003-2017 | -0.21 | -5.37, 5.24 | - | - | - | - | - | - | -0.21 |
| 50+y | 2003-2005 | 24.54 | -9.26, 70.95 | 2005-2017 | -1.48 | -3.31, 0.38 | - | - | - | 1.88 |
| New Zealand | | | | | | | | | | |
| 20-49y | 2003-2017 | 0.40 | -0.67, 1.48 | - | - | - | - | - | - | 0.40 |
| 50+y | 2003-2017 | 2.49 | 1.49, 3.5 | - | - | - | - | - | - | 2.49 |
| Norway | | | | | | | | | | |
| 20-49y | 2003-2017 | 1.98 | 0.31, 3.68 | - | - | - | - | - | - | 1.98 |
| 50+y | 2003-2017 | 1.86 | 1.3, 2.42 | - | - | - | - | - | - | 1.86 |
| Philippines | | | | | | | | | | |
| 20-49y | 2003-2017 | -3.30 | -5.03, -1.54 | - | - | - | - | - | - | -3.30 |

**Appendix Table 12: Segment specific annual percentage change (APC) for cancer incidence trends from 2003 to 2017 by country: oral cancer**

| | Joinpoint trend 1 | | | Joinpoint trend 2 | | | Joinpoint trend 3 | | | |
|---|---|---|---|---|---|---|---|---|---|---|
| **Country** | **Years** | **APC** | **95% CI** | **Years** | **APC** | **95% CI** | **Years** | **APC** | **95% CI** | **AAPC** |
| 50+y | 2003-2017 | -3.68 | -5.27, -2.07 | - | - | - | - | - | - | -3.68 |
| Poland | | | | | | | | | | |
| 20-49y | 2003-2015 | 4.06 | 1.44, 6.74 | 2015-2017 | -32.79 | -56.31, 3.39 | - | - | - | -2.24 |
| 50+y | 2003-2017 | 2.23 | 1.1, 3.36 | - | - | - | - | - | - | 2.23 |
| Qatar | | | | | | | | | | |
| 20-49y | - | - | - | - | - | - | - | - | - | - |
| 50+y | - | - | - | - | - | - | - | - | - | - |
| Republic of Korea | | | | | | | | | | |
| 20-49y | 2003-2017 | 1.97 | 1.52, 2.43 | - | - | - | - | - | - | 1.97 |
| 50+y | 2003-2017 | 0.63 | 0.38, 0.87 | - | - | - | - | - | - | 0.63 |
| Slovenia | | | | | | | | | | |
| 20-49y | 2003-2017 | -2.04 | -3.73, -0.31 | - | - | - | - | - | - | -2.04 |
| 50+y | 2003-2017 | -0.27 | -1.03, 0.5 | - | - | - | - | - | - | -0.27 |
| Sweden | | | | | | | | | | |
| 20-49y | 2003-2017 | 0.73 | -0.25, 1.72 | - | - | - | - | - | - | 0.73 |
| 50+y | 2003-2017 | 2.31 | 1.84, 2.79 | - | - | - | - | - | - | 2.31 |
| Switzerland | | | | | | | | | | |
| 20-49y | 2003-2017 | -4.28 | -6.59, -1.91 | - | - | - | - | - | - | -4.28 |
| 50+y | 2003-2005 | -4.14 | -12.76, 5.32 | 2005-2008 | 4.22 | -5.14, 14.51 | 2008-2017 | -1.90 | -2.73, -1.05 | -0.94 |
| Thailand | | | | | | | | | | |
| 20-49y | 2003-2017 | 2.17 | 0.99, 3.35 | - | - | - | - | - | - | 2.17 |
| 50+y | 2003-2011 | -3.36 | -4.74, -1.97 | 2011-2014 | 7.84 | -5.43, 22.97 | 2014-2017 | -5.41 | -11.42, 1.01 | -1.52 |
| The Netherlands | | | | | | | | | | |
| 20-49y | 2003-2017 | -2.32 | -3.48, -1.14 | - | - | - | - | - | - | -2.32 |
| 50+y | 2003-2014 | 1.11 | 0.66, 1.56 | 2014-2017 | -2.07 | -5.28, 1.24 | - | - | - | 0.42 |
| Türkiye | | | | | | | | | | |
| 20-49y | 2003-2017 | -0.49 | -1.86, 0.89 | - | - | - | - | - | - | -0.49 |
| 50+y | 2003-2017 | -1.04 | -2.24, 0.18 | - | - | - | - | - | - | -1.04 |
| UK | | | | | | | | | | |
| 20-49y | 2003-2012 | 2.91 | 2.17, 3.66 | 2012-2017 | -0.03 | -1.79, 1.76 | - | - | - | 1.85 |
| 50+y | 2003-2013 | 3.84 | 3.34, 4.34 | 2013-2017 | 0.86 | -1.1, 2.86 | - | - | - | 2.98 |
| USA | | | | | | | | | | |
| 20-49y | 2003-2017 | -0.42 | -1.01, 0.17 | - | - | - | - | - | - | -0.42 |
| 50+y | 2003-2017 | 0.88 | 0.72, 1.04 | - | - | - | - | - | - | 0.88 |
| Uganda | | | | | | | | | | |
| 20-49y | 2003-2017 | 4.77 | 0.18, 9.58 | - | - | - | - | - | - | 4.77 |
| 50+y | 2003-2017 | -0.67 | -4.6, 3.41 | - | - | - | - | - | - | -0.67 |

#### Appendix Table 13: Segment specific annual percentage change (APC) for cancer incidence trends from 2003 to 2017 by country: pancreas cancer

| | Joinpoint trend 1 | | | Joinpoint trend 2 | | | Joinpoint trend 3 | | | |
|---|---|---|---|---|---|---|---|---|---|---|
| Country | Years | APC | 95% CI | Years | APC | 95% CI | Years | APC | 95% CI | AAPC |
| Argentina | | | | | | | | | | |
| 20-49y | 2003-2017 | 0.90 | -2.56, 4.48 | - | - | - | - | - | - | 0.90 |
| 50+y | 2003-2005 | 18.43 | -2.87, 44.41 | 2005-2017 | 0.04 | -1.12, 1.22 | - | - | - | 2.49 |
| Australia | | | | | | | | | | |
| 20-49y | 2003-2017 | 3.14 | 2.02, 4.27 | - | - | - | - | - | - | 3.14 |
| 50+y | 2003-2017 | 1.18 | 0.81, 1.55 | - | - | - | - | - | - | 1.18 |
| Austria | | | | | | | | | | |
| 20-49y | 2003-2017 | 0.09 | -1.66, 1.86 | - | - | - | - | - | - | 0.09 |
| 50+y | 2003-2017 | 0.96 | 0.58, 1.34 | - | - | - | - | - | - | 0.96 |
| Bahrain | | | | | | | | | | |
| 20-49y | - | - | - | - | - | - | - | - | - | - |
| 50+y | 2003-2017 | -1.66 | -4.58, 1.36 | - | - | - | - | - | - | -1.66 |
| Belarus | | | | | | | | | | |
| 20-49y | 2003-2017 | 1.01 | -0.37, 2.4 | - | - | - | - | - | - | 1.01 |
| 50+y | 2003-2017 | 2.49 | 2.01, 2.96 | - | - | - | - | - | - | 2.49 |
| Canada | | | | | | | | | | |
| 20-49y | 2003-2011 | 4.66 | 2.45, 6.92 | 2011-2017 | -1.15 | -4.36, 2.17 | - | - | - | 2.13 |
| 50+y | 2003-2010 | -0.64 | -1.71, 0.44 | 2010-2013 | 5.88 | -2.35, 14.8 | 2013-2017 | -0.27 | -2.79, 2.31 | 0.83 |
| Chile | | | | | | | | | | |
| 20-49y | - | - | - | - | - | - | - | - | - | - |
| 50+y | 2003-2017 | 0.40 | -3.71, 4.69 | - | - | - | - | - | - | 0.40 |
| China | | | | | | | | | | |
| 20-49y | 2003-2017 | -1.25 | -3.15, 0.68 | - | - | - | - | - | - | -1.25 |
| 50+y | 2003-2017 | 1.07 | 0.67, 1.46 | - | - | - | - | - | - | 1.07 |
| Colombia | | | | | | | | | | |
| 20-49y | 2003-2006 | -23.49 | -43.15, 2.96 | 2006-2017 | 7.53 | 3.3, 11.92 | - | - | - | -0.04 |
| 50+y | 2003-2012 | 0.29 | -1.38, 1.98 | 2012-2017 | 5.86 | 1.6, 10.3 | - | - | - | 2.24 |
| Croatia | | | | | | | | | | |
| 20-49y | 2003-2017 | -1.56 | -4.02, 0.97 | - | - | - | - | - | - | -1.56 |
| 50+y | 2003-2017 | 1.50 | 0.95, 2.04 | - | - | - | - | - | - | 1.50 |
| Cyprus | | | | | | | | | | |
| 20-49y | 2003-2017 | 11.86 | 4.32, 19.94 | - | - | - | - | - | - | 11.86 |
| 50+y | 2003-2007 | 11.18 | -0.59, 24.35 | 2007-2017 | 1.41 | -1.35, 4.24 | - | - | - | 4.11 |
| Czechia | | | | | | | | | | |
| 20-49y | 2003-2017 | -0.16 | -1.37, 1.07 | - | - | - | - | - | - | -0.16 |
| 50+y | 2003-2017 | 0.39 | -0.03, 0.82 | - | - | - | - | - | - | 0.39 |
| Denmark | | | | | | | | | | |
| 20-49y | 2003-2017 | -0.53 | -2.54, 1.53 | - | - | - | - | - | - | -0.53 |
| 50+y | 2003-2009 | 1.40 | -0.66, 3.5 | 2009-2017 | -1.16 | -2.46, 0.16 | - | - | - | -0.07 |
| Ecuador | | | | | | | | | | |
| 20-49y | - | - | - | - | - | - | - | - | - | - |
| 50+y | 2003-2017 | 1.14 | -1.04, 3.36 | - | - | - | - | - | - | 1.14 |
| Estonia | | | | | | | | | | |
| 20-49y | 2003-2015 | 0.11 | -2.89, 3.19 | 2015-2017 | -26.38 | -55.95, 23.05 | - | - | - | -4.19 |

**Appendix Table 13: Segment specific annual percentage change (APC) for cancer incidence trends from 2003 to 2017 by country: pancreas cancer**

| | Joinpoint trend 1 | | | Joinpoint trend 2 | | | Joinpoint trend 3 | | | |
|---|---|---|---|---|---|---|---|---|---|---|
| **Country** | **Years** | **APC** | **95% CI** | **Years** | **APC** | **95% CI** | **Years** | **APC** | **95% CI** | **AAPC** |
| 50+y | 2003-2017 | 1.02 | -0.12, 2.18 | - | - | - | - | - | - | 1.02 |
| Finland | | | | | | | | | | |
| 20-49y | 2003-2017 | -1.09 | -2.73, 0.57 | - | - | - | - | - | - | -1.09 |
| 50+y | 2003-2017 | 0.57 | 0.05, 1.1 | - | - | - | - | - | - | 0.57 |
| France | | | | | | | | | | |
| 20-49y | 2003-2017 | 3.07 | 1.08, 5.09 | - | - | - | - | - | - | 3.07 |
| 50+y | 2003-2005 | 9.05 | 1.59, 17.04 | 2005-2012 | 3.20 | 1.98, 4.45 | 2012-2017 | 0.67 | -0.92, 2.27 | 3.10 |
| Germany | | | | | | | | | | |
| 20-49y | 2003-2015 | 2.74 | 0.23, 5.32 | 2015-2017 | -19.09 | -46.78, 23.02 | - | - | - | -0.71 |
| 50+y | 2003-2017 | 0.82 | 0.35, 1.31 | - | - | - | - | - | - | 0.82 |
| Iceland | | | | | | | | | | |
| 20-49y | - | - | - | - | - | - | - | - | - | - |
| 50+y | 2003-2017 | 0.48 | -1.85, 2.86 | - | - | - | - | - | - | 0.48 |
| India | | | | | | | | | | |
| 20-49y | 2003-2017 | 1.14 | -0.62, 2.94 | - | - | - | - | - | - | 1.14 |
| 50+y | 2003-2017 | 3.70 | 2.68, 4.72 | - | - | - | - | - | - | 3.70 |
| Ireland | | | | | | | | | | |
| 20-49y | 2003-2017 | 1.93 | -1.11, 5.07 | - | - | - | - | - | - | 1.93 |
| 50+y | 2003-2006 | 7.40 | 1.11, 14.07 | 2006-2017 | -0.64 | -1.44, 0.17 | - | - | - | 1.03 |
| Israel | | | | | | | | | | |
| 20-49y | 2003-2017 | -2.21 | -3.8, -0.6 | - | - | - | - | - | - | -2.21 |
| 50+y | 2003-2013 | 1.16 | 0.13, 2.19 | 2013-2017 | -2.70 | -6.64, 1.4 | - | - | - | 0.04 |
| Italy | | | | | | | | | | |
| 20-49y | 2003-2017 | 0.17 | -2.65, 3.07 | - | - | - | - | - | - | 0.17 |
| 50+y | 2003-2017 | 1.21 | 0.65, 1.78 | - | - | - | - | - | - | 1.21 |
| Kuwait | | | | | | | | | | |
| 20-49y | 2003-2017 | 4.40 | -3.25, 12.66 | - | - | - | - | - | - | 4.40 |
| 50+y | 2003-2017 | 1.53 | -1.42, 4.56 | - | - | - | - | - | - | 1.53 |
| Latvia | | | | | | | | | | |
| 20-49y | 2003-2017 | -0.29 | -3.25, 2.75 | - | - | - | - | - | - | -0.29 |
| 50+y | 2003-2017 | 1.45 | 0.83, 2.08 | - | - | - | - | - | - | 1.45 |
| Lithuania | | | | | | | | | | |
| 20-49y | 2003-2017 | 0.21 | -2.16, 2.64 | - | - | - | - | - | - | 0.21 |
| 50+y | 2003-2017 | 0.97 | 0.45, 1.5 | - | - | - | - | - | - | 0.97 |
| Malta | | | | | | | | | | |
| 20-49y | - | - | - | - | - | - | - | - | - | - |
| 50+y | 2003-2015 | 3.93 | 0.52, 7.46 | 2015-2017 | -21.98 | -55.63, 37.2 | - | - | - | -0.24 |
| New Zealand | | | | | | | | | | |
| 20-49y | 2003-2017 | 2.68 | 0.06, 5.38 | - | - | - | - | - | - | 2.68 |
| 50+y | 2003-2017 | 1.18 | 0.52, 1.86 | - | - | - | - | - | - | 1.18 |
| Norway | | | | | | | | | | |
| 20-49y | 2003-2017 | -2.18 | -4.66, 0.36 | - | - | - | - | - | - | -2.18 |
| 50+y | 2003-2017 | -0.07 | -0.82, 0.68 | - | - | - | - | - | - | -0.07 |
| Philippines | | | | | | | | | | |
| 20-49y | 2003-2017 | -1.64 | -4.18, 0.97 | - | - | - | - | - | - | -1.64 |

**Appendix Table 13: Segment specific annual percentage change (APC) for cancer incidence trends from 2003 to 2017 by country: pancreas cancer**

| | Joinpoint trend 1 | | | Joinpoint trend 2 | | | Joinpoint trend 3 | | | |
|---|---|---|---|---|---|---|---|---|---|---|
| Country | Years | APC | 95% CI | Years | APC | 95% CI | Years | APC | 95% CI | AAPC |
| 50+y | 2003-2009 | -3.03 | -7.13, 1.24 | 2009-2017 | 3.03 | 0.2, 5.94 | - | - | - | 0.39 |
| Poland | | | | | | | | | | |
| 20-49y | 2003-2017 | 3.14 | -0.95, 7.39 | - | - | - | - | - | - | 3.14 |
| 50+y | 2003-2017 | -2.90 | -4.03, -1.76 | - | - | - | - | - | - | -2.90 |
| Qatar | | | | | | | | | | |
| 20-49y | - | - | - | - | - | - | - | - | - | - |
| 50+y | - | - | - | - | - | - | - | - | - | - |
| Republic of Korea | | | | | | | | | | |
| 20-49y | 2003-2009 | -1.05 | -3.89, 1.87 | 2009-2017 | 4.44 | 2.5, 6.42 | - | - | - | 2.05 |
| 50+y | 2003-2005 | 3.30 | -0.9, 7.69 | 2005-2017 | 1.14 | 0.9, 1.39 | - | - | - | 1.45 |
| Slovenia | | | | | | | | | | |
| 20-49y | 2003-2017 | -2.35 | -6.43, 1.92 | - | - | - | - | - | - | -2.35 |
| 50+y | 2003-2007 | 5.18 | -0.22, 10.88 | 2007-2017 | 0.12 | -1.17, 1.43 | - | - | - | 1.54 |
| Sweden | | | | | | | | | | |
| 20-49y | 2003-2008 | -3.07 | -9.81, 4.18 | 2008-2015 | 8.22 | 2.48, 14.28 | 2015-2017 | -11.17 | -35.66, 22.64 | 1.15 |
| 50+y | 2003-2008 | -1.02 | -2.79, 0.78 | 2008-2017 | 3.84 | 3.08, 4.61 | - | - | - | 2.08 |
| Switzerland | | | | | | | | | | |
| 20-49y | 2003-2017 | 3.89 | 1.19, 6.67 | - | - | - | - | - | - | 3.89 |
| 50+y | 2003-2007 | -3.10 | -6.25, 0.16 | 2007-2017 | 2.53 | 1.7, 3.37 | - | - | - | 0.89 |
| Thailand | | | | | | | | | | |
| 20-49y | 2003-2017 | 2.21 | -0.19, 4.68 | - | - | - | - | - | - | 2.21 |
| 50+y | 2003-2017 | 3.52 | 2.23, 4.82 | - | - | - | - | - | - | 3.52 |
| The Netherlands | | | | | | | | | | |
| 20-49y | 2003-2017 | 1.90 | 0.38, 3.44 | - | - | - | - | - | - | 1.90 |
| 50+y | 2003-2008 | 3.68 | 1.26, 6.15 | 2008-2017 | 1.22 | 0.25, 2.2 | - | - | - | 2.09 |
| Türkiye | | | | | | | | | | |
| 20-49y | 2003-2013 | 4.67 | 1.44, 8 | 2013-2017 | -5.71 | -16.98, 7.09 | - | - | - | 1.59 |
| 50+y | 2003-2017 | 2.71 | 1.69, 3.74 | - | - | - | - | - | - | 2.71 |
| UK | | | | | | | | | | |
| 20-49y | 2003-2017 | 2.25 | 1.59, 2.92 | - | - | - | - | - | - | 2.25 |
| 50+y | 2003-2017 | 1.04 | 0.88, 1.2 | - | - | - | - | - | - | 1.04 |
| USA | | | | | | | | | | |
| 20-49y | 2003-2017 | 1.32 | 0.4, 2.25 | - | - | - | - | - | - | 1.32 |
| 50+y | 2003-2017 | 0.73 | 0.49, 0.98 | - | - | - | - | - | - | 0.73 |
| Uganda | | | | | | | | | | |
| 20-49y | 2003-2005 | -46.78 | -76.56, 20.84 | 2005-2008 | 61.89 | -28.7, 267.6 | 2008-2017 | -9.95 | -16.45, -2.95 | -5.28 |
| 50+y | 2003-2017 | 0.06 | -6.57, 7.16 | - | - | - | - | - | - | 0.06 |

**Appendix Table 14: Segment specific annual percentage change (APC) for cancer incidence trends from 2003 to 2017 by country: prostate cancer**

| | Joinpoint trend 1 | | | Joinpoint trend 2 | | | Joinpoint trend 3 | | | |
|---|---|---|---|---|---|---|---|---|---|---|
| **Country** | **Years** | **APC** | **95% CI** | **Years** | **APC** | **95% CI** | **Years** | **APC** | **95% CI** | **AAPC** |
| Argentina | | | | | | | | | | |
| 20-49y | 2003-2017 | 2.34 | -5.33, 10.63 | - | - | - | - | - | - | 2.34 |
| 50+y | 2003-2017 | -1.13 | -2.05, -0.21 | - | - | - | - | - | - | -1.13 |
| Australia | | | | | | | | | | |
| 20-49y | 2003-2009 | 21.29 | 15.88, 26.96 | 2009-2017 | -5.19 | -7.94, -2.35 | - | - | - | 5.37 |
| 50+y | 2003-2008 | 6.05 | 3.47, 8.7 | 2008-2015 | -4.66 | -6.42, -2.87 | 2015-2017 | 1.74 | -8.88, 13.61 | -0.04 |
| Austria | | | | | | | | | | |
| 20-49y | 2003-2017 | -4.31 | -5.3, -3.32 | - | - | - | - | - | - | -4.31 |
| 50+y | 2003-2013 | -4.68 | -5.28, -4.08 | 2013-2017 | 3.81 | 1.19, 6.49 | - | - | - | -2.33 |
| Bahrain | | | | | | | | | | |
| 20-49y | - | - | - | - | - | - | - | - | - | - |
| 50+y | 2003-2017 | 1.73 | -2.61, 6.26 | - | - | - | - | - | - | 1.73 |
| Belarus | | | | | | | | | | |
| 20-49y | 2003-2017 | 10.28 | 7.89, 12.73 | - | - | - | - | - | - | 10.28 |
| 50+y | 2003-2014 | 9.56 | 8.65, 10.49 | 2014-2017 | 2.61 | -3.58, 9.2 | - | - | - | 8.03 |
| Canada | | | | | | | | | | |
| 20-49y | 2003-2009 | 3.98 | 0.15, 7.96 | 2009-2017 | -6.71 | -8.94, -4.42 | - | - | - | -2.27 |
| 50+y | 2003-2007 | 2.33 | -2.64, 7.55 | 2007-2015 | -5.72 | -7.68, -3.71 | 2015-2017 | 4.60 | -10.62, 22.43 | -2.04 |
| Chile | | | | | | | | | | |
| 20-49y | - | - | - | - | - | - | - | - | - | - |
| 50+y | 2003-2017 | -1.88 | -2.67, -1.08 | - | - | - | - | - | - | -1.88 |
| China | | | | | | | | | | |
| 20-49y | 2003-2017 | 12.49 | 5.3, 20.17 | - | - | - | - | - | - | 12.49 |
| 50+y | 2003-2017 | 5.03 | 4.39, 5.67 | - | - | - | - | - | - | 5.03 |
| Colombia | | | | | | | | | | |
| 20-49y | 2003-2017 | -4.73 | -7.98, -1.36 | - | - | - | - | - | - | -4.73 |
| 50+y | 2003-2017 | -0.57 | -1.17, 0.02 | - | - | - | - | - | - | -0.57 |
| Croatia | | | | | | | | | | |
| 20-49y | 2003-2017 | 7.49 | 2.67, 12.54 | - | - | - | - | - | - | 7.49 |
| 50+y | 2003-2009 | 3.85 | 1.8, 5.94 | 2009-2014 | -0.27 | -3.92, 3.51 | 2014-2017 | 12.66 | 6.21, 19.49 | 4.16 |
| Cyprus | | | | | | | | | | |
| 20-49y | 2003-2017 | -2.15 | -9.03, 5.25 | - | - | - | - | - | - | -2.15 |
| 50+y | 2003-2005 | 10.76 | -6.42, 31.09 | 2005-2017 | 0.04 | -0.95, 1.04 | - | - | - | 1.50 |
| Czechia | | | | | | | | | | |
| 20-49y | 2003-2010 | 18.87 | 10.21, 28.22 | 2010-2017 | 3.05 | -4.46, 11.16 | - | - | - | 10.68 |
| 50+y | 2003-2010 | 5.55 | 3.44, 7.7 | 2010-2017 | -0.51 | -2.5, 1.52 | - | - | - | 2.47 |
| Denmark | | | | | | | | | | |
| 20-49y | 2003-2007 | 35.51 | 2.29, 79.53 | 2007-2017 | 0.79 | -5.95, 8.02 | - | - | - | 9.69 |
| 50+y | 2003-2008 | 8.93 | 5.23, 12.77 | 2008-2017 | -2.23 | -3.6, -0.84 | - | - | - | 1.62 |
| Ecuador | | | | | | | | | | |
| 20-49y | 2003-2013 | 20.00 | 11.06, 29.66 | 2013-2017 | -21.82 | -42.91, 7.06 | - | - | - | 6.17 |
| 50+y | 2003-2017 | 1.55 | 0.3, 2.83 | - | - | - | - | - | - | 1.55 |
| Estonia | | | | | | | | | | |
| 20-49y | 2003-2005 | 126.86 | -2.04, 425.36 | 2005-2017 | 6.68 | 1.51, 12.11 | - | - | - | 18.82 |

**Appendix Table 14: Segment specific annual percentage change (APC) for cancer incidence trends from 2003 to 2017 by country: prostate cancer**

| | Joinpoint trend 1 | | | Joinpoint trend 2 | | | Joinpoint trend 3 | | | |
|---|---|---|---|---|---|---|---|---|---|---|
| Country | Years | APC | 95% CI | Years | APC | 95% CI | Years | APC | 95% CI | AAPC |
| 50+y | 2003-2011 | 8.79 | 5.94, 11.71 | 2011-2017 | -2.77 | -6.68, 1.3 | - | - | - | 3.67 |
| Finland | | | | | | | | | | |
| 20-49y | 2003-2017 | -0.15 | -2.52, 2.29 | - | - | - | - | - | - | -0.15 |
| 50+y | 2003-2017 | -1.95 | -3.01, -0.88 | - | - | - | - | - | - | -1.95 |
| France | | | | | | | | | | |
| 20-49y | 2003-2007 | 15.00 | -3.8, 37.47 | 2007-2017 | -5.68 | -9.73, -1.44 | - | - | - | -0.18 |
| 50+y | 2003-2005 | 7.15 | -7.7, 24.39 | 2005-2013 | -6.11 | -7.97, -4.22 | 2013-2017 | 1.35 | -3.32, 6.24 | -2.21 |
| Germany | | | | | | | | | | |
| 20-49y | 2003-2017 | 0.52 | -1.6, 2.68 | - | - | - | - | - | - | 0.52 |
| 50+y | 2003-2017 | -2.15 | -2.9, -1.39 | - | - | - | - | - | - | -2.15 |
| Iceland | | | | | | | | | | |
| 20-49y | - | - | - | - | - | - | - | - | - | - |
| 50+y | 2003-2017 | -3.08 | -4.37, -1.77 | - | - | - | - | - | - | -3.08 |
| India | | | | | | | | | | |
| 20-49y | 2003-2017 | 2.31 | -1.16, 5.91 | - | - | - | - | - | - | 2.31 |
| 50+y | 2003-2017 | 4.35 | 3.87, 4.83 | - | - | - | - | - | - | 4.35 |
| Ireland | | | | | | | | | | |
| 20-49y | 2003-2013 | 10.66 | 6.35, 15.15 | 2013-2017 | -10.65 | -23.96, 5 | - | - | - | 4.10 |
| 50+y | 2003-2011 | 2.69 | 0.9, 4.51 | 2011-2017 | -2.46 | -5.08, 0.24 | - | - | - | 0.45 |
| Israel | | | | | | | | | | |
| 20-49y | 2003-2017 | -5.95 | -9.66, -2.08 | - | - | - | - | - | - | -5.95 |
| 50+y | 2003-2007 | 3.02 | -3.55, 10.04 | 2007-2017 | -6.23 | -7.74, -4.7 | - | - | - | -3.68 |
| Italy | | | | | | | | | | |
| 20-49y | 2003-2017 | -0.73 | -5.32, 4.08 | - | - | - | - | - | - | -0.73 |
| 50+y | 2003-2017 | -0.38 | -0.86, 0.11 | - | - | - | - | - | - | -0.38 |
| Kuwait | | | | | | | | | | |
| 20-49y | - | - | - | - | - | - | - | - | - | - |
| 50+y | 2003-2007 | -6.68 | -12.75, -0.18 | 2007-2010 | 22.00 | -1.39, 50.93 | 2010-2017 | -3.44 | -6.15, -0.65 | 0.54 |
| Latvia | | | | | | | | | | |
| 20-49y | 2003-2017 | 9.25 | 3.16, 15.7 | - | - | - | - | - | - | 9.25 |
| 50+y | 2003-2017 | 4.00 | 3.56, 4.44 | - | - | - | - | - | - | 4.00 |
| Lithuania | | | | | | | | | | |
| 20-49y | 2003-2007 | 49.73 | 15.56, 94.02 | 2007-2017 | 3.14 | -3.23, 9.94 | - | - | - | 14.73 |
| 50+y | 2003-2007 | 22.41 | 5.62, 41.87 | 2007-2017 | -2.80 | -6.27, 0.8 | - | - | - | 3.82 |
| Malta | | | | | | | | | | |
| 20-49y | - | - | - | - | - | - | - | - | - | - |
| 50+y | 2003-2015 | 1.81 | -0.67, 4.36 | 2015-2017 | -16.24 | -44.81, 27.13 | - | - | - | -0.99 |
| New Zealand | | | | | | | | | | |
| 20-49y | 2003-2005 | -27.71 | -46.33, -2.62 | 2005-2009 | 22.04 | 5.15, 41.65 | 2009-2017 | -3.50 | -6.59, -0.31 | -0.97 |
| 50+y | 2003-2017 | -0.62 | -1.48, 0.25 | - | - | - | - | - | - | -0.62 |
| Norway | | | | | | | | | | |
| 20-49y | 2003-2017 | 5.70 | 3.94, 7.5 | - | - | - | - | - | - | 5.70 |
| 50+y | 2003-2007 | 4.98 | 0.22, 9.98 | 2007-2017 | -0.38 | -1.52, 0.76 | - | - | - | 1.12 |
| Philippines | | | | | | | | | | |
| 20-49y | 2003-2017 | -4.62 | -10.19, 1.29 | - | - | - | - | - | - | -4.62 |

**Appendix Table 14: Segment specific annual percentage change (APC) for cancer incidence trends from 2003 to 2017 by country: prostate cancer**

| | Joinpoint trend 1 | | | Joinpoint trend 2 | | | Joinpoint trend 3 | | | |
|---|---|---|---|---|---|---|---|---|---|---|
| **Country** | **Years** | **APC** | **95% CI** | **Years** | **APC** | **95% CI** | **Years** | **APC** | **95% CI** | **AAPC** |
| 50+y | 2003-2006 | 5.74 | -1.51, 13.53 | 2006-2009 | -12.44 | -24.04, 0.93 | 2009-2017 | -0.22 | -1.76, 1.34 | -1.76 |
| Poland | | | | | | | | | | |
| 20-49y | 2003-2017 | 5.21 | -3.1, 14.23 | - | - | - | - | - | - | 5.21 |
| 50+y | 2003-2010 | -2.28 | -5.06, 0.58 | 2010-2017 | 10.95 | 7.8, 14.2 | - | - | - | 4.12 |
| Qatar | | | | | | | | | | |
| 20-49y | - | - | - | - | - | - | - | - | - | - |
| 50+y | 2003-2017 | 6.74 | 1.07, 12.73 | - | - | - | - | - | - | 6.74 |
| Republic of Korea | | | | | | | | | | |
| 20-49y | 2003-2008 | 21.41 | 7.43, 37.22 | 2008-2017 | 0.79 | -4.12, 5.96 | - | - | - | 7.72 |
| 50+y | 2003-2009 | 12.62 | 10.25, 15.03 | 2009-2017 | 1.06 | -0.31, 2.46 | - | - | - | 5.86 |
| Slovenia | | | | | | | | | | |
| 20-49y | 2003-2009 | 27.86 | 5.11, 55.53 | 2009-2017 | -5.55 | -16.77, 7.18 | - | - | - | 7.54 |
| 50+y | 2003-2009 | 9.22 | 5.55, 13.01 | 2009-2017 | -0.54 | -2.71, 1.68 | - | - | - | 3.53 |
| Sweden | | | | | | | | | | |
| 20-49y | 2003-2017 | 4.14 | 2.07, 6.25 | - | - | - | - | - | - | 4.14 |
| 50+y | 2003-2017 | -0.68 | -1.46, 0.12 | - | - | - | - | - | - | -0.68 |
| Switzerland | | | | | | | | | | |
| 20-49y | 2003-2005 | 27.61 | -11.39, 83.77 | 2005-2017 | -0.51 | -2.64, 1.65 | - | - | - | 3.09 |
| 50+y | 2003-2011 | 0.19 | -1.32, 1.72 | 2011-2014 | -8.04 | -19.97, 5.67 | 2014-2017 | 6.43 | -0.71, 14.09 | -0.36 |
| Thailand | | | | | | | | | | |
| 20-49y | - | - | - | - | - | - | - | - | - | - |
| 50+y | 2003-2017 | 3.80 | 2.81, 4.81 | - | - | - | - | - | - | 3.80 |
| The Netherlands | | | | | | | | | | |
| 20-49y | 2003-2006 | 19.90 | 2.39, 40.39 | 2006-2017 | -0.99 | -3.07, 1.14 | - | - | - | 3.16 |
| 50+y | 2003-2011 | 0.50 | -0.89, 1.9 | 2011-2017 | -2.59 | -4.67, -0.48 | - | - | - | -0.84 |
| Türkiye | | | | | | | | | | |
| 20-49y | 2003-2017 | 3.18 | -0.63, 7.14 | - | - | - | - | - | - | 3.18 |
| 50+y | 2003-2007 | 12.76 | 7.43, 18.36 | 2007-2017 | -2.30 | -3.45, -1.12 | - | - | - | 1.79 |
| UK | | | | | | | | | | |
| 20-49y | 2003-2010 | 8.97 | 6.45, 11.55 | 2010-2017 | 3.77 | 1.37, 6.22 | - | - | - | 6.33 |
| 50+y | 2003-2013 | 1.68 | 1.09, 2.27 | 2013-2017 | -0.84 | -3.17, 1.54 | - | - | - | 0.95 |
| USA | | | | | | | | | | |
| 20-49y | 2003-2009 | 3.91 | 0.89, 7.02 | 2009-2017 | -8.54 | -10.27, -6.79 | - | - | - | -3.40 |
| 50+y | 2003-2009 | -0.17 | -2.86, 2.59 | 2009-2014 | -8.76 | -13.29, -3.98 | 2014-2017 | 4.64 | -3.47, 13.43 | -2.35 |
| Uganda | | | | | | | | | | |
| 20-49y | 2003-2017 | -1.13 | -6.57, 4.64 | - | - | - | - | - | - | -1.13 |
| 50+y | 2003-2012 | 5.08 | 1.75, 8.52 | 2012-2017 | -3.13 | -10.48, 4.82 | - | - | - | 2.07 |

#### Appendix Table 15: Segment specific annual percentage change (APC) for cancer incidence trends from 2003 to 2017 by country: stomach cancer

| | Joinpoint trend 1 | | | Joinpoint trend 2 | | | Joinpoint trend 3 | | | |
|---|---|---|---|---|---|---|---|---|---|---|
| Country | Years | APC | 95% CI | Years | APC | 95% CI | Years | APC | 95% CI | AAPC |
| Argentina | | | | | | | | | | |
| 20-49y | 2003-2012 | 2.33 | -4.83, 10.03 | 2012-2017 | -13.71 | -27.76, 3.08 | - | - | - | -3.71 |
| 50+y | 2003-2017 | -0.94 | -1.97, 0.1 | - | - | - | - | - | - | -0.94 |
| Australia | | | | | | | | | | |
| 20-49y | 2003-2017 | 0.93 | -0.56, 2.44 | - | - | - | - | - | - | 0.93 |
| 50+y | 2003-2009 | -2.06 | -3.23, -0.87 | 2009-2017 | -0.11 | -0.88, 0.67 | - | - | - | -0.95 |
| Austria | | | | | | | | | | |
| 20-49y | 2003-2017 | -1.77 | -2.9, -0.63 | - | - | - | - | - | - | -1.77 |
| 50+y | 2003-2017 | -2.97 | -3.37, -2.56 | - | - | - | - | - | - | -2.97 |
| Bahrain | | | | | | | | | | |
| 20-49y | - | - | - | - | - | - | - | - | - | - |
| 50+y | 2003-2017 | -4.11 | -7.4, -0.7 | - | - | - | - | - | - | -4.11 |
| Belarus | | | | | | | | | | |
| 20-49y | 2003-2017 | -3.32 | -4.1, -2.54 | - | - | - | - | - | - | -3.32 |
| 50+y | 2003-2005 | 1.20 | -5, 7.8 | 2005-2017 | -2.57 | -2.93, -2.2 | - | - | - | -2.04 |
| Canada | | | | | | | | | | |
| 20-49y | 2003-2017 | -0.07 | -1.37, 1.24 | - | - | - | - | - | - | -0.07 |
| 50+y | 2003-2011 | -2.29 | -3.14, -1.42 | 2011-2015 | 2.59 | -1.45, 6.8 | 2015-2017 | -3.34 | -10.82, 4.76 | -1.07 |
| Chile | | | | | | | | | | |
| 20-49y | 2003-2017 | -4.01 | -10.4, 2.84 | - | - | - | - | - | - | -4.01 |
| 50+y | 2003-2017 | -1.34 | -2.73, 0.08 | - | - | - | - | - | - | -1.34 |
| China | | | | | | | | | | |
| 20-49y | 2003-2013 | -5.84 | -7.8, -3.84 | 2013-2017 | 0.44 | -7.77, 9.39 | - | - | - | -4.09 |
| 50+y | 2003-2008 | -3.74 | -5.08, -2.39 | 2008-2017 | -1.82 | -2.37, -1.25 | - | - | - | -2.51 |
| Colombia | | | | | | | | | | |
| 20-49y | 2003-2017 | 1.05 | -0.16, 2.28 | - | - | - | - | - | - | 1.05 |
| 50+y | 2003-2005 | 1.08 | -8.82, 12.06 | 2005-2009 | -7.69 | -12.33, -2.81 | 2009-2017 | 0.08 | -1.04, 1.21 | -2.06 |
| Croatia | | | | | | | | | | |
| 20-49y | 2003-2017 | -3.04 | -4.44, -1.61 | - | - | - | - | - | - | -3.04 |
| 50+y | 2003-2006 | -5.91 | -11.89, 0.46 | 2006-2017 | -1.90 | -2.77, -1.03 | - | - | - | -2.78 |
| Cyprus | | | | | | | | | | |
| 20-49y | 2003-2017 | -2.78 | -8.65, 3.47 | - | - | - | - | - | - | -2.78 |
| 50+y | 2003-2008 | 8.00 | 0.97, 15.51 | 2008-2017 | -3.02 | -5.65, -0.32 | - | - | - | 0.78 |
| Czechia | | | | | | | | | | |
| 20-49y | 2003-2017 | -2.60 | -4.01, -1.16 | - | - | - | - | - | - | -2.60 |
| 50+y | 2003-2017 | -3.13 | -3.5, -2.76 | - | - | - | - | - | - | -3.13 |
| Denmark | | | | | | | | | | |
| 20-49y | 2003-2017 | -1.46 | -3.8, 0.93 | - | - | - | - | - | - | -1.46 |
| 50+y | 2003-2013 | -1.23 | -2.16, -0.29 | 2013-2017 | 3.29 | -0.6, 7.34 | - | - | - | 0.04 |
| Ecuador | | | | | | | | | | |
| 20-49y | 2003-2017 | 0.75 | -1.7, 3.26 | - | - | - | - | - | - | 0.75 |
| 50+y | 2003-2005 | 17.17 | -11.49, 55.11 | 2005-2017 | -2.45 | -4.06, -0.82 | - | - | - | 0.14 |
| Estonia | | | | | | | | | | |
| 20-49y | 2003-2017 | -3.39 | -5.25, -1.49 | - | - | - | - | - | - | -3.39 |

**Appendix Table 15: Segment specific annual percentage change (APC) for cancer incidence trends from 2003 to 2017 by country: stomach cancer**

| | Joinpoint trend 1 | | | Joinpoint trend 2 | | | Joinpoint trend 3 | | | |
|---|---|---|---|---|---|---|---|---|---|---|
| **Country** | **Years** | **APC** | **95% CI** | **Years** | **APC** | **95% CI** | **Years** | **APC** | **95% CI** | **AAPC** |
| 50+y | 2003-2017 | -1.94 | -2.51, -1.37 | - | - | - | - | - | - | -1.94 |
| Finland | | | | | | | | | | |
| 20-49y | 2003-2017 | -3.15 | -4.89, -1.38 | - | - | - | - | - | - | -3.15 |
| 50+y | 2003-2017 | -3.21 | -3.79, -2.64 | - | - | - | - | - | - | -3.21 |
| France | | | | | | | | | | |
| 20-49y | 2003-2011 | -2.68 | -5.15, -0.14 | 2011-2017 | 3.95 | -0.11, 8.17 | - | - | - | 0.11 |
| 50+y | 2003-2017 | -1.56 | -2.11, -1.01 | - | - | - | - | - | - | -1.56 |
| Germany | | | | | | | | | | |
| 20-49y | 2003-2017 | -1.44 | -3.31, 0.46 | - | - | - | - | - | - | -1.44 |
| 50+y | 2003-2017 | -1.67 | -2.14, -1.21 | - | - | - | - | - | - | -1.67 |
| Iceland | | | | | | | | | | |
| 20-49y | - | - | - | - | - | - | - | - | - | - |
| 50+y | 2003-2017 | -5.13 | -7.52, -2.68 | - | - | - | - | - | - | -5.13 |
| India | | | | | | | | | | |
| 20-49y | 2003-2017 | -0.97 | -1.84, -0.08 | - | - | - | - | - | - | -0.97 |
| 50+y | 2003-2017 | -0.15 | -1.04, 0.75 | - | - | - | - | - | - | -0.15 |
| Ireland | | | | | | | | | | |
| 20-49y | 2003-2017 | -0.10 | -2.02, 1.87 | - | - | - | - | - | - | -0.10 |
| 50+y | 2003-2015 | -0.07 | -0.65, 0.51 | 2015-2017 | -13.57 | -21.6, -4.72 | - | - | - | -2.12 |
| Israel | | | | | | | | | | |
| 20-49y | 2003-2017 | -1.72 | -3.18, -0.24 | - | - | - | - | - | - | -1.72 |
| 50+y | 2003-2017 | -2.25 | -3.03, -1.46 | - | - | - | - | - | - | -2.25 |
| Italy | | | | | | | | | | |
| 20-49y | 2003-2017 | -3.57 | -6.02, -1.06 | - | - | - | - | - | - | -3.57 |
| 50+y | 2003-2017 | -2.77 | -3.4, -2.14 | - | - | - | - | - | - | -2.77 |
| Kuwait | | | | | | | | | | |
| 20-49y | 2003-2005 | 80.36 | -12.99, 273.86 | 2005-2014 | -6.60 | -13.74, 1.13 | 2014-2017 | 43.15 | -0.57, 106.1 | 12.44 |
| 50+y | 2003-2017 | 2.22 | -2.94, 7.66 | - | - | - | - | - | - | 2.22 |
| Latvia | | | | | | | | | | |
| 20-49y | 2003-2017 | -0.55 | -2.75, 1.7 | - | - | - | - | - | - | -0.55 |
| 50+y | 2003-2011 | -2.91 | -4.09, -1.71 | 2011-2017 | -0.49 | -2.36, 1.42 | - | - | - | -1.88 |
| Lithuania | | | | | | | | | | |
| 20-49y | 2003-2005 | -19.36 | -43.47, 15.04 | 2005-2010 | 5.59 | -5.63, 18.14 | 2010-2017 | -4.79 | -9.2, -0.16 | -3.52 |
| 50+y | 2003-2017 | -2.36 | -2.87, -1.85 | - | - | - | - | - | - | -2.36 |
| Malta | | | | | | | | | | |
| 20-49y | - | - | - | - | - | - | - | - | - | - |
| 50+y | 2003-2017 | -1.38 | -3.08, 0.35 | - | - | - | - | - | - | -1.38 |
| New Zealand | | | | | | | | | | |
| 20-49y | 2003-2017 | -2.38 | -4.04, -0.69 | - | - | - | - | - | - | -2.38 |
| 50+y | 2003-2017 | -1.48 | -2.12, -0.84 | - | - | - | - | - | - | -1.48 |
| Norway | | | | | | | | | | |
| 20-49y | 2003-2017 | -1.08 | -3.29, 1.17 | - | - | - | - | - | - | -1.08 |
| 50+y | 2003-2017 | -2.78 | -3.31, -2.24 | - | - | - | - | - | - | -2.78 |
| Philippines | | | | | | | | | | |
| 20-49y | 2003-2017 | -5.89 | -8.23, -3.49 | - | - | - | - | - | - | -5.89 |

**Appendix Table 15: Segment specific annual percentage change (APC) for cancer incidence trends from 2003 to 2017 by country: stomach cancer**

| | Joinpoint trend 1 | | | Joinpoint trend 2 | | | Joinpoint trend 3 | | | |
|---|---|---|---|---|---|---|---|---|---|---|
| **Country** | **Years** | **APC** | **95% CI** | **Years** | **APC** | **95% CI** | **Years** | **APC** | **95% CI** | **AAPC** |
| 50+y | 2003-2011 | -8.34 | -11.91, -4.62 | 2011-2017 | -0.54 | -6.48, 5.78 | - | - | - | -5.07 |
| Poland | | | | | | | | | | |
| 20-49y | 2003-2017 | -3.53 | -7.94, 1.1 | - | - | - | - | - | - | -3.53 |
| 50+y | 2003-2005 | -14.15 | -29.73, 4.89 | 2005-2017 | -1.69 | -2.84, -0.52 | - | - | - | -3.57 |
| Qatar | | | | | | | | | | |
| 20-49y | 2003-2017 | 1.89 | -4.04, 8.19 | - | - | - | - | - | - | 1.89 |
| 50+y | 2003-2017 | -3.37 | -10.24, 4.01 | - | - | - | - | - | - | -3.37 |
| Republic of Korea | | | | | | | | | | |
| 20-49y | 2003-2010 | -0.24 | -1.16, 0.69 | 2010-2017 | -4.75 | -5.63, -3.86 | - | - | - | -2.52 |
| 50+y | 2003-2011 | -0.08 | -0.94, 0.79 | 2011-2017 | -4.42 | -5.69, -3.12 | - | - | - | -1.96 |
| Slovenia | | | | | | | | | | |
| 20-49y | 2003-2006 | -15.34 | -32.03, 5.43 | 2006-2011 | 6.51 | -7.29, 22.37 | 2011-2017 | -13.73 | -19.9, -7.08 | -7.36 |
| 50+y | 2003-2015 | -2.04 | -2.44, -1.64 | 2015-2017 | -5.93 | -12.17, 0.76 | - | - | - | -2.61 |
| Sweden | | | | | | | | | | |
| 20-49y | 2003-2017 | 0.43 | -1.78, 2.7 | - | - | - | - | - | - | 0.43 |
| 50+y | 2003-2017 | -2.31 | -2.86, -1.76 | - | - | - | - | - | - | -2.31 |
| Switzerland | | | | | | | | | | |
| 20-49y | 2003-2017 | -0.45 | -4.09, 3.32 | - | - | - | - | - | - | -0.45 |
| 50+y | 2003-2017 | -0.82 | -1.83, 0.21 | - | - | - | - | - | - | -0.82 |
| Thailand | | | | | | | | | | |
| 20-49y | 2003-2017 | 1.94 | 0.23, 3.67 | - | - | - | - | - | - | 1.94 |
| 50+y | 2003-2017 | 1.30 | 0.12, 2.5 | - | - | - | - | - | - | 1.30 |
| The Netherlands | | | | | | | | | | |
| 20-49y | 2003-2017 | -1.62 | -2.89, -0.34 | - | - | - | - | - | - | -1.62 |
| 50+y | 2003-2011 | -2.12 | -3.14, -1.08 | 2011-2017 | -4.59 | -6.14, -3.02 | - | - | - | -3.18 |
| Türkiye | | | | | | | | | | |
| 20-49y | 2003-2017 | -0.65 | -1.87, 0.58 | - | - | - | - | - | - | -0.65 |
| 50+y | 2003-2007 | 1.01 | -2.08, 4.21 | 2007-2017 | -1.28 | -2.03, -0.52 | - | - | - | -0.63 |
| UK | | | | | | | | | | |
| 20-49y | 2003-2017 | -0.83 | -1.48, -0.17 | - | - | - | - | - | - | -0.83 |
| 50+y | 2003-2017 | -3.52 | -3.7, -3.34 | - | - | - | - | - | - | -3.52 |
| USA | | | | | | | | | | |
| 20-49y | 2003-2017 | 0.39 | -0.4, 1.19 | - | - | - | - | - | - | 0.39 |
| 50+y | 2003-2017 | -1.52 | -1.8, -1.24 | - | - | - | - | - | - | -1.52 |
| Uganda | | | | | | | | | | |
| 20-49y | 2003-2017 | -3.04 | -9.09, 3.42 | - | - | - | - | - | - | -3.04 |
| 50+y | 2003-2017 | -3.87 | -7.5, -0.1 | - | - | - | - | - | - | -3.87 |

**Appendix Table 16: Segment specific annual percentage change (APC) for cancer incidence trends from 2003 to 2017 by country: thyroid cancer**

| | Joinpoint trend 1 | | | Joinpoint trend 2 | | | Joinpoint trend 3 | | | |
|---|---|---|---|---|---|---|---|---|---|---|
| Country | Years | APC | 95% CI | Years | APC | 95% CI | Years | APC | 95% CI | AAPC |
| Argentina | | | | | | | | | | |
| 20-49y | 2003-2017 | 2.35 | -1.21, 6.05 | - | - | - | - | - | - | 2.35 |
| 50+y | 2003-2017 | 0.92 | -1.03, 2.9 | - | - | - | - | - | - | 0.92 |
| Australia | | | | | | | | | | |
| 20-49y | 2003-2013 | 4.06 | 3.45, 4.68 | 2013-2017 | 1.57 | -0.83, 4.02 | - | - | - | 3.34 |
| 50+y | 2003-2014 | 6.02 | 5.58, 6.46 | 2014-2017 | 0.65 | -2.42, 3.82 | - | - | - | 4.85 |
| Austria | | | | | | | | | | |
| 20-49y | 2003-2008 | 11.41 | 6.78, 16.23 | 2008-2017 | -1.74 | -3.43, -0.02 | - | - | - | 2.77 |
| 50+y | 2003-2008 | 8.34 | 4.53, 12.3 | 2008-2017 | -4.29 | -5.68, -2.88 | - | - | - | 0.04 |
| Bahrain | | | | | | | | | | |
| 20-49y | 2003-2007 | -27.71 | -50.05, 4.62 | 2007-2017 | 13.70 | 3.81, 24.53 | - | - | - | -0.10 |
| 50+y | 2003-2009 | -12.48 | -28.29, 6.82 | 2009-2017 | 9.87 | -3.39, 24.94 | - | - | - | -0.34 |
| Belarus | | | | | | | | | | |
| 20-49y | 2003-2014 | 1.19 | 0.42, 1.96 | 2014-2017 | 7.89 | 1.97, 14.16 | - | - | - | 2.59 |
| 50+y | 2003-2017 | -0.15 | -0.87, 0.57 | - | - | - | - | - | - | -0.15 |
| Canada | | | | | | | | | | |
| 20-49y | 2003-2014 | 4.78 | 4.15, 5.42 | 2014-2017 | -6.88 | -10.99, -2.58 | - | - | - | 2.16 |
| 50+y | 2003-2013 | 6.89 | 5.82, 7.97 | 2013-2017 | -4.83 | -8.64, -0.85 | - | - | - | 3.40 |
| Chile | | | | | | | | | | |
| 20-49y | 2003-2017 | 10.68 | 5.38, 16.24 | - | - | - | - | - | - | 10.68 |
| 50+y | 2003-2013 | 1.36 | -4.54, 7.63 | 2013-2017 | 31.76 | 3.25, 68.16 | - | - | - | 9.25 |
| China | | | | | | | | | | |
| 20-49y | 2003-2008 | 15.46 | 9.63, 21.6 | 2008-2013 | 28.23 | 19.17, 37.98 | 2013-2017 | 9.25 | 1.53, 17.56 | 17.99 |
| 50+y | 2003-2014 | 18.61 | 16.81, 20.44 | 2014-2017 | 3.07 | -8, 15.47 | - | - | - | 15.09 |
| Colombia | | | | | | | | | | |
| 20-49y | 2003-2009 | 2.82 | -1.8, 7.65 | 2009-2017 | 9.20 | 6.01, 12.49 | - | - | - | 6.42 |
| 50+y | 2003-2017 | 7.37 | 5.93, 8.82 | - | - | - | - | - | - | 7.37 |
| Croatia | | | | | | | | | | |
| 20-49y | 2003-2017 | 6.62 | 5.54, 7.7 | - | - | - | - | - | - | 6.62 |
| 50+y | 2003-2015 | 4.38 | 3.63, 5.14 | 2015-2017 | -4.90 | -15.86, 7.48 | - | - | - | 3.00 |
| Cyprus | | | | | | | | | | |
| 20-49y | 2003-2017 | 10.94 | 8.81, 13.1 | - | - | - | - | - | - | 10.94 |
| 50+y | 2003-2017 | 15.71 | 13.11, 18.36 | - | - | - | - | - | - | 15.71 |
| Czechia | | | | | | | | | | |
| 20-49y | 2003-2017 | 5.38 | 4.73, 6.04 | - | - | - | - | - | - | 5.38 |
| 50+y | 2003-2005 | 16.07 | -3.87, 40.15 | 2005-2015 | 1.72 | -0.02, 3.49 | 2015-2017 | -6.74 | -22.77, 12.6 | 2.38 |
| Denmark | | | | | | | | | | |
| 20-49y | 2003-2006 | 1.46 | -7.64, 11.45 | 2006-2017 | 7.88 | 6.52, 9.25 | - | - | - | 6.47 |
| 50+y | 2003-2017 | 7.39 | 5.46, 9.34 | - | - | - | - | - | - | 7.39 |
| Ecuador | | | | | | | | | | |
| 20-49y | 2003-2012 | 13.95 | 9.8, 18.26 | 2012-2017 | 4.82 | -4.29, 14.8 | - | - | - | 10.60 |
| 50+y | 2003-2017 | 10.67 | 8.84, 12.52 | - | - | - | - | - | - | 10.67 |
| Estonia | | | | | | | | | | |
| 20-49y | 2003-2017 | 0.30 | -2.39, 3.07 | - | - | - | - | - | - | 0.30 |

**Appendix Table 16: Segment specific annual percentage change (APC) for cancer incidence trends from 2003 to 2017 by country: thyroid cancer**

| | Joinpoint trend 1 | | | Joinpoint trend 2 | | | Joinpoint trend 3 | | | |
|---|---|---|---|---|---|---|---|---|---|---|
| Country | Years | APC | 95% CI | Years | APC | 95% CI | Years | APC | 95% CI | AAPC |
| 50+y | 2003-2017 | -0.09 | -2.27, 2.15 | - | - | - | - | - | - | -0.09 |
| Finland | | | | | | | | | | |
| 20-49y | 2003-2005 | -9.17 | -23.49, 7.83 | 2005-2017 | 3.84 | 2.8, 4.9 | - | - | - | 1.88 |
| 50+y | 2003-2005 | -8.84 | -25.23, 11.14 | 2005-2017 | 4.77 | 3.55, 6.01 | - | - | - | 2.71 |
| France | | | | | | | | | | |
| 20-49y | 2003-2013 | 4.17 | 2.93, 5.42 | 2013-2017 | -4.72 | -9.23, 0.02 | - | - | - | 1.55 |
| 50+y | 2003-2009 | 4.97 | 2.3, 7.71 | 2009-2015 | -0.04 | -3.39, 3.43 | 2015-2017 | -11.44 | -23.97, 3.15 | 0.33 |
| Germany | | | | | | | | | | |
| 20-49y | 2003-2017 | 6.16 | 4.42, 7.93 | - | - | - | - | - | - | 6.16 |
| 50+y | 2003-2017 | 3.41 | 2.17, 4.66 | - | - | - | - | - | - | 3.41 |
| Iceland | | | | | | | | | | |
| 20-49y | 2003-2017 | 2.09 | -2.73, 7.15 | - | - | - | - | - | - | 2.09 |
| 50+y | 2003-2017 | -0.56 | -5.86, 5.04 | - | - | - | - | - | - | -0.56 |
| India | | | | | | | | | | |
| 20-49y | 2003-2017 | 1.34 | 0.25, 2.44 | - | - | - | - | - | - | 1.34 |
| 50+y | 2003-2017 | 2.17 | 0.61, 3.76 | - | - | - | - | - | - | 2.17 |
| Ireland | | | | | | | | | | |
| 20-49y | 2003-2013 | 11.33 | 8.21, 14.54 | 2013-2017 | -6.24 | -16.46, 5.24 | - | - | - | 6.00 |
| 50+y | 2003-2013 | 9.85 | 6.88, 12.91 | 2013-2017 | -5.56 | -15.52, 5.59 | - | - | - | 5.21 |
| Israel | | | | | | | | | | |
| 20-49y | 2003-2014 | 3.86 | 2.54, 5.2 | 2014-2017 | -8.16 | -16.48, 1 | - | - | - | 1.16 |
| 50+y | 2003-2013 | 3.06 | 1.97, 4.16 | 2013-2017 | -7.39 | -11.29, -3.31 | - | - | - | -0.04 |
| Italy | | | | | | | | | | |
| 20-49y | 2003-2017 | 1.66 | 0.32, 3.02 | - | - | - | - | - | - | 1.66 |
| 50+y | 2003-2017 | 2.20 | 0.87, 3.55 | - | - | - | - | - | - | 2.20 |
| Kuwait | | | | | | | | | | |
| 20-49y | 2003-2007 | -8.45 | -14.65, -1.79 | 2007-2017 | 8.67 | 6.8, 10.56 | - | - | - | 3.47 |
| 50+y | 2003-2005 | 65.65 | -22.71, 255.06 | 2005-2017 | 3.22 | -1.33, 7.98 | - | - | - | 10.44 |
| Latvia | | | | | | | | | | |
| 20-49y | 2003-2013 | 14.04 | 9.72, 18.53 | 2013-2017 | 0.48 | -14.11, 17.56 | - | - | - | 9.99 |
| 50+y | 2003-2008 | 14.83 | 9, 20.98 | 2008-2017 | 4.71 | 2.5, 6.96 | - | - | - | 8.22 |
| Lithuania | | | | | | | | | | |
| 20-49y | 2003-2017 | 1.96 | 0.73, 3.21 | - | - | - | - | - | - | 1.96 |
| 50+y | 2003-2008 | 7.06 | 2.78, 11.52 | 2008-2017 | -2.24 | -3.86, -0.6 | - | - | - | 0.98 |
| Malta | | | | | | | | | | |
| 20-49y | 2003-2017 | 6.77 | 2.84, 10.85 | - | - | - | - | - | - | 6.77 |
| 50+y | 2003-2017 | 10.47 | 4.83, 16.41 | - | - | - | - | - | - | 10.47 |
| New Zealand | | | | | | | | | | |
| 20-49y | 2003-2017 | 3.66 | 2.29, 5.06 | - | - | - | - | - | - | 3.66 |
| 50+y | 2003-2017 | 2.68 | 1.77, 3.6 | - | - | - | - | - | - | 2.68 |
| Norway | | | | | | | | | | |
| 20-49y | 2003-2017 | 5.05 | 3.8, 6.31 | - | - | - | - | - | - | 5.05 |
| 50+y | 2003-2009 | 1.66 | -1.62, 5.05 | 2009-2017 | 6.89 | 4.65, 9.18 | - | - | - | 4.62 |
| Philippines | | | | | | | | | | |
| 20-49y | 2003-2012 | -4.07 | -6.59, -1.49 | 2012-2015 | 15.08 | -14, 53.99 | 2015-2017 | -8.32 | -31.49, 22.67 | -0.90 |

**Appendix Table 16: Segment specific annual percentage change (APC) for cancer incidence trends from 2003 to 2017 by country: thyroid cancer**

| | Joinpoint trend 1 | | | Joinpoint trend 2 | | | Joinpoint trend 3 | | | |
|---|---|---|---|---|---|---|---|---|---|---|
| Country | Years | APC | 95% CI | Years | APC | 95% CI | Years | APC | 95% CI | AAPC |
| 50+y | 2003-2012 | -3.48 | -6.12, -0.76 | 2012-2015 | 15.57 | -14.74, 56.66 | 2015-2017 | -12.92 | -35.75, 18.04 | -1.15 |
| Poland | | | | | | | | | | |
| 20-49y | 2003-2010 | 3.11 | -1.45, 7.88 | 2010-2015 | 19.37 | 7.26, 32.85 | 2015-2017 | -4.56 | -31.95, 33.85 | 7.45 |
| 50+y | 2003-2017 | 2.13 | 0.5, 3.78 | - | - | - | - | - | - | 2.13 |
| Qatar | | | | | | | | | | |
| 20-49y | 2003-2012 | -7.13 | -16.42, 3.2 | 2012-2017 | 19.28 | -7.87, 54.43 | - | - | - | 1.55 |
| 50+y | - | - | - | - | - | - | - | - | - | - |
| Republic of Korea | | | | | | | | | | |
| 20-49y | 2003-2011 | 22.14 | 18.37, 26.03 | 2011-2017 | -9.12 | -13.43, -4.6 | - | - | - | 7.60 |
| 50+y | 2003-2011 | 21.91 | 18, 25.96 | 2011-2017 | -14.20 | -18.43, -9.75 | - | - | - | 4.88 |
| Slovenia | | | | | | | | | | |
| 20-49y | 2003-2017 | 2.45 | 0.94, 3.99 | - | - | - | - | - | - | 2.45 |
| 50+y | 2003-2017 | 2.56 | 1.15, 3.98 | - | - | - | - | - | - | 2.56 |
| Sweden | | | | | | | | | | |
| 20-49y | 2003-2017 | 5.50 | 4.47, 6.54 | - | - | - | - | - | - | 5.50 |
| 50+y | 2003-2017 | 4.05 | 3.29, 4.82 | - | - | - | - | - | - | 4.05 |
| Switzerland | | | | | | | | | | |
| 20-49y | 2003-2013 | 7.30 | 4.6, 10.07 | 2013-2017 | -4.77 | -14.15, 5.62 | - | - | - | 3.70 |
| 50+y | 2003-2017 | 4.45 | 2.52, 6.42 | - | - | - | - | - | - | 4.45 |
| Thailand | | | | | | | | | | |
| 20-49y | 2003-2005 | -13.71 | -43.53, 31.84 | 2005-2017 | 6.25 | 3.62, 8.95 | - | - | - | 3.14 |
| 50+y | 2003-2017 | 2.73 | 1.26, 4.22 | - | - | - | - | - | - | 2.73 |
| The Netherlands | | | | | | | | | | |
| 20-49y | 2003-2017 | 4.63 | 3.77, 5.49 | - | - | - | - | - | - | 4.63 |
| 50+y | 2003-2017 | 4.38 | 3.36, 5.4 | - | - | - | - | - | - | 4.38 |
| Türkiye | | | | | | | | | | |
| 20-49y | 2003-2005 | 57.15 | 28.61, 92.03 | 2005-2017 | 6.44 | 5.19, 7.71 | - | - | - | 12.53 |
| 50+y | 2003-2008 | 17.14 | 11.91, 22.61 | 2008-2017 | 3.02 | 1.12, 4.96 | - | - | - | 7.86 |
| UK | | | | | | | | | | |
| 20-49y | 2003-2013 | 7.26 | 6.79, 7.73 | 2013-2017 | 3.36 | 1.52, 5.24 | - | - | - | 6.13 |
| 50+y | 2003-2014 | 6.38 | 5.57, 7.2 | 2014-2017 | 2.23 | -3.39, 8.18 | - | - | - | 5.48 |
| USA | | | | | | | | | | |
| 20-49y | 2003-2009 | 6.09 | 5.12, 7.08 | 2009-2015 | 2.03 | 0.8, 3.28 | 2015-2017 | -4.48 | -9.56, 0.87 | 2.78 |
| 50+y | 2003-2009 | 7.46 | 5.79, 9.15 | 2009-2015 | 0.69 | -1.38, 2.79 | 2015-2017 | -6.48 | -14.76, 2.6 | 2.45 |
| Uganda | | | | | | | | | | |
| 20-49y | 2003-2013 | -0.55 | -6.95, 6.3 | 2013-2017 | -25.44 | -43.11, -2.29 | - | - | - | -8.41 |
| 50+y | 2003-2017 | -4.25 | -10.46, 2.38 | - | - | - | - | - | - | -4.25 |

Table 17: Number and % of eligible countries in Europe with increasing cancer rates (AAPC>0) in younger adults, and also in older adults and the number of countries with cancer rates in younger adults increasing at a faster rate than in older adults

| | **Eligible countries (age 20–49y)** | **↑ age 20–49y** | **↑ age 20–49y & ↑ 50+y** | **↑ age 20–49y > 50+y** |
|---|---|---|---|---|
| **Cancer** | **n (%)[1]** | **n (%)** | **n (%)** | **n (%)** |
| Breast | 22 (100) | 22 (100) | 16 (73) | 18 (82) |
| Thyroid | 22 (100) | 22 (100) | 19 (86) | 17 (77) |
| Colorectum | 22 (100) | 20 (91) | 11 (50) | 18 (82) |
| Kidney | 22 (100) | 18 (82) | 17 (77) | 12 (55) |
| Leukaemia | 22 (100) | 18 (82) | 12 (55) | 15 (68) |
| Prostate | 20 (100) | 16 (80) | 12 (60) | 12 (60) |
| Endometrium | 21 (100) | 14 (67) | 9 (43) | 12 (57) |
| Gallbladder | 15 (100) | 9 (60) | 7 (47) | 6 (40) |
| Liver | 20 (100) | 11 (55) | 11 (55) | 4 (20) |
| Pancreas | 20 (100) | 11 (55) | 10 (50) | 4 (20) |
| Oesophagus | 19 (100) | 9 (47) | 7 (37) | 5 (26) |
| Oral | 21 (100) | 9 (43) | 9 (43) | 6 (29) |
| Stomach | 20 (100) | 2 (10) | 0 (0) | 2 (10) |

[1]AAPC was not estimable if countries had 0 cases in any year.

Table 18: Number and % of eligible countries in Americas with increasing cancer rates (AAPC>0) in younger adults, and also in older adults and the number of countries with cancer rates in younger adults increasing at a faster rate than in older adults

| | **Eligible countries (age 20–49y)** | **↑ age 20–49y** | **↑ age 20–49y & ↑ 50+y** | **↑ age 20–49y > 50+y** |
|---|---|---|---|---|
| **Cancer** | **n (%)**[1] | **n (%)** | **n (%)** | **n (%)** |
| Colorectum | 6 (100) | 6 (100) | 4 (67) | 5 (83) |
| Endometrium | 5 (100) | 5 (100) | 5 (100) | 4 (80) |
| Kidney | 6 (100) | 6 (100) | 4 (67) | 5 (83) |
| Thyroid | 6 (100) | 6 (100) | 6 (100) | 2 (33) |
| Breast | 6 (100) | 5 (83) | 4 (67) | 2 (33) |
| Leukaemia | 5 (100) | 4 (80) | 4 (80) | 2 (40) |
| Pancreas | 4 (100) | 3 (75) | 3 (75) | 2 (50) |
| Gallbladder | 5 (100) | 3 (60) | 1 (20) | 3 (60) |
| Stomach | 6 (100) | 3 (50) | 1 (17) | 3 (50) |
| Liver | 5 (100) | 2 (40) | 2 (40) | 1 (20) |
| Oral | 5 (100) | 2 (40) | 2 (40) | 1 (20) |
| Prostate | 5 (100) | 2 (40) | 1 (20) | 2 (40) |
| Oesophagus | 3 (100) | 0 (0) | 0 (0) | 0 (0) |

[1]AAPC was not estimable if countries had 0 cases in any year.

Table 19: Number and % of eligible countries in Asia with increasing cancer rates (AAPC>0) in younger adults, and also in older adults and the number of countries with cancer rates in younger adults increasing at a faster rate than in older adults

| | **Eligible countries (age 20–49y)** | **↑ age 20–49y** | **↑ age 20–49y & ↑ 50+y** | **↑ age 20–49y > 50+y** |
|---|---|---|---|---|
| **Cancer** | **n (%)[1]** | **n (%)** | **n (%)** | **n (%)** |
| Kidney | 10 (100) | 8 (80) | 6 (60) | 7 (70) |
| Thyroid | 10 (100) | 8 (80) | 7 (70) | 5 (50) |
| Endometrium | 8 (100) | 6 (75) | 6 (75) | 4 (50) |
| Breast | 11 (100) | 8 (73) | 8 (73) | 2 (18) |
| Colorectum | 11 (100) | 8 (73) | 7 (64) | 4 (36) |
| Pancreas | 9 (100) | 6 (67) | 6 (67) | 2 (22) |
| Leukaemia | 10 (100) | 6 (60) | 4 (40) | 4 (40) |
| Gallbladder | 7 (100) | 4 (57) | 3 (43) | 1 (14) |
| Prostate | 7 (100) | 4 (57) | 4 (57) | 2 (29) |
| Oral | 10 (100) | 5 (50) | 3 (30) | 5 (50) |
| Stomach | 10 (100) | 3 (30) | 2 (20) | 2 (20) |
| Liver | 7 (100) | 2 (29) | 2 (29) | 1 (14) |
| Oesophagus | 7 (100) | 1 (14) | 0 (0) | 1 (14) |

[1]AAPC was not estimable if countries had 0 cases in any year.

Table 20: Number and % of eligible countries in Europe with increasing cancer rates (AAPC>0) in younger adults, and also in older adults and the number of countries with cancer rates in younger adults increasing at a faster rate than in older adults: females

| | **Eligible countries (age 20-49y)** | **↑ age 20-49y** | **↑ age 20-49y & ↑ 50+y** | **↑ age 20-49y > 50+y** |
|---|---|---|---|---|
| **Cancer** | **n (%)**[1] | **n (%)** | **n (%)** | **n (%)** |
| Thyroid | 41 (100) | 38 (93) | 35 (85) | 22 (54) |
| Breast | 42 (100) | 38 (90) | 30 (71) | 23 (55) |
| Colorectum | 41 (100) | 37 (90) | 21 (51) | 32 (78) |
| Kidney | 35 (100) | 27 (77) | 24 (69) | 20 (57) |
| Pancreas | 33 (100) | 25 (76) | 23 (70) | 17 (52) |
| Endometrium | 36 (100) | 27 (75) | 22 (61) | 22 (61) |
| Leukaemia | 38 (100) | 22 (58) | 16 (42) | 13 (34) |
| Liver | 28 (100) | 16 (57) | 13 (46) | 6 (21) |
| Gallbladder | 27 (100) | 15 (56) | 8 (30) | 11 (41) |
| Oral | 36 (100) | 18 (50) | 14 (39) | 11 (31) |
| Oesophagus | 15 (100) | 5 (33) | 4 (27) | 3 (20) |
| Stomach | 35 (100) | 10 (29) | 3 (9) | 10 (29) |

[1]AAPC was not estimable if countries had 0 cases in any year.

Table 21: Number and % of eligible countries in Europe with increasing cancer rates (AAPC>0) in younger adults, and also in older adults and the number of countries with cancer rates in younger adults increasing at a faster rate than in older adults

| | Eligible countries (age 20-49y) | ↑ age 20-49y | ↑ age 20-49y & ↑ 50+y | ↑ age 20-49y > 50+y |
|---|---|---|---|---|
| **Cancer** | **n countries[1]** | **n (%)** | **n (%)** | **n (%)** |
| Thyroid | 34 (100) | 34 (100) | 32 (94) | 22 (65) |
| Kidney | 35 (100) | 29 (83) | 27 (77) | 24 (69) |
| Colorectum | 41 (100) | 32 (78) | 20 (49) | 25 (61) |
| Leukaemia | 39 (100) | 28 (72) | 22 (56) | 20 (51) |
| Prostate | 35 (100) | 23 (66) | 17 (49) | 17 (49) |
| Gallbladder | 22 (100) | 13 (59) | 13 (59) | 8 (36) |
| Pancreas | 33 (100) | 19 (58) | 18 (55) | 10 (30) |
| Oral | 37 (100) | 17 (46) | 15 (41) | 12 (32) |
| Liver | 32 (100) | 12 (38) | 12 (38) | 2 (6) |
| Oesophagus | 32 (100) | 10 (31) | 10 (31) | 4 (12) |
| Stomach | 38 (100) | 5 (13) | 2 (5) | 5 (13) |

[1]AAPC was not estimable if countries had 0 cases in any year.
